# Anatomical comparison populations and threshold transport in vascular transcriptomic classification

**DOI:** 10.64898/2026.10.29.26366564

**Authors:** Lingsen You, Zisu Peng, Yujun Guo, Li Shen, Junbo Ge

**Author notes:** Lingsen You, Zisu Peng, and Yujun Guo contributed equally to this work. Correspondence: Lingsen You; Junbo Ge.

## Abstract

**Background:** High vascular transcriptomic discrimination may obscure dependence on comparison populations, gene selection, and score scale. We examined these properties separately.

**Methods:** GSE100927 compared surgical atherosclerotic tissue with lesion-free deceased-donor arteries: 104 arrays formed 95 complete-label groups, versus 96 arteries reported originally; independent donor identities remained unresolved. Within held-out folds, pooled area under the receiver-operating-characteristic curve (AUROC) was decomposed into nine case-bed–control-bed comparisons. Seven fixed rules underwent ten repeated three-fold splits. The primary ridge model used sample-wise gene ranks and training-only selection of 200 genes. External tests involved 32 paired plaque/intact patients in GSE43292 and 29 early/advanced specimens in GSE28829. Post-result explorations added random and disjoint gene-panel controls, feature stability, frozen-score accounting, source-only threshold learning, and individual-failure analyses.

**Results:** Primary mean held-out-fold pooled AUROC was 0.9968, versus a within-bed mean of 1.0000 and contrast −0.0032; the empirical ceiling constrained this contrast to be nonpositive. Paired external AUROC was 0.877 (conditional 95% interval 0.792–0.947), with plaque ranked higher in 29/32 patients. Histological AUROC was 0.986 (0.942–1.000). All primary external probabilities fell below 0.5. Both source-derived thresholds also gave zero external sensitivity. Twenty outcome-independent random panels retained source AUROCs of 0.9932–0.9992, while external ranking varied substantially. Across 30 primary fits, 619 genes were selected at least once and 45 every time. Fixed-score failure analyses distinguished shared from model-specific discordance.

**Conclusions:** Strong source ranking did not establish a unique gene signature or transferable technical thresholds. Procurement confounding, uncertain donor identities, inherited preprocessing, changed endpoints, and the exploratory status of added analyses limit interpretation. This is a comparison-population audit, not a clinically calibrated risk model.

## 1 Introduction

Panvascular research seeks to understand disease across anatomically distinct vascular territories. Transcriptomic studies have identified both shared and bed-associated expression patterns in human arteries, providing a basis for investigating common disease signals and local biology [1]. Recent conceptual frameworks also emphasize this dual requirement. FLOW proposes integrating heterogeneous vascular beds within a systemic modeling framework, whereas the tri-ecological account of device–vessel matching makes the relevant biological context dependent on the particular bed, lesion, and patient [2, 3]. These proposals motivate explicit tests of transfer; they do not establish that a molecular classifier trained in one tissue comparison will retain its meaning in another.

A recent immune-world-model preprint evaluates intervention-dependent state transitions across biological scales using computational benchmarks [4]. Our static comparison-population audit examines a complementary measurement problem and does not test intervention forecasts or clinical efficacy.

The availability of public arterial expression cohorts has already enabled multi-bed biomarker studies. Wu and colleagues combined arterial datasets to derive candidate genes, while Cui and colleagues evaluated C1Q-related signatures across carotid and other arterial tissues [5, 6]. Consequently, another gene list or another high pooled AUROC using these cohorts would provide limited independent information. A separate question concerns the comparisons that create the reported performance: which diseased specimen is being ranked above which comparator, and under what anatomical and sampling distribution?

AUROC can be interpreted as the probability that a positive observation receives a higher score than a negative observation, with half credit for ties [7]. In a multi-bed cohort, that probability combines comparisons within the same bed and comparisons between different beds. Its anatomical composition is therefore part of the estimand. Related work on matched classification studies shows why the comparison population must be considered when interpreting accuracy [8]. This issue is distinct from training leakage: correctly nested feature selection does not by itself determine whether the resulting comparisons answer the intended biological question.

A second distinction separates ranking from the transport of a decision threshold. A model may preserve tissue ordering after a change of platform, recruitment, or comparator while shifting its entire score distribution. Moreover, a healthy-donor comparison, plaque versus relatively intact tissue from the same patient, and early versus advanced histology represent different endpoints. Treating them as interchangeable external replications can obscure both preserved information and important failures.

We therefore conducted an anatomical comparison-population audit. The primary question concerned the difference between pooled disease-versus-donor discrimination and within-bed discrimination; the prespecified contrast was pooled AUROC minus the equally weighted mean of the three within-bed AUROCs. An exact nine-cell decomposition under a common held-out-fold scoring model evaluated that comparison. Anatomical holdouts, patient-paired lesion ranking, and an external histological contrast then examined distinct transport questions. The protocol identified strong diagonal discrimination with a narrow reweighting range as evidence against marked composition sensitivity in the evaluated samples, without specifying a one-sided positive-inflation hypothesis or an equivalence margin. Preserved external ranking accompanied by threshold failure would identify a separate limitation of score transport.

## 2 Methods

### 2.1 Study design, prior exposure, and analysis record

This was a secondary computational analysis of publicly deposited human arterial tissue measurements obtained from Gene Expression Omnibus (GEO) [9]. It used neither simulated patients nor synthetic expression observations. The internal protocol and a pre-fit amendment were recorded on 25 October 2026 before the first fitted model in this project. They were not a registry-based preregistration. The matrices had been obtained in earlier work, and dataset metadata, source publications, and published secondary findings were already known. GSE43292 had also supported descriptive module contrasts in the separate VascFlux-GEO project. Those results were not reused as new findings here; external cohorts were not described as previously unseen.

The related VascBridge-Instability project evaluated GSE98278-derived rupture-status classifiers across symptom, rupture-related, and censored-event endpoints; it did not analyze GSE100927. Conventional rank-based modeling and assessment of transfer failures are therefore shared approaches. The present question instead concerns anatomical case–control comparisons, followed by separately interpreted paired lesion ranking.

An implementation note written after initial results corrected the feature-ranking terminology: the executed statistic was a Welch-type standardized difference, not a Cohen standardized mean difference. The executed algorithm was retained and is defined below. The note also documented a stricter decomposition check: a failed algebraic identity raises an error, while a missing bed-by-status cell is recorded explicitly. This checking change did not alter saved predictions. No externally best-performing model was substituted for the prespecified primary model.

### 2.2 Cohorts and analysis units

GSE100927 was the development and anatomical-holdout dataset, measured on Agilent GPL17077 [1]. Its 104 deposited arrays comprised 69 diseased and 35 control arterial specimens across carotid, femoral, and infrapopliteal beds. Disease samples were surgically obtained atherosclerotic tissue; controls were arteries without atherosclerotic lesions from deceased organ donors. The source paper’s expression methods reported 65 diseased and 31 healthy arteries, totaling 96, which did not match the deposited column count.

The complete specimen label following the colon in each deposited title occurred twice for nine labels. We conservatively aggregated each complete-label group, yielding 95 groups: 65 disease and 30 control. These groups comprised 27/10 carotid, 25/10 femoral, and 13/10 infrapopliteal disease/control units. The distinction between 95 grouped labels and the source paper’s 96 arteries, including the femoral control discrepancy, remains unresolved. Aggregation is an analysis convention, not proof of technical replication or independent donor identity. Numerical suffixes were not treated as universally unique donor identifiers. Alternative identity conventions were examined rather than selecting a single interpretation as established fact.

GSE43292, measured on Affymetrix GPL6244, supplied 64 carotid tissues from 32 hypertensive endarterectomy patients [10]. Each patient contributed plaque core/shoulder tissue with advanced histology and distant, macroscopically intact tissue classified as Stary I/II. The comparator was therefore neither a healthy donor nor a disease-free patient. Patient identifiers supported paired comparisons and patient-block resampling.

GSE28829, measured on Affymetrix GPL570, supplied 29 autopsy carotid specimens: 13 early lesions and 16 advanced lesions [11]. Early lesions included pathological intimal thickening or intimal xanthoma; advanced lesions included thin- or thick-fibrous-cap atheroma. Public metadata did not establish an independent donor count. This dataset tested a cross-sectional histological contrast, not longitudinal progression. No cohort contributed analyzed follow-up or censoring outcomes, and sample counts were not added to claim a combined independent-patient total. Dataset roles and source-unit structure are summarized in Figure 1.

**Figure 1:**
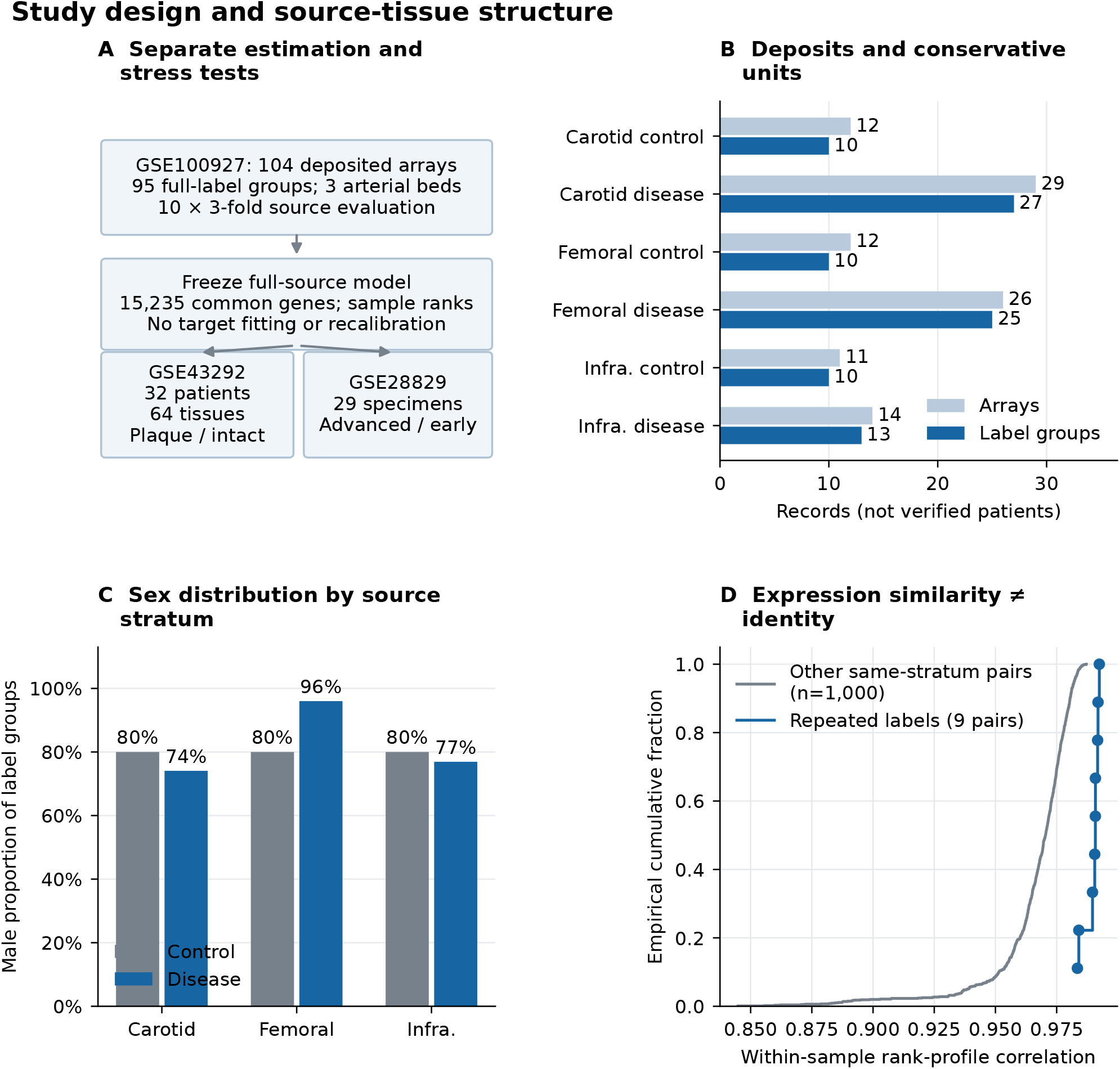
Study design and source-tissue structure. A, Study structure; the full-source frozen model is applied without target fitting. B, 104 deposited arrays are conservatively aggregated into 95 full-label groups (65 disease, 30 control), not authenticated independent patients. C, Male proportions illustrate a covariate available to the bed-and-sex control. D, Rank-profile similarity of the nine repeated-label pairs versus other pairs within the same bed and status. Pairwise comparisons are dependent and descriptive; high correlation does not authenticate donor identity. Infra., infrapopliteal.

### 2.3 Expression processing and common vocabulary

Official processed matrices, sample metadata, and platform annotations were downloaded from GEO on 25 October 2026, with source URLs and SHA256 hashes retained. The deposited GSE100927 matrix contained 47,264 probe rows before our mapping and filtering; the source paper described a 20,000-probe filter. This study therefore analyzed the deposited processed matrix anew rather than exactly reimplementing that filtered analysis. Probes were kept only when their annotation mapped unambiguously to one gene symbol. Multiple eligible probes for a gene were summarized by the median within each sample. The common vocabulary contained 15,235 genes present in the three platform annotations and available matrix probes. Thus, target input availability informed the vocabulary, although target expression distributions and outcome associations did not inform feature selection. This is declared use of target input information, not a claim of completely untouched inductive development.

The primary representation was each sample’s fractional gene ranks within that vocabulary. Ranks were calculated before median aggregation of repeated complete-label groups. No gene-level missingness remained in the analyzed matrices, so fitted imputation was unnecessary. For the native-expression sensitivity, all matrices were retained on their deposited scale because their values already occupied a finite log-intensity range; no additional logarithmic transformation was performed. Gene centering and scaling were fitted on each training set and applied unchanged to its held-out or external data.

These operations did not reverse the original normalization. GSE100927 had been normalized against an across-array reference; the Affymetrix datasets had undergone cohort-level RMA processing. Consequently, the study did not reproduce a prospective raw-array pipeline in which every preprocessing operation was learned without held-out samples. Source-only secondary scaling and selection address a narrower leakage risk. Methodological work has shown that batch and source confounding can remain problematic even when cross-validation is otherwise carefully organized [12]. We did not pool cohorts for batch correction, fit a target transformation, or recalibrate on external outcomes.

### 2.4 Fixed models and training-only operations

Seven fixed specifications were evaluated: a training-prevalence intercept; logistic regression using arterial-bed indicators and sex; the mean ranks of C1QA and C1QC; logistic regression using the published six-gene panel ADAP2, CSF3R, FABP5, ITGAX, MYOC, and SPP1; the primary ridge logistic model; a signed standardized centroid; and a random forest. The two literature panels were retrospective controls: Wu’s selection included GSE100927, whereas Cui’s C1QA/C1QC selection used GSE43292, with published evaluation also including GSE100927 and GSE28829 [5, 6]. Their historical information exposure therefore differed from the fold-selected primary model. Our simple implementations were not represented as exact reproductions of those publications or as independent biomarker confirmation.

For the three 200-gene models, genes were ranked within the training set by

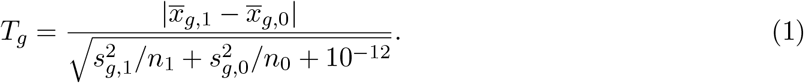

This absolute Welch-type statistic was used for feature ordering, not hypothesis testing. The primary model selected the top 200 genes, standardized them with training-set means and standard deviations, and fitted L2 logistic regression with *C* = 0.1 and balanced class weights. The six-gene and bed/sex logistic controls used the same regularization setting. The centroid averaged standardized selected features after multiplying by the sign of their training-set disease/control difference. The forest used 300 trees, maximum depth 3, minimum leaf size 3, square-root feature sampling, and balanced class weights.

All data-driven feature ranking, scaling, and model fitting performed in this secondary analysis used the relevant training set; the fixed literature panels retained the prior cohort exposure described above. Hyperparameters were fixed rather than selected using held-out performance, addressing the distinction between fitting a prespecified model and optimizing a model-selection procedure [13, 14]. These secondary fitting boundaries did not remove the inherited cohort-wide normalization. The C1Q and centroid outputs were treated as scores: they received ranking metrics but no probability-threshold or Brier interpretation.

### 2.5 Primary comparison-population estimand

For a single held-out-fold scoring function, let *A_ab_* denote the fraction of all case-bed-*a*/control-bed-*b* pairs in which the case has the higher score, counting a tie as one half. For the three beds, the nine values form a matrix. If *n*_1_*_a_* and *n*_0_*_b_* are the held-out counts, then

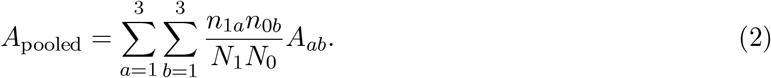

This is an empirical partition of the ordinary AUROC, not a new estimator. We verified its equality numerically within every complete held-out fold. The primary contrast was

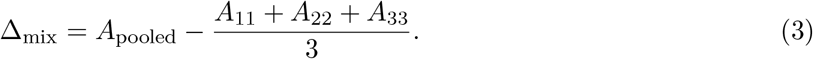

It compares the observed pair mixture with an equal-weight mean of within-bed comparisons. It does not estimate a causal effect of anatomy.

A secondary grid varied case-bed and control-bed proportions independently in increments of 0.1. Each three-bed simplex had 66 compositions, giving 4,356 pair-mixture scenarios per fold. For case and control composition vectors *p* and *q*, the reweighted value *p*^T^*Aq* = Σ*_a,b_ p_a_q_b_A_ab_* is a convex combination of the nine cells. Because the grid includes all simplex vertices, its minimum and maximum equal min*_a,b_ A_ab_* and max*_a,b_ A_ab_*; interior grid points describe additional scenarios without adding information about these extrema. Extreme scenarios can place all cases in one bed and all controls in another and are not asserted to be plausible deployment mixtures. We summarized the minimum, maximum, and span within each fold before averaging. Figure 2 additionally displays reweighting of the mean cell matrix; its extrema need not equal means of per-fold extrema. Grid scenarios and their histogram are deterministic descriptions, not observations, bootstrap draws, or a probability distribution over plausible target populations. A small Δ_mix_ alone cannot establish mixture invariance; the full matrix and span describe comparisons that this one weighted contrast can obscure. No clinical equivalence margin was specified.

**Figure 2:**
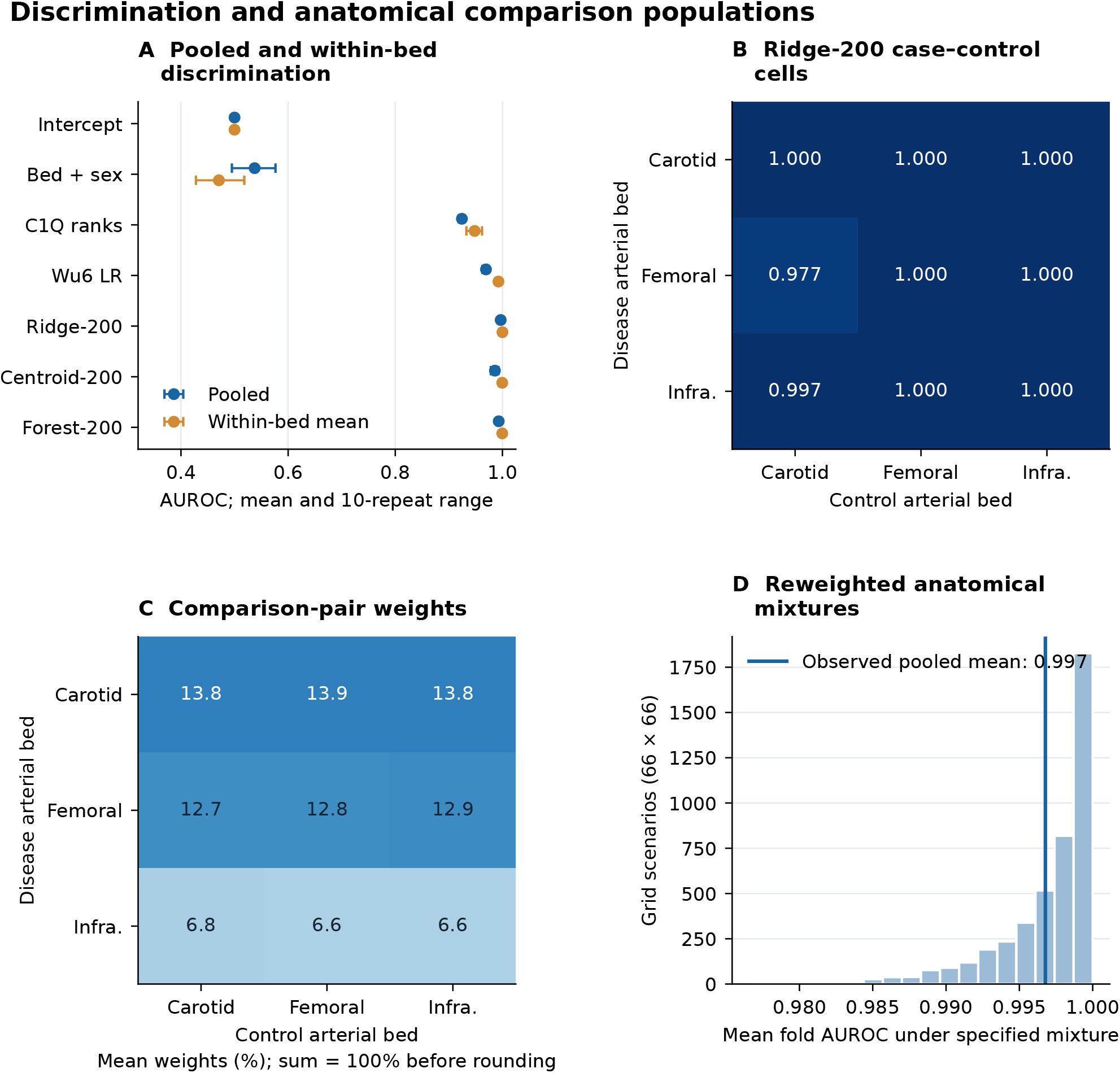
Discrimination and anatomical comparison populations. A, Fold metrics are averaged within each repetition, then summarized across ten repetitions; bars are the observed split range, not confidence intervals. B, Each cell compares case and control scores generated by the same held-out-fold model; cells are averaged over 30 folds. C, Mean fractions of all case–control pairs; within each fold the pair-weighted sum of cell AUROCs exactly equals pooled AUROC. D, The two 0.1-step three-bed simplexes each contain 66 mixtures, giving 4,356 deterministic reweighting scenarios. This histogram is the distribution of scenario values from the mean cell matrix, not a sampling distribution or observed population. It differs from the protocol’s mean per-fold extrema, reported numerically in results. C1Q and Wu6 are retrospective literature controls with prior cohort exposure, not independent biomarker discovery validation.

### 2.6 Validation, uncertainty, and sensitivity analyses

Mixed-bed evaluation used ten repetitions of three-fold splitting, stratified by the six bed-by-status strata, with seeds 20261025 + *r* for repetition *r*. Complete-label groups could not cross folds. Fold statistics were first averaged within a repetition and then across repetitions. Reported repetition ranges describe split sensitivity, not confidence intervals or 30 independent studies. The decomposition never compared scores from independently fitted folds on an assumed common scale. A separately labeled descriptive output averages each label group’s ten saved held-out scores within each model and calculates its pooled AUROC; it is neither the primary fold-based statistic nor an independent validation. Group-aware evaluation follows the general need to respect structure in predictive data [15].

The five expression-based specifications were evaluated in each of the three leave-one-bed-out tasks and in both external cohorts; the prevalence and bed/sex controls were restricted to mixed-bed evaluation. In each anatomical holdout, the four trainable specifications used only the other two beds, while the fixed C1Q mean-rank score was applied directly. The primary ridge model additionally underwent six directional single-source-bed transfers and control-proxy exclusion analyses. The latter excluded source controls sharing a numeric control suffix and sex with target controls; this proxy was not treated as authenticated linkage. For external evaluation, the four trainable specifications were fitted once to all 95 source label groups and applied unchanged to both targets, alongside the fixed C1Q score.

In GSE43292, the principal external statistic was the fraction of patients whose plaque score exceeded their intact-tissue score, with half credit for ties. It differs from marginal AUROC, which compares every positive tissue with every negative tissue. Patient clustering was retained when resampling the latter, consistent with the need to distinguish clustered from independent ROC observations [16]. We used 2,000 patient-block bootstrap draws for conditional intervals and an exact binomial interval for pair concordance when there were no ties. GSE28829 intervals used 2,000 class-stratified specimen bootstrap draws; unresolved donor dependence was not incorporated.

Additional metrics were average precision, sensitivity, specificity, balanced accuracy at the unchanged score threshold 0.5, and Brier score for probability-producing models. The threshold was a technical operating point, not an established clinical decision rule. Brier scores described the selected tissue samples; no population-risk calibration or clinical net benefit was inferred. Bootstrap intervals were conditional on the fitted source model and excluded training, feature-selection, annotation, and preprocessing uncertainty. At empirical boundaries, fixed-score resampling can give degenerate intervals: sensitivity [0, 0] when every observed positive scores below 0.5, specificity [1, 1] when every observed negative scores below it, and AUROC [1, 1] under perfect observed separation. These intervals reflect the support of the observed scores and do not establish population certainty.

Eight prespecified sensitivity specifications changed feature count to 50 or 500, ridge *C* to 0.01 or 1, representation to native expression, or identity handling to exclusion of all ambiguous records, all 104 arrays with complete-label grouping in folds, or possible-shared-control proxy grouping. Negative controls reran selection, scaling, and fitting for 99 within-bed label permutations and 99 unrestricted permutations. They diagnosed pipeline behavior under different label nulls; they were not tests of anatomical causation. Thirty stratified 80% subsets, using within-stratum flooring, each contained 75 label groups and reran the primary pipeline. Their ranges provided separate fitting-stability evidence, not confidence intervals.

### 2.7 Post-result exploratory extensions

After inspecting the original results, we recorded an extension on 26 October 2026 to examine representation specificity, feature stability, score geometry, source-only threshold selection, and individual failures. These analyses were not part of the pre-fit primary plan. A preliminary threshold based on ten-repetition averaged source scores had already been inspected and is disclosed in the extension record; it is distinct from the first-repetition recipe below. The original model, primary estimand, and external predictions were retained.

We drew 20 sets of 200 genes from the 15,235-gene vocabulary without consulting outcomes, using seed 20261026. The same sets were used throughout the original 30 folds and in full-source external fits. A disjoint control selected training-set feature ranks 201–400, excluding that fit’s top 200. All controls used the original scaling, ridge penalty, class weighting, and splits. Panel-level fold means and their distribution describe variation across representations, not independent cohorts or confidence intervals. No externally favorable panel was selected for promotion. We also replayed the 30 original primary fits to recover membership and coefficients, requiring agreement with their saved held-out scores. Pairwise Jaccard overlap and selection frequency were descriptive; we did not train a new “stable” signature using all-fold selections.

Ranking metrics for the new logistic controls used unrounded linear logits. Independent checking found that one random panel returned probabilities rounded to 1 for every GSE43292 tissue, creating artificial ranking ties. A dated correction applied logit-based AUROC, average precision, and pair ordering uniformly to all new controls. Probabilities remained the inputs to threshold and Brier diagnostics. The pre-correction outputs were retained; this numerical correction neither changed the random panels nor selected a model from target outcomes.

For the unchanged full-source primary model, we decomposed each logit into its intercept and the sum of coefficient-weighted, source-standardized feature values. Source predictions in this display were in-sample references, not validation estimates. We summarized class-specific contributions and the fraction of selected values with absolute source-standardized *z >* 3, a descriptive displacement measure rather than a validated out-of-distribution detector. Named-feature displays used the largest absolute full-source coefficients, without target-outcome selection. Exact paired logit differences additionally cancel the intercept and source center; their attribution describes the classifier, not a causal mechanism.

Two technical thresholds were learned from source cross-fitted scores: maximum Youden *J* and the largest candidate attaining source sensitivity at least 0.90. Candidates were adjacent distinct-score midpoints plus 0 and 1. Youden ties within 10^−12^ were resolved by proximity to 0.5, then the smaller threshold. Full-source thresholds used one held-out score per group from the original first three-fold repetition and were applied to the original frozen external probabilities. Scores from three separately fitted fold models were pooled only for threshold learning; this procedure remains sensitive to fold-to-fold scale and does not replace the within-fold AUROC estimand. For each anatomical holdout, a separate inner three-fold cross-fit was confined to the other two beds; the outer fit then scored the excluded bed. Source sensitivity constraints are optimization criteria, not clinical targets or unbiased source performance estimates. Target threshold-response curves were descriptive, and no target-best rule was adopted.

Finally, we displayed ordering signs for all 32 patient pairs under five expression rules. Fixed-score deletion of both tissues from each patient assessed influence on marginal AUROC and pair concordance; deletion of individual GSE28829 specimens assessed a separate histological contrast. These operations did not refit a model and are not cross-validation or confidence intervals. Failures selected for detailed contribution displays were explicitly retrospective. Supplementary Figures S1 and S2 show additional fold-level cell variation and repeated source-score behavior from the original saved results.

### 2.8 Reproducibility and software assistance

The analysis record retains acquisition manifests, annotation mappings, grouped-label membership, split assignments, predictions, frozen coefficients, logs, protocol versions, and plotting code. Saved prediction checks independently recompute the decomposition and external paired statistics. Applicable TRIPOD+AI reporting principles informed the explicit reporting of populations, comparators, preprocessing, validation, and uncertainty, without treating a reporting checklist as evidence of clinical readiness [17].

Codex assisted with literature retrieval, specification review, analysis-code development and execution, figure generation from computed results, and manuscript preparation. Quantitative results were produced by numerical scripts operating on deposited measurements; language-model-generated observations were not substituted for data. AI assistance itself was not an evaluated intervention, and no autonomous-agent performance, reinforcement-learning improvement, or human–AI collaboration experiment was conducted.

## 3 Results

### 3.1 Source structure and identity uncertainty

The 104 deposited source arrays produced 95 complete-label groups under the primary convention (Figure 1). Nine repeated-label pairs had rank-profile correlations of 0.9835–0.9924. This similarity was consistent with closely related profiles but could not identify the duplication mechanism or authenticate a donor. Sex composition also differed across disease strata: male proportions were 74% in carotid, 96% in femoral, and 77% in infrapopliteal disease groups, compared with 80% in each control stratum. These are properties of the recorded groups, not estimates of disease prevalence.

### 3.2 The primary model did not show pooling inflation

Primary mean held-out-fold pooled AUROC was 0.9968, with ten-repetition means ranging from 0.9939 to 0.9985. Mean within-bed AUROC was 1.0000, giving Δ_mix_ = −0.0032; repetition values ranged from −0.0061 to −0.0015. The prespecified contrast therefore showed pooled discrimination slightly below within-bed discrimination, with no positive pooling inflation in these evaluated samples (Figure 2). All three diagonal AUROCs were 1 in every primary-model fold, so *A*_pooled_ *≤* 1 mathematically constrained Δ_mix_ = *A*_pooled_ −1 *≤* 0. Its sign is consequently not independent evidence of population-level anatomical invariance. The mean cross-bed share of comparison pairs was 66.8%; the observed pooled AUROC remained 0.0032 below the diagonal mean.

The nine-cell mean matrix localized the small shortfall: femoral cases compared with carotid controls had mean AUROC 0.9766, and infrapopliteal cases compared with carotid controls had 0.9967; the other cells were 1.0000. Mean per-fold mixture extrema were 0.9743 and 1.0000, giving a mean span of 0.0257. The extrema of the mean matrix were instead 0.9766 and 1.0000, and the smallest individual fold cell was 0.8750. Individual primary cells contained only 12–36 case–control pairs sharing observations. These results support strong observed within-bed separation, not a formal claim of anatomical equivalence. The separate pooled AUROC after averaging each label group’s ten held-out primary-model scores was 0.996923; the supplement distinguishes this descriptive value from the primary equal-fold mean of 0.996775.

The estimand distinction was more apparent for some simpler controls. Bed/sex logistic regression had pooled AUROC 0.5375 and within-bed mean 0.4709. C1Q scores had corresponding values 0.9244 and 0.9482; the six-gene model had 0.9693 and 0.9926. Centroid and forest pooled AUROCs were 0.9860 and 0.9931, respectively, with within-bed means near 1.0000. The prevalence intercept remained at 0.5000. Primary balanced accuracy was 0.9549 and Brier score 0.0358 in the source evaluation.

### 3.3 Anatomical ranking transported more consistently than thresholds

For the primary model, leave-one-bed-out AUROCs were 1.000 for carotid, 0.996 for femoral, and 1.000 for infrapopliteal targets. Corresponding balanced accuracies were 0.850, 0.820, and 0.962 (Figure 3). The six directional single-bed transfers had AUROCs from 0.980 to 1.000, but balanced accuracies ranged from 0.500 to 0.962. In particular, carotid-to-infrapopliteal transfer achieved AUROC 1.000 while assigning every target score below 0.5.

**Figure 3:**
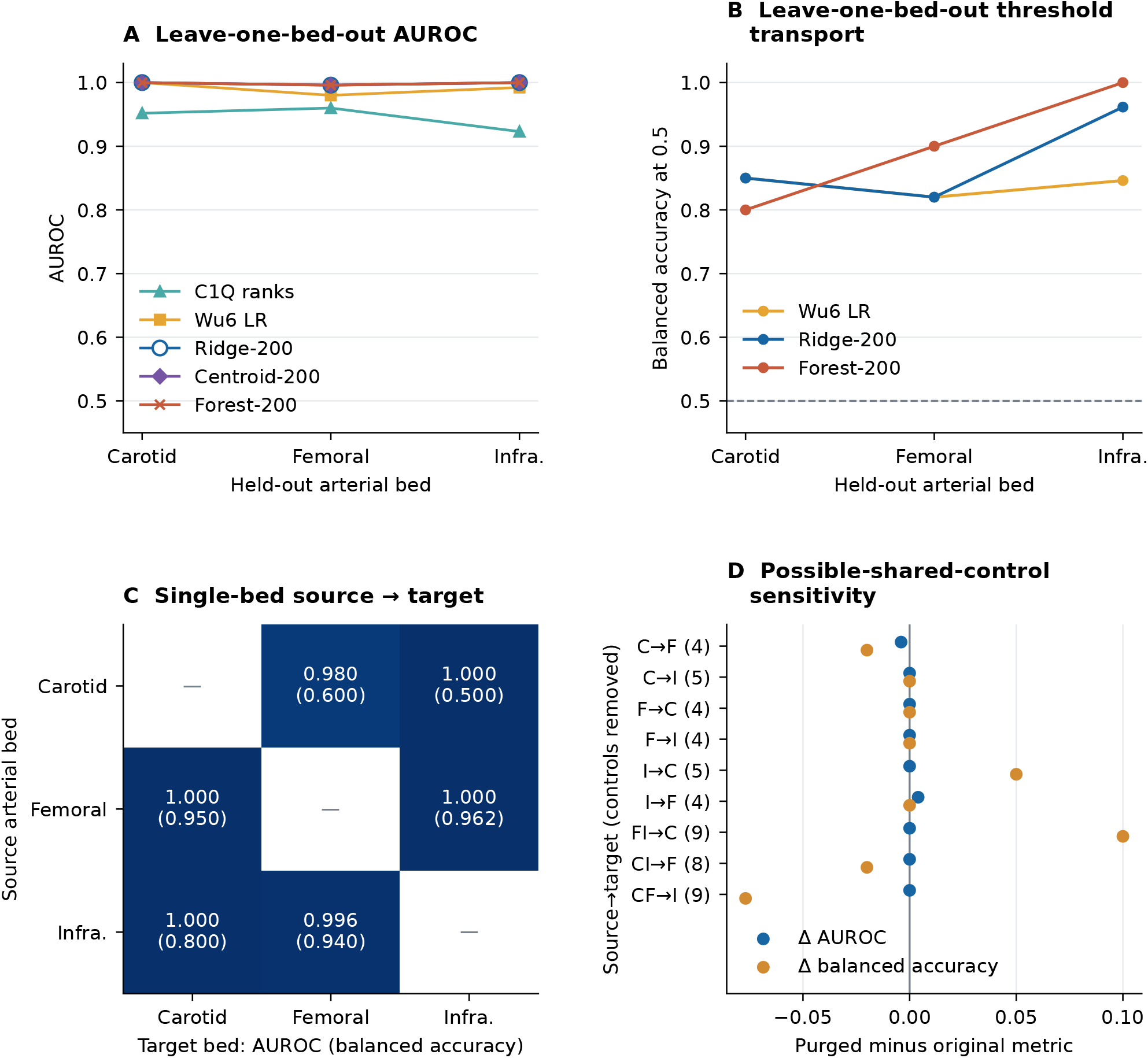
Transport to held-out arterial beds. A–B, For each trainable expression specification, a separate model is fitted on two source beds and evaluated in the remaining bed; the fixed C1Q score is applied without fitting. C, Ridge-200 is trained in a single bed for each of six directed source–target comparisons; cells show AUROC and balanced accuracy at the fixed 0.5 threshold. D, Changes after excluding source controls with a numeric suffix/sex proxy matching target controls; labels give the number removed. This deliberately conservative sensitivity does not authenticate shared donors. C, carotid; F, femoral; I, infrapopliteal. C1Q and centroid scores are not probabilities and are excluded from fixed-probability-threshold diagnostics. No uncertainty bars are inferred from these three or six dependent tasks.

Excluding possible shared controls removed eight or nine source controls in the leave-one-bed-out scenarios. The primary AUROCs remained 1.000, 0.996, and 1.000; balanced accuracies changed to 0.950, 0.800, and 0.885. Directional exclusions removed four or five controls. These analyses show sensitivity of operating characteristics to source composition while leaving the uncertainty about donor linkage unresolved.

### 3.4 Paired lesion ranking and the unchanged technical threshold diverged

In GSE43292, the frozen primary model achieved marginal AUROC 0.877 (conditional 95% interval 0.792–0.947) and average precision 0.870. Plaque scored above intact tissue in 29 of 32 patients, with no ties: concordance 90.6% (exact 95% interval 75.0–98.0%). The mean paired probability-score difference was 0.00728 (patient-bootstrap interval 0.00187–0.01665). Three patients had reversed ordering (Figure 4).

**Figure 4:**
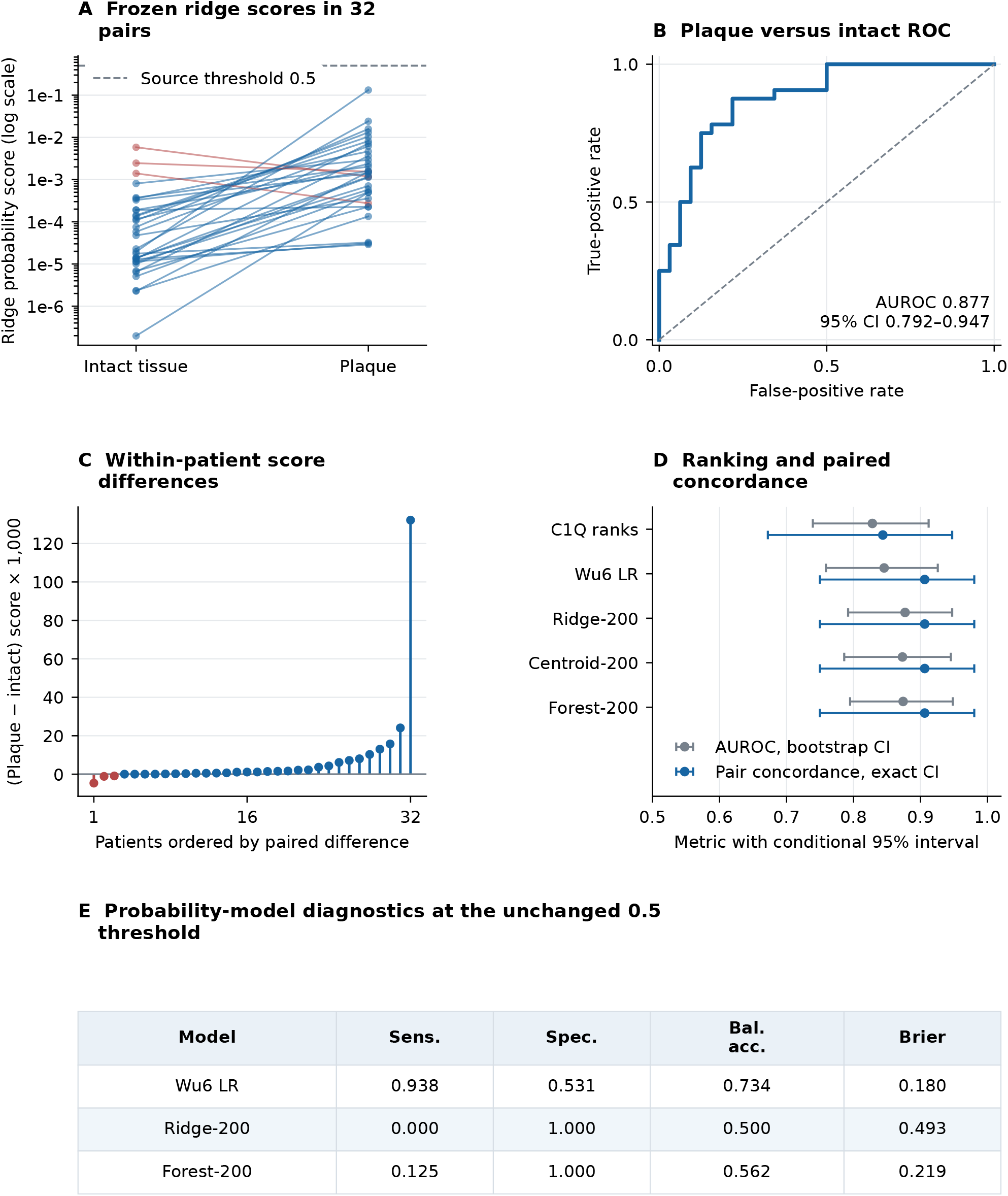
Paired external tissue discrimination and threshold failure. Frozen source models are applied to GSE43292: 64 carotid tissues from 32 paired hypertensive endarterectomy patients, plaque versus macroscopically intact Stary I/II tissue. A, Lines join tissues from the same patient; red indicates a lower plaque score. The log axis displays the original positive scores and the unchanged threshold. B, The primary-model AUROC interval uses 2,000 patient-block bootstrap draws. C, Raw paired differences, scaled only by 1,000 for legibility. D, AUROC intervals use the patient bootstrap; pair-concordance intervals are exact binomial intervals because there are no ties. These are conditional on the frozen model. E, All primary-model scores fall below 0.5 despite ranking discrimination. Brier scores describe selected tissue samples and are not clinical-risk calibration. Nonprobability scores have no threshold/Brier entries.

All 64 primary scores were below 0.5: 0/32 plaque tissues were classified positive and 32/32 intact tissues negative. Sensitivity was therefore zero, specificity 1.000, balanced accuracy 0.500, and Brier score 0.4926. The degenerate fixed-score bootstrap intervals at these boundaries do not establish future sensitivity or specificity with certainty. Other models illustrated that ranking and operating-point behavior were separable. C1Q marginal AUROC was 0.828; six-gene, centroid, and forest AUROCs were 0.846, 0.873, and 0.874. The six-gene model retained sensitivity 0.938 with specificity 0.531, whereas forest sensitivity was 0.125 with specificity 1.000. No threshold metrics were assigned to C1Q or centroid scores.

### 3.5 Histological discrimination did not establish progression prediction

In GSE28829, primary advanced-versus-early lesion AUROC was 0.986 (conditional specimen-bootstrap interval 0.942–1.000), with average precision 0.989. Unknown donor dependence was not incorporated in this interval. Again, every score was below 0.5: 0/16 advanced specimens were classified positive and 13/13 early specimens negative, producing sensitivity zero, specificity 1.000, balanced accuracy 0.500, and Brier score 0.4182 (Figure 5). C1Q, six-gene, centroid, and forest AUROCs were 0.952, 0.971, 1.000, and 0.846. The centroid’s perfect observed ordering and the boundary threshold metrics have degenerate empirical bootstrap intervals; these do not establish zero uncertainty outside these specimens. This contrast contains no longitudinal observations from which to infer disease progression.

**Figure 5:**
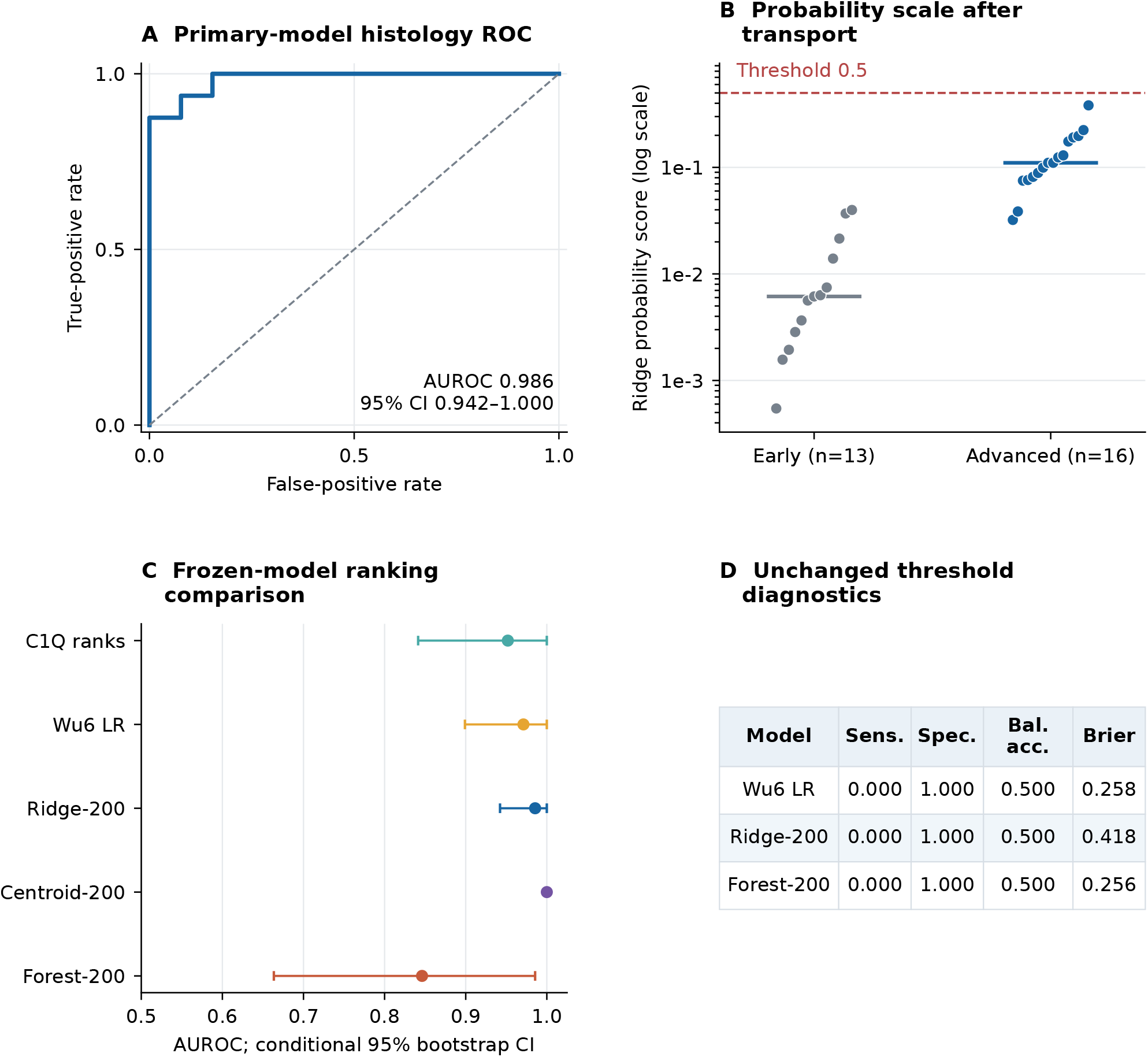
External histology contrast: advanced versus early lesions. GSE28829 contains 29 autopsy carotid specimens (13 early, 16 advanced lesions); deposited metadata do not establish an independent donor count. This is a changed histological contrast, not healthy-control discrimination. A, Primary frozen-model ROC. B, Original probability scores on a log axis; horizontal marks are within-class medians. C, Conditional percentile intervals from 2,000 class-stratified specimen bootstraps; unresolved donor dependence is not captured. A perfect observed AUROC can have a degenerate fixed-score empirical interval, which does not establish zero uncertainty. D, Threshold diagnostics without target recalibration; Brier values do not estimate clinical-risk calibration. No best external model was selected for the primary conclusion.

### 3.6 Specification and pipeline checks

Mean pooled AUROC across the eight alternative specifications ranged from 0.9937 to 0.9981; all mean composition contrasts remained nonpositive (Figure 6). Excluding every ambiguous-label record left 86 arrays and mean pooled AUROC 0.9937. Retaining all 104 arrays with complete-label groups held together yielded 0.9958; conservative control-proxy grouping yielded 0.9957. Native-expression AUROC was 0.9972.

**Figure 6:**
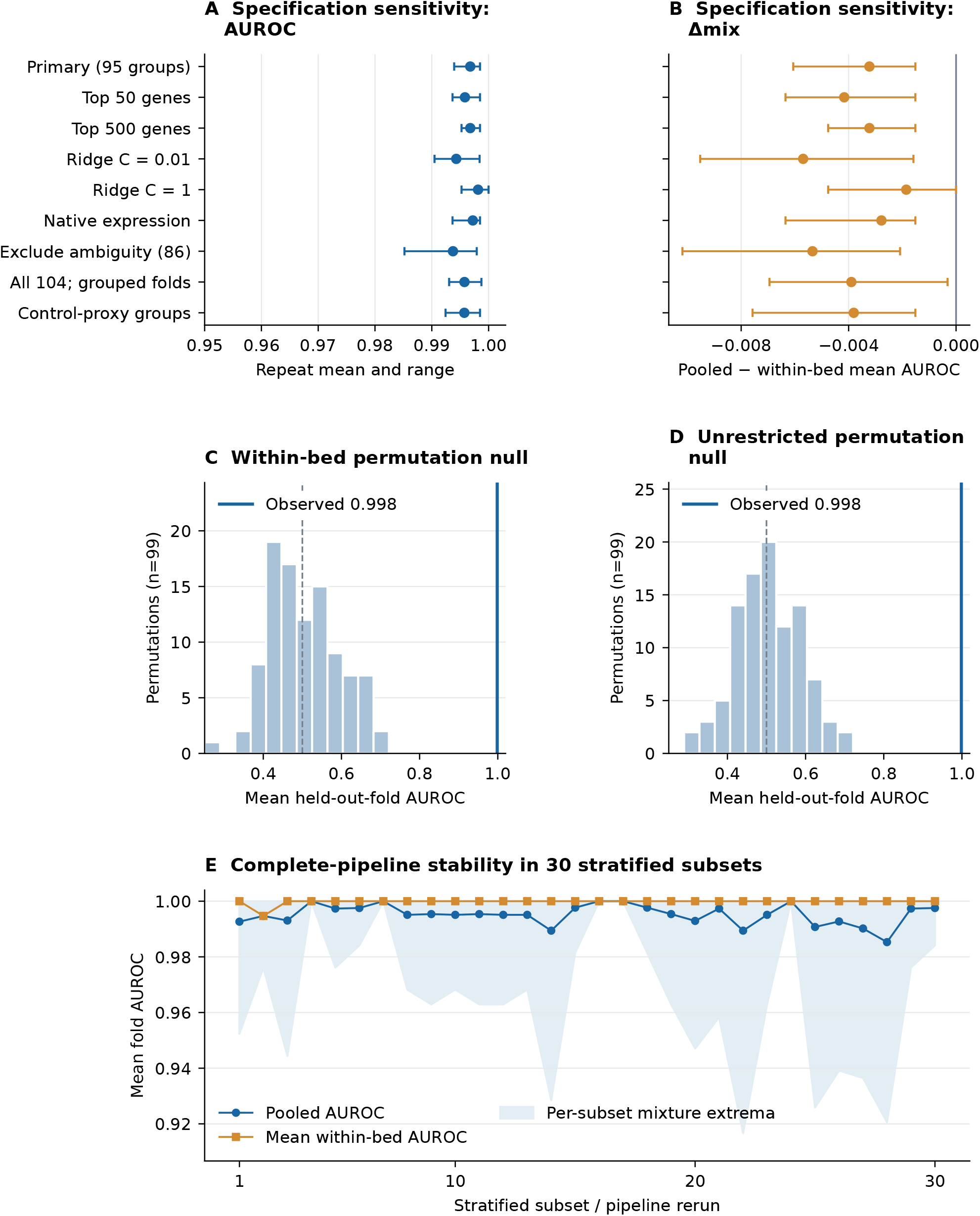
Specification, negative-control and subset stability analyses. A–B, Each point is the mean of ten repetition means, each formed from three held-out folds; bars show the ten-repeat range, not confidence intervals. Each alternative was specified before its dependent analysis. Identity conventions include excluding all repeated-label records (86 arrays), retaining 104 arrays with full-label groups kept in one fold, and conservatively grouping possible shared-control suffix/sex proxies. C–D, Ninety-nine label permutations per scheme rerun feature selection, scaling and fitting; the observed line is the first fixed three-fold scheme. These are pipeline-null diagnostics, not tests of anatomical causation or independent replications. E, Thirty stratified 80% subsets (flooring within six strata gives 75 label groups) rerun the complete primary pipeline. Shading shows mean per-fold mixture extrema within each subset, not a confidence band.

Within-bed and unrestricted permutation means were 0.5027 and 0.5057, with maxima 0.7154 and 0.6853, respectively. Both maxima were below the observed first-repetition mean AUROC of 0.9985. Across 30 complete-pipeline subset runs, pooled AUROC ranged from 0.9853 to 1.0000 and within-bed mean AUROC from 0.9947 to 1.0000. These diagnostics support reproducible source ranking under the tested perturbations; they do not remove original-study selection, processing, or identity limitations.

### 3.7 Random panels preserved source discrimination but differed after transport

The exploratory representation controls exposed information not apparent from source model comparisons alone (Figure 7). The 20 random panels had mean-fold source AUROCs of 0.9932–0.9992 (median 0.9961), spanning the primary value 0.9968. The disjoint next-ranked panel yielded 0.9955. These results show that high source discrimination was not unique to the particular top-200 panel in these data; random feature choice is not a label-permutation null, because the selected genes can still contain real or confounded class-associated information.

**Figure 7:**
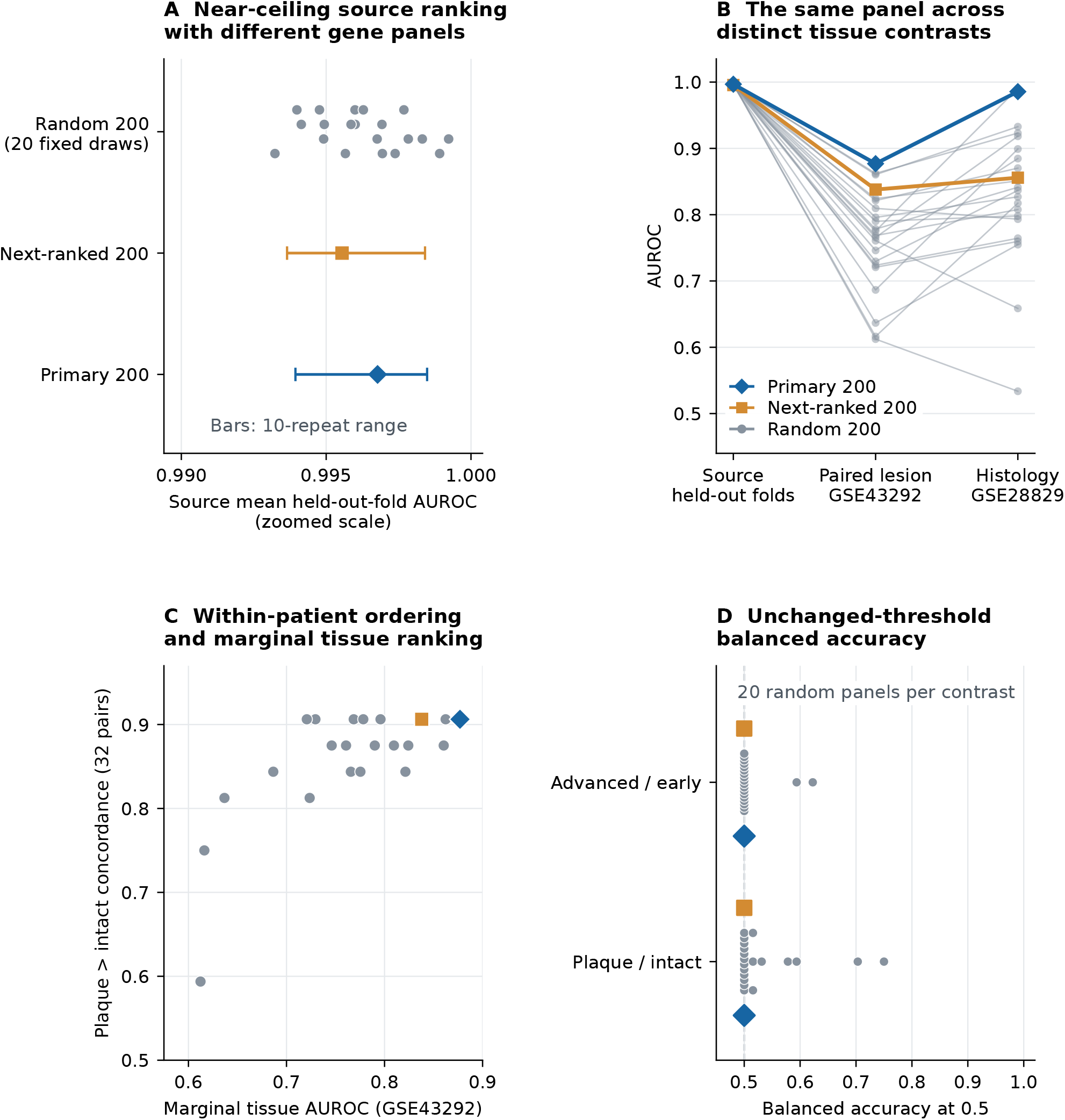
Gene-panel controls separate source ranking from external transport. Exploratory analyses added after inspection of the original results. Twenty 200-gene panels were drawn once without using outcomes (seed 20261026); all draws are retained. The next-ranked control selects training ranks 201-400 after excluding each training set’s top 200 genes. All trainable rules use source-only scaling and the same fixed ridge specification. New-control ranking metrics use raw linear logits to avoid artificial ties from floating-point sigmoid saturation; probabilities remain the inputs to threshold and Brier diagnostics. A, Each random point is its panel’s mean across the original 30 held-out folds. Bars for primary and next-ranked panels show the ten-repetition range, not a confidence interval; the horizontal axis is zoomed. B, Each grey line follows the same random panel; source cross-validation and external full-source fits are different evaluations, and the three contrasts are not interchangeable endpoints. C, For each panel, marginal tissue AUROC and within-patient plaque > intact concordance answer different comparison questions in GSE43292; half credit is assigned to ties. No external winner is selected. D, Tied random-panel values are stacked only for visibility. The unchanged 0.5 rule is a technical classifier diagnostic, not a validated clinical decision threshold. Panel draws and overlapping folds are not independent cohorts; variations across panels are not confidence intervals.

External random-panel AUROCs ranged from 0.6123 to 0.8623 in GSE43292 and from 0.5337 to 0.9904 in GSE28829. Their medians were 0.7671 and 0.8317. The next-ranked panel yielded 0.8379 and 0.8558, respectively. Thus, similar near-ceiling source performance concealed much wider variation under changed tissue comparisons. These descriptive draws do not establish an optimized or universally superior signature. For the saturated random panel, raw-logit AUROC was 0.7207 and pair concordance 29/32 despite all 64 probabilities rounding to 1; this illustrates why numerical score precision must be distinguished from biological ranking failure.

### 3.8 Stable ranking coexisted with variable selected-gene membership

Across the 30 recovered primary fits, 619 different genes were selected at least once; 14,616 were never selected, 103 appeared in at least 24 fits, and 45 appeared in all 30 (Figure 8). The 435 pairwise feature-set Jaccard values ranged from 0.3468 to 0.7167, with mean 0.4850. These overlapping fits are dependent, so the distribution is not an uncertainty interval. Agreement between recovered and original held-out predictions verified that this display describes the original fitting procedure. Membership and coefficient direction of source-selected named features were shown without reusing their stability to build a new classifier. High source ranking therefore did not imply a uniquely identified molecular signature.

**Figure 8:**
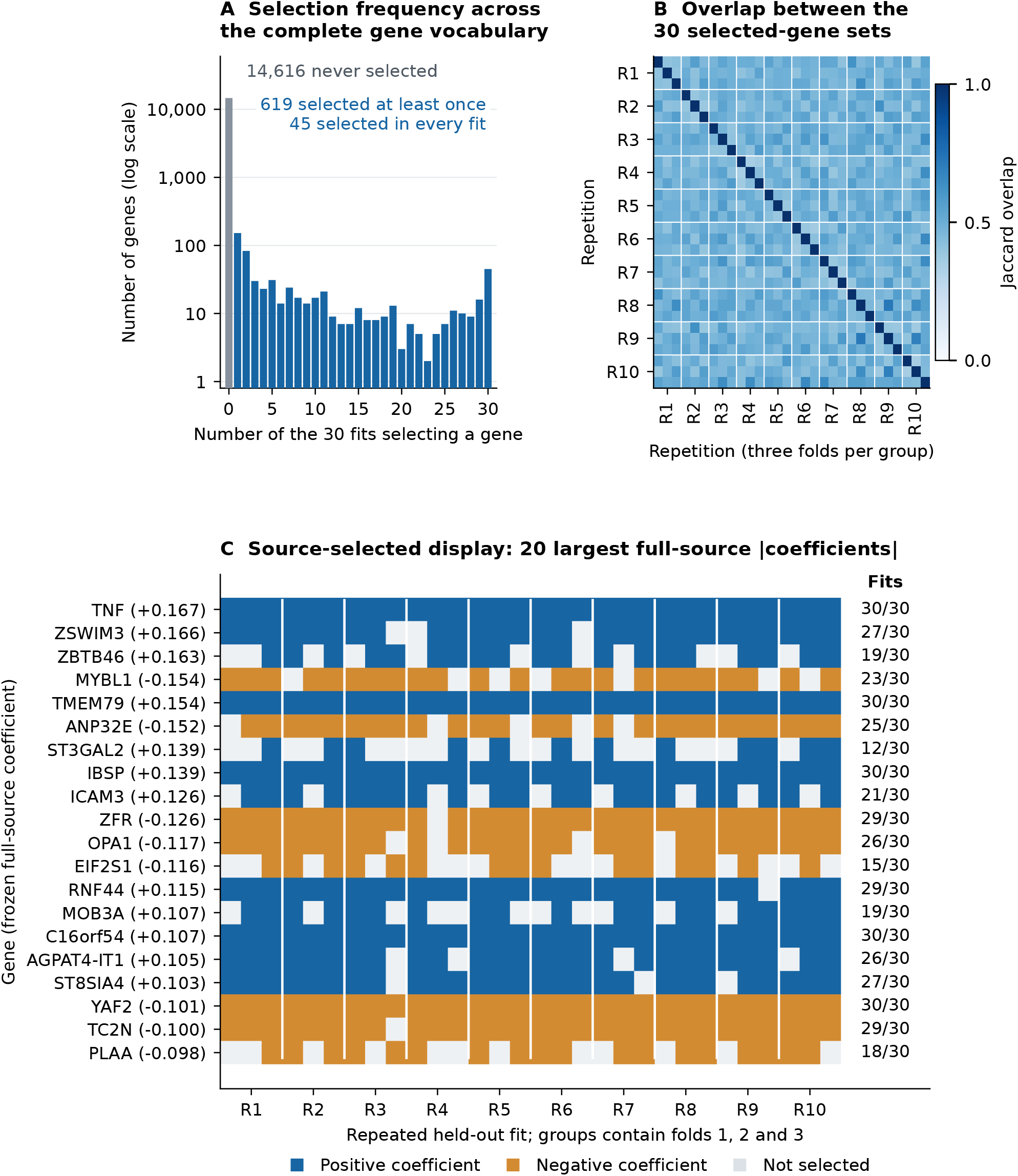
Feature membership varies across fits despite stable source ranking. Exploratory feature-recovery analysis after the original results. The 30 primary source fits were replayed to recover selected genes and coefficients; held-out predictions were checked against the saved original predictions before interpretation. A, All 15,235 candidate genes are included; the count axis is logarithmic. Each fit selects 200 genes, with 619 distinct genes selected across fits, including 45 selected in every fit. B, Jaccard overlap is intersection divided by union. Three adjacent rows/columns constitute the three folds of a repetition; diagonal values equal one. Off-diagonal comparisons reuse training observations and are descriptive, not independent replications or confidence intervals. C, Named genes are the 20 largest absolute coefficients of the unchanged full-source primary model, selected without using external outcomes; full-source coefficients appear beside names. Each tile shows absence or coefficient sign in one recovered fold fit, with selection counts at right. These high-weight genes range from intermittent to persistent membership, while A-B retain the full vocabulary. No stable signature is refitted or revalidated using these summaries. Correlated genes and fold-specific standardization prevent interpreting coefficient magnitude or selection frequency as causal biological importance.

### 3.9 Frozen-score accounting localized scale displacement without identifying its cause

The same full-source model placed mean logits at 0.2808 in the source in-sample reference, −8.3072 in GSE43292, and −3.4185 in GSE28829 (Figure 9). Because overall means depend on class composition, the figure also separates every cohort’s two classes. The intercept plus signed feature contributions exactly reconstructed these logits. Across the selected 200 genes, the proportion of gene–sample values with absolute source-standardized *z >* 3 was 0.12% in the source, 23.21% in GSE43292, and 21.71% in GSE28829. These are fractions of correlated feature values, not percentages of independent patients or clinical error rates. They document displacement relative to source scaling while leaving platform, procurement, normalization, and comparator changes inseparable.

**Figure 9:**
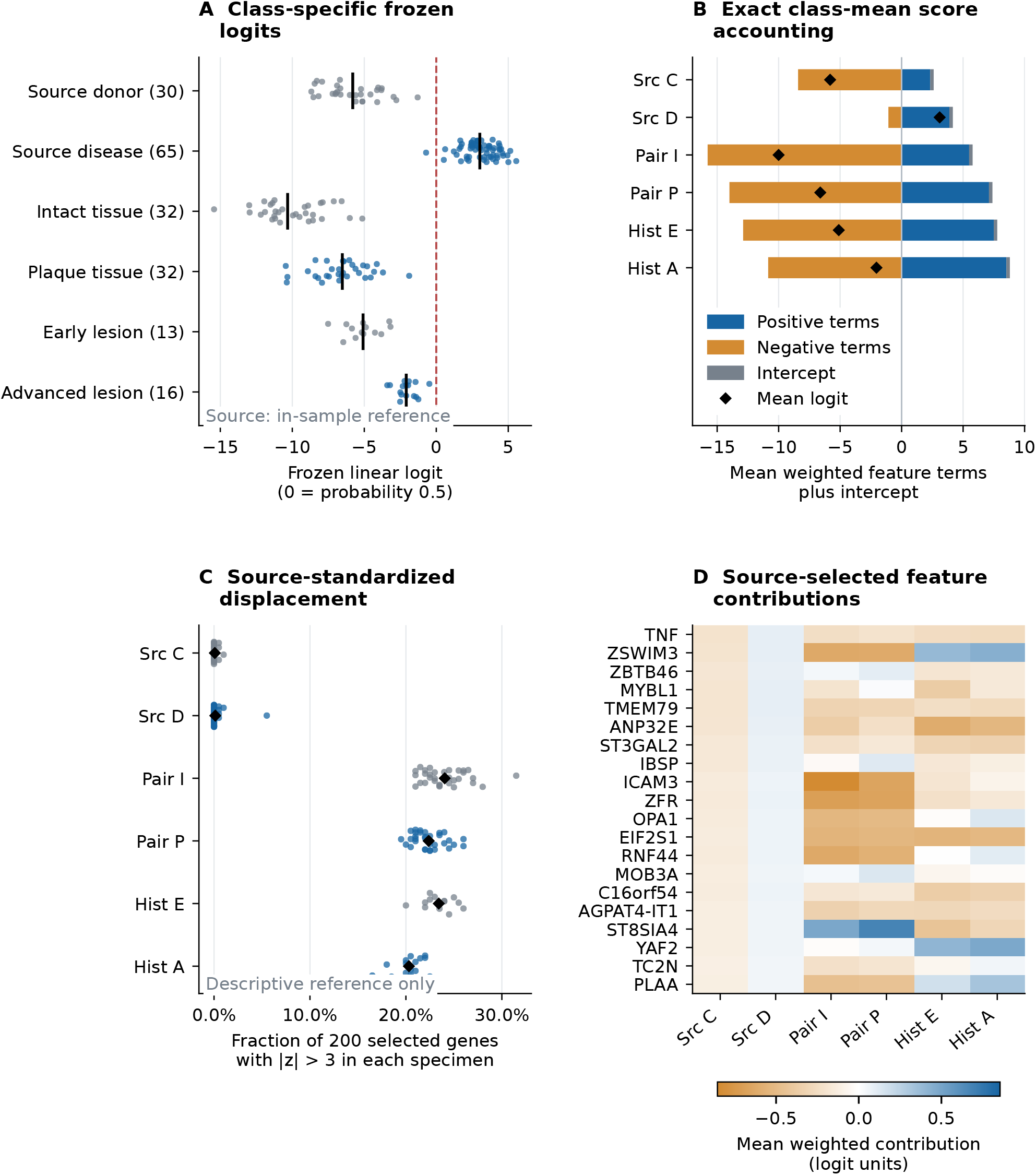
Frozen score geometry under changed tissue comparisons. Post-result exploratory diagnostics. A, Each point is one specimen or source label group scored with the same original full-source ridge model; source values are in-sample descriptive references, not held-out performance. Black marks denote class medians. B, Positive and negative sums of the 200 class-mean coefficient-weighted standardized features, plus the intercept, exactly reconstruct the black mean-logit markers. Classes are shown separately because overall cohort means also depend on class composition. C, Each point gives the fraction of 200 selected feature values outside absolute source-standardized z = 3; diamonds show class means. This descriptive reference does not establish an out-of-distribution detector or patient error rate. D, Genes are displayed in descending absolute full-source coefficient order, fixed without target outcomes; the heatmap shows only these 20 genes, whereas B uses all 200. Src C/D, source donor control/surgical disease; Pair I/P, paired intact/plaque tissues; Hist E/A, early/advanced histology. These are distinct endpoints. The accounting does not identify causal genes or isolate platform, procurement, normalization or comparator effects. GSE28829 donor independence is unresolved.

### 3.10 Source-only threshold learning did not restore external sensitivity

The first-repetition source cross-fit produced thresholds of 0.6173 for maximum Youden *J* and 0.6922 for the source sensitivity constraint. Both remained above every primary-model probability in both external datasets, yielding zero sensitivity and balanced accuracy 0.500, as did 0.5 (Figure 10). Nested anatomical evaluations showed that threshold effects were context-dependent: maximum-*J* balanced accuracy changed from 0.850 to 0.800 in carotid, from 0.820 to 0.880 in femoral, and remained 0.962 in infrapopliteal targets. The corresponding sensitivity-constrained values were 0.850, 0.780, and 0.962. Thus, these particular source-only recipes did not solve external transport; the experiment does not establish that every possible source recipe must fail or that target recalibration would be clinically valid.

**Figure 10:**
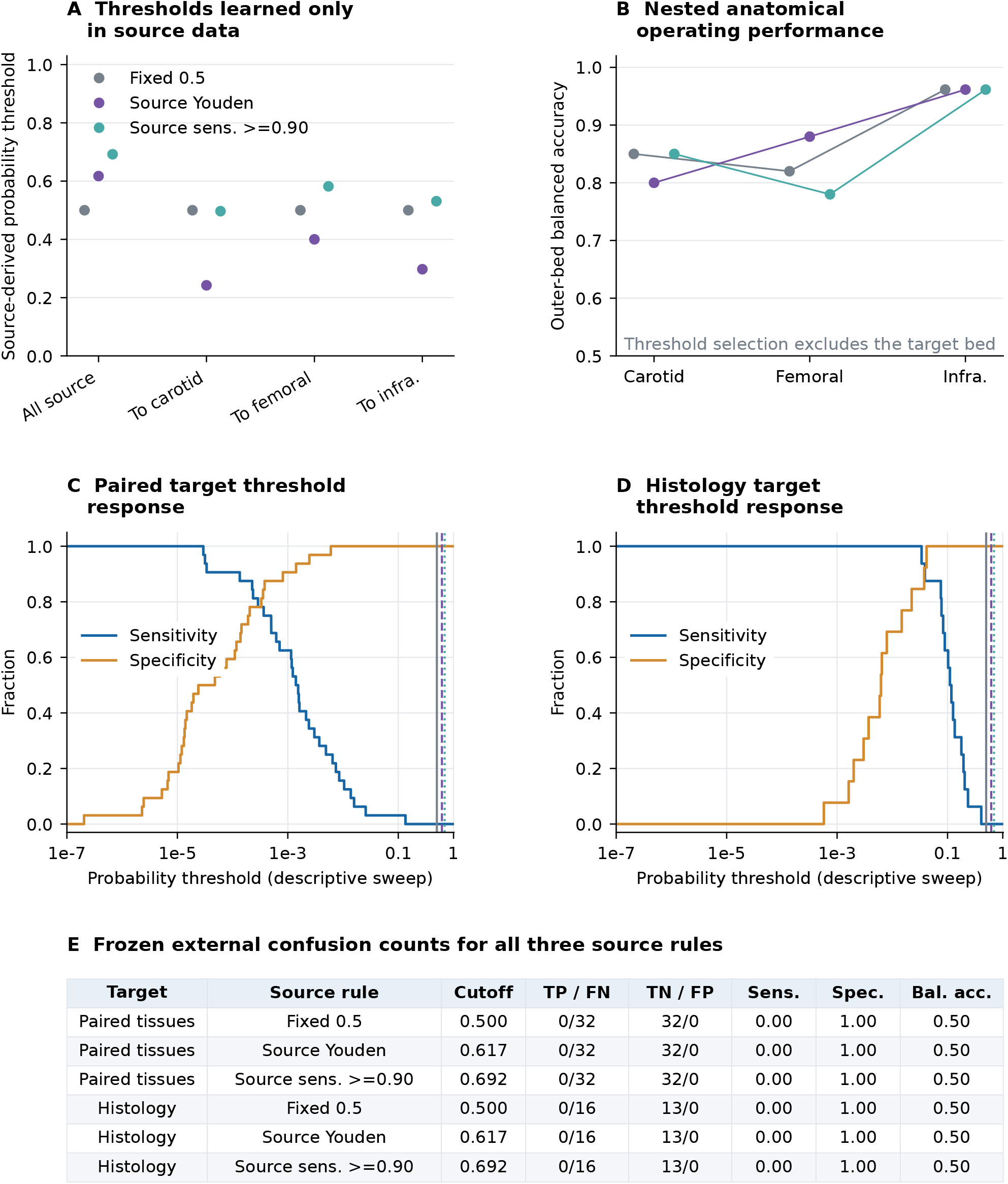
Tested source thresholds leave external sensitivity at zero. Post-result exploratory diagnostics; no target threshold optimization or recalibration. A, Source threshold rules are fixed 0.5, maximum Youden J and the largest candidate meeting source sensitivity at least 0.90. Candidates are adjacent score midpoints plus endpoints 0 and 1; Youden ties choose the value closest to 0.5, then the smaller value. The 0.90 constraint is illustrative, not a clinical target. The all-source rule uses the original first three-fold repetition, one held-out score per label group. Each anatomical rule uses a new inner three-fold cross-fit confined to the other two beds. B, Each full outer-training fit is evaluated only in its excluded bed; no source fitting or threshold selection used that bed. C–D, Outcome-dependent sweeps describe the already observed target scores; vertical lines mark the three unchanged source rules, whose values are shown in A/E. No target-best cutoff is promoted. E, TP/FN and TN/FP are tissue counts, not numbers of independent patients across cohorts. Source maxima/constraints are optimization quantities, not unbiased source performance estimates. The frozen external model and original primary 0.5 results are unchanged. Previously inspected ten-repeat averaged-source threshold calculations are separately disclosed and are not this first-repetition recipe.

### 3.11 Patient-level failures were partly shared across scoring rules

All five expression rules ranked plaque above intact tissue in 26 of 32 patients; at least one rule reversed the ordering in the remaining six (Figure 11). Primary ridge reversals involved deposited patient identifiers 12, 17, and 21. Fixed-score deletion of one known patient changed primary marginal AUROC to values between 0.8689 and 0.8959 and pair concordance to 0.9032–0.9355. Deleting one histology specimen yielded primary AUROC 0.9844–0.9949. These ranges describe sensitivity to individual records, not uncertainty bounds or additional validation. In the source repeated predictions, 87 groups had no 0.5-threshold error, five had errors in some repetitions, and three were misclassified in every repetition (Supplementary Figure S2). Exact contribution displays for the three reversed patient pairs expose the fitted score arithmetic without attributing failure to an unmeasured clinical characteristic.

**Figure 11:**
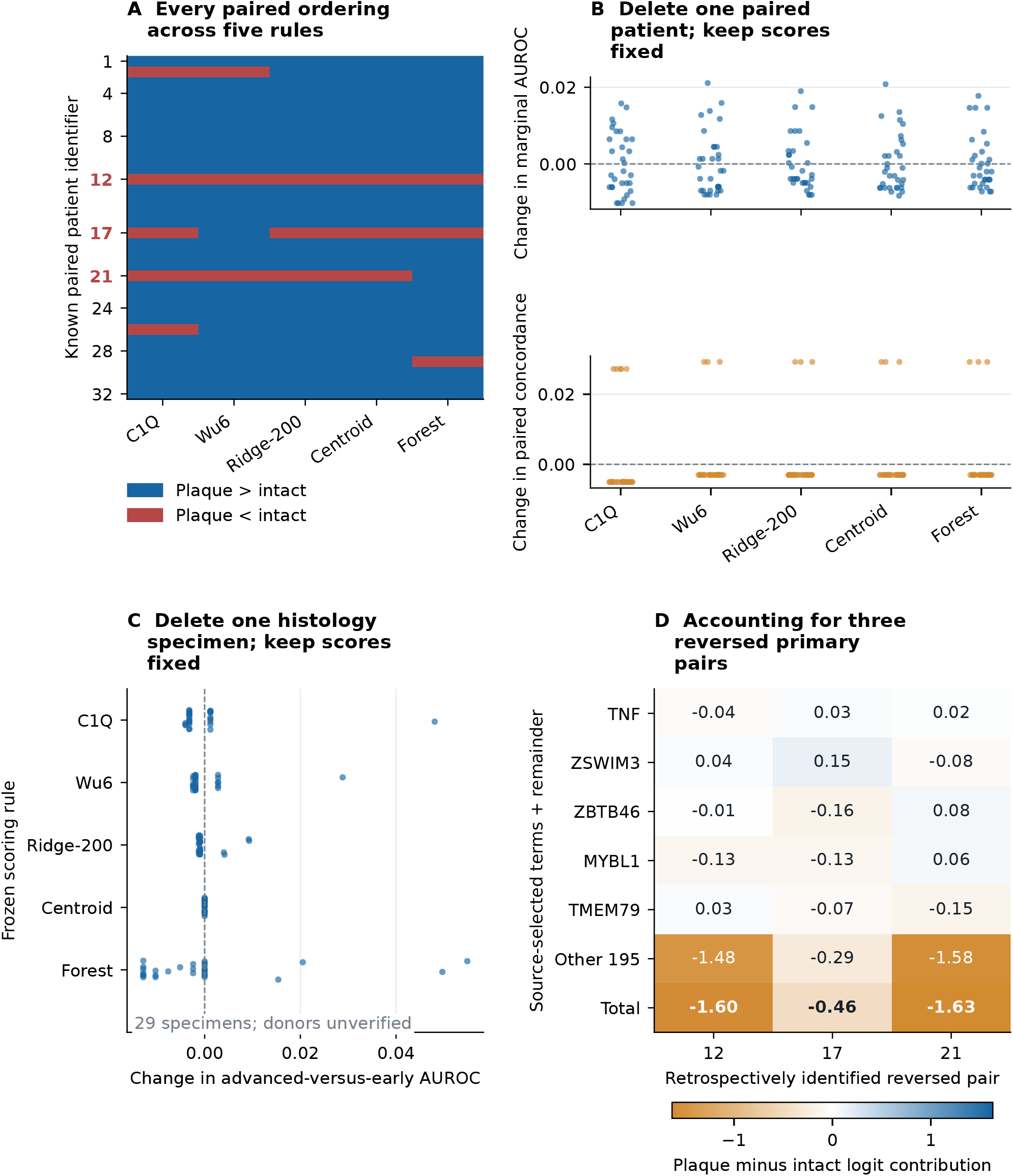
Individual failures and fixed-score influence boundaries. Post-result exploratory failure diagnostics. A, All 32 known patient pairs and five fixed expression rules are shown in numerical patient order; a subset of axis identifiers is labeled for legibility. Blue means plaque scores above intact tissue and red means the reverse. No ties occurred. Bold red identifiers mark the three primary ridge reversals, selected retrospectively as failures; colors compare ordering signs, not raw score scales across models. B, Each point deletes both tissues from one patient while retaining every other frozen score; upper and lower displays concern marginal AUROC and within-patient concordance, respectively. These are influence descriptions, not refits, bootstrap draws or confidence intervals. C, Analogous fixed-score deletion removes one of 29 histology specimens; donor dependence remains unknown. D, Coefficient-weighted paired rank differences exactly reconstruct the plaque-minus-intact logit margin. Five named genes are the largest absolute coefficients in the full-source model, not chosen from these failed patients. The other 195 genes are summed; the intercept and source center cancel in paired differences. This algebra does not establish biological causes of failure. The three failed pairs do not represent an independently selected validation set.

## 4 Discussion

This study separated three properties often compressed into a single external AUROC: the anatomical composition of comparisons, preservation of tissue ordering, and transfer of an operating threshold. For the fixed primary model, the prespecified comparison placed pooled AUROC 0.0032 below the equally weighted within-bed mean. Leave-one-bed-out ranking also remained high. A separate limitation concerned score transport: both external contrasts retained ranking information while the unchanged 0.5 threshold classified every primary-model target tissue as negative.

The composition contrast describes how the comparisons contributed to the observed statistic. Approximately two thirds of held-out case–control pairs crossed beds, yet the nine-cell decomposition localized the small departure from perfect separation to particular off-diagonal comparisons. Because the observed diagonal mean was already 1, a positive Δ_mix_ was mathematically impossible in these primary-model folds. The nonpositive contrast therefore cannot by itself demonstrate population-level mixture invariance; the cell matrix and reweighting span supply complementary descriptive information. A high pooled result warrants an estimand audit, while near-ceiling source data limit resolution for smaller composition effects and cannot establish absence of anatomical effects in other populations, disease stages, or measurement systems.

Our analysis does not introduce a new AUROC formula or demonstrate that shared vascular signatures are novel. Earlier work has already explored both multi-bed gene discovery and C1Q-related cross-cohort discrimination [5, 6]. The contribution is the combination of an explicit empirical comparison-population decomposition, transparent identity conventions, and separately interpreted anatomical and tissue-comparator stress tests. The source results favor a strong transferable ranking signal within this selected donor-versus-surgical-tissue setting. The repeated-label alternatives help show that its magnitude did not depend entirely on one grouping convention, while stopping short of proving patient independence.

The paired external analysis adds a different biological perspective. Marginal AUROC 0.877 and 29/32 concordant patient pairs both indicate retained plaque-associated information, but they answer different questions. The first aggregates comparisons across all plaque and intact tissues; the second asks whether the lesion is ranked higher within the same person. The latter reduces some between-person differences by design, but the relatively intact sample remains tissue from a patient with established vascular disease. Neither statistic establishes diagnostic accuracy for an asymptomatic population or identifies an intervention-responsive molecular state.

The unchanged technical 0.5 boundary assigned every specimen to the negative class in one source-study arterial-bed transfer and in both changed external contrasts. The exploratory source-only threshold experiment extended this observation beyond an arbitrary single cutoff: two source-derived rules also had zero external sensitivity, while their effects differed across anatomical holdouts. These are failures to separate changed tissue labels, not failure tests of a validated clinical rule for an unchanged endpoint. A balanced, penalized classifier’s probability is not automatically an individual’s clinical risk. Target intercept adjustment or recalibration would answer a different question; neither was performed or established as clinically effective.

The random-panel and feature-membership results further constrain molecular interpretation. Near-perfect source discrimination survived outcome-independent feature sampling and replacement by a disjoint ranked panel, while external rankings and selected-gene sets varied. A broad expression contrast can therefore make a particular panel appear compelling without showing that it is necessary or biologically specific. The present controls do not distinguish a distributed disease-associated program from cellular composition, procurement, or other shared class differences. Likewise, coefficient accounting connects numerical displacement to actual prediction failures but does not establish the biological cause of that displacement. Both observations favor evaluating representations against explicit alternatives before assigning significance to named genes.

Earlier breast-cancer expression analyses provide a methodological precedent for comparing named signatures with size-matched random gene sets [18]; those results do not validate vascular biology or the present classifier.

The external differences cannot be attributed specifically to comparator definition. Platforms, source normalization, tissue handling, recruitment, sampling, and lesion spectrum changed together. Samplewise ranking removed a common within-sample scale dependence but could not remove gene-specific platform behavior or make different tissue definitions exchangeable. Strong ordering after transport is therefore evidence of an association shared across these particular contrasts, not proof of a universal vascular representation. Poor threshold transport likewise cannot identify which component of the domain change was responsible.

This distinction matters for broader panvascular and interventional proposals. The full-watershed perspective recognizes organ-specific vascular architectures within a systemic framework [19]. Our findings are compatible with retaining that local context when evaluating molecular measurements. They do not validate a device–vessel compatibility score. Similarly, proposals linking intravascular imaging and digital twins emphasize multimodal interpretation and acknowledge unresolved generalization across vascular regions [20]. A tissue classifier supplies neither a patient-specific dynamical model nor evidence that choosing an interventional device from its output improves outcomes. Such applications would require relevant imaging, hemodynamics, longitudinal measurements, and prospective clinical evaluation.

Several limitations govern the strength of the evidence. First, the source unit discrepancy is unresolved: 104 deposited arrays, 95 complete labels, and 96 arteries reported in the original methods cannot be treated as equivalent counts. Unknown cross-bed donor overlap may remain after all tested conventions. Second, source-wide original normalization means that training-only secondary operations do not deliver a fully inductive raw-data validation. The common vocabulary also used target annotation and probe availability. Third, surgical versus deceased-donor procurement is inseparable from class in the source data and remains so under arterial-bed holdout. Bulk expression may reflect cellular composition, tissue collection, preservation, and other unrecorded factors; a sample-level preservation map was unavailable. The analysis neither isolates the contribution of these factors nor identifies a causal molecular mechanism.

Fourth, the external sample sizes were modest. GSE43292 uncertainty respected known pairing, whereas GSE28829 donor dependence could not be reconstructed. Fixed-score bootstrap intervals omit source-model and preprocessing uncertainty. Perfect empirical ordering and boundary sensitivity or specificity can produce degenerate intervals, which do not imply population certainty. Subset refitting addressed a separate source-stability question without supplying a comprehensive confidence interval. Fifth, small source strata and near-ceiling discrimination limit resolution for distinguishing models. Fixed baselines reduced selection flexibility but do not constitute an exhaustive comparison of modern representation learning. Finally, prior access to public cohorts and publication-informed panels limits claims of untouched external validation. The random-panel, stability, threshold, geometry, and influence analyses were added after original results and must be interpreted as exploratory. Their internal records, including the sigmoid-saturation correction, improve traceability without converting them into preregistered confirmation.

The appropriate next step is to evaluate the same estimands in datasets with authenticated donor identifiers, explicitly separated acquisition batches, and comparable tissue definitions. Raw-data prospective preprocessing and truly locked multicenter sampling would test whether source ranking survives beyond the present archive. If a clinical application is proposed, the target population, decision threshold, and consequences of errors should be specified independently of these selected tissue labels.

In conclusion, pooled AUROC was slightly below the equally weighted within-bed mean in this anatomical comparison-population analysis. Source and external tissue ranking remained strong while the unchanged technical threshold did not separate either changed external contrast. Exploratory random-panel and membership analyses further showed that strong source ranking did not identify a unique gene signature. Reporting these properties separately, together with the empirical ceiling, sample-identity uncertainty, and preprocessing provenance, gives a more defensible account of vascular transcriptomic transport than pooled AUROC alone.

## Supporting information

Reproducibility archive: code, provenance and results

LaTeX source and editable figures

## Data and code availability

Official processed data are available from GEO accessions GSE100927, GSE43292, and GSE28829, with platforms GPL17077, GPL6244, and GPL570. The accompanying reproducibility archive contains acquisition code, source hashes, annotation mappings, the internal protocol and amendments, sample-label audits, analysis code, frozen coefficients, predictions, figures, and machine-readable result tables. Inputs can be reacquired from the recorded official URLs. The archive does not confer rights beyond those applying to the original data. A separate public archival identifier for the reproducibility archive has not yet been assigned.

## Ethics, consent, and study registration

This work analyzes existing public tissue-expression records and does not recruit participants, intervene, or access direct personal identifiers. Original cohort ethics and consent are described in the source publications. The authors confirm that applicable ethical and consent requirements for this secondary analysis have been met. No new institutional approval or exemption is claimed. This is not a clinical trial, and no trial registration is claimed. The internal analysis plan was not a prospective registry entry.

## Author contributions

Lingsen You, Zisu Peng, and Yujun Guo contributed equally to this work. All authors reviewed and approved the manuscript.

## DECLARATION OF GENERATIVE AI AND AI-ASSISTED TECHNOLOGIES

Codex assisted with LaTeX assembly, prose editing, and consistency checks. The authors reviewed and revised all AI-assisted text and take responsibility for the submitted manuscript.

