## Supplementary Methods, Tables S1-S10 and Figures S1-S2 for "Anatomical comparison populations and threshold transport in vascular transcriptomic classification"

Computational tables and provenance. No independent-patient total is constructed across cohorts.

### Supplementary methods and results

This supplement reports the saved outputs of a retrospective computational tissue-classification audit. Counts refer to arrays, conservative specimen-label groups, or explicitly paired patients as specified; they must not be summed into a total of independent patients. AUROC denotes area under the receiver-operating-characteristic curve; AP denotes the non-interpolated average precision calculated by `average_precision_score`, not a trapezoidal precision–recall area. BA denotes balanced accuracy. The internal repeat ranges describe partition sensitivity and are not confidence intervals. External intervals condition on the fitted source model and fixed scores. The probability threshold of 0.5 and Brier scores are technical diagnostics of selected tissue contrasts, not population-risk calibration or validated clinical decision thresholds.

### Exact computational specification

For each platform, only probes mapping to one unambiguous gene symbol were retained; multiple probes for a gene were combined by the median within each array. The common vocabulary was the intersection of the three platform mappings and available probe identifiers, without use of target expression distributions or target outcomes. Fractional ranks used average ranks for ties, divided by 15,235. Ranks were computed within each array before the median over repeated full specimen labels. No missing ranks occurred. Native-expression sensitivity used the already processed deposited values unchanged. No joint cohort normalization, target-derived scaling, ComBat correction, external threshold selection or external recalibration was performed by the present pipeline.

The executed training-only feature score for gene $g$ was

$$T_g = \frac{|\bar{x}_{1g} - \bar{x}_{0g}|}{\sqrt{s_{1g}^2/n_1 + s_{0g}^2/n_0 + 10^{-12}}},$$

with unbiased within-class variances. The largest 200 scores were selected using a stable sort over the alphabetically ordered vocabulary. This is a Welch-type statistic, not Cohen's standardized mean difference. Training means and population standard deviations were then fitted by `StandardScaler`; the same fitted transformation was applied to held-out samples.

For case bed $a$ and control bed $b$, $A_{ab}$ is the proportion of score pairs with the case higher, with one-half credit for ties. Within each held-out fold, with $n_{1a}$ cases and $n_{0b}$ controls,

$$A_{\text{pooled}} = \frac{\sum_{a,b} n_{1a} n_{0b} A_{ab}}{\sum_{a,b} n_{1a} n_{0b}}, \qquad A_{\text{within}} = \frac{1}{3}\sum_a A_{aa}, \qquad \Delta_{\text{mix}} = A_{\text{pooled}} - A_{\text{within}}.$$

The identity is checked against the directly calculated fold AUROC. Case and control bed proportions were varied independently over the 66-point simplex grid in steps of 0.1 (4,356 combinations). Because this grid includes simplex vertices, its extrema equal the minimum and maximum of the nine observed pair-cell AUROCs; the grid is an algebraic reweighting, not a validation cohort. Scores from separately fitted folds were never pooled for this decomposition.

**Table S1:** Dataset units, contrasts and analytical roles

| Dataset | Arrays | Independent-patient status | Tissue contrast | Platform | Role |
|---|---|---|---|---|---|
| GSE100927 | 104 | Unknown; 95 distinct full specimen labels do not establish independent donors | Surgically obtained atherosclerotic arterial specimens; comparator: Arteries without atherosclerotic lesions from deceased organ donors | GPL17077; Agilent SurePrint G3 Human GE v2 8x60K | Development, repeated-label grouping sensitivity, arterial-bed holdout and restricted permutation |
| GSE43292 | 64 | 32 patients, each contributing two tissue samples (explicit GEO pairing) | Plaque core/shoulders, Stary IV or above, from hypertensive endarterectomy patients; comparator: Distant macroscopically intact same-patient tissue, Stary I/II; not healthy donors | GPL6244; Affymetrix Human Gene 1.0 ST | Locked external paired lesion-stage transport; donor-level bootstrap and within-pair concordance |
| GSE28829 | 29 | Unverified; 29 tissue arrays and no explicit donor IDs in deposited metadata | Advanced lesion: thin- or thick-fibrous-cap atheroma; comparator: Early lesion: pathological intimal thickening/intimal xanthoma; not healthy volunteers | GPL570; Affymetrix Human Genome U133 Plus 2.0 | Locked external early-versus-advanced lesion-stage challenge; sample-level uncertainty with independence limitation |



GSE100927 contains 69 disease and 35 control arrays. Conservative full-label aggregation produces 65 disease and 30 control groups: carotid 27/10, femoral 25/10, infrapopliteal 13/10. The original report states 65 disease and 31 healthy arteries; the femoral-control discrepancy remains unresolved. GSE43292 comprises 32 plaque/intact pairs. GSE28829 comprises 16 advanced and 13 early lesions. None of these endpoints has a follow-up time or censoring variable.

**Table S1:** Dataset provenance and preprocessing (continued)

| Dataset | Download completion | Bytes | SHA256 (four consecutive blocks) | Matrix probes / matched probes / genes | Inherited preprocessing |
|---|---|---|---|---|---|
| GSE100927 | 2026-10-25 15:54:14.648675 UTC | 7,149,412 | `1f096514b8668716`<br>`6c2271efd8e197f9`<br>`a11c522059b7b77e`<br>`8504ef66a7b82c42` | 47,264 / 43,144 / 30,556 | Depositor LOWESS against virtual median of all arrays; replicate probes averaged; low-intensity filtering |
| GSE43292 | 2026-10-25 15:54:13.183185 UTC | 7,231,220 | `9bf92c40eccbe359`<br>`4f0b4550e54e81a5`<br>`8365ba4a2251a0d6`<br>`bd7fda439a20f573` | 33,297 / 22,151 / 21,406 | Expression Console v1.1 default RMA summarization, as stated in deposited sample metadata |

Continued on next page

**Table S1:** Dataset provenance and preprocessing (continued)

| Dataset | Download completion | Bytes | SHA256 (four consecutive blocks) | Matrix probes / matched probes / genes | Inherited preprocessing |
|---|---|---|---|---|---|
| GSE28829 | 2026-10-25 15:54:13.558867 UTC | 9,527,080 | `022546a68ab6f2c6` `ceab3b9d9d57e9eb` `d0aa592185e4c51d` `6eeead9e6539ce48` | 54,675 / 43,161 / 21,813 | RMA with background subtraction, median-polish summarization and quantile normalization (affy) |

The common annotation-defined vocabulary contains 15,235 genes. All matrices had zero missing probe values, and none required the originally specified log transformation. Source-normalization procedures used whole deposited cohorts; training-only downstream fitting cannot remove that inherited transductive aspect. Exact URLs and downloaded-file hashes are in `evidence/acquisition.json`. GEO permits unrestricted download but does not certify absence of submitter intellectual-property claims (`https://www.ncbi.nlm.nih.gov/geo/info/disclaimer.html`). Source ethics approvals and consent statements refer to the original studies. Public availability does not itself create a new ethics approval or exemption for the present analysis.

**Table S2:** All nine repeated specimen-label pairs and profile similarity

| Full label after colon | Array A | Source prefix A | Array B | Source prefix B | Sex | Rank-profile r | Same numeric prefix |
|---|---|---|---|---|---|---|---|
| Carotid artery_10 | GSM2696620 | H112_26 | GSM2696680 | H34_26b | male | 0.99243 | Yes |
| Carotid artery_44 | GSM2696645 | H144_81 | GSM2696686 | H52_141 | female | 0.98348 | No |
| Carotid artery_Control 10 | GSM2696615 | H106_89b | GSM2696700 | H79_89 | male | 0.99173 | Yes |
| Carotid artery_Control 22 | GSM2696627 | H123_1 | GSM2696697 | H72_198 | male | 0.98965 | No |
| Femoral artery_27 | GSM2696638 | H138_185b | GSM2696665 | H182_185 | male | 0.99084 | Yes |
| Femoral artery_Control 05 | GSM2696635 | H133_44b | GSM2696669 | H190_44 | male | 0.99190 | Yes |
| Femoral artery_Control 25 | GSM2696660 | H17_200 | GSM2696691 | H6_4 | male | 0.98406 | No |
| Infra-popliteal artery_17 | GSM2696666 | H184_196 | GSM2696684 | H40_24 | male | 0.99081 | No |
| Infra-popliteal artery_Control 11 | GSM2696651 | H154_194 | GSM2696671 | H194_201 | female | 0.99055 | No |

Correlations are Pearson correlations between sample-wise fractional-rank vectors over the 15,235 common genes. High similarity does not authenticate donor identity or establish technical replication. Four pairs share a numeric pre-colon component with a b suffix in one array; five share only the post-colon specimen label. The original metadata label `Femoral artery_41G` is retained. All 18 arrays belonging to these nine pairs are excluded in the discard-ambiguous sensitivity, leaving 86 arrays.

**Table S3:** All models: mean and range of repetition-level mean fold statistics

| Model | Statistic | AUROC | AP | Within-bed AUROC | $\Delta_{\text{mix}}$ | Mix span | BA | Sensitivity | Specificity | Brier |
|---|---|---|---|---|---|---|---|---|---|---|
| Prevalence | Mean | 0.500 | 0.684 | 0.500 | 0.000 | 0.000 | 0.500 | 1.000 | 0.000 | 0.216 |
| | Min | 0.500 | 0.684 | 0.500 | 0.000 | 0.000 | 0.500 | 1.000 | 0.000 | 0.216 |
| | Max | 0.500 | 0.684 | 0.500 | 0.000 | 0.000 | 0.500 | 1.000 | 0.000 | 0.216 |
| Bed + sex | Mean | 0.538 | 0.705 | 0.471 | 0.067 | 1.000 | 0.549 | 0.719 | 0.380 | 0.249 |
| | Min | 0.495 | 0.682 | 0.428 | 0.039 | 1.000 | 0.494 | 0.556 | 0.333 | 0.246 |
| | Max | 0.576 | 0.731 | 0.518 | 0.082 | 1.000 | 0.577 | 0.800 | 0.433 | 0.254 |
| C1Q | Mean | 0.924 | 0.960 | 0.948 | -0.024 | 0.353 | – | – | – | – |
| | Min | 0.918 | 0.957 | 0.933 | -0.034 | 0.311 | – | – | – | – |
| | Max | 0.929 | 0.963 | 0.962 | -0.012 | 0.417 | – | – | – | – |
| Wu6 | Mean | 0.969 | 0.986 | 0.993 | -0.023 | 0.190 | 0.879 | 0.865 | 0.893 | 0.094 |
| | Min | 0.963 | 0.982 | 0.988 | -0.033 | 0.143 | 0.856 | 0.846 | 0.867 | 0.092 |
| | Max | 0.974 | 0.988 | 0.997 | -0.017 | 0.299 | 0.889 | 0.878 | 0.900 | 0.095 |
| Ridge-200 | Mean | 0.997 | 0.999 | 1.000 | -0.003 | 0.026 | 0.955 | 0.943 | 0.967 | 0.036 |
| | Min | 0.994 | 0.997 | 1.000 | -0.006 | 0.012 | 0.928 | 0.890 | 0.967 | 0.029 |
| | Max | 0.998 | 0.999 | 1.000 | -0.002 | 0.042 | 0.967 | 0.968 | 0.967 | 0.049 |
| Centroid-200 | Mean | 0.986 | 0.994 | 1.000 | -0.014 | 0.094 | – | – | – | – |
| | Min | 0.978 | 0.990 | 0.997 | -0.022 | 0.052 | – | – | – | – |
| | Max | 0.992 | 0.997 | 1.000 | -0.008 | 0.142 | – | – | – | – |
| Forest-200 | Mean | 0.993 | 0.997 | 1.000 | -0.007 | 0.047 | 0.960 | 0.974 | 0.947 | 0.037 |
| | Min | 0.991 | 0.996 | 0.997 | -0.009 | 0.028 | 0.927 | 0.953 | 0.900 | 0.033 |
| | Max | 0.995 | 0.998 | 1.000 | -0.004 | 0.063 | 0.975 | 0.985 | 0.967 | 0.040 |

Each repetition first averages its three held-out folds equally; this table then summarizes the ten repetition means. Minimum and maximum are split-sensitivity ranges, not confidence limits. Within-bed AUROC is the equal mean of the three diagonal case-bed/control-bed cells. $\Delta_{\text{mix}}$ equals pooled minus within-bed AUROC. Mix span is the maximum minus minimum over the independently varied case/control bed-proportion grid. C1Q and centroid are uncalibrated ranking scores, so threshold and Brier metrics are inapplicable. The retrospective literature panels were informed by these public cohorts and are not independent biomarker confirmations.

**Table S4:** All 18 primary-model anatomical transport settings

| Training bed(s) | Held-out bed | Proxy purge | Train n | Removed n | Test n | AUROC | AP | BA | Sensitivity | Specificity | Brier |
|---|---|---|---|---|---|---|---|---|---|---|---|
| Other two | Carotid | No | 58 | 0 | 37 | 1.000 | 1.000 | 0.850 | 1.000 | 0.700 | 0.048 |
| Other two | Carotid | Yes | 49 | 9 | 37 | 1.000 | 1.000 | 0.950 | 1.000 | 0.900 | 0.030 |
| Femoral | Carotid | No | 35 | 0 | 37 | 1.000 | 1.000 | 0.950 | 1.000 | 0.900 | 0.035 |
| Femoral | Carotid | Yes | 31 | 4 | 37 | 1.000 | 1.000 | 0.950 | 1.000 | 0.900 | 0.012 |
| Infrapopliteal | Carotid | No | 23 | 0 | 37 | 1.000 | 1.000 | 0.800 | 1.000 | 0.600 | 0.063 |
| Infrapopliteal | Carotid | Yes | 18 | 5 | 37 | 1.000 | 1.000 | 0.850 | 1.000 | 0.700 | 0.054 |
| Other two | Femoral | No | 60 | 0 | 35 | 0.996 | 0.998 | 0.820 | 0.640 | 1.000 | 0.187 |
| Other two | Femoral | Yes | 52 | 8 | 35 | 0.996 | 0.998 | 0.800 | 0.600 | 1.000 | 0.198 |
| Carotid | Femoral | No | 37 | 0 | 35 | 0.980 | 0.993 | 0.600 | 0.200 | 1.000 | 0.522 |
| Carotid | Femoral | Yes | 33 | 4 | 35 | 0.976 | 0.992 | 0.580 | 0.160 | 1.000 | 0.562 |
| Infrapopliteal | Femoral | No | 23 | 0 | 35 | 0.996 | 0.998 | 0.940 | 0.880 | 1.000 | 0.049 |
| Infrapopliteal | Femoral | Yes | 19 | 4 | 35 | 1.000 | 1.000 | 0.940 | 0.880 | 1.000 | 0.066 |
| Other two | Infrapopliteal | No | 72 | 0 | 23 | 1.000 | 1.000 | 0.962 | 0.923 | 1.000 | 0.057 |
| Other two | Infrapopliteal | Yes | 63 | 9 | 23 | 1.000 | 1.000 | 0.885 | 0.769 | 1.000 | 0.087 |
| Carotid | Infrapopliteal | No | 37 | 0 | 23 | 1.000 | 1.000 | 0.500 | 0.000 | 1.000 | 0.535 |
| Carotid | Infrapopliteal | Yes | 32 | 5 | 23 | 1.000 | 1.000 | 0.500 | 0.000 | 1.000 | 0.549 |
| Femoral | Infrapopliteal | No | 35 | 0 | 23 | 1.000 | 1.000 | 0.962 | 0.923 | 1.000 | 0.026 |
| Femoral | Infrapopliteal | Yes | 31 | 4 | 23 | 1.000 | 1.000 | 0.962 | 0.923 | 1.000 | 0.047 |

Counts denote conservative label groups. Proxy purging removes source controls with the same control-number suffix and sex as a target control; it is a sensitivity scenario for unresolved shared-donor identity, not confirmed donor linkage. Each row fits only its stated training data, including feature selection and scaling. No target recalibration is applied. The 12 additional unpurged leave-one-bed-out comparator rows are retained in `results/transport_metrics.csv`.

**Table S5:** External ranking metrics for five expression rules (four source-fitted; C1Q fit-free)

| Dataset | Model | Tissue arrays | AUROC [95% interval] | AP [95% interval] |
|---|---|---|---|---|
| GSE43292 | C1Q | 64 | 0.828 [0.739, 0.912] | 0.837 [0.750, 0.921] |
| GSE28829 | C1Q | 29 | 0.952 [0.841, 1.000] | 0.976 [0.924, 1.000] |
| GSE43292 | Wu6 | 64 | 0.846 [0.759, 0.926] | 0.827 [0.720, 0.934] |
| GSE28829 | Wu6 | 29 | 0.971 [0.899, 1.000] | 0.983 [0.943, 1.000] |
| GSE43292 | Ridge-200 | 64 | 0.877 [0.792, 0.947] | 0.870 [0.772, 0.952] |
| GSE28829 | Ridge-200 | 29 | 0.986 [0.942, 1.000] | 0.989 [0.961, 1.000] |
| GSE43292 | Centroid-200 | 64 | 0.873 [0.786, 0.945] | 0.866 [0.763, 0.950] |
| GSE28829 | Centroid-200 | 29 | 1.000 [1.000, 1.000] | 1.000 [1.000, 1.000] |
| GSE43292 | Forest-200 | 64 | 0.874 [0.795, 0.948] | 0.871 [0.781, 0.954] |
| GSE28829 | Forest-200 | 29 | 0.846 [0.663, 0.986] | 0.838 [0.686, 0.990] |

Intervals use 2,000 fixed-score bootstrap resamples (seed 20261025). GSE43292 is resampled by patient, retaining both tissues; GSE28829 is resampled by specimen within lesion class because donor IDs are unavailable. Intervals exclude source-training, model-choice, annotation and unresolved donor-dependence uncertainty. A boundary interval of [1,1] reflects the finite-sample bootstrap distribution, not certainty about population performance.



**Table S5:** Paired GSE43292 contrasts (continued)

| Model | Plaque > intact | Ties | Concordance [exact 95% CI] | Concordance [bootstrap 95% interval] | Mean paired score difference [bootstrap 95% interval] |
|---|---|---|---|---|---|
| C1Q | 27/32 | 0 | 0.844 [0.672, 0.947] | 0.844 [0.719, 0.969] | 0.08292 [0.05052, 0.12208] |
| Wu6 | 29/32 | 0 | 0.906 [0.750, 0.980] | 0.906 [0.812, 1.000] | 0.28805 [0.20223, 0.37735] |
| Ridge-200 | 29/32 | 0 | 0.906 [0.750, 0.980] | 0.906 [0.781, 1.000] | 0.00728 [0.00187, 0.01665] |
| Centroid-200 | 29/32 | 0 | 0.906 [0.750, 0.980] | 0.906 [0.781, 1.000] | 0.76594 [0.55985, 0.97136] |
| Forest-200 | 29/32 | 0 | 0.906 [0.750, 0.980] | 0.906 [0.781, 1.000] | 0.11371 [0.08670, 0.14340] |

Concordance assigns one to a higher plaque score and one-half to a tie within the same patient. The exact interval is Clopper–Pearson because all five models have zero ties. Score differences use each model's native score scale and cannot be compared as calibrated biological effect sizes. This within-patient estimand differs from marginal AUROC, which compares all positive and negative tissues.

**Table S5:** External probability-output diagnostics at the fixed 0.5 threshold (continued)

| Dataset | Model | BA [95% interval] | Sensitivity [95% interval] | Specificity [95% interval] | Brier [95% interval] |
|---|---|---|---|---|---|
| GSE43292 | Wu6 | 0.734 [0.641, 0.828] | 0.938 [0.844, 1.000] | 0.531 [0.344, 0.688] | 0.180 [0.141, 0.220] |
| GSE28829 | Wu6 | 0.500 [0.500, 0.500] | 0.000 [0.000, 0.000] | 1.000 [1.000, 1.000] | 0.258 [0.217, 0.303] |
| GSE43292 | Ridge-200 | 0.500 [0.500, 0.500] | 0.000 [0.000, 0.000] | 1.000 [1.000, 1.000] | 0.493 [0.484, 0.498] |
| GSE28829 | Ridge-200 | 0.500 [0.500, 0.500] | 0.000 [0.000, 0.000] | 1.000 [1.000, 1.000] | 0.418 [0.379, 0.452] |
| GSE43292 | Forest-200 | 0.562 [0.516, 0.625] | 0.125 [0.031, 0.250] | 1.000 [1.000, 1.000] | 0.219 [0.205, 0.232] |
| GSE28829 | Forest-200 | 0.500 [0.500, 0.500] | 0.000 [0.000, 0.000] | 1.000 [1.000, 1.000] | 0.256 [0.240, 0.273] |

Only models with probability outputs are shown. These class-balanced classifier outputs were not recalibrated externally. Good ranking can coexist with failure of a fixed threshold. Brier scores here describe selected histological contrasts; they do not estimate calibration for population disease or future clinical events.

**Table S6:** All eight prespecified primary-model sensitivities

| Scenario | Units | Complete folds | Mean AUROC | AUROC repeat range | Mean AP | Within-bed AUROC | $\Delta_{\text{mix}}$ | Mix span | BA | Brier |
|---|---|---|---|---|---|---|---|---|---|---|
| Top 50 genes | 95 | 30/30 | 0.996 | 0.994–0.998 | 0.998 | 1.000 | -0.004 | 0.032 | 0.948 | 0.048 |
| Top 500 genes | 95 | 30/30 | 0.997 | 0.995–0.998 | 0.999 | 1.000 | -0.003 | 0.029 | 0.962 | 0.030 |
| C = 0.01 | 95 | 30/30 | 0.994 | 0.990–0.998 | 0.997 | 1.000 | -0.006 | 0.042 | 0.935 | 0.068 |
| C = 1 | 95 | 30/30 | 0.998 | 0.995–1.000 | 0.999 | 1.000 | -0.002 | 0.018 | 0.966 | 0.026 |
| Native processed values | 95 | 30/30 | 0.997 | 0.994–0.998 | 0.999 | 1.000 | -0.003 | 0.025 | 0.958 | 0.035 |
| Exclude ambiguous arrays | 86 | 30/30 | 0.994 | 0.985–0.998 | 0.997 | 0.999 | -0.005 | 0.038 | 0.949 | 0.042 |
| 104 arrays, grouped folds | 104 | 30/30 | 0.996 | 0.993–0.999 | 0.998 | 1.000 | -0.004 | 0.031 | 0.965 | 0.028 |
| Possible shared controls | 95 | 30/30 | 0.996 | 0.992–0.998 | 0.998 | 1.000 | -0.004 | 0.036 | 0.960 | 0.034 |

Other specifications remain fixed. Means average the three folds within each repetition and then ten repetitions. Grouped-array and possible-control scenarios use stratified group folds; all 30 test folds in every scenario retained the six bed-by-status strata needed for the nine-cell decomposition. Units are arrays in the 104-array and exclusion scenarios and conservative labels otherwise. These results are sensitivity analyses, not independent studies.

**Table S7:** Full-pipeline label-permutation controls

| Scheme | Permutations | Observed first-repeat AUROC | Null mean | Null min | Null max | Null >= observed | Plus-one tail fraction |
|---|---|---|---|---|---|---|---|
| within bed | 99 | 0.998 | 0.503 | 0.271 | 0.715 | 0 | 0.010 |
| unrestricted | 99 | 0.998 | 0.506 | 0.299 | 0.685 | 0 | 0.010 |

Each scheme has 99 permutations at the 95-label unit. Within-bed permutations preserve class counts by bed; unrestricted permutations do not. Every permutation regenerates stratified folds and repeats feature selection, source scaling and fitting. The plus-one tail fraction is $(1 + \#\{A_{\text{null}} \geq A_{\text{observed}}\})/(99 + 1)$; its minimum resolution is 0.01. This is a pipeline-null diagnostic, not a significance test of anatomical transport or clinical usefulness.

**Table S7:** Thirty complete-pipeline subsamples (continued)

| Statistic | Mean | Minimum | Maximum |
|---|---|---|---|
| auc | 0.995 | 0.985 | 1.000 |
| within bed auc | 1.000 | 0.995 | 1.000 |
| delta mix | -0.005 | -0.015 | 0.000 |
| mix min | 0.965 | 0.917 | 1.000 |
| mix max | 1.000 | 1.000 | 1.000 |
| mix span | 0.035 | 0.000 | 0.083 |
| cross bed pair fraction | 0.667 | 0.667 | 0.667 |

Every subsample selects without replacement the floor of 80% of each of the six bed-by-status groups: 75 conservative label groups in every run. The full three-fold primary procedure is refitted. Ranges are stability descriptions, not confidence intervals. Repeated partitions and subsamples share observations.

**Table S8:** Model recipes, seeds and reproducibility specification

| Component | Executed specification |
|---|---|
| Prevalence | Constant training prevalence; no expression features. |
| Bed + sex | Three fixed bed indicators and one male indicator; training scaling and class-balanced L2 logistic regression, C=0.1. |
| C1Q | Mean of C1QA and C1QC fractional ranks; no fitting or probability conversion. Retrospective literature control. |
| Wu6 | ADAP2, CSF3R, FABP5, ITGAX, MYOC, SPP1; all six are in the common vocabulary; scaled class-balanced L2 logistic regression, C=0.1. |
| Ridge-200 | Top 200 Welch-type features, StandardScaler, class-balanced L2 logistic regression; solver=liblinear, C=0.1, max_iter=2000. |
| Centroid-200 | Same top-200 training-selected features and scaling. Score = mean of standardized features multiplied by the sign of the training case-control mean difference. |
| Forest-200 | Same top-200 features and scaling; 300 trees, max_depth=3, min_samples_leaf=3, max_features=sqrt, class_weight=balanced, n_jobs=2. |
| Repeated validation | Three folds stratified by bed and status; ten repeats; random_state=20261025+r, r=0,...,9. Fixed model specifications; no outcome-driven hyperparameter search. |
| Grouped sensitivities | StratifiedGroupKFold with the same repeat seeds; groups are full specimen labels or the explicit control-suffix-plus-sex uncertainty proxy. |
| Frozen external models | Fit each specified model to all 95 source label groups; random_state=20261025. No target adaptation. |
| External uncertainty | 2,000 bootstrap resamples per model/cohort; seed=20261025. Complete patient blocks for GSE43292; class-stratified specimen resampling for GSE28829. |
| Null and subset randomness | NumPy default_rng(20261116), shared sequentially through 99 within-bed permutations, 99 unrestricted permutations, then 30 stratified subsamples. Null fold seed=20261025; subset fold seed=20261025+iteration. |

All hyperparameters are fixed specifications, not results of nested tuning. The 200-gene final source model is reconstructed from `results/frozen_primary_coefficients.csv` and `results/frozen_primary_model.json`; the former contains gene, coefficient, source mean and source standard deviation for every selected gene. The fitted intercept is 0.28075375142531145. These coefficients permit score reconstruction but are not interpreted as causal gene effects.

For the frozen primary model, the reproducible score is

$$p(x) = \mathrm{logit}^{-1}\left(0.28075375142531145 + \sum_{g=1}^{200} \beta_g \frac{x_g - \mu_g}{\sigma_g}\right).$$

The CSV preserves full numerical precision and the order of selected genes; duplicating this 200-row machine-readable table in the PDF is unnecessary. The source-only classifier probabilities must not be interpreted as population disease probabilities.

**Table S8:** Machine-readable result and code index (continued)

| Table | Relative file path | Contents |
|---|---|---|
| S1 | `evidence/dataset_registry.csv` | Dataset units, official sources, usage conditions, ethical boundaries, downloads and hashes. |
| S1 | `evidence/acquisition.json` | Complete matrix and extracted platform-table provenance; full huge platform families were not downloaded. |
| S1 | `evidence/preprocessing_qc.json` | Probe mapping, expression ranges, missingness and common-gene count. |
| S2 | `results/ambiguous_label_profile_similarity.csv` | All nine pair correlations and exact labels/source prefixes. |
| S2 | `results/unmatched_within_stratum_similarity.csv` | Nonmatching-label profile similarities within bed and status. |
| S3 | `results/primary_fold_metrics.csv` | All 210 fold/model records. |
| S3 | `results/primary_predictions.csv;results/primary_pair_cells.csv` | Held-out scores/partitions and all nine comparison cells per fold/model. |
| S4 | `results/transport_metrics.csv;results/transport_predictions.csv` | All 30 transport metric rows, including the 18 primary-model settings, with predictions. |
| S5 | `results/external_metrics.csv;results/external_predictions.csv;results/external_pairs.csv` | Ten cohort/model summaries, all individual scores and 160 paired score contrasts. |
| S6 | `results/sensitivity_all_fold_metrics.csv` | 240 fold records across eight specifications; corresponding scenario predictions and pair-cell tables are also retained. |
| S7 | `results/permutation_nulls.csv;results/full_pipeline_subsampling.csv` | 198 complete-pipeline null statistics and 30 subsampling records. |
| S8 | `results/frozen_primary_coefficients.csv;results/frozen_primary_model.json` | All 200 coefficients, training scaling and intercept for the frozen primary model. |
| S8 | `protocol/analysis_protocol_v1.txt;protocol/amendment_01_before_fit.txt;protocol/implementation_note_02.txt` | Internal chronology, pre-fit clarification and post-result terminology/error-handling note. |
| S8 | `evidence/protocol_freeze_sha256.json;evidence/amendment_01_sha256.json` | Protocol hashes; not registration certificates. |
| S8 | `evidence/independent_saved_results_validation.json` | Saved-prediction metric/decomposition checks and primary-score reconstruction. |
| S8 | `code/acquire.py;code/prepare.py;code/analyze.py;code/validate_results.py` | Acquisition, deterministic preparation, fitting/validation and independent saved-result checks. |
| S8 | `code/plot_figures.py;code/build_supplement.py` | Programmatic figure and supplementary-table generation. |

##### Protocol chronology and disclosure of prior exposure

The internally time-stamped protocol was recorded on 25 October 2026 before new-study expression model fitting; it was not preregistration. Metadata, original papers and several previously published results were already known. GSE43292 had previously supported descriptive module contrasts in a separate VascFlux-GEO manuscript; the present work does not claim that this public cohort was previously untouched. The current endpoint, fitted models, anatomical decomposition and transport results were newly calculated here.

The pre-fit amendment, after outcome-agnostic preprocessing QC, specified that probability diagnostics apply only to probability-output models, retained deposited native values without transformation, added sex to the conservative control-linkage proxy, confirmed the 15,235-gene vocabulary, fixed rank-before-label-aggregation ordering, and clarified interval interpretation. After main and robustness results were observed, an implementation note corrected the feature-ranking terminology to the executed Welch-type statistic. It also narrowed decomposition error handling so a failed algebraic identity raises an error, while a missing stratum is explicitly recorded. These changes did not alter saved predictions or select a model on external outcomes. The saved-result audit independently verified all complete-fold decompositions and reconstructed the primary external scores. This is an audit of recorded computations, not independent clinical validation or proof that all possible biases are absent.

##### Execution environment and commands

Snapshot: Python 3.11.9; numpy 2.4.6, pandas 2.3.3, scipy 1.17.1, scikit-learn 1.9.0. The working directory is the project root. The supported analysis sequence is:

```
python -m pip install -r requirements.txt
python code/acquire.py
python code/metadata.py
python code/prepare.py
python code/analyze.py main
python code/analyze.py robust
python code/validate_results.py
python code/export_descriptive_scores.py
python code/plot_figures.py
python code/build_supplement.py
python code/assemble.py --compile
```

Acquisition is unnecessary when the verified source files are already present. Figure generation and supplementary assembly do not refit the models. Official processed matrix files, data hashes, fitted-source parameters, saved predictions, protocol records and dependency snapshots should accompany any reproducibility release. Public source data remain subject to their original repository conditions.

#### Statistical reporting clarifications and descriptive score aggregation

##### Descriptive averages of repeated held-out scores

The internal pre-fit protocol promised a separate pooled AUROC based on averaged cross-fitted scores. This display was assembled after internal review from the original saved predictions, without refitting or selecting a model. The unit is a complete-title label group, not an authenticated subject or patient. Each of the 95 label groups contributes exactly one held-out score in each of the ten repetitions for each of seven models. Averaging these ten scores produces 665 label-group/model records, exported in `results/descriptive_mean_crossfit_scores.csv`; the corresponding metrics are in `results/descriptive_mean_crossfit_metrics.csv`.

| Model | Averaged-score pooled AUROC | Primary equal-fold mean AUROC |
|---|---:|---:|
| Prevalence | 0.483590 | 0.500000 |
| Bed + sex | 0.491282 | 0.537518 |
| C1Q | 0.922564 | 0.924372 |
| Wu6 | 0.966154 | 0.969329 |
| Ridge-200 | 0.996923 | 0.996775 |
| Centroid-200 | 0.988718 | 0.985981 |
| Forest-200 | 0.990769 | 0.993074 |

The averaged-score AUROCs are descriptive and are not substituted for the primary within-fold nine-cell estimand or treated as independent validation. Different label groups receive predictions from different fitted models, and the ten repetitions share observations and training data. In particular, the prevalence model has AUROC 0.5 in every fold because its score is constant within that fold, while different training-fold prevalences can produce unequal label-group averages and a different descriptive pooled AUROC. C1Q and centroid outputs are uncalibrated scores; the other model outputs are technical classifier probabilities, not population disease risks. No interval is attached to this descriptive display. The no-refit export and checks are implemented in `code/export_descriptive_scores.py`, with input/output hashes and validation details in `evidence/descriptive_mean_crossfit_export_audit.json`.

#### Composition contrast, ceiling, and grid extrema

The protocol specified the signed contrast between pooled AUROC and the equally weighted diagonal mean; it did not specify a one-sided primary positive-inflation hypothesis and was not a registry-based preregistration. All three primary-model diagonal cells were 1 in every held-out fold, so $\Delta_{\text{mix}} = A_{\text{pooled}} - 1 \leq 0$ necessarily held in these data. The observed negative sign cannot establish population-level mixture invariance. For any case and control compositions $p$ and $q$, $p^{\mathsf{T}}Aq$ is a convex combination of the nine cells. Because the 0.1 grid includes all simplex vertices, its extrema are exactly the smallest and largest cells. Mean per-fold extrema were 0.974290 and 1.000000 (mean span 0.025710), whereas extrema of the mean cell matrix were 0.976582 and 1.000000. The smallest individual fold cell was 0.875; individual cells contained 12–36 overlapping case–control pairs. Grid scenarios are deterministic descriptions, including extreme single-bed case/control mixtures; their histogram is not a probability distribution over plausible deployment populations.

#### Boundary intervals and subsample denominators

The frozen primary model classified 0/32 plaque tissues as positive and 32/32 intact tissues as negative in GSE43292, and 0/16 advanced specimens as positive and 13/13 early specimens as negative in GSE28829. Fixed-score empirical resampling therefore necessarily returns sensitivity $[0, 0]$ and specificity $[1, 1]$ in these settings, with balanced accuracy $[0.5, 0.5]$. Likewise, perfect observed ordering can yield AUROC $[1, 1]$. These degenerate conditional intervals describe the resampled empirical support; they do not imply certainty about future or population performance. They exclude source fitting and preprocessing uncertainty, and the GSE28829 intervals additionally retain the unresolved donor-independence limitation. The originally specified intervals are retained; no post hoc interval replacement or threshold calibration was performed.

The 30 source-stability subsets used the floor of 80% within each of six strata: disease counts $\lfloor 0.8(27) \rfloor + \lfloor 0.8(25) \rfloor + \lfloor 0.8(13) \rfloor = 21 + 20 + 10 = 51$, plus $3 \times 8 = 24$ controls. Thus each subset contained 75 of 95 label groups (78.9% overall), rather than 76 groups from applying 80% once to the total. These overlapping refitted subsets describe stability and are not independent replications or a full-pipeline confidence interval.

#### Post-result exploratory extensions

These analyses were specified on 26 October 2026 after the original results had been examined. They add diagnostics; they do not replace the primary analysis or constitute preregistered confirmation. The original result files were retained byte-for-byte. The dated record is `protocol/exploratory_extension_04_20261026.txt`; the numerical correction is documented in `implementation_note_05_logit_ranking.txt`.

##### Representation controls and recovered feature membership

Twenty panels of 200 genes were drawn without replacement within each panel from the 15,235-gene common vocabulary using NumPy `default_rng(20261026)`. Panels may overlap each other. Their genes were drawn without using labels and fixed across the original 30 folds and the full-source fits; supervised coefficient fitting still used the source outcomes. All panels are reported, with no external winner selected. A disjoint alternative selected training-only absolute Welch-type ranks 201–400. Scaling, regularization ($C = 0.1$), class weighting and folds matched the primary ridge procedure. There were 630 additional source-fold fits and 21 full-source fits. Thirty primary fits were separately recovered to obtain feature membership and signed coefficients; recovered held-out probabilities agreed with the saved original values. The 435 Jaccard coefficients compare overlapping, dependent fits and are not replicates for confidence-interval estimation. Feature sampling is not a label-permutation null.

One panel's sigmoid probabilities saturated to exactly 1 for all 64 GSE43292 tissues. The saved unsquashed linear scores retained 64 distinct values. We therefore compute AUROC, average precision and paired ordering uniformly from the linear logit for all new logistic representation controls. Probabilities remain the inputs to Brier scores and threshold diagnostics. The original primary outputs did not change. For this panel (random 18), logit AUROC was 0.7207 and paired concordance was 29/32; a probability implementation would have introduced artificial ties. Both score forms are archived.

##### Threshold recipes and prior pilot exposure

Before the formal exploratory record, a pilot used each group's average of ten held-out source probabilities. Its maximum-balanced-accuracy midpoint was 0.2251206725 and produced external balanced accuracies 0.5000 and 0.53125. This prior observation is disclosed and is not replaced by a claim that the later analyses were blind. The reported full-source recipes use one score per group from the original first three-fold repetition, rather than averaging probabilities from ten fitted models. Candidate thresholds are 0, 1, and the midpoints between consecutive distinct scores; classification uses score $\geq$ threshold. Maximum Youden $J$ is selected with an absolute tie tolerance of $10^{-12}$, preferring the candidate closest to 0.5 and then the lower value. The sensitivity rule chooses the largest candidate with source sensitivity at least 0.90. The first-repetition source split seed is 20261025.

For anatomical holdouts, three-fold inner cross-fitting is stratified by the four available bed–status cells, using seeds 20261026, 20261027 and 20261028 for the carotid, femoral and infrapopliteal targets. Gene selection, scaling and fitting occur inside each inner training fold. These nine inner fits choose thresholds; three outer fits on the remaining beds generate held-out-bed probabilities. Thus target outcomes do not choose the thresholds. Source selection quantities are optimization results, not unbiased performance estimates. Target-label threshold curves are descriptive only. Pooling cross-fitted source scores to choose a threshold does not alter the primary within-fold AUROC estimand and can remain sensitive to fold-to-fold scale.

#### Score arithmetic, failure maps and display summaries

The frozen score is the intercept plus the sum of its 200 signed standardized gene contributions. Source values from this full-source model are in-sample references. Overall means depend on class proportions, so class-specific means are also supplied. The fraction of feature values with $|z| > 3$ is a scale-displacement diagnostic, not a patient-level error rate or a validated out-of-distribution detector. Named display features are selected by source coefficients rather than target labels. Failure maps include all 32 known patient pairs and all five expression rules. Delete-one-patient and delete-one-specimen calculations keep predictions fixed; they are influence analyses, not refitting, new validation or uncertainty intervals. Displays of the three reversed primary pairs are explicitly selected after observing their failures and give arithmetic explanations rather than causal mechanisms. Supplementary Figures S1–S2 show the original nine-cell fold values and repeated source-group probabilities; repeated partitions are dependent.

**Table S9:** All exploratory representation controls. Source values are means of the same 30 held-out folds. External values use full-source fits and linear-logit ordering. The primary row is the original reference model. All 20 random draws are retained.

| Panel | Source AUC | GSE43292 | | | GSE28829 | |
|---|---|---|---|---|---|---|
| | | AUC | Pair concord. | BA | AUC | BA |
| Primary | 0.9968 | 0.8770 | 0.9062 | 0.5000 | 0.9856 | 0.5000 |
| Ranks 201–400 | 0.9955 | 0.8379 | 0.9062 | 0.5000 | 0.8558 | 0.5000 |
| Random 01 | 0.9989 | 0.8213 | 0.8438 | 0.5000 | 0.8702 | 0.5000 |
| Random 02 | 0.9983 | 0.7656 | 0.8438 | 0.7031 | 0.9183 | 0.5000 |
| Random 03 | 0.9941 | 0.6865 | 0.8438 | 0.5156 | 0.8990 | 0.5000 |
| Random 04 | 0.9960 | 0.8623 | 0.9062 | 0.5000 | 0.9231 | 0.5000 |
| Random 05 | 0.9957 | 0.7754 | 0.8438 | 0.5000 | 0.9904 | 0.5938 |
| Random 06 | 0.9978 | 0.7607 | 0.8750 | 0.7500 | 0.6587 | 0.5000 |
| Random 07 | 0.9960 | 0.6367 | 0.8125 | 0.5000 | 0.7548 | 0.5000 |
| Random 08 | 0.9948 | 0.7236 | 0.8125 | 0.5938 | 0.7644 | 0.5000 |
| Random 09 | 0.9974 | 0.6123 | 0.5938 | 0.5000 | 0.5337 | 0.5000 |
| Random 10 | 0.9968 | 0.6162 | 0.7500 | 0.5156 | 0.8173 | 0.5000 |
| Random 11 | 0.9949 | 0.7686 | 0.9062 | 0.5000 | 0.8077 | 0.5000 |
| Random 12 | 0.9963 | 0.7461 | 0.8750 | 0.5312 | 0.8846 | 0.5000 |
| Random 13 | 0.9969 | 0.7783 | 0.9062 | 0.5000 | 0.8413 | 0.5000 |
| Random 14 | 0.9949 | 0.7295 | 0.9062 | 0.5781 | 0.8365 | 0.5000 |
| Random 15 | 0.9969 | 0.8604 | 0.8750 | 0.5000 | 0.9327 | 0.5000 |
| Random 16 | 0.9977 | 0.7959 | 0.9062 | 0.5000 | 0.8269 | 0.5000 |
| Random 17 | 0.9932 | 0.8242 | 0.8750 | 0.5000 | 0.8510 | 0.5000 |
| Random 18 | 0.9992 | 0.7207 | 0.9062 | 0.5000 | 0.7596 | 0.5000 |
| Random 19 | 0.9959 | 0.7900 | 0.8750 | 0.5000 | 0.7981 | 0.5000 |
| Random 20 | 0.9940 | 0.8096 | 0.8750 | 0.5156 | 0.7933 | 0.6226 |

Pair concordance compares plaque with relatively intact tissue within each of 32 known patients; marginal AUC compares all tissues across classes. BA is balanced accuracy at the technical probability threshold 0.5. No draw ranges are confidence intervals, and near-ceiling source performance does not identify a unique panel.

**Table S10:** Evaluation of source-only threshold rules. Each anatomical holdout uses nested source-only threshold selection; external cohorts use unchanged full-source thresholds. The table reports every evaluated rule, not a target-selected winner.

| Target | Rule | Threshold | TP | FN | TN/FP | Sens. | BA |
|---|---|---|---|---|---|---|---|
| Carotid | Fixed 0.5 | 0.5000 | 27 | 0 | 7/3 | 1.000 | 0.850 |
| Carotid | Max. J | 0.2423 | 27 | 0 | 6/4 | 1.000 | 0.800 |
| Carotid | Sens. 0.90 | 0.4968 | 27 | 0 | 7/3 | 1.000 | 0.850 |
| Femoral | Fixed 0.5 | 0.5000 | 16 | 9 | 10/0 | 0.640 | 0.820 |
| Femoral | Max. J | 0.4002 | 19 | 6 | 10/0 | 0.760 | 0.880 |
| Femoral | Sens. 0.90 | 0.5820 | 14 | 11 | 10/0 | 0.560 | 0.780 |
| Infrapopliteal | Fixed 0.5 | 0.5000 | 12 | 1 | 10/0 | 0.923 | 0.962 |
| Infrapopliteal | Max. J | 0.2976 | 12 | 1 | 10/0 | 0.923 | 0.962 |
| Infrapopliteal | Sens. 0.90 | 0.5310 | 12 | 1 | 10/0 | 0.923 | 0.962 |
| GSE43292 | Fixed 0.5 | 0.5000 | 0 | 32 | 32/0 | 0.000 | 0.500 |
| GSE43292 | Max. J | 0.6173 | 0 | 32 | 32/0 | 0.000 | 0.500 |
| GSE43292 | Sens. 0.90 | 0.6922 | 0 | 32 | 32/0 | 0.000 | 0.500 |
| GSE28829 | Fixed 0.5 | 0.5000 | 0 | 16 | 13/0 | 0.000 | 0.500 |
| GSE28829 | Max. J | 0.6173 | 0 | 16 | 13/0 | 0.000 | 0.500 |
| GSE28829 | Sens. 0.90 | 0.6922 | 0 | 16 | 13/0 | 0.000 | 0.500 |

TP, FN, TN and FP denote true positives, false negatives, true negatives and false positives. Sens. is sensitivity, BA is balanced accuracy; specificity can be recovered as TN/(TN+FP). These labels concern sampled tissue contrasts, not patient risk. The full candidate curves and all 12 learning-context/rule records, including source optimization quantities, are retained in the accompanying CSV files.

#### Reproduction and complete tabular outputs

`results/exploratory_features/` contains panel memberships, all source and external predictions, fold metrics, all 6,000 primary selections, stability values, and 435 Jaccard comparisons. `results/exploratory_threshold/` contains all threshold candidates/rules/evaluations, class-specific logit accounting, 37,600 gene contributions, all paired and specimen deletion values, and failure maps. `results/exploratory_display/` contains repeated source-score summaries. No per-feature contribution row is an independent biological replicate.

Starting from the saved inputs, `code/explore_feature_dependence.py` and `code/explore_threshold_shift.py` rebuild the new fitted analyses. `code/validate_exploratory.py` independently recomputes metrics and performs limited independent fitting checks. The three `plot_*_extension.py` scripts render Figures 7–11 and S1–S2. Then run `assemble_figure_catalog.py`, `build_exploratory_supplement.py`, and `assemble.py -compile`. These commands are run from the project root with the `code/` prefix. The archive README gives the ordered full workflow, original analysis commands, dependencies, and the limits of the relocated smoke check.

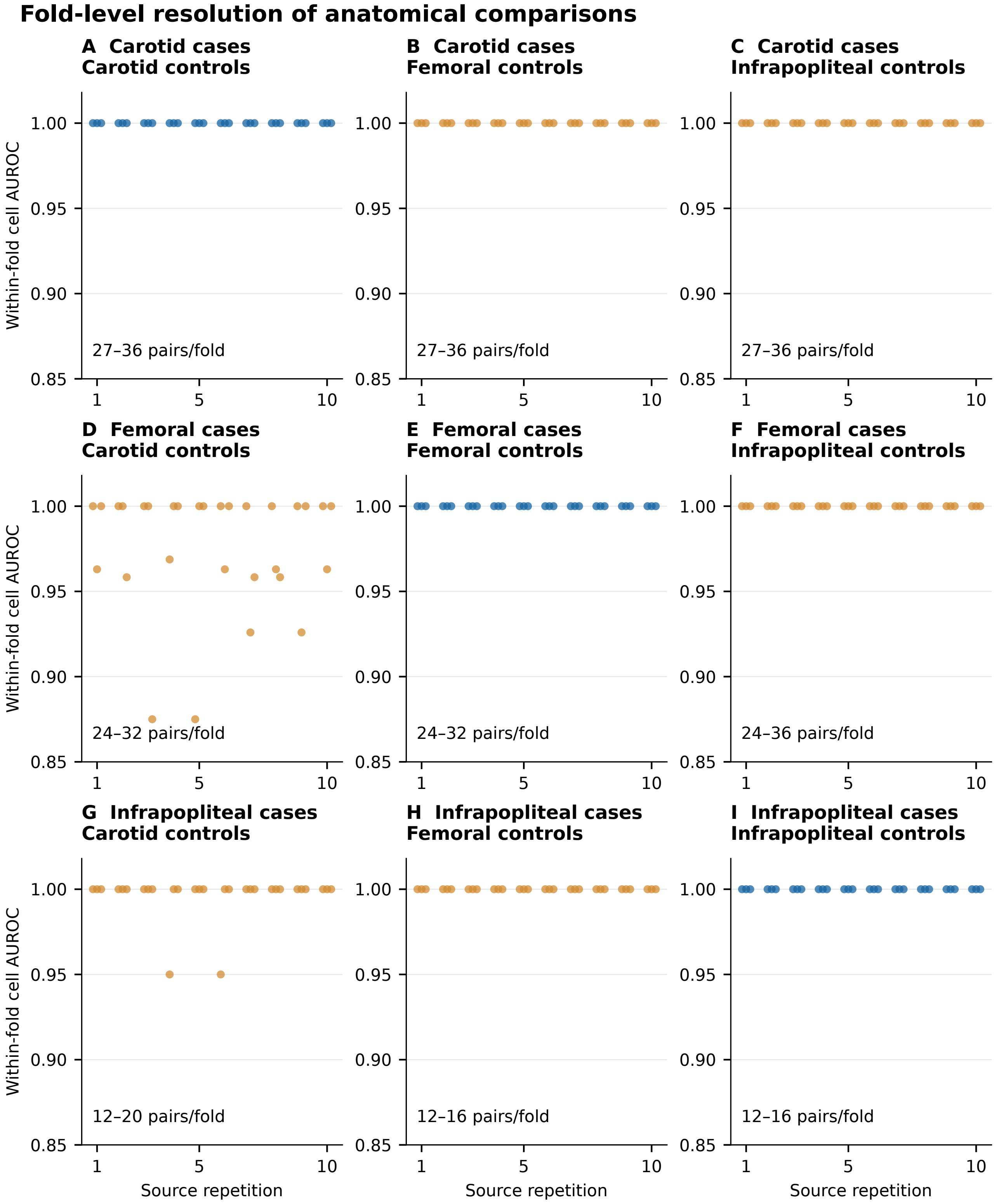


**Figure S1: Fold-level resolution of anatomical comparisons**. A–I, All 30 held-out folds for each primary-model case-bed/control-bed comparison. Horizontal jitter separates the three folds of each repetition. Blue denotes diagonal within-bed comparisons; orange denotes cross-bed comparisons. The AUROC axis is deliberately truncated at 0.85 to display the smallest observed cell (0.875). Labels give the actual pair-count range in each cell; pairs share observations, and folds/repetitions share source data. Points are not independent studies, and no confidence interval is implied. These original saved results complement the mean matrix in Figure 2; no new model was fitted for this display.

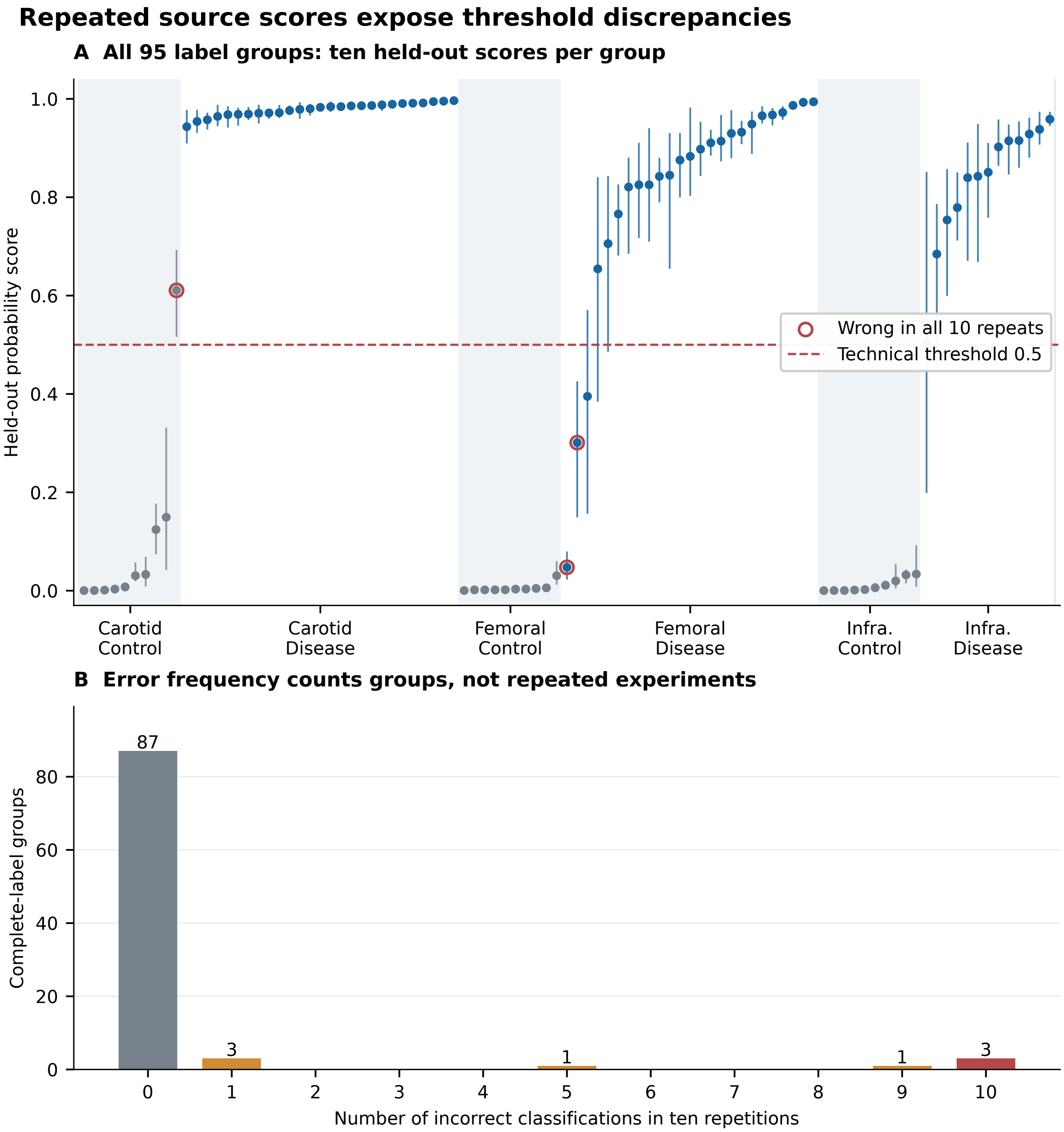


**Figure S2: Repeated source scores expose threshold discrepancies**. A, Each point is one group's mean of ten original held-out primary scores, with its minimum-to-maximum range. Groups are ordered within bed/status by mean score; this ordering is descriptive. Red outlines identify groups misclassified in every repetition at 0.5. The display-position-to-label mapping is supplied in results/exploratory_display/source_score_stability.csv. B, Counts of groups by their number of threshold errors. Source groups are unverified donor units, repeated fits are dependent, and ranges are not confidence intervals. These pooled score summaries do not replace the original within-fold AUROC estimand. No new fitting or threshold selection was performed for this display.
