## Supplementary material for "Anatomical comparison populations and threshold transport in vascular transcriptomic classification": Reproducibility archive: code, provenance and results: acquire.py

```
"""Verify/reuse frozen GEO inputs; download missing inputs without rewriting provenance.

Default reproduction refuses changed bytes. A changed upstream version requires a
separate, deliberately versioned analysis project; this script never blesses it.
Only the first complete platform table is downloaded from each platform archive.
"""
from pathlib import Path
import argparse, concurrent.futures, datetime, gzip, hashlib, json, requests

ROOT = Path(__file__).resolve().parents[1]
EXPECTED = ROOT / 'evidence' / 'expected_inputs_v1.json'

def now(): return datetime.datetime.now(datetime.timezone.utc).isoformat()
def sha(path):
    h = hashlib.sha256()
    with path.open('rb') as f:
        for chunk in iter(lambda: f.read(2**20), b''): h.update(chunk)
    return h.hexdigest()

def verify(path, record):
    return path.stat().st_size == record['bytes'] and sha(path) == record['sha256']

def acquire(record, offline=False):
    relative = Path(record['path'].replace('\\', '/'))
    path = (ROOT / relative).resolve()
    if not path.is_relative_to((ROOT / 'data').resolve()):
        raise ValueError('Input path is outside this project data directory')
    if path.exists():
        if not verify(path, record):
            raise ValueError(f'{relative}: local bytes differ from the frozen input manifest; nothing overwritten')
        return {'accession': record['accession'], 'status': 'verified_existing', 'sha256': record['sha256']}
    if offline:
        raise FileNotFoundError(f'{relative}: unavailable in offline verification')
    path.parent.mkdir(parents=True, exist_ok=True)
    temporary = path.with_name(path.name + '.download-partial')
    start = now()
    try:
        with requests.get(record['url'], stream=True, timeout=(30,180)) as response:
            response.raise_for_status()
            if record['kind'] == 'complete_processed_matrix':
                with temporary.open('wb') as out:
                    for chunk in response.iter_content(2**20): out.write(chunk)
                with gzip.open(temporary, 'rb') as stream: content = stream.read()
                if not all(x in content for x in [b'!series_matrix_table_begin', b'!series_matrix_table_end']):
                    raise ValueError('Incomplete matrix')
            elif record['kind'] == 'complete_first_platform_table_only':
                response.raw.decode_content = False
                seen = False
                with gzip.GzipFile(fileobj=response.raw) as stream, temporary.open('wb') as out:
                    for line in stream:
                        if line.startswith(b'!platform_table_begin'): seen = True
                        if seen:
                            out.write(line)
                            if line.startswith(b'!platform_table_end'): break
                    else: raise ValueError('Incomplete first platform table')
            else: raise ValueError('Unexpected input kind')
        if not verify(temporary, record):
            raise ValueError(f'{relative}: downloaded bytes differ from the frozen manifest; original expectation retained')
        temporary.replace(path)
        return {'accession': record['accession'], 'status': 'downloaded_and_verified',
                'start_utc': start, 'finish_utc': now(), 'sha256': record['sha256']}
    finally:
        if temporary.exists(): temporary.unlink()

def main():
    parser = argparse.ArgumentParser(description=__doc__)
    parser.add_argument('--offline', action='store_true', help='Verify bundled inputs only; never access the network')
    args = parser.parse_args()
    records = json.loads(EXPECTED.read_text(encoding='utf8'))
    if {r['accession'] for r in records} != {'GSE100927','GSE43292','GSE28829','GPL17077','GPL6244','GPL570'} or len(records) != 6:
        raise ValueError('Frozen manifest must contain all six exact inputs')
    results = []
    with concurrent.futures.ThreadPoolExecutor(max_workers=3) as pool:
        futures = {pool.submit(acquire,r,args.offline):r['accession'] for r in records}
        for future in concurrent.futures.as_completed(futures):
            try: result = future.result()
            except Exception as exc: result = {'accession':futures[future], 'error':repr(exc)}
            results.append(result)
            print(json.dumps(result), flush=True)
    audit = {'created_utc': now(), 'offline': args.offline, 'expected_manifest_sha256': sha(EXPECTED),
             'original_acquisition_record_unchanged': True, 'results': results,
             'passed': all('error' not in r for r in results)}
    (ROOT/'evidence'/'input_verification_latest.json').write_text(json.dumps(audit,indent=2), encoding='utf8')
    raise SystemExit(0 if audit['passed'] else 1)

if __name__ == '__main__': main()
```
