## Supplementary material for "Anatomical comparison populations and threshold transport in vascular transcriptomic classification": Reproducibility archive: code, provenance and results: analyze.py

```
"""Prespecified anatomical-comparison audit. See frozen protocol for estimands."""
import os
os.environ.setdefault('OMP_NUM_THREADS','2')
os.environ.setdefault('OPENBLAS_NUM_THREADS','2')
from pathlib import Path
import json,time,datetime,itertools,warnings
import numpy as np,pandas as pd
from scipy.special import expit
from scipy.stats import beta,spearmanr
from sklearn.model_selection import StratifiedKFold,StratifiedGroupKFold
from sklearn.linear_model import LogisticRegression
from sklearn.preprocessing import StandardScaler
from sklearn.ensemble import RandomForestClassifier
from sklearn.metrics import roc_auc_score,average_precision_score,balanced_accuracy_score,brier_score_loss

ROOT=Path(__file__).resolve().parents[1];OUT=ROOT/'results';OUT.mkdir(exist_ok=True)
SEED=20261025;BEDS=['Carotid','Femoral','Infrapopliteal']
MODELS=['prevalence','bed_sex','C1Q','Wu6','ridge200','centroid200','forest200']
PANEL=['ADAP2','CSF3R','FABP5','ITGAX','MYOC','SPP1']
def save(name,rows):pd.DataFrame(rows).to_csv(OUT/name,index=False)
def log(s):print(datetime.datetime.now(datetime.timezone.utc).isoformat(),s,flush=True)

def load(acc,rep='rank',unit='labels'):
    x=pd.read_pickle(ROOT/'data'/f'{acc}_{rep}.pkl')
    m=pd.read_csv(ROOT/'data'/f'{acc}_samples.csv',index_col='sample').fillna('unknown')
    if acc=='GSE100927' and unit=='labels':
        x=x.groupby(m.label_group,sort=True).median()
        cols=['bed','y','sex','identity_proxy','patient']
        assert (m.groupby('label_group')[['bed','y','sex']].nunique().max()<=1).all()
        first=m.groupby('label_group',sort=True).first()
        first['array_count']=m.groupby('label_group').size()
        first['sample_members']=m.groupby('label_group').apply(lambda z:';'.join(z.index),include_groups=False) if pd.__version__>='2.2' else m.groupby('label_group').apply(lambda z:';'.join(z.index))
        m=first; m['label_group']=m.index
    elif acc=='GSE100927' and unit=='exclude_ambiguous':
        keep=~m.label_group.duplicated(keep=False);m=m.loc[keep];x=x.loc[m.index]
    assert list(x.index)==list(m.index)
    return x,m

def bedsex(m):
    return np.column_stack([(m.bed==b).to_numpy(dtype=float) for b in BEDS]+[(m.sex=='male').to_numpy(dtype=float)])

class Fit:
    def __init__(self,kind,k=200,C=.1,seed=SEED):self.kind=kind;self.k=k;self.C=C;self.seed=seed
    def fit(self,x,m):
        y=m.y.to_numpy(dtype=int);self.prev=y.mean();self.features=[]
        if self.kind in ['prevalence','C1Q']:return self
        if self.kind=='bed_sex':v=bedsex(m)
        else:
            a=x.to_numpy(dtype=float)
            if self.kind=='Wu6':self.features=[g for g in PANEL if g in x.columns]
            else:
                d=a[y==1].mean(0)-a[y==0].mean(0)
                se=np.sqrt(a[y==1].var(0,ddof=1)/(y==1).sum()+a[y==0].var(0,ddof=1)/(y==0).sum()+1e-12)
                order=np.argsort(-np.abs(d/se),kind='stable')[:self.k]
                self.features=x.columns[order].tolist()
            v=x[self.features].to_numpy()
        self.scaler=StandardScaler().fit(v);z=self.scaler.transform(v)
        if self.kind=='centroid200':
            self.sign=np.sign(z[y==1].mean(0)-z[y==0].mean(0));return self
        if self.kind=='forest200':
            self.model=RandomForestClassifier(n_estimators=300,max_depth=3,min_samples_leaf=3,max_features='sqrt',class_weight='balanced',random_state=self.seed,n_jobs=2)
        else:self.model=LogisticRegression(C=self.C,class_weight='balanced',solver='liblinear',max_iter=2000,random_state=self.seed)
        self.model.fit(z,y);return self
    def predict(self,x,m):
        if self.kind=='prevalence':return np.full(len(m),self.prev)
        if self.kind=='C1Q':return x[['C1QA','C1QC']].mean(axis=1).to_numpy()
        v=bedsex(m) if self.kind=='bed_sex' else x[self.features].to_numpy()
        z=self.scaler.transform(v)
        if self.kind=='centroid200':return (z*self.sign).mean(axis=1)
        return self.model.predict_proba(z)[:,1]

def metrics(y,s,prob=True):
    y=np.array(y,dtype=int);s=np.array(s)
    r={'auc':roc_auc_score(y,s),'ap':average_precision_score(y,s)}
    if prob:
        p=s>=.5;r.update(balanced_accuracy=balanced_accuracy_score(y,p),sensitivity=p[y==1].mean(),specificity=(~p[y==0]).mean(),brier=brier_score_loss(y,s))
    return r

GRID=np.array([[i/10,j/10,(10-i-j)/10] for i in range(11) for j in range(11-i)])
def decompose(y,s,bed):
    y=np.asarray(y);s=np.asarray(s);bed=np.asarray(bed);cells=[];mat=np.empty((3,3));w=np.empty((3,3))
    for a,b in itertools.product(range(3),repeat=2):
        case=s[(y==1)&(bed==BEDS[a])];ctrl=s[(y==0)&(bed==BEDS[b])]
        assert len(case)>0 and len(ctrl)>0
        auc=((case[:,None]>ctrl).astype(float)+.5*(case[:,None]==ctrl)).mean()
        mat[a,b]=auc;w[a,b]=len(case)*len(ctrl)
        cells.append({'case_bed':BEDS[a],'control_bed':BEDS[b],'auc':auc,'pairs':int(w[a,b])})
    pooled=(mat*w).sum()/w.sum();diag=np.diag(mat).mean()
    assert abs(pooled-roc_auc_score(y,s))<1e-10
    mixed=GRID@mat@GRID.T
    return dict(auc=pooled,within_bed_auc=diag,delta_mix=pooled-diag,mix_min=mixed.min(),mix_max=mixed.max(),mix_span=mixed.max()-mixed.min(),cross_bed_pair_fraction=1-np.trace(w)/w.sum()),cells

def splits(m,seed,group=None):
    strata=m.bed.astype(str)+'_'+m.y.astype(str)
    if group is None:
        return list(StratifiedKFold(3,shuffle=True,random_state=seed).split(m,strata))
    return list(StratifiedGroupKFold(3,shuffle=True,random_state=seed).split(m,strata,groups=group))

def cv(x,m,kinds=MODELS,repeats=10,k=200,C=.1,scenario='primary',group=None,record=True):
    rows=[];pred=[];cellrows=[]
    for rep in range(repeats):
        for fold,(tr,te) in enumerate(splits(m,SEED+rep,group)):
            for kind in kinds:
                fit=Fit(kind,k,C,SEED+rep).fit(x.iloc[tr],m.iloc[tr]);s=fit.predict(x.iloc[te],m.iloc[te])
                base=dict(scenario=scenario,model=kind,repeat=rep,fold=fold,n_train=len(tr),n_test=len(te))
                met=metrics(m.y.iloc[te],s,kind not in ['C1Q','centroid200'])
                complete=all(np.any((m.y.iloc[te].to_numpy()==yy)&(m.bed.iloc[te].to_numpy()==bb)) for yy in [0,1] for bb in BEDS)
                if complete:dec,cells=decompose(m.y.iloc[te],s,m.bed.iloc[te])
                else:dec={};cells=[] # Missing cells are recorded, while an algebra check failure is fatal.
                base['complete_six_strata']=complete
                rows.append(dict(**base,**(met|dec)))
                for c in cells:cellrows.append(base|c)
                for idx,score in zip(te,s):pred.append(base|dict(sample=str(m.index[idx]),bed=m.bed.iloc[idx],y=int(m.y.iloc[idx]),score=float(score)))
        log(f'CV {scenario} repeat {rep+1}/{repeats}')
    if record:
        save(f'{scenario}_fold_metrics.csv',rows);save(f'{scenario}_predictions.csv',pred);save(f'{scenario}_pair_cells.csv',cellrows)
    return pd.DataFrame(rows),pd.DataFrame(pred),pd.DataFrame(cellrows)

def transport(x,m):
    rows=[];pred=[];features=[]
    for target in BEDS:
        te=np.flatnonzero(m.bed==target)
        for source in ['other_two']+[b for b in BEDS if b!=target]:
            tr=np.flatnonzero(m.bed!=target) if source=='other_two' else np.flatnonzero(m.bed==source)
            for purge in [False,True]:
                tr2=tr
                if purge:
                    proxies=set((m.identity_proxy.astype(str)+'_'+m.sex.astype(str)).iloc[te][m.y.iloc[te]==0])
                    p=m.identity_proxy.astype(str)+'_'+m.sex.astype(str)
                    tr2=np.array([i for i in tr if not (m.y.iloc[i]==0 and p.iloc[i] in proxies)])
                for kind in (['C1Q','Wu6','ridge200','centroid200','forest200'] if source=='other_two' and not purge else ['ridge200']):
                    fit=Fit(kind).fit(x.iloc[tr2],m.iloc[tr2]);s=fit.predict(x.iloc[te],m.iloc[te])
                    base=dict(source=source,target=target,purge_possible_controls=purge,model=kind,n_train=len(tr2),n_removed=len(tr)-len(tr2),n_test=len(te))
                    rows.append(base|metrics(m.y.iloc[te],s,kind not in ['C1Q','centroid200']))
                    for i,ss in zip(te,s):pred.append(base|dict(sample=str(m.index[i]),y=int(m.y.iloc[i]),score=float(ss)))
    save('transport_metrics.csv',rows);save('transport_predictions.csv',pred)

def bootstrap_metrics(m,s,prob,paired=False,n=2000,seed=SEED):
    if paired:
        # The paired reshaping below requires complete two-tissue patient blocks.
        # Validate that contract before drawing, rather than relying on row count.
        if not {'patient','y'}.issubset(m.columns):
            raise ValueError('Paired bootstrap requires patient and y columns')
        if m.patient.isna().any() or not m.y.isin([0,1]).all():
            raise ValueError('Paired bootstrap requires known patient IDs and binary labels')
        blocks=m.groupby('patient',sort=False).y.agg(['size','sum'])
        if blocks.empty or not ((blocks['size']==2)&(blocks['sum']==1)).all():
            raise ValueError('Each paired bootstrap patient must have exactly two tissues, one per class')
    rng=np.random.default_rng(seed);vals=[]
    for j in range(n):
        if paired:
            pats=m.patient.unique();draw=rng.choice(pats,len(pats),replace=True)
            idx=np.concatenate([np.flatnonzero(m.patient==p) for p in draw])
        else:
            idx=np.concatenate([rng.choice(np.flatnonzero(m.y==c),sum(m.y==c),replace=True) for c in [0,1]])
        r=metrics(m.y.iloc[idx],s[idx],prob)
        if paired:
            # Each sampled block is a two-tissue patient with one of each class.
            z=s[idx].reshape(-1,2);yy=m.y.iloc[idx].to_numpy().reshape(-1,2)
            d=z[yy==1]-z[yy==0];r['pair_concordance']=np.mean((d>0)+.5*(d==0));r['paired_mean_difference']=d.mean()
        vals.append(r)
    q=pd.DataFrame(vals).quantile([.025,.975])
    return {k:(float(q.loc[.025,k]),float(q.loc[.975,k])) for k in q}

def external(x,m):
    rows=[];pred=[];coefs=[];pairedrows=[]
    for kind in ['C1Q','Wu6','ridge200','centroid200','forest200']:
        fit=Fit(kind).fit(x,m)
        if kind=='ridge200':
            for gene,coef,mu,scale in zip(fit.features,fit.model.coef_[0],fit.scaler.mean_,fit.scaler.scale_):coefs.append(dict(gene=gene,coefficient=coef,source_mean=mu,source_sd=scale))
            (OUT/'frozen_primary_model.json').write_text(json.dumps({'intercept':float(fit.model.intercept_[0]),'C':.1,'k':200,'source_units':len(m),'vocabulary_count':x.shape[1],'fit_utc':datetime.datetime.now(datetime.timezone.utc).isoformat()},indent=2))
        for acc in ['GSE43292','GSE28829']:
            xt,mt=load(acc);s=fit.predict(xt,mt);prob=kind not in ['C1Q','centroid200'];met=metrics(mt.y,s,prob)
            if acc=='GSE43292':
                ds=[]
                for p in mt.patient.unique():
                    ii=np.flatnonzero(mt.patient==p);yy=mt.y.iloc[ii].to_numpy();d=float(s[ii][yy==1][0]-s[ii][yy==0][0]);ds.append(d)
                    pairedrows.append(dict(model=kind,patient=p,plaque_score=float(s[ii][yy==1][0]),intact_score=float(s[ii][yy==0][0]),difference=d))
                d=np.array(ds);met['pair_concordance']=np.mean((d>0)+.5*(d==0));met['paired_mean_difference']=d.mean()
                if not np.any(d==0):
                    wins=sum(d>0);n=len(d);met['pair_exact_low']=0 if wins==0 else beta.ppf(.025,wins,n-wins+1);met['pair_exact_high']=1 if wins==n else beta.ppf(.975,wins+1,n-wins)
            cis=bootstrap_metrics(mt,s,prob,acc=='GSE43292')
            for key,(lo,hi) in cis.items():met[key+'_low']=lo;met[key+'_high']=hi
            rows.append(dict(dataset=acc,model=kind,n=len(mt))|met)
            for i,ss in enumerate(s):pred.append(dict(dataset=acc,model=kind,sample=mt.index[i],patient=mt.patient.iloc[i],y=int(mt.y.iloc[i]),score=float(ss)))
            log(f'external {kind} {acc} AUC {met["auc"]:.3f}')
    save('external_metrics.csv',rows);save('external_predictions.csv',pred);save('external_pairs.csv',pairedrows);save('frozen_primary_coefficients.csv',coefs)

def robustness(x,m):
    rows=[]
    for name,k,C,rep,unit,groupmode in [
        ('top50',50,.1,'rank','labels',None),('top500',500,.1,'rank','labels',None),('C001',200,.01,'rank','labels',None),('C1',200,1,'rank','labels',None),
        ('native',200,.1,'native','labels',None),('exclude_ambiguous',200,.1,'rank','exclude_ambiguous',None),('104_grouped_arrays',200,.1,'rank','arrays','title'),('possible_shared_controls',200,.1,'rank','labels','control')]:
        xx,mm=load('GSE100927',rep,unit)
        group=None
        if groupmode=='title':group=mm.label_group
        if groupmode=='control':group=pd.Series(['C_'+str(r.identity_proxy)+'_'+str(r.sex) if r.y==0 else str(i) for i,r in mm.iterrows()],index=mm.index)
        f,p,c=cv(xx,mm,['ridge200'],10,k,C,name,group,record=True)
        f['units']=len(mm);rows.append(f)
    pd.concat(rows).to_csv(OUT/'sensitivity_all_fold_metrics.csv',index=False)

def nulls_and_subsample(x,m):
    rng=np.random.default_rng(SEED+91);rows=[]
    for scheme in ['within_bed','unrestricted']:
        for it in range(99):
            mm=m.copy()
            if scheme=='within_bed':
                for b in BEDS:
                    idx=mm.bed==b;mm.loc[idx,'y']=rng.permutation(mm.loc[idx,'y'].to_numpy())
            else:mm['y']=rng.permutation(mm.y.to_numpy())
            vals=[]
            for fold,(tr,te) in enumerate(splits(mm,SEED)):
                fit=Fit('ridge200').fit(x.iloc[tr],mm.iloc[tr]);s=fit.predict(x.iloc[te],mm.iloc[te]);vals.append(roc_auc_score(mm.y.iloc[te],s))
            rows.append(dict(scheme=scheme,permutation=it,mean_fold_auc=np.mean(vals)))
            if (it+1)%25==0:log(f'null {scheme} {it+1}/99')
    save('permutation_nulls.csv',rows)
    sub=[]
    for it in range(30):
        ids=[]
        for _,gg in m.groupby(['bed','y']):ids.extend(rng.choice(gg.index,int(np.floor(.8*len(gg))),replace=False))
        mm=m.loc[ids];xx=x.loc[ids];vals=[]
        for fold,(tr,te) in enumerate(splits(mm,SEED+it)):
            fit=Fit('ridge200').fit(xx.iloc[tr],mm.iloc[tr]);s=fit.predict(xx.iloc[te],mm.iloc[te]);de,c=decompose(mm.y.iloc[te],s,mm.bed.iloc[te]);vals.append(de)
        means=pd.DataFrame(vals).mean().to_dict();sub.append(dict(iteration=it,n=len(mm))|means)
    save('full_pipeline_subsampling.csv',sub)

def identity_qc():
    x,m=load('GSE100927',unit='arrays');rows=[]
    corr=np.corrcoef(x.to_numpy());non=[]
    for lab,gg in m.groupby('label_group'):
        if len(gg)>1:
            for a,b in itertools.combinations(gg.index,2):
                i=m.index.get_loc(a);j=m.index.get_loc(b)
                rows.append(dict(label_group=lab,array_a=a,array_b=b,title_a=m.title.iloc[i],title_b=m.title.iloc[j],bed=m.bed.iloc[i],y=m.y.iloc[i],rank_profile_correlation=corr[i,j]))
    for i,j in itertools.combinations(range(len(m)),2):
        if m.label_group.iloc[i]!=m.label_group.iloc[j] and m.bed.iloc[i]==m.bed.iloc[j] and m.y.iloc[i]==m.y.iloc[j]:non.append(dict(bed=m.bed.iloc[i],y=m.y.iloc[i],correlation=corr[i,j]))
    save('ambiguous_label_profile_similarity.csv',rows);save('unmatched_within_stratum_similarity.csv',non)

if __name__=='__main__':
    import sys
    mode=sys.argv[1] if len(sys.argv)>1 else 'main'
    x,m=load('GSE100927');m.to_csv(OUT/'primary_analysis_units.csv',index_label='unit_id')
    assert len(m)==95 and x.shape[1]==15235
    if mode=='main':
        identity_qc();cv(x,m);transport(x,m);external(x,m)
    elif mode=='robust':robustness(x,m);nulls_and_subsample(x,m)
    else:raise ValueError(mode)
    log('finished '+mode)
```
