## Supplementary material for "Anatomical comparison populations and threshold transport in vascular transcriptomic classification": Reproducibility archive: code, provenance and results: assemble.py

```
"""Assemble author-confirmed identity, scientific body, figures and audit supplement."""
from pathlib import Path
import json,re,subprocess,hashlib,datetime,shutil,importlib.metadata,sys
ROOT=Path(__file__).resolve().parents[1];M=ROOT/'manuscript'
TITLE='Anatomical comparison populations and threshold transport in vascular transcriptomic classification'
AI_TITLE='DECLARATION OF GENERATIVE AI AND AI-ASSISTED TECHNOLOGIES'
AI_TEXT='Codex assisted with LaTeX assembly, prose editing, and consistency checks. The authors reviewed and revised all AI-assisted text and take responsibility for the submitted manuscript.'
def esc(s):
    for a,b in [('\\',r'\textbackslash{}'),('&',r'\&'),('%',r'\%'),('_',r'\_'),('#',r'\#')]:s=s.replace(a,b)
    s=s.replace('–','--').replace('—','---').replace('−','-').replace('×',r'$\times$').replace('≥',r'$\geq$').replace('≤',r'$\leq$').replace('≠',r'$\ne$')
    return s
PREAMBLE=r"""\documentclass[11pt,a4paper]{article}
\usepackage[margin=20mm]{geometry}
\usepackage[T1]{fontenc}
\usepackage[utf8]{inputenc}
\usepackage{lmodern,amsmath,amssymb,graphicx,booktabs,longtable,array,xcolor,pdflscape}
\usepackage{microtype,setspace,caption,float}
\usepackage[hidelinks]{hyperref}
\usepackage[numbers,sort&compress]{natbib}
\setstretch{1.12}
\setlength{\parindent}{0pt}
\setlength{\parskip}{5pt}
\captionsetup{font=small,labelfont=bf}
\begin{document}
"""
def generate():
    old=json.loads((M/'author_metadata.json').read_text(encoding='utf8'))
    portal_path=ROOT/'submission/portal_state.json'
    portal=json.loads(portal_path.read_text(encoding='utf-8-sig')) if portal_path.exists() else {}
    if portal and portal.get('title')!=TITLE:raise ValueError('Portal authorization belongs to another manuscript')
    final_authorized=portal.get('final_submit_authorized', old.get('final_submission_authorized'))
    keep={k:old[k] for k in ['authors','affiliations','equal_contribution_statement','corresponding_authors','submitting_author']}
    keep.update(title=TITLE,license='CC BY 4.0')
    if isinstance(final_authorized,bool):keep['final_submission_authorized']=final_authorized
    # Public packages deliberately omit private authorization fields and portal
    # records. A standalone build must preserve that minimized metadata exactly.
    if (portal_path.exists() and 'final_submit_authorized' in portal
            and 'final_submission_authorized' in old
            and isinstance(final_authorized,bool)
            and old['final_submission_authorized'] is not final_authorized):
        old['final_submission_authorized']=final_authorized
        (M/'author_metadata.json').write_text(json.dumps(old,ensure_ascii=False,indent=2)+'\n',encoding='utf8')
    # Identity/authorization metadata is a source record, not a build artifact.
    # Do not overwrite later confirmations during a document-only rebuild.
    pdf_authors=', '.join(a['given_name']+' '+a['family_name'] for a in keep['authors'])
    def preamble(pdf_title):
        metadata=r'\hypersetup{pdftitle={'+esc(pdf_title)+r'},pdfauthor={'+esc(pdf_authors)+r'},pdfsubject={medRxiv submission manuscript}}'+'\n'
        return PREAMBLE.replace(r'\begin{document}',metadata+r'\begin{document}')
    names=[]
    for a in keep['authors']:
        sup=','.join(a['affiliations'])+(',$\\dagger$' if a['equal_contribution'] else '')+(',$*$' if a['corresponding'] else '')
        names.append(esc(a['given_name']+' '+a['family_name'])+r'\textsuperscript{'+sup+'}')
    title=r'\begin{titlepage}\vspace*{2mm}{\Large\bfseries\raggedright\hyphenpenalty=10000 '+TITLE+r'\par}\vspace{5mm}'+'\n'
    title+=', '.join(names)+r'\par\vspace{3mm}'+'\n'+r'{\small'+'\n'
    for key,text in keep['affiliations'].items():title+=r'\textsuperscript{'+key+'}'+esc(text)+r'\par'+'\n'
    title+='}\n'+r'\vspace{4mm}\textsuperscript{$\dagger$}'+keep['equal_contribution_statement']+r'\par'+'\n'
    title+=r'\textsuperscript{$*$}Correspondence: Lingsen You (\href{mailto:}{}); Junbo Ge (\href{mailto:}{}).\par'+'\n'
    if (M/'body.tex').exists():
        body=(M/'body.tex').read_text(encoding='utf8')
        abstract,rest=body.split(r'\section{Introduction}',1)
        abstract=abstract.replace(r'\section*{Abstract}',r'\noindent\textbf{Abstract}\par')
        title+=r'\vspace{3mm}\begingroup\small\setstretch{1}'+'\n'+abstract+r'\endgroup'+'\n'
        (M/'body_for_compilation.tex').write_text(r'\section{Introduction}'+rest,encoding='utf8')
    title+=r'\end{titlepage}'+'\n'
    (M/'titlepage.tex').write_text(title,encoding='utf8')
    caps=json.loads((ROOT/'figures/figure_captions.json').read_text(encoding='utf8'))
    fs=[r'\newgeometry{left=20mm,right=20mm,top=12mm,bottom=14mm}']
    # Full 17-cm width preserves the designed minimum 8-pt labels.
    for i in range(1,len(caps)+1):
        c=caps[f'Figure_{i}']
        fs.append(r'\clearpage\begin{figure}[p]\centering\setstretch{1}'+'\n'+rf'\includegraphics[width=17cm]{{../figures/Figure_{i}.pdf}}'+'\n'+rf'\caption{{\textbf{{{esc(c["title"])}}}. {esc(c["caption"])}}}\label{{fig:{i}}}'+'\n'+r'\end{figure}\clearpage'+'\n')
    fs.append(r'\restoregeometry')
    (M/'figure_pages.tex').write_text('\n'.join(fs),encoding='utf8')
    team=(ROOT/'evidence/citations/recommended_team_references.bib').read_text(encoding='utf8')
    independent=(ROOT/'evidence/independent_references_verified.bib').read_text(encoding='utf8')
    (M/'references.bib').write_text(independent+'\n'+team,encoding='utf8')
    admin=(M/'declarations.tex').read_text(encoding='utf8') if (M/'declarations.tex').exists() else ''
    main=preamble(TITLE)+r'\hypersetup{pageanchor=false}\input{titlepage.tex}\hypersetup{pageanchor=true}'+'\n'+r'\input{body_for_compilation.tex}'+'\n'+r'\clearpage'+'\n'+admin+'\n'+r'\section*{\raggedright '+AI_TITLE.replace('TECHNOLOGIES',r'\mbox{TECHNOLOGIES}')+'}\n'+AI_TEXT+'\n'
    main+=r'\clearpage\bibliographystyle{unsrtnat}\bibliography{references}\clearpage\input{figure_pages.tex}'+'\n'+r'\end{document}'
    (M/'main.tex').write_text(main,encoding='utf8')
    supp=preamble('Supplementary material: '+TITLE)+r'{\Large\bfseries Supplementary material\par}'+'\n'+TITLE+r'\par\vspace{3mm}'+'\n'+r'Computational tables and provenance. No independent-patient total is constructed across cohorts.\par'+'\n'+r'\input{supplement_body.tex}'+'\n'
    if (M/'exploratory_supplement.tex').exists():supp+=r'\clearpage\input{exploratory_supplement.tex}'+'\n'
    sc=json.loads((ROOT/'figures/supplement_extension_captions.json').read_text(encoding='utf8'))
    supp+=r'\clearpage\renewcommand{\thefigure}{S\arabic{figure}}\setcounter{figure}{0}\newgeometry{left=20mm,right=20mm,top=12mm,bottom=14mm}'+'\n'
    for i in range(1,len(sc)+1):
        c=sc[f'Figure_S{i}']
        supp+=r'\clearpage\begin{figure}[p]\centering\setstretch{1}'+'\n'+rf'\includegraphics[width=17cm]{{../figures/Figure_S{i}.pdf}}'+'\n'+rf'\caption{{\textbf{{{esc(c["title"])}}}. {esc(c["caption"])}}}\label{{fig:s{i}}}'+'\n'+r'\end{figure}\clearpage'+'\n'
    supp+=r'\restoregeometry\end{document}'
    (M/'supplement.tex').write_text(supp,encoding='utf8')
    pkgs=['numpy','pandas','scipy','scikit-learn','matplotlib','requests','Pillow','pypdf']
    env={p:importlib.metadata.version(p) for p in pkgs}
    env.update(python=sys.version,executable=sys.executable,recorded_utc=datetime.datetime.now(datetime.timezone.utc).isoformat())
    env.update(scope='Document assembly environment, not the historical analysis environment',
        analysis_environment_sha256=hashlib.sha256((ROOT/'evidence/analysis_environment_original.json').read_bytes()).hexdigest(),
        analysis_requirements_sha256=hashlib.sha256((ROOT/'requirements.txt').read_bytes()).hexdigest())
    (ROOT/'evidence/manuscript_build_environment.json').write_text(json.dumps(env,indent=2),encoding='utf8')
    print('Assembled source files')
def compile_all():
    engine=subprocess.run(['pdflatex','--version'],capture_output=True,text=True,check=True).stdout
    engine_flags=['--disable-installer'] if 'miktex' in engine.lower() else []
    for name in ['main','supplement']:
        latex=['pdflatex']+engine_flags+['-interaction=nonstopmode','-halt-on-error',name+'.tex']
        seq=[latex]
        if name=='main':seq.append(['bibtex',name])
        seq += [latex]*2
        for j,cmd in enumerate(seq):
            p=subprocess.run(cmd,cwd=M,stdout=subprocess.PIPE,stderr=subprocess.STDOUT,text=True,encoding='utf8',errors='replace',timeout=180)
            (ROOT/'evidence'/f'compile_{name}_{j}.log').write_text(p.stdout,encoding='utf8')
            if p.returncode:print(p.stdout[-6500:]);raise RuntimeError(cmd)
        print(name,'compiled',flush=True)
if __name__=='__main__':
    generate()
    if '--compile' in sys.argv:compile_all()
```
