## Supplementary material for "Anatomical comparison populations and threshold transport in vascular transcriptomic classification": Reproducibility archive: code, provenance and results: assemble_figure_catalog.py

```
"""Merge independently generated figure sets and verify every referenced input."""
from pathlib import Path
import hashlib,json
ROOT=Path(__file__).resolve().parents[1];F=ROOT/'figures'
def sha(p):return hashlib.sha256(p.read_bytes()).hexdigest()
base=json.loads((F/'figure_captions.json').read_text(encoding='utf8'))
caps={f'Figure_{i}':base[f'Figure_{i}'] for i in range(1,7)}
for name in ['feature_extension_captions.json','threshold_extension_captions.json']:
    incoming=json.loads((F/name).read_text(encoding='utf8'))
    assert not set(caps)&set(incoming)
    caps.update(incoming)
assert set(caps)=={f'Figure_{i}' for i in range(1,12)}
inputs={}
for key,record in caps.items():
    normalized=[]
    for name in record['source_tables']:
        rel=name if name.startswith('results/') else 'results/'+name
        assert (ROOT/rel).is_file(),rel
        normalized.append(rel);inputs[rel]=sha(ROOT/rel)
    record['source_tables']=normalized
    assert record['panel_count']==len(record['panels'])
(F/'figure_captions.json').write_text(json.dumps(caps,ensure_ascii=False,indent=2),encoding='utf8')
supp=json.loads((F/'supplement_extension_captions.json').read_text(encoding='utf8'))
sp=json.loads((F/'supplement_extension_provenance.json').read_text(encoding='utf8'))
for rel,digest in sp['input_sha256'].items():
    assert (ROOT/rel).is_file() and sha(ROOT/rel)==digest,rel
    inputs[rel]=digest
rel='results/exploratory_display/source_score_stability.csv'
inputs[rel]=sha(ROOT/rel)
files={}
for key in list(caps)+list(supp):
    for ext in ['pdf','svg','png']:
        p=F/f'{key}.{ext}';assert p.is_file()
        files[p.name]={'bytes':p.stat().st_size,'sha256':sha(p)}
manifest={'script':'code/assemble_figure_catalog.py','input_sha256':inputs,'figures':files,
    'main_figures':len(caps),'main_panels':sum(c['panel_count'] for c in caps.values()),
    'supplementary_figures':len(supp),'supplementary_panels':sum(c['panel_count'] for c in supp.values()),
    'note':'Figures1-6 retain original analyses; Figures7-11 are explicitly post-result exploratory analyses. Supplementary displays show original saved score variation.',
    'caption_sha256':{'figure_captions.json':sha(F/'figure_captions.json'),
                      'supplement_extension_captions.json':sha(F/'supplement_extension_captions.json')}}
(F/'figure_provenance.json').write_text(json.dumps(manifest,indent=2),encoding='utf8')
print(json.dumps({k:manifest[k] for k in ['main_figures','main_panels','supplementary_figures','supplementary_panels']},indent=2))
```
