## Supplementary material for "Anatomical comparison populations and threshold transport in vascular transcriptomic classification": Reproducibility archive: code, provenance and results: build_exploratory_supplement.py

```
"""Generate post-result supplementary methods and Tables S9-S10 from saved values."""
from pathlib import Path
import datetime, hashlib, json
import pandas as pd

ROOT=Path(__file__).resolve().parents[1]
R=ROOT/'results'; used=[]
def read(name):
    p=R/name;used.append(p);return pd.read_csv(p)
def esc(s):
    return str(s).replace('_',r'\_').replace('%',r'\%').replace('&',r'\&')
def num(x,n=4):return f'{float(x):.{n}f}'

fold=read('exploratory_features/representation_fold_metrics.csv')
ext=read('exploratory_features/representation_external_metrics.csv')
primary=read('primary_fold_metrics.csv');primary=primary[primary.model=='ridge200']
pe=read('external_metrics.csv');pe=pe[pe.model=='ridge200'].set_index('dataset')
pairs=read('external_pairs.csv');pairs=pairs[pairs.model=='ridge200']
ev=read('exploratory_threshold/evaluation_metrics.csv')
rules=read('exploratory_threshold/threshold_rules.csv')

text=[r'''% Generated from saved outputs by code/build_exploratory_supplement.py.
\section*{Post-result exploratory extensions}
These analyses were specified on 26 October 2026 after the original results had been
examined. They add diagnostics; they do not replace the primary analysis or constitute
preregistered confirmation. The original result files were retained byte-for-byte.
The dated record is \texttt{protocol/exploratory\_extension\_04\_20261026.txt};
the numerical correction is documented in \texttt{implementation\_note\_05\_logit\_ranking.txt}.

\subsection*{Representation controls and recovered feature membership}
Twenty panels of 200 genes were drawn without replacement within each panel from the
15,235-gene common vocabulary using NumPy \texttt{default\_rng(20261026)}.
Panels may overlap each other. Their genes were drawn without using labels and fixed
across the original 30 folds and the full-source fits; supervised coefficient fitting
still used the source outcomes. All panels are reported, with no external winner selected.
A disjoint alternative selected training-only absolute Welch-type ranks 201--400.
Scaling, regularization ($C=0.1$), class weighting and folds matched the primary ridge
procedure. There were 630 additional source-fold fits and 21 full-source fits.
Thirty primary fits were separately recovered to obtain feature membership and signed
coefficients; recovered held-out probabilities agreed with the saved original values.
The 435 Jaccard coefficients compare overlapping, dependent fits and are not replicates
for confidence-interval estimation. Feature sampling is not a label-permutation null.

One panel's sigmoid probabilities saturated to exactly 1 for all 64 GSE43292 tissues.
The saved unsquashed linear scores retained 64 distinct values. We therefore compute
AUROC, average precision and paired ordering uniformly from the linear logit for all
new logistic representation controls. Probabilities remain the inputs to Brier scores
and threshold diagnostics. The original primary outputs did not change. For this
panel (random 18), logit AUROC was 0.7207 and paired concordance was 29/32; a probability
implementation would have introduced artificial ties. Both score forms are archived.

\subsection*{Threshold recipes and prior pilot exposure}
Before the formal exploratory record, a pilot used each group's average of ten held-out
source probabilities. Its maximum-balanced-accuracy midpoint was 0.2251206725 and
produced external balanced accuracies 0.5000 and 0.53125. This prior observation is
disclosed and is not replaced by a claim that the later analyses were blind.
The reported full-source recipes use one score per group from the original first
three-fold repetition, rather than averaging probabilities from ten fitted models.
Candidate thresholds are 0, 1, and the midpoints between consecutive distinct scores;
classification uses score $\geq$ threshold. Maximum Youden $J$ is selected with an
absolute tie tolerance of $10^{-12}$, preferring the candidate closest to 0.5 and then
the lower value. The sensitivity rule chooses the largest candidate with source
sensitivity at least 0.90. The first-repetition source split seed is 20261025.

For anatomical holdouts, three-fold inner cross-fitting is stratified by the four
available bed--status cells, using seeds 20261026, 20261027 and 20261028 for the
carotid, femoral and infrapopliteal targets. Gene selection, scaling and fitting occur
inside each inner training fold. These nine inner fits choose thresholds; three outer
fits on the remaining beds generate held-out-bed probabilities. Thus target outcomes
do not choose the thresholds. Source selection quantities are optimization results,
not unbiased performance estimates. Target-label threshold curves are descriptive
only. Pooling cross-fitted source scores to choose a threshold does not alter the
primary within-fold AUROC estimand and can remain sensitive to fold-to-fold scale.

\subsection*{Score arithmetic, failure maps and display summaries}
The frozen score is the intercept plus the sum of its 200 signed standardized gene
contributions. Source values from this full-source model are in-sample references.
Overall means depend on class proportions, so class-specific means are also supplied.
The fraction of feature values with $|z|>3$ is a scale-displacement diagnostic,
not a patient-level error rate or a validated out-of-distribution detector.
Named display features are selected by source coefficients rather than target labels.
Failure maps include all 32 known patient pairs and all five expression rules.
Delete-one-patient and delete-one-specimen calculations keep predictions fixed;
they are influence analyses, not refitting, new validation or uncertainty intervals.
Displays of the three reversed primary pairs are explicitly selected after observing
their failures and give arithmetic explanations rather than causal mechanisms.
Supplementary Figures S1--S2 show the original nine-cell fold values and repeated
source-group probabilities; repeated partitions are dependent.

\setcounter{table}{8}
\begin{longtable}{lrrrrrr}
\caption{All exploratory representation controls. Source values are means of the
same 30 held-out folds. External values use full-source fits and linear-logit ordering.
The primary row is the original reference model. All 20 random draws are retained.}
\label{tab:randompanels}\\
\toprule
Panel & Source AUC & \multicolumn{3}{c}{GSE43292} & \multicolumn{2}{c}{GSE28829}\\
\cmidrule(lr){3-5}\cmidrule(lr){6-7}
 & & AUC & Pair concord. & BA & AUC & BA\\
\midrule\endfirsthead
\toprule Panel & Source AUC & AUC & Pair concord. & BA & AUC & BA\\
\midrule\endhead
\bottomrule\endfoot
''']
pc=((pairs.difference>0).sum()+.5*(pairs.difference==0).sum())/len(pairs)
vals=[primary.auc.mean(),pe.loc['GSE43292','auc'],pc,pe.loc['GSE43292','balanced_accuracy'],pe.loc['GSE28829','auc'],pe.loc['GSE28829','balanced_accuracy']]
text.append('Primary & '+' & '.join(num(x) for x in vals)+r'\\'+'\n')
for panel in ['next200']+[f'random_{i:02d}' for i in range(1,21)]:
    ee=ext[ext.panel==panel].set_index('dataset')
    vals=[fold.loc[fold.panel==panel,'auc'].mean(),ee.loc['GSE43292','auc'],ee.loc['GSE43292','pair_concordance'],ee.loc['GSE43292','balanced_accuracy'],ee.loc['GSE28829','auc'],ee.loc['GSE28829','balanced_accuracy']]
    label='Ranks 201--400' if panel=='next200' else panel.replace('random_','Random ')
    text.append(label+' & '+' & '.join(num(x) for x in vals)+r'\\'+'\n')
text.append(r'''\end{longtable}
Pair concordance compares plaque with relatively intact tissue within each of 32
known patients; marginal AUC compares all tissues across classes. BA is balanced
accuracy at the technical probability threshold 0.5. No draw ranges are confidence
intervals, and near-ceiling source performance does not identify a unique panel.

\clearpage
\begin{longtable}{llrrrrrr}
\caption{Evaluation of source-only threshold rules. Each anatomical holdout uses
nested source-only threshold selection; external cohorts use unchanged full-source
thresholds. The table reports every evaluated rule, not a target-selected winner.}
\label{tab:thresholdrecipes}\\
\toprule Target & Rule & Threshold & TP & FN & TN/FP & Sens. & BA\\
\midrule\endfirsthead
\toprule Target & Rule & Threshold & TP & FN & TN/FP & Sens. & BA\\
\midrule\endhead
\bottomrule\endfoot
''')
labels={'LOBO_Carotid':'Carotid','LOBO_Femoral':'Femoral','LOBO_Infrapopliteal':'Infrapopliteal'}
rulelabels={'fixed_0.5':'Fixed 0.5','source_max_youden':'Max. J','source_sensitivity_0.90':'Sens. 0.90'}
for row in ev.itertuples():
    vals=[labels.get(row.dataset,row.dataset),rulelabels[row.rule],num(row.threshold),int(row.tp),int(row.fn),f'{int(row.tn)}/{int(row.fp)}',num(row.sensitivity,3),num(row.balanced_accuracy,3)]
    text.append(' & '.join(esc(x) for x in vals)+r'\\'+'\n')
text.append(r'''\end{longtable}
TP, FN, TN and FP denote true positives, false negatives, true negatives and false
positives. Sens. is sensitivity, BA is balanced accuracy; specificity can be
recovered as TN/(TN+FP). These labels concern sampled tissue contrasts, not patient
risk. The full candidate curves and all 12 learning-context/rule records, including
source optimization quantities, are retained in the accompanying CSV files.

\subsection*{Reproduction and complete tabular outputs}
\texttt{results/exploratory\_features/} contains panel memberships, all source and
external predictions, fold metrics, all 6,000 primary selections, stability values,
and 435 Jaccard comparisons. \texttt{results/exploratory\_threshold/} contains
all threshold candidates/rules/evaluations, class-specific logit accounting,
37,600 gene contributions, all paired and specimen deletion values, and failure maps.
\texttt{results/exploratory\_display/} contains repeated source-score summaries.
No per-feature contribution row is an independent biological replicate.

\begingroup\raggedright
Starting from the saved inputs, \texttt{code/explore\_feature\_dependence.py} and
\texttt{code/explore\_threshold\_shift.py} rebuild the new fitted analyses.
\texttt{code/validate\_exploratory.py} independently recomputes metrics and performs
limited independent fitting checks. The three \texttt{plot\_*\_extension.py} scripts
render Figures 7--11 and S1--S2. Then run \texttt{assemble\_figure\_catalog.py},
\texttt{build\_exploratory\_supplement.py}, and \texttt{assemble.py --compile}.
These commands are run from the project root with the \texttt{code/} prefix.
The archive README gives the ordered full workflow, original analysis commands,
dependencies, and the limits of the relocated smoke check.\par\endgroup
''')
import re
source=''.join(text)
# Hyperref path formatting permits line breaks in long reproducibility filenames.
source=re.sub(r'\\texttt\{([^{}]+)\}',lambda m:r'\path{'+m.group(1).replace(r'\_','_')+'}' if ('/' in m.group(1) or r'\_' in m.group(1)) else m.group(0),source)
out=ROOT/'manuscript/exploratory_supplement.tex';out.write_text(source,encoding='utf8')
audit={'created_utc':datetime.datetime.now(datetime.timezone.utc).isoformat(),'status':'Saved-result table generation; no fitting','table_ids':['S9','S10'],'panel_rows':22,'threshold_evaluation_rows':len(ev),'input_sha256':{str(p.relative_to(ROOT)).replace('\\','/'):hashlib.sha256(p.read_bytes()).hexdigest() for p in used},'output_sha256':hashlib.sha256(out.read_bytes()).hexdigest()}
(ROOT/'evidence/figure_revision_20261026/exploratory_supplement_audit.json').write_text(json.dumps(audit,indent=2),encoding='utf8')
print(json.dumps(audit,indent=2))
```
