## Supplementary material for "Anatomical comparison populations and threshold transport in vascular transcriptomic classification": Reproducibility archive: code, provenance and results: build_supplement.py

```
"""Build Supplementary Tables S1-S8 from saved outputs; no model fitting."""
from pathlib import Path
from datetime import datetime, timezone
import hashlib
import importlib.metadata
import json
import platform
import re
import sys
import numpy as np
import pandas as pd

ROOT = Path(__file__).resolve().parents[1]
OUT = ROOT / 'manuscript'
OUT.mkdir(exist_ok=True)
R = ROOT / 'results'
E = ROOT / 'evidence'
used = []

def read_csv(name):
    path = R / name
    used.append(path)
    return pd.read_csv(path)

def read_json(path):
    used.append(path)
    return json.loads(path.read_text(encoding='utf-8'))

def esc(value):
    return ''.join({'\\': r'\textbackslash{}', '&': r'\&', '%': r'\%', '$': r'\$',
                    '#': r'\#', '_': r'\_', '{': r'\{', '}': r'\}', '~': r'\textasciitilde{}',
                    '^': r'\textasciicircum{}'}.get(c, c) for c in str(value))

class Raw(str):
    pass

def cell(value):
    return str(value) if isinstance(value, Raw) else esc(value)

def num(value, digits=3):
    if pd.isna(value):
        return '--'
    value = 0 if abs(float(value)) < 0.5 * 10 ** (-digits) else float(value)
    return f'{value:.{digits}f}'

def interval(row, key, digits=3):
    return f"{num(row[key], digits)} [{num(row[key+'_low'], digits)}, {num(row[key+'_high'], digits)}]"

parts = [r'''% Generated by code/build_supplement.py. Required packages: longtable, booktabs,
% array, pdflscape, hyperref, amsmath. No standalone preamble or document environment.
\renewcommand{\thetable}{S\arabic{table}}
\section*{Supplementary methods and results}
This supplement reports the saved outputs of a retrospective computational tissue-classification
audit. Counts refer to arrays, conservative specimen-label groups, or explicitly paired patients as
specified; they must not be summed into a total of independent patients. AUROC denotes area
under the receiver-operating-characteristic curve; AP denotes the non-interpolated average
precision calculated by \texttt{average\_precision\_score}, not a trapezoidal precision--recall area.
BA denotes balanced accuracy. The internal repeat ranges describe partition sensitivity and are
not confidence intervals. External intervals condition on the fitted source model and fixed scores.
The probability threshold of 0.5 and Brier scores are technical diagnostics of selected tissue
contrasts, not population-risk calibration or validated clinical decision thresholds.
''']

in_landscape = False
table_instance = 0

def set_orientation(landscape):
    global in_landscape
    if landscape != in_landscape:
        parts.append(r'\begin{landscape}' if landscape else r'\end{landscape}')
        in_landscape = landscape

def table(number, title, headers, rows, widths, foot='', landscape=False, label=None):
    global parts, table_instance
    table_instance += 1
    # Keep related S1-S7 tables in one landscape section; no page per table.
    landscape = landscape or number <= 7
    set_orientation(landscape)
    # Start the external-results group together, avoiding a two-row orphan.
    if number == 5 and label is not None:
        parts.append(r'\clearpage')
    parts.append(r'\begingroup\footnotesize\setlength{\tabcolsep}{2pt}\renewcommand{\arraystretch}{1.15}')
    parts.append(r'\setcounter{table}{' + str(number-1) + '}')
    parts.append(r'\renewcommand{\theHtable}{supplement.' + str(table_instance) + '}')
    spec = '@{}' + ''.join(r'>{\raggedright\arraybackslash}p{' + f'{w:.3f}' + r'\linewidth}' for w in widths) + '@{}'
    parts.append(r'\begin{longtable}{' + spec + '}')
    cap = r'\caption{' + esc(title) + '}'
    if label:
        cap += r'\label{' + label + '}'
    parts.append(cap + r'\\')
    heading = ' & '.join(cell(x) for x in headers) + r' \\'
    parts += [r'\toprule', heading, r'\midrule\endfirsthead',
              r'\caption[]{' + esc(title if title.endswith('(continued)') else title + ' (continued)') + r'}\\',
              r'\toprule', heading, r'\midrule\endhead', r'\midrule',
              r'\multicolumn{' + str(len(headers)) + r'}{r}{Continued on next page}\\\endfoot',
              r'\bottomrule\endlastfoot']
    for row in rows:
        assert len(row) == len(headers), (number, row)
        parts.append(' & '.join(cell(x) for x in row) + r' \\')
    parts.append(r'\end{longtable}\endgroup')
    if foot:
        parts.append(r'{\footnotesize ' + foot + r'\par}')
    parts.append(r'\par\medskip')

registry = read_json(E / 'dataset_registry.json')
qc = read_json(E / 'preprocessing_qc.json')
acquisition = read_json(E / 'acquisition.json')
datasets = []
for row in registry:
    acc = row['accession']
    datasets.append([Raw(r'\href{' + row['official_source'] + '}{' + acc + '}'),
                     row['arrays'], row['independent_patients_or_donors'],
                     row['disease_definition'] + '; comparator: ' + row['comparator_definition'],
                     row['platform'], row['role']])
table(1, 'Dataset units, contrasts and analytical roles',
      ['Dataset', 'Arrays', 'Independent-patient status', 'Tissue contrast', 'Platform', 'Role'],
      datasets, [.09,.05,.18,.27,.13,.20],
      foot=r'GSE100927 contains 69 disease and 35 control arrays. Conservative full-label aggregation produces '
           r'65 disease and 30 control groups: carotid 27/10, femoral 25/10, infrapopliteal 13/10. '
           r'The original report states 65 disease and 31 healthy arteries; the femoral-control discrepancy remains unresolved. '
           r'GSE43292 comprises 32 plaque/intact pairs. GSE28829 comprises 16 advanced and 13 early lesions. '
           r'None of these endpoints has a follow-up time or censoring variable.', landscape=True, label='tab:s1')

provenance_rows = []
for row in registry:
    acc=row['accession']; q=qc[acc]
    digest=Raw(r'\shortstack[l]{'+r'\\'.join(r'\texttt{'+row['sha256'][i:i+16]+'}' for i in range(0,64,16))+'}')
    provenance_rows.append([acc, row['download_completed_utc'].replace('T',' ').replace('+00:00',' UTC'),
                            f"{int(row['bytes']):,}", digest,
                            f"{q['matrix_probes']:,} / {q['matched_probes']:,} / {q['unique_genes']:,}",
                            row['inherited_preprocessing']])
table(1, 'Dataset provenance and preprocessing (continued)',
      ['Dataset', 'Download completion', 'Bytes', 'SHA256 (four consecutive blocks)',
       'Matrix probes / matched probes / genes', 'Inherited preprocessing'],
      provenance_rows,[.09,.15,.08,.18,.16,.26], landscape=True,
      foot=r'The common annotation-defined vocabulary contains 15,235 genes. All matrices had zero missing probe values, '
           r'and none required the originally specified log transformation. Source-normalization procedures used whole deposited cohorts; '
           r'training-only downstream fitting cannot remove that inherited transductive aspect. Exact URLs and downloaded-file hashes are in '
           r'\path{evidence/acquisition.json}. GEO permits unrestricted download but does not certify absence of submitter intellectual-property claims '
           r'(\url{https://www.ncbi.nlm.nih.gov/geo/info/disclaimer.html}). Source ethics approvals and consent statements refer to the original studies. '
           r'Public availability does not itself create a new ethics approval or exemption for the present analysis.')

pairs = read_csv('ambiguous_label_profile_similarity.csv')
metadata = read_json(E / 'GSE100927_sample_metadata.json')
sex = {r['geo_accession']:r['characteristics_ch1_2'].split(': ')[-1] for r in metadata}
pair_rows=[]
for _,r in pairs.iterrows():
    pa=r.title_a.split(':')[0];pb=r.title_b.split(':')[0]
    same=re.search(r'_(\d+)',pa).group(1)==re.search(r'_(\d+)',pb).group(1)
    pair_rows.append([r.label_group,r.array_a,pa,r.array_b,pb,sex[r.array_a],num(r.rank_profile_correlation,5),'Yes' if same else 'No'])
table(2,'All nine repeated specimen-label pairs and profile similarity',
      ['Full label after colon','Array A','Source prefix A','Array B','Source prefix B','Sex','Rank-profile r','Same numeric prefix'],
      pair_rows,[.22,.10,.10,.10,.10,.06,.09,.10],landscape=True,label='tab:s2',
      foot=r'Correlations are Pearson correlations between sample-wise fractional-rank vectors over the 15,235 common genes. '
           r'High similarity does not authenticate donor identity or establish technical replication. Four pairs share a numeric '
           r'pre-colon component with a b suffix in one array; five share only the post-colon specimen label. '
           r'The original metadata label \texttt{Femoral artery\_41G} is retained. All 18 arrays belonging to these nine pairs '
           r'are excluded in the discard-ambiguous sensitivity, leaving 86 arrays.')

primary=read_csv('primary_fold_metrics.csv')
model_order=['prevalence','bed_sex','C1Q','Wu6','ridge200','centroid200','forest200']
model_names={'prevalence':'Prevalence','bed_sex':'Bed + sex','C1Q':'C1Q','Wu6':'Wu6','ridge200':'Ridge-200','centroid200':'Centroid-200','forest200':'Forest-200'}
metric_keys=['auc','ap','within_bed_auc','delta_mix','mix_span','balanced_accuracy','sensitivity','specificity','brier']
repeat_means=primary.groupby(['model','repeat'])[metric_keys].mean()
summary_rows=[]
for model in model_order:
    for stat,func in [('Mean','mean'),('Min','min'),('Max','max')]:
        values=getattr(repeat_means.loc[model],func)()
        summary_rows.append([model_names[model] if stat=='Mean' else '',stat]+[num(values[k]) for k in metric_keys])
table(3,'All models: mean and range of repetition-level mean fold statistics',
      ['Model','Statistic','AUROC','AP','Within-bed AUROC',Raw(r'$\Delta_{\rm mix}$'),'Mix span','BA','Sensitivity','Specificity','Brier'],
      summary_rows,[.12,.06,.075,.075,.09,.075,.075,.075,.075,.075,.075],landscape=True,label='tab:s3',
      foot=r'Each repetition first averages its three held-out folds equally; this table then summarizes the ten repetition means. '
           r'Minimum and maximum are split-sensitivity ranges, not confidence limits. Within-bed AUROC is the equal mean of the three diagonal '
           r'case-bed/control-bed cells. $\Delta_{\rm mix}$ equals pooled minus within-bed AUROC. Mix span is the maximum minus minimum '
           r'over the independently varied case/control bed-proportion grid. C1Q and centroid are uncalibrated ranking scores, '
           r'so threshold and Brier metrics are inapplicable. The retrospective literature panels were informed by these public cohorts '
           r'and are not independent biomarker confirmations.')

transport=read_csv('transport_metrics.csv')
tr=transport[transport.model=='ridge200']
assert len(tr)==18
tr_rows=[]
for _,r in tr.iterrows():
    tr_rows.append(['Other two' if r.source=='other_two' else r.source,r.target,'Yes' if r.purge_possible_controls else 'No',
                    int(r.n_train),int(r.n_removed),int(r.n_test)]+[num(r[k]) for k in ['auc','ap','balanced_accuracy','sensitivity','specificity','brier']])
table(4,'All 18 primary-model anatomical transport settings',
      ['Training bed(s)','Held-out bed','Proxy purge','Train n','Removed n','Test n','AUROC','AP','BA','Sensitivity','Specificity','Brier'],
      tr_rows,[.115,.115,.06,.05,.06,.05,.07,.07,.07,.07,.07,.07],landscape=True,label='tab:s4',
      foot=r'Counts denote conservative label groups. Proxy purging removes source controls with the same control-number suffix and sex '
           r'as a target control; it is a sensitivity scenario for unresolved shared-donor identity, not confirmed donor linkage. '
           r'Each row fits only its stated training data, including feature selection and scaling. No target recalibration is applied. '
           r'The 12 additional unpurged leave-one-bed-out comparator rows are retained in \path{results/transport_metrics.csv}.')

external=read_csv('external_metrics.csv')
external_pairs=read_csv('external_pairs.csv')
ranking=[]
for _,r in external.iterrows():
    ranking.append([r.dataset,model_names[r.model],int(r.n),interval(r,'auc'),interval(r,'ap')])
table(5,'External ranking metrics for five expression rules (four source-fitted; C1Q fit-free)',
      ['Dataset','Model','Tissue arrays','AUROC [95% interval]','AP [95% interval]'],ranking,[.15,.18,.10,.25,.25],label='tab:s5',
      foot=r'Intervals use 2,000 fixed-score bootstrap resamples (seed 20261025). GSE43292 is resampled by patient, retaining both tissues; '
           r'GSE28829 is resampled by specimen within lesion class because donor IDs are unavailable. Intervals exclude source-training, '
           r'model-choice, annotation and unresolved donor-dependence uncertainty. A boundary interval of [1,1] reflects the finite-sample '
           r'bootstrap distribution, not certainty about population performance.')
pair_result=[]
for _,r in external[external.dataset=='GSE43292'].iterrows():
    ds=external_pairs[external_pairs.model==r.model].difference
    pair_result.append([model_names[r.model],f'{int((ds>0).sum())}/32',int((ds==0).sum()),
                        f"{num(r.pair_concordance)} [{num(r.pair_exact_low)}, {num(r.pair_exact_high)}]",
                        interval(r,'pair_concordance'),interval(r,'paired_mean_difference',5)])
table(5,'Paired GSE43292 contrasts (continued)',
      ['Model','Plaque > intact','Ties','Concordance [exact 95% CI]','Concordance [bootstrap 95% interval]',
       'Mean paired score difference [bootstrap 95% interval]'],pair_result,[.12,.12,.06,.20,.20,.24],landscape=True,
      foot=r'Concordance assigns one to a higher plaque score and one-half to a tie within the same patient. '
           r"The exact interval is Clopper--Pearson because all five models have zero ties. Score differences use each model's "
           r'native score scale and cannot be compared as calibrated biological effect sizes. This within-patient estimand differs '
           r'from marginal AUROC, which compares all positive and negative tissues.')
prob_rows=[]
for _,r in external[external.model.isin(['Wu6','ridge200','forest200'])].iterrows():
    prob_rows.append([r.dataset,model_names[r.model]]+[interval(r,k) for k in ['balanced_accuracy','sensitivity','specificity','brier']])
table(5,'External probability-output diagnostics at the fixed 0.5 threshold (continued)',
      ['Dataset','Model','BA [95% interval]','Sensitivity [95% interval]','Specificity [95% interval]','Brier [95% interval]'],
      prob_rows,[.12,.14,.17,.17,.17,.17],landscape=True,
      foot=r'Only models with probability outputs are shown. These class-balanced classifier outputs were not recalibrated externally. '
           r'Good ranking can coexist with failure of a fixed threshold. Brier scores here describe selected histological contrasts; '
           r'they do not estimate calibration for population disease or future clinical events.')

sensitivity=read_csv('sensitivity_all_fold_metrics.csv')
validation=read_json(E/'independent_saved_results_validation.json')
scenarios=['top50','top500','C001','C1','native','exclude_ambiguous','104_grouped_arrays','possible_shared_controls']
scenario_names={'top50':'Top 50 genes','top500':'Top 500 genes','C001':'C = 0.01','C1':'C = 1',
                'native':'Native processed values','exclude_ambiguous':'Exclude ambiguous arrays',
                '104_grouped_arrays':'104 arrays, grouped folds','possible_shared_controls':'Possible shared controls'}
sens_rows=[]
for scenario in scenarios:
    g=sensitivity[sensitivity.scenario==scenario]
    means=g.groupby('repeat')[metric_keys].mean();mu=means.mean()
    complete=validation['checks']['independent_fold_recomputation'][scenario]['complete_cell_records']
    sens_rows.append([scenario_names[scenario],int(g.units.iloc[0]),f'{complete}/30',num(mu.auc),
                     f'{num(means.auc.min())}--{num(means.auc.max())}']+[num(mu[k]) for k in ['ap','within_bed_auc','delta_mix','mix_span','balanced_accuracy','brier']])
table(6,'All eight prespecified primary-model sensitivities',
      ['Scenario','Units','Complete folds','Mean AUROC','AUROC repeat range','Mean AP','Within-bed AUROC',Raw(r'$\Delta_{\rm mix}$'),'Mix span','BA','Brier'],
      sens_rows,[.16,.045,.065,.07,.115,.07,.085,.07,.07,.07,.07],landscape=True,label='tab:s6',
      foot=r'Other specifications remain fixed. Means average the three folds within each repetition and then ten repetitions. '
           r'Grouped-array and possible-control scenarios use stratified group folds; all 30 test folds in every scenario retained '
           r'the six bed-by-status strata needed for the nine-cell decomposition. Units are arrays in the 104-array and exclusion '
           r'scenarios and conservative labels otherwise. These results are sensitivity analyses, not independent studies.')

nulls=read_csv('permutation_nulls.csv');sub=read_csv('full_pipeline_subsampling.csv')
observed=primary[(primary.model=='ridge200')&(primary.repeat==0)].auc.mean()
null_rows=[]
for scheme,g in nulls.groupby('scheme',sort=False):
    exceed=int((g.mean_fold_auc>=observed).sum())
    null_rows.append([scheme.replace('_',' '),len(g),num(observed),num(g.mean_fold_auc.mean()),num(g.mean_fold_auc.min()),
                     num(g.mean_fold_auc.max()),exceed,num((1+exceed)/(1+len(g)))])
table(7,'Full-pipeline label-permutation controls',
      ['Scheme','Permutations','Observed first-repeat AUROC','Null mean','Null min','Null max','Null >= observed','Plus-one tail fraction'],
      null_rows,[.15,.09,.17,.10,.10,.10,.10,.12],landscape=True,label='tab:s7',
      foot=r'Each scheme has 99 permutations at the 95-label unit. Within-bed permutations preserve class counts by bed; unrestricted '
           r'permutations do not. Every permutation regenerates stratified folds and repeats feature selection, source scaling and fitting. '
           r'The plus-one tail fraction is $(1+\#\{A_{\rm null}\ge A_{\rm observed}\})/(99+1)$; its minimum resolution is 0.01. '
           r'This is a pipeline-null diagnostic, not a significance test of anatomical transport or clinical usefulness.')
sub_keys=['auc','within_bed_auc','delta_mix','mix_min','mix_max','mix_span','cross_bed_pair_fraction']
sub_rows=[[key.replace('_',' '),num(sub[key].mean()),num(sub[key].min()),num(sub[key].max())] for key in sub_keys]
table(7,'Thirty complete-pipeline subsamples (continued)',
      ['Statistic','Mean','Minimum','Maximum'],sub_rows,[.43,.16,.16,.16],
      foot=r'Every subsample selects without replacement the floor of 80\% of each of the six bed-by-status groups: '
           r'75 conservative label groups in every run. The full three-fold primary procedure is refitted. Ranges are stability '
           r'descriptions, not confidence intervals. Repeated partitions and subsamples share observations.')

set_orientation(False)
# Place compact computational methods with the title/intro before the wide tables.
parts.insert(1,r'''\begingroup\small
\section*{Exact computational specification}
For each platform, only probes mapping to one unambiguous gene symbol were retained; multiple probes
for a gene were combined by the median within each array. The common vocabulary was the intersection
of the three platform mappings and available probe identifiers, without use of target expression
distributions or target outcomes. Fractional ranks used average ranks for ties, divided by 15,235.
Ranks were computed within each array before the median over repeated full specimen labels.
No missing ranks occurred. Native-expression sensitivity used the already processed deposited values
unchanged. No joint cohort normalization, target-derived scaling, ComBat correction, external
threshold selection or external recalibration was performed by the present pipeline.

The executed training-only feature score for gene $g$ was
\[
 T_g=\frac{|\bar{x}_{1g}-\bar{x}_{0g}|}
 {\sqrt{s^2_{1g}/n_1+s^2_{0g}/n_0+10^{-12}}},
\]
with unbiased within-class variances. The largest 200 scores were selected using a stable sort
over the alphabetically ordered vocabulary. This is a Welch-type statistic, not Cohen's standardized
mean difference. Training means and population standard deviations were then fitted by
\texttt{StandardScaler}; the same fitted transformation was applied to held-out samples.

For case bed $a$ and control bed $b$, $A_{ab}$ is the proportion of score pairs with the case higher,
with one-half credit for ties. Within each held-out fold, with $n_{1a}$ cases and $n_{0b}$ controls,
\[
 A_{\rm pooled}=\frac{\sum_{a,b} n_{1a}n_{0b}A_{ab}}{\sum_{a,b}n_{1a}n_{0b}},\qquad
 A_{\rm within}=\frac{1}{3}\sum_a A_{aa},\qquad
 \Delta_{\rm mix}=A_{\rm pooled}-A_{\rm within}.
\]
The identity is checked against the directly calculated fold AUROC. Case and control bed proportions
were varied independently over the 66-point simplex grid in steps of 0.1 (4,356 combinations).
Because this grid includes simplex vertices, its extrema equal the minimum and maximum of the
nine observed pair-cell AUROCs; the grid is an algebraic reweighting, not a validation cohort.
Scores from separately fitted folds were never pooled for this decomposition.
\par\endgroup
''')

settings=[
 ['Prevalence','Constant training prevalence; no expression features.'],
 ['Bed + sex','Three fixed bed indicators and one male indicator; training scaling and class-balanced L2 logistic regression, C=0.1.'],
 ['C1Q','Mean of C1QA and C1QC fractional ranks; no fitting or probability conversion. Retrospective literature control.'],
 ['Wu6','ADAP2, CSF3R, FABP5, ITGAX, MYOC, SPP1; all six are in the common vocabulary; scaled class-balanced L2 logistic regression, C=0.1.'],
 ['Ridge-200','Top 200 Welch-type features, StandardScaler, class-balanced L2 logistic regression; solver=liblinear, C=0.1, max_iter=2000.'],
 ['Centroid-200','Same top-200 training-selected features and scaling. Score = mean of standardized features multiplied by the sign of the training case-control mean difference.'],
 ['Forest-200','Same top-200 features and scaling; 300 trees, max_depth=3, min_samples_leaf=3, max_features=sqrt, class_weight=balanced, n_jobs=2.'],
 ['Repeated validation','Three folds stratified by bed and status; ten repeats; random_state=20261025+r, r=0,...,9. Fixed model specifications; no outcome-driven hyperparameter search.'],
 ['Grouped sensitivities','StratifiedGroupKFold with the same repeat seeds; groups are full specimen labels or the explicit control-suffix-plus-sex uncertainty proxy.'],
 ['Frozen external models','Fit each specified model to all 95 source label groups; random_state=20261025. No target adaptation.'],
 ['External uncertainty','2,000 bootstrap resamples per model/cohort; seed=20261025. Complete patient blocks for GSE43292; class-stratified specimen resampling for GSE28829.'],
 ['Null and subset randomness','NumPy default_rng(20261116), shared sequentially through 99 within-bed permutations, 99 unrestricted permutations, then 30 stratified subsamples. Null fold seed=20261025; subset fold seed=20261025+iteration.'],
]
table(8,'Model recipes, seeds and reproducibility specification',['Component','Executed specification'],settings,[.23,.70],label='tab:s8',
      foot=r'All hyperparameters are fixed specifications, not results of nested tuning. The 200-gene final source model is reconstructed '
           r'from \path{results/frozen_primary_coefficients.csv} and \path{results/frozen_primary_model.json}; the former contains gene, '
           r'coefficient, source mean and source standard deviation for every selected gene. The fitted intercept is 0.28075375142531145. '
           r'These coefficients permit score reconstruction but are not interpreted as causal gene effects.')

parts.append(r'''For the frozen primary model, the reproducible score is
\[
 p(x)=\operatorname{logit}^{-1}\left(0.28075375142531145+
 \sum_{g=1}^{200}\beta_g\frac{x_g-\mu_g}{\sigma_g}\right).
\]
The CSV preserves full numerical precision and the order of selected genes; duplicating this
200-row machine-readable table in the PDF is unnecessary. The source-only classifier probabilities
must not be interpreted as population disease probabilities.

\subsection*{Protocol chronology and disclosure of prior exposure}
The internally time-stamped protocol was recorded on 25 October 2026 before new-study expression
model fitting; it was not preregistration. Metadata, original papers and several previously published
results were already known. GSE43292 had previously supported descriptive module contrasts in a
separate VascFlux-GEO manuscript; the present work does not claim that this public cohort was
previously untouched. The current endpoint, fitted models, anatomical decomposition and transport
results were newly calculated here.

The pre-fit amendment, after outcome-agnostic preprocessing QC, specified that probability diagnostics
apply only to probability-output models, retained deposited native values without transformation,
added sex to the conservative control-linkage proxy, confirmed the 15,235-gene vocabulary, fixed
rank-before-label-aggregation ordering, and clarified interval interpretation. After main and robustness
results were observed, an implementation note corrected the feature-ranking terminology to the executed
Welch-type statistic. It also narrowed decomposition error handling so a failed algebraic identity
raises an error, while a missing stratum is explicitly recorded. These changes did not alter saved
predictions or select a model on external outcomes. The saved-result audit independently verified all
complete-fold decompositions and reconstructed the primary external scores. This is an audit of
recorded computations, not independent clinical validation or proof that all possible biases are absent.
''')

env=dict(python=platform.python_version(),packages={x:importlib.metadata.version(x) for x in ['numpy','pandas','scipy','scikit-learn']})
(E/'supplement_environment_snapshot.json').write_text(json.dumps(env,indent=2),encoding='utf-8')
files=[
 ['S1','evidence/dataset_registry.csv','Dataset units, official sources, usage conditions, ethical boundaries, downloads and hashes.'],
 ['S1','evidence/acquisition.json','Complete matrix and extracted platform-table provenance; full huge platform families were not downloaded.'],
 ['S1','evidence/preprocessing_qc.json','Probe mapping, expression ranges, missingness and common-gene count.'],
 ['S2','results/ambiguous_label_profile_similarity.csv','All nine pair correlations and exact labels/source prefixes.'],
 ['S2','results/unmatched_within_stratum_similarity.csv','Nonmatching-label profile similarities within bed and status.'],
 ['S3','results/primary_fold_metrics.csv','All 210 fold/model records.'],
 ['S3','results/primary_predictions.csv; results/primary_pair_cells.csv','Held-out scores/partitions and all nine comparison cells per fold/model.'],
 ['S4','results/transport_metrics.csv; results/transport_predictions.csv','All 30 transport metric rows, including the 18 primary-model settings, with predictions.'],
 ['S5','results/external_metrics.csv; results/external_predictions.csv; results/external_pairs.csv','Ten cohort/model summaries, all individual scores and 160 paired score contrasts.'],
 ['S6','results/sensitivity_all_fold_metrics.csv','240 fold records across eight specifications; corresponding scenario predictions and pair-cell tables are also retained.'],
 ['S7','results/permutation_nulls.csv; results/full_pipeline_subsampling.csv','198 complete-pipeline null statistics and 30 subsampling records.'],
 ['S8','results/frozen_primary_coefficients.csv; results/frozen_primary_model.json','All 200 coefficients, training scaling and intercept for the frozen primary model.'],
 ['S8','protocol/analysis_protocol_v1.txt; protocol/amendment_01_before_fit.txt; protocol/implementation_note_02.txt','Internal chronology, pre-fit clarification and post-result terminology/error-handling note.'],
 ['S8','evidence/protocol_freeze_sha256.json; evidence/amendment_01_sha256.json','Protocol hashes; not registration certificates.'],
 ['S8','evidence/independent_saved_results_validation.json','Saved-prediction metric/decomposition checks and primary-score reconstruction.'],
 ['S8','code/acquire.py; code/prepare.py; code/analyze.py; code/validate_results.py','Acquisition, deterministic preparation, fitting/validation and independent saved-result checks.'],
 ['S8','code/plot_figures.py; code/build_supplement.py','Programmatic figure and supplementary-table generation.'],
]
file_rows=[[a,Raw(r'\path{'+b+'}'),c] for a,b,c in files]
table(8,'Machine-readable result and code index (continued)',['Table','Relative file path','Contents'],file_rows,[.06,.47,.40],landscape=True)
set_orientation(False)
parts.append(r'\subsection*{Execution environment and commands}')
parts.append(esc('Snapshot: Python '+env['python']+'; '+', '.join(k+' '+v for k,v in env['packages'].items()))+'.')
parts.append(r'''The working directory is the project root. The supported analysis sequence is:
\begin{verbatim}
python -m pip install -r requirements.txt
python code/acquire.py
python code/metadata.py
python code/prepare.py
python code/analyze.py main
python code/analyze.py robust
python code/validate_results.py
python code/export_descriptive_scores.py
python code/plot_figures.py
python code/build_supplement.py
python code/assemble.py --compile
\end{verbatim}
Acquisition is unnecessary when the verified source files are already present. Figure generation
and supplementary assembly do not refit the models. Official processed matrix files, data hashes, fitted-source
parameters, saved predictions, protocol records and dependency snapshots should accompany any
reproducibility release. Public source data remain subject to their original repository conditions.
''')

set_orientation(False)
addendum = OUT / 'statistical_addendum.tex'
if addendum.exists():
    used.append(addendum)
    parts.append(r'\begingroup\small' + '\n' + addendum.read_text(encoding='utf-8').replace(r'\clearpage', '', 1) + '\n' + r'\par\endgroup')
# Keep the protocol chronology in the final continuous portrait-methods section.
for i, block in enumerate(parts):
    marker = r'\subsection*{Protocol chronology and disclosure of prior exposure}'
    if marker in block:
        before, chronology = block.split(marker, 1)
        parts[i] = before
        dest = parts.index(r'\subsection*{Execution environment and commands}')
        parts.insert(dest, marker + chronology)
        break
text='\n'.join(parts)+'\n'
assert 'DECLARATION OF GENERATIVE AI' not in text
(OUT/'supplement_body.tex').write_text(text,encoding='utf-8')
manifest={str(p.relative_to(ROOT)).replace('\\','/'):hashlib.sha256(p.read_bytes()).hexdigest() for p in set(used)}
audit=dict(created_utc=datetime.now(timezone.utc).isoformat(),output='manuscript/supplement_body.tex',
           output_sha256=hashlib.sha256(text.encode()).hexdigest(),input_sha256=manifest,
           table_groups=list(range(1,9)),primary_models=7,transport_ridge_rows=len(tr),external_model_cohort_rows=len(external),
           paired_model_rows=len(external_pairs),sensitivity_scenarios=len(scenarios),permutations=len(nulls),subsamples=len(sub),
           benchmark_note='The 4-figure/26-panel benchmark comparison is administrative manuscript QA and is intentionally excluded from scientific supplement content.',
           compilation_status='Body source authored; root agent supplies preamble and performs compilation/visual QA.')
(E/'supplement_build_audit.json').write_text(json.dumps(audit,indent=2),encoding='utf-8')
print(json.dumps({k:audit[k] for k in ['output','primary_models','transport_ridge_rows','external_model_cohort_rows','sensitivity_scenarios','permutations','subsamples']}))
```
