## Supplementary material for "Anatomical comparison populations and threshold transport in vascular transcriptomic classification": Reproducibility archive: code, provenance and results: check_paired_bootstrap.py

```
"""Check paired-bootstrap input contract and retained primary intervals; no refit."""
from pathlib import Path
import datetime
import hashlib
import json
import numpy as np
import pandas as pd
from analyze import bootstrap_metrics, load, metrics, SEED

ROOT = Path(__file__).resolve().parents[1]
OUT = ROOT / 'results'

def hashes():
    return {p.name: hashlib.sha256(p.read_bytes()).hexdigest()
            for p in sorted(OUT.iterdir()) if p.is_file()}

before = hashes()
_, meta = load('GSE43292')
pred = pd.read_csv(OUT / 'external_predictions.csv')
block = pred[(pred.dataset == 'GSE43292') & (pred.model == 'ridge200')].set_index('sample')
scores = block.loc[meta.index, 'score'].to_numpy()

def legacy_paired_bootstrap(m, s, prob, n=23, seed=SEED):
    # Literal pre-guard sampling/metric algorithm retained solely for parity check.
    rng = np.random.default_rng(seed)
    vals = []
    for _ in range(n):
        pats = m.patient.unique()
        draw = rng.choice(pats, len(pats), replace=True)
        idx = np.concatenate([np.flatnonzero(m.patient == p) for p in draw])
        r = metrics(m.y.iloc[idx], s[idx], prob)
        z = s[idx].reshape(-1, 2)
        yy = m.y.iloc[idx].to_numpy().reshape(-1, 2)
        d = z[yy == 1] - z[yy == 0]
        r['pair_concordance'] = np.mean((d > 0) + .5 * (d == 0))
        r['paired_mean_difference'] = d.mean()
        vals.append(r)
    q = pd.DataFrame(vals).quantile([.025, .975])
    return {k: (float(q.loc[.025, k]), float(q.loc[.975, k])) for k in q}

old = legacy_paired_bootstrap(meta, scores, True)
new = bootstrap_metrics(meta, scores, True, paired=True, n=23)
assert old == new, 'Input guard changed valid-bootstrap output'

malformed = {}
patient = meta.patient.iloc[0]
same_class = meta.copy()
same_class.loc[same_class.patient == patient, 'y'] = 1
missing_id = meta.copy()
missing_id.loc[missing_id.index[0], 'patient'] = np.nan
nonbinary = meta.copy()
nonbinary['y'] = nonbinary.y.astype(float)
nonbinary.loc[nonbinary.index[0], 'y'] = .5
for name, bad, values in [
        ('incomplete_pair', meta.iloc[1:].copy(), scores[1:]),
        ('two_positive_tissues', same_class, scores),
        ('missing_patient_id', missing_id, scores),
        ('nonbinary_label', nonbinary, scores)]:
    try:
        bootstrap_metrics(bad, values, True, paired=True, n=1)
    except ValueError as exc:
        malformed[name] = str(exc)
    else:
        raise AssertionError('Malformed paired input was not rejected: ' + name)

actual = bootstrap_metrics(meta, scores, True, paired=True, n=2000)
saved = pd.read_csv(OUT / 'external_metrics.csv')
saved = saved[(saved.dataset == 'GSE43292') & (saved.model == 'ridge200')].iloc[0]
differences = {k: max(abs(actual[k][0] - saved[k + '_low']),
                      abs(actual[k][1] - saved[k + '_high'])) for k in actual}
assert max(differences.values()) < 1e-12, 'Saved primary intervals changed'
assert before == hashes(), 'A numerical results file changed'
report = {
    'checked_utc': datetime.datetime.now(datetime.timezone.utc).isoformat(),
    'scope': 'Input validation and saved-score resampling only; no model training or new analysis selection.',
    'passed': True,
    'paired_patients': int(meta.patient.nunique()),
    'tissues': len(meta),
    'legacy_parity_draws': 23,
    'legacy_parity_exact': old == new,
    'saved_interval_reproduction_draws': 2000,
    'maximum_interval_error': float(max(differences.values())),
    'interval_errors': differences,
    'rejected_inputs': malformed,
    'result_hashes_unchanged': True,
    'result_file_sha256': before,
    'analysis_code_sha256': hashlib.sha256((ROOT / 'code/analyze.py').read_bytes()).hexdigest()
}
path = ROOT / 'evidence/revision05_bootstrap_checks.json'
path.write_text(json.dumps(report, indent=2), encoding='utf-8')
print(json.dumps({k: v for k, v in report.items() if k != 'result_file_sha256'}, indent=2))
```
