## Supplementary material for "Anatomical comparison populations and threshold transport in vascular transcriptomic classification": Reproducibility archive: code, provenance and results: explore_feature_dependence.py

```
"""Exploratory representation controls after the original results; no target tuning.

See protocol/exploratory_extension_04_20261026.txt. Original result files are read-only.
"""
from pathlib import Path
import datetime, hashlib, json, itertools
import numpy as np
import pandas as pd
from sklearn.preprocessing import StandardScaler
from sklearn.linear_model import LogisticRegression
from sklearn.metrics import roc_auc_score,average_precision_score
import analyze as core

ROOT=Path(__file__).resolve().parents[1]
OUT=ROOT/'results/exploratory_features';OUT.mkdir(parents=True,exist_ok=True)
E=ROOT/'evidence/figure_revision_20261026';E.mkdir(parents=True,exist_ok=True)
SEED=20261026
def sha(p):return hashlib.sha256(p.read_bytes()).hexdigest()
original={str(p.relative_to(ROOT)).replace('\\','/'):sha(p) for p in (ROOT/'results').glob('*') if p.is_file()}
def save(name, rows):
    frame=rows if isinstance(rows,pd.DataFrame) else pd.DataFrame(rows)
    frame.to_csv(OUT/name,index=False)
    return frame
def order(x,y):
    a=x.to_numpy(); y=np.asarray(y)
    d=a[y==1].mean(0)-a[y==0].mean(0)
    t=d/np.sqrt(a[y==1].var(0,ddof=1)/(y==1).sum()+a[y==0].var(0,ddof=1)/(y==0).sum()+1e-12)
    return np.argsort(-np.abs(t),kind='stable'), t
def fit_panel(x,y,genes,seed):
    scale=StandardScaler().fit(x[genes])
    model=LogisticRegression(C=.1,class_weight='balanced',solver='liblinear',max_iter=2000,random_state=seed)
    model.fit(scale.transform(x[genes]),y)
    return scale,model
def predict(fitted,x,genes):
    z=fitted[0].transform(x[genes])
    return fitted[1].predict_proba(z)[:,1],fitted[1].decision_function(z)
def panel_metrics(y,probability,linear_logit):
    # A monotone sigmoid can create artificial ties at machine precision.
    # Ranking uses the unrounded linear score; probability diagnostics remain unchanged.
    return core.metrics(y,probability)|dict(auc=roc_auc_score(y,linear_logit),
        ap=average_precision_score(y,linear_logit),ranking_score='linear_logit',
        probability_unique=len(np.unique(probability)),linear_logit_unique=len(np.unique(linear_logit)),
        saturated_probability_count=int(np.sum((probability==0)|(probability==1))))

x,m=core.load('GSE100927')
assert x.shape==(95,15235)
allgenes=x.columns.tolist()
rng=np.random.default_rng(SEED)
panels={f'random_{i+1:02d}':sorted(rng.choice(allgenes,200,replace=False)) for i in range(20)}
save('random_panel_genes.csv',[dict(panel=p,gene=g,seed=SEED) for p,gs in panels.items() for g in gs])
old=pd.read_csv(ROOT/'results/primary_predictions.csv')
old=old[old.model=='ridge200']
features=[];metrics=[];predictions=[];sets={};maxerr=0.
for rep in range(10):
    for fold,(tr,te) in enumerate(core.splits(m,core.SEED+rep)):
        xx,mm=x.iloc[tr],m.iloc[tr]
        rank,t=order(xx,mm.y)
        primary=core.Fit('ridge200',seed=core.SEED+rep).fit(xx,mm)
        ss=primary.predict(x.iloc[te],m.iloc[te])
        expected=old[(old.repeat==rep)&(old.fold==fold)].set_index('sample').loc[m.index[te],'score'].to_numpy()
        err=float(np.max(np.abs(ss-expected)));maxerr=max(maxerr,err)
        assert err<1e-10, (rep,fold,err)
        fit_id=f'r{rep+1:02d}_f{fold+1}'
        sets[fit_id]=set(primary.features)
        tmap=dict(zip(allgenes,t))
        for j,g in enumerate(primary.features):
            features.append(dict(fit_id=fit_id,repeat=rep,fold=fold,gene=g,
                coefficient=primary.model.coef_[0,j],source_mean=primary.scaler.mean_[j],
                source_sd=primary.scaler.scale_[j],signed_training_t=tmap[g],rank=j+1))
        candidates={'next200':x.columns[rank[200:400]].tolist(),**panels}
        assert not set(candidates['next200']) & set(primary.features)
        for name,genes in candidates.items():
            fitted=fit_panel(xx,mm.y,genes,core.SEED+rep)
            score,linear=predict(fitted,x.iloc[te],genes)
            row=dict(panel=name,repeat=rep,fold=fold,n_train=len(tr),n_test=len(te))
            metric=panel_metrics(m.y.iloc[te],score,linear)
            decomp,_=core.decompose(m.y.iloc[te],linear,m.bed.iloc[te])
            metrics.append(row|metric|decomp)
            for label,s,z in zip(m.index[te],score,linear):
                predictions.append(row|dict(sample=label,bed=m.loc[label,'bed'],y=int(m.loc[label,'y']),score=s,linear_logit=z))
    print('Completed exploratory source repeat',rep+1,flush=True)
f=save('primary_fold_features.csv',features)
fm=save('representation_fold_metrics.csv',metrics)
save('representation_cv_predictions.csv',predictions)
save('feature_jaccard.csv',[dict(fit_a=a,fit_b=b,jaccard=len(sets[a]&sets[b])/len(sets[a]|sets[b]),
    shared_genes=len(sets[a]&sets[b]),same_repeat=a[:3]==b[:3]) for a,b in itertools.combinations(sets,2)])
frozen=pd.read_csv(ROOT/'results/frozen_primary_coefficients.csv').set_index('gene')
stats=f.groupby('gene').agg(selected_folds=('fit_id','size'),mean_coefficient_when_selected=('coefficient','mean'),
    mean_abs_coefficient_when_selected=('coefficient',lambda s:np.abs(s).mean()),
    positive_coefficient_fraction=('coefficient',lambda s:(s>0).mean())).reindex(allgenes)
stats['selected_folds']=stats.selected_folds.fillna(0).astype(int)
stats['selection_fraction']=stats.selected_folds/30
stats['full_source_selected']=stats.index.isin(frozen.index)
stats['full_source_coefficient']=frozen.coefficient.reindex(stats.index)
stats=stats.rename_axis('gene').reset_index()
save('feature_stability.csv',stats)
rank,_=order(x,m.y)
full_panels={'next200':x.columns[rank[200:400]].tolist(),**panels}
save('full_source_panel_genes.csv',[dict(panel=p,gene=g,source_only_selection=p=='next200') for p,gs in full_panels.items() for g in gs])
ep=[];em=[]
for name,genes in full_panels.items():
    fitted=fit_panel(x,m.y,genes,core.SEED)
    for acc in ['GSE43292','GSE28829']:
        xx,mm=core.load(acc)
        score,linear=predict(fitted,xx,genes)
        met=panel_metrics(mm.y,score,linear)
        if acc=='GSE43292':
            diffs=[]
            for patient in mm.patient.unique():
                yy=mm.patient==patient
                diffs.append(float(linear[yy & (mm.y==1)][0]-linear[yy & (mm.y==0)][0]))
            diffs=np.asarray(diffs)
            met['pair_concordance']=float(np.mean((diffs>0)+.5*(diffs==0)))
        em.append(dict(panel=name,dataset=acc,n=len(mm))|met)
        ep += [dict(panel=name,dataset=acc,sample=sample,patient=mm.loc[sample,'patient'],y=int(mm.loc[sample,'y']),score=s,linear_logit=z)
               for sample,s,z in zip(mm.index,score,linear)]
save('representation_external_predictions.csv',ep)
em=save('representation_external_metrics.csv',em)
panelmeans=fm.groupby('panel').mean(numeric_only=True)
summary={'source_mean_fold_auc':panelmeans.auc.to_dict(),
    'random_panel_source_auc_range':[float(panelmeans.loc[list(panels),'auc'].min()),float(panelmeans.loc[list(panels),'auc'].max())],
    'random_panel_source_auc_median':float(panelmeans.loc[list(panels),'auc'].median()),
    'next200_source_mean_fold_auc':float(panelmeans.loc['next200','auc']),
    'primary_feature_union':int((stats.selected_folds>0).sum()),
    'primary_features_in_all_30_folds':int((stats.selected_folds==30).sum()),
    'primary_features_in_at_least_24_folds':int((stats.selected_folds>=24).sum()),
    'external':em.to_dict('records')}
(OUT/'summary.json').write_text(json.dumps(summary,indent=2),encoding='utf8')
assert all(sha(ROOT/p)==h for p,h in original.items()),'Original numerical result file changed'
assert len(f)==6000 and len(fm)==630 and len(em)==42
assert fm.groupby('panel').size().eq(30).all()
for (_,rep),block in pd.DataFrame(predictions).groupby(['panel','repeat']):
    assert block['sample'].nunique()==95 and len(block)==95
report={'recorded_utc':datetime.datetime.now(datetime.timezone.utc).isoformat(),
    'scope':'Exploratory post-result representation controls; no target feature/threshold selection; original primary retained',
    'seed':SEED,'random_panels':20,'genes_per_panel':200,'source_fits':630,'external_source_fits':21,
    'ranking_numeric_correction':'All new linear classifiers use decision_function for AUROC/AP/pair concordance to avoid sigmoid saturation ties; see implementation_note_05_logit_ranking.txt. Probabilities retained for threshold and Brier.',
    'primary_recovery_fits':30,'max_saved_primary_prediction_error':maxerr,'passed':True,
    'original_result_sha256_unchanged':original,
    'new_output_sha256':{p.name:sha(p) for p in OUT.iterdir() if p.is_file()},
    'limitations':['Panel draws and overlapping folds are not independent cohorts or uncertainty intervals',
                   'Near-ceiling source data and inherited all-array normalization remain',
                   'No random-panel winner selected; no causal gene importance or new biomarker claim']}
(E/'feature_analysis_audit.json').write_text(json.dumps(report,indent=2),encoding='utf8')
print(json.dumps({k:v for k,v in summary.items() if k!='external'},indent=2),flush=True)
```
