## Supplementary material for "Anatomical comparison populations and threshold transport in vascular transcriptomic classification": Reproducibility archive: code, provenance and results: explore_threshold_shift.py

```
"""Post-result exploratory source-only thresholds, score accounting and influence.

See protocol/exploratory_extension_04_20261026.txt. This adds diagnostics without
replacing the original model, threshold result, prediction files or primary estimand.
"""
from pathlib import Path
import datetime, hashlib, json, sys, time
import numpy as np
import pandas as pd
from scipy.special import expit
from sklearn.metrics import roc_auc_score, average_precision_score, roc_curve, confusion_matrix

ROOT = Path(__file__).resolve().parents[1]
sys.path.insert(0, str(ROOT/'code'))
from analyze import load, Fit, splits, BEDS, SEED

OUT = ROOT/'results/exploratory_threshold'
OUT.mkdir(parents=True, exist_ok=True)
EVIDENCE = ROOT/'evidence/figure_revision_20261026'
EVIDENCE.mkdir(parents=True, exist_ok=True)
RULES = ['fixed_0.5', 'source_max_youden', 'source_sensitivity_0.90']
INNER_SEED = 20261026
started = time.perf_counter()

def sha(path):
    return hashlib.sha256(path.read_bytes()).hexdigest()

original = {str(p.relative_to(ROOT)).replace('\\','/'):sha(p)
            for p in sorted((ROOT/'results').iterdir()) if p.is_file()}
input_paths = [ROOT/'code/analyze.py', ROOT/'protocol/exploratory_extension_04_20261026.txt']
input_paths += [ROOT/'data'/f'{a}_{s}' for a in ['GSE100927','GSE43292','GSE28829']
                for s in ['rank.pkl','samples.csv']]
inputs = {str(p.relative_to(ROOT)).replace('\\','/'):sha(p) for p in input_paths}

def save(name, records):
    df = records if isinstance(records, pd.DataFrame) else pd.DataFrame(records)
    df.to_csv(OUT/(name+'.csv'), index=False)
    return df

def operating(y, score, threshold):
    y, score = np.asarray(y, dtype=int), np.asarray(score, dtype=float)
    pred = score >= threshold
    tp, fn = int(np.sum(pred & (y==1))), int(np.sum(~pred & (y==1)))
    tn, fp = int(np.sum(~pred & (y==0))), int(np.sum(pred & (y==0)))
    sen, spe = tp/(tp+fn), tn/(tn+fp)
    return dict(tp=tp, fn=fn, tn=tn, fp=fp, sensitivity=sen,
                specificity=spe, balanced_accuracy=(sen+spe)/2, youden_j=sen+spe-1,
                positive_predictions=int(pred.sum()), n=len(y))

def candidates(score):
    unique = np.unique(np.asarray(score, dtype=float))
    return np.unique(np.r_[0., (unique[:-1]+unique[1:])/2, 1.])

threshold_rows, candidate_rows, inner_rows = [], [], []

def choose(context, y, score, seed, description):
    cc = pd.DataFrame([dict(threshold=float(t), **operating(y, score, t)) for t in candidates(score)])
    maximum = cc.youden_j.max()
    tied = cc[np.isclose(cc.youden_j, maximum, rtol=0, atol=1e-12)].copy()
    tied['distance_to_half'] = abs(tied.threshold-.5)
    youden = tied.sort_values(['distance_to_half','threshold']).iloc[0].threshold
    high_sens = cc[cc.sensitivity >= .90-1e-12].threshold.max()
    selected = dict(zip(RULES, [.5, float(youden), float(high_sens)]))
    for row in cc.to_dict('records'):
        candidate_rows.append(dict(learning_context=context, **row))
    for rule, threshold in selected.items():
        threshold_rows.append(dict(learning_context=context, rule=rule, threshold=threshold,
            inner_seed=seed, learning_recipe=description, candidates=len(cc),
            source_optimization_quantity=True, **operating(y, score, threshold)))
    return selected

x, m = load('GSE100927')
saved_primary = pd.read_csv(ROOT/'results/primary_predictions.csv')
first = saved_primary[(saved_primary.model=='ridge200') & (saved_primary['repeat']==0)].copy()
assert len(first)==95 and first['sample'].nunique()==95 and set(first['sample'])==set(m.index)
first = first.set_index('sample').loc[m.index].rename_axis('sample').reset_index()
assert np.array_equal(first.y.to_numpy(), m.y.to_numpy())
all_thresholds = choose('all_source', first.y, first.score, SEED,
    'Original repetition0: one held-out probability per label group from three different fitted folds; pooled only to learn a technical threshold, not to replace the primary within-fold AUROC estimand')
for row in first.itertuples(index=False):
    inner_rows.append(dict(learning_context='all_source', sample=row.sample, bed=row.bed,
                           y=int(row.y), fold=int(row.fold), score=row.score, seed=SEED,
                           origin='original_saved_repeat0'))

ext = pd.read_csv(ROOT/'results/external_predictions.csv')
ext_metrics = pd.read_csv(ROOT/'results/external_metrics.csv')
epairs = pd.read_csv(ROOT/'results/external_pairs.csv')
old_transport = pd.read_csv(ROOT/'results/transport_predictions.csv')
eval_rows, pred_rows, curve_rows, replay = [], [], [], {}

def evaluate(dataset, context, mm, ss, thresholds, source_fit_kind):
    yy = mm.y.to_numpy(dtype=int)
    auc, ap = roc_auc_score(yy,ss), average_precision_score(yy,ss)
    for rule, threshold in thresholds.items():
        eval_rows.append(dict(dataset=dataset, learning_context=context, rule=rule,
            threshold=threshold, auc=auc, average_precision=ap, source_fit_kind=source_fit_kind,
            **operating(yy,ss,threshold)))
    for i, (sample, row) in enumerate(mm.iterrows()):
        item = dict(dataset=dataset, learning_context=context, sample=str(sample),
                    bed=str(row.bed), patient=str(row.patient), y=int(row.y), score=float(ss[i]))
        item.update({rule+'_prediction':int(ss[i]>=thresholds[rule]) for rule in RULES})
        pred_rows.append(item)
    grid = np.unique(np.r_[0., np.logspace(-12,0,401), candidates(ss), list(thresholds.values())])
    for t in grid:
        curve_rows.append(dict(dataset=dataset, threshold=float(t),
            curve_role='Descriptive outcome-dependent curve; not target threshold selection', **operating(yy,ss,t)))

for bi, target in enumerate(BEDS):
    training = np.flatnonzero(m.bed != target)
    test = np.flatnonzero(m.bed == target)
    xx, mm = x.iloc[training], m.iloc[training]
    oo = np.full(len(mm), np.nan)
    inner_seed = INNER_SEED+bi
    for fold, (tr, te) in enumerate(splits(mm, inner_seed)):
        fit = Fit('ridge200',seed=SEED).fit(xx.iloc[tr],mm.iloc[tr])
        ss = fit.predict(xx.iloc[te],mm.iloc[te])
        oo[te] = ss
        assert not set(mm.index[tr]) & set(mm.index[te])
        assert not set(mm.index) & set(m.index[test])
        for i, s in zip(te,ss):
            inner_rows.append(dict(learning_context='LOBO_to_'+target, sample=str(mm.index[i]),
                bed=str(mm.bed.iloc[i]), y=int(mm.y.iloc[i]), fold=fold, score=float(s),
                seed=inner_seed, origin='new_inner_crossfit_only_outer_training_beds'))
    assert np.isfinite(oo).all()
    thresholds = choose('LOBO_to_'+target, mm.y, oo, inner_seed,
        'Single three-fold inner cross-fit stratified by four available bed/status cells; all feature selection/scaling/fitting inside inner training')
    fit = Fit('ridge200',seed=SEED).fit(xx,mm)
    ss = fit.predict(x.iloc[test],m.iloc[test])
    old = old_transport[(old_transport.model=='ridge200') & (old_transport.source=='other_two') &
                        (old_transport.target==target) & (~old_transport.purge_possible_controls)].set_index('sample')
    err = float(np.max(abs(ss-old.loc[m.index[test],'score'].to_numpy())))
    assert err < 1e-10
    replay[target] = err
    evaluate('LOBO_'+target, 'LOBO_to_'+target, m.iloc[test], ss, thresholds,
             'Full outer-training fit; its held-out bed does not inform threshold')

for acc in ['GSE43292','GSE28829']:
    xt, mt = load(acc)
    saved = ext[(ext.dataset==acc)&(ext.model=='ridge200')].set_index('sample').loc[mt.index]
    assert np.array_equal(saved.y.to_numpy(),mt.y.to_numpy())
    evaluate(acc, 'all_source', mt, saved.score.to_numpy(), all_thresholds,
             'Unchanged original frozen full-source model; saved target probabilities')

save('threshold_rules',threshold_rows)
save('source_threshold_candidates',candidate_rows)
save('source_crossfit_learning_scores',inner_rows)
evaluations = save('evaluation_metrics',eval_rows)
save('evaluation_predictions',pred_rows)
save('threshold_response_curves',curve_rows)

# Exact frozen-model accounting. Only source coefficients determine display genes.
coef = pd.read_csv(ROOT/'results/frozen_primary_coefficients.csv')
frozen = json.loads((ROOT/'results/frozen_primary_model.json').read_text())
intercept = frozen['intercept']
top = coef.assign(abs_coefficient=abs(coef.coefficient)).sort_values(
    ['abs_coefficient','gene'],ascending=[False,True]).head(20).copy()
top['source_absolute_coefficient_rank'] = np.arange(1,21)
save('top20_source_coefficient_genes',top)
top_rank = dict(zip(top.gene,top.source_absolute_coefficient_rank))
samples, all_contributions, mean_contributions, decompositions, standardization = [], [], [], [], []
reconstruction, caches = {}, {}
for acc in ['GSE100927','GSE43292','GSE28829']:
    xx, mm = load(acc)
    a = xx[coef.gene].to_numpy()
    z = (a-coef.source_mean.to_numpy())/coef.source_sd.to_numpy()
    c = z*coef.coefficient.to_numpy()
    eta = intercept+c.sum(axis=1)
    pp = expit(eta)
    assert np.isfinite(z).all() and np.isfinite(eta).all()
    if acc != 'GSE100927':
        saved = ext[(ext.dataset==acc)&(ext.model=='ridge200')].set_index('sample').loc[mm.index]
        reconstruction[acc] = float(np.max(abs(pp-saved.score.to_numpy())))
        assert reconstruction[acc] < 1e-10
    caches[acc]=(mm,z,c,eta,pp)
    for i,(sample,row) in enumerate(mm.iterrows()):
        samples.append(dict(dataset=acc,sample=str(sample),patient=str(row.patient),bed=str(row.bed),
            y=int(row.y),linear_logit=eta[i],probability=pp[i],intercept=intercept,
            positive_contribution_sum=np.maximum(c[i],0).sum(),negative_contribution_sum=np.minimum(c[i],0).sum(),
            selected_gene_count=200,z_abs_gt3_fraction=np.mean(abs(z[i])>3),median_abs_z=np.median(abs(z[i])),
            source_in_sample_reference=acc=='GSE100927'))
        for j,gene in enumerate(coef.gene):
            all_contributions.append(dict(dataset=acc,sample=str(sample),patient=str(row.patient),y=int(row.y),
                gene=gene,source_top20_rank=top_rank.get(gene,np.nan),standardized_value=z[i,j],weighted_contribution=c[i,j]))
    for state,mask in [('all',np.ones(len(mm),dtype=bool)),('0',mm.y.to_numpy()==0),('1',mm.y.to_numpy()==1)]:
        cm,zm=c[mask].mean(axis=0),z[mask].mean(axis=0)
        mean_eta=float(eta[mask].mean())
        assert abs(mean_eta-(intercept+cm.sum()))<1e-10
        for j,gene in enumerate(coef.gene):
            mean_contributions.append(dict(dataset=acc,class_state=state,gene=gene,source_top20_rank=top_rank.get(gene,np.nan),
                mean_standardized_value=zm[j],mean_weighted_contribution=cm[j],coefficient=coef.coefficient.iloc[j],
                n_specimens=int(mask.sum())))
        decompositions.append(dict(dataset=acc,class_state=state,n_specimens=int(mask.sum()),intercept=intercept,
            positive_mean_feature_sum=np.maximum(cm,0).sum(),negative_mean_feature_sum=np.minimum(cm,0).sum(),
            mean_logit=mean_eta,mean_probability=float(pp[mask].mean()),
            source_in_sample_reference=acc=='GSE100927',aggregation='Observed class composition' if state=='all' else 'One deposited class'))
        standardization.append(dict(dataset=acc,class_state=state,n_specimens=int(mask.sum()),n_gene_values=int(mask.sum()*200),
            n_abs_z_gt3=int(np.sum(abs(z[mask])>3)),fraction_abs_z_gt3=float(np.mean(abs(z[mask])>3)),
            median_abs_z=float(np.median(abs(z[mask]))),p90_abs_z=float(np.quantile(abs(z[mask]),.9))))
    # Class-balanced aggregate removes differences in counts, not endpoint mismatch.
    cm=(c[mm.y.to_numpy()==0].mean(axis=0)+c[mm.y.to_numpy()==1].mean(axis=0))/2
    decompositions.append(dict(dataset=acc,class_state='class_balanced',n_specimens=len(mm),intercept=intercept,
        positive_mean_feature_sum=np.maximum(cm,0).sum(),negative_mean_feature_sum=np.minimum(cm,0).sum(),
        mean_logit=intercept+cm.sum(),mean_probability=(pp[mm.y.to_numpy()==0].mean()+pp[mm.y.to_numpy()==1].mean())/2,
        source_in_sample_reference=acc=='GSE100927',aggregation='Equal weight to two class means; not observed prevalence and not exchangeable endpoints'))

save('score_geometry_samples',samples)
save('feature_contributions_all',all_contributions)
save('mean_feature_contributions',mean_contributions)
decomp = save('class_logit_decomposition',decompositions)
zstats = save('standardization_displacement',standardization)

mm,z,c,eta,pp=caches['GSE43292']
pair_differences, pair_margins = [],[]
for patient in mm.patient.unique():
    ii=np.flatnonzero((mm.patient==patient)&(mm.y==0))[0]
    ip=np.flatnonzero((mm.patient==patient)&(mm.y==1))[0]
    dc=c[ip]-c[ii]
    assert abs(dc.sum()-(eta[ip]-eta[ii]))<1e-10
    original_pair=epairs[(epairs.model=='ridge200')&(epairs.patient.astype(str)==str(patient))].iloc[0]
    assert abs((pp[ip]-pp[ii])-original_pair.difference)<1e-10
    pair_margins.append(dict(patient=patient,intact_logit=eta[ii],plaque_logit=eta[ip],logit_difference=eta[ip]-eta[ii],
        probability_difference=pp[ip]-pp[ii],paired_ordering=int(np.sign(eta[ip]-eta[ii])),
        retrospective_reversed_pair=bool(eta[ip]<eta[ii])))
    for j,gene in enumerate(coef.gene):
        pair_differences.append(dict(patient=patient,gene=gene,source_top20_rank=top_rank.get(gene,np.nan),
            weighted_logit_difference=dc[j]))
save('paired_feature_logit_differences',pair_differences)
save('paired_primary_margins',pair_margins)
signs=epairs.copy()
signs['paired_ordering']=np.sign(signs.difference).astype(int)
save('paired_five_rule_sign_map',signs)

# Fixed-score case deletion: no fitting, no bootstrap, no new independent evidence.
paired_delete,hist_delete=[],[]
for acc in ['GSE43292','GSE28829']:
    for model, block in ext[ext.dataset==acc].groupby('model'):
        full_auc=roc_auc_score(block.y,block.score)
        recorded=ext_metrics[(ext_metrics.dataset==acc)&(ext_metrics.model==model)].iloc[0]
        assert abs(full_auc-recorded.auc)<1e-10
        if acc=='GSE43292':
            pairs=epairs[epairs.model==model]
            full_concordance=float(((pairs.difference>0)+.5*(pairs.difference==0)).mean())
            assert abs(full_concordance-recorded.pair_concordance)<1e-10
            assert block.groupby('patient').size().eq(2).all() and block.groupby('patient').y.sum().eq(1).all()
            for patient in block.patient.unique():
                keep=block[block.patient!=patient]
                kept_pairs=pairs[pairs.patient.astype(str)!=str(patient)]
                auc=roc_auc_score(keep.y,keep.score)
                concordance=float(((kept_pairs.difference>0)+.5*(kept_pairs.difference==0)).mean())
                paired_delete.append(dict(model=model,omitted_patient=patient,n_patients=31,n_tissues=len(keep),
                    auc=auc,auc_change=auc-full_auc,pair_concordance=concordance,
                    pair_concordance_change=concordance-full_concordance,
                    interpretation='Fixed-score leave-one-known-patient-out influence, not refitting or a confidence interval'))
        else:
            for sample in block['sample']:
                keep=block[block['sample']!=sample]
                auc=roc_auc_score(keep.y,keep.score)
                hist_delete.append(dict(model=model,omitted_sample=sample,
                    omitted_class=int(block.loc[block['sample']==sample,'y'].iloc[0]),n_specimens=len(keep),
                    n_early=int(sum(keep.y==0)),n_advanced=int(sum(keep.y==1)),auc=auc,auc_change=auc-full_auc,
                    interpretation='Fixed-score leave-one-specimen-out influence; donor independence unknown'))
paired_delete=save('paired_deletion_influence',paired_delete)
hist_delete=save('histology_deletion_influence',hist_delete)

# Re-read exported records and cross-check with distinct library operations.
learning_check=pd.read_csv(OUT/'source_crossfit_learning_scores.csv')
rules_check=pd.read_csv(OUT/'threshold_rules.csv')
for context,bb in learning_check.groupby('learning_context'):
    fpr,tpr,_=roc_curve(bb.y,bb.score,drop_intermediate=False)
    rr=rules_check[rules_check.learning_context==context]
    assert abs(rr.loc[rr.rule=='source_max_youden','youden_j'].iloc[0]-np.max(tpr-fpr))<1e-12
    cc=pd.read_csv(OUT/'source_threshold_candidates.csv')
    cc=cc[cc.learning_context==context]
    assert abs(rr.loc[rr.rule=='source_sensitivity_0.90','threshold'].iloc[0]-
               cc.loc[cc.sensitivity>=.90-1e-12,'threshold'].max())<1e-12
pred_check=pd.read_csv(OUT/'evaluation_predictions.csv')
metrics_check=pd.read_csv(OUT/'evaluation_metrics.csv')
for row in metrics_check.itertuples(index=False):
    bb=pred_check[pred_check.dataset==row.dataset]
    tn,fp,fn,tp=confusion_matrix(bb.y,bb[row.rule+'_prediction'],labels=[0,1]).ravel()
    assert (tn,fp,fn,tp)==(row.tn,row.fp,row.fn,row.tp)
assert len(learning_check)==285 and len(pred_check)==188
assert len(paired_delete)==160 and len(hist_delete)==145

after = {str(p.relative_to(ROOT)).replace('\\','/'):sha(p)
         for p in sorted((ROOT/'results').iterdir()) if p.is_file()}
assert original==after, 'Original core result files changed during exploratory analysis'
summary = dict(
    study_status='Post-result exploratory diagnostic; prior threshold/logit inspection disclosed in extension04',
    full_source_threshold_recipe='Original saved repetition0 three-fold cross-fitted scores, not averages over ten repetitions; the latter was the preliminary feasibility check only',
    full_source_thresholds=all_thresholds,
    nested_inner_seeds={b:INNER_SEED+i for i,b in enumerate(BEDS)},
    nested_outer_model_seed=SEED,
    threshold_rules='Candidate endpoints0,1 and adjacent unique-score midpoints. Youden ties within1e-12: closest0.5 then smaller. Sensitivity rule: largest candidate reaching>=0.90 source sensitivity; not a clinical target.',
    evaluation_records=evaluations.to_dict('records'),
    score_geometry_class_means=decomp.to_dict('records'),
    standardization_displacement=zstats.to_dict('records'),
    primary_reversed_patients=[str(p['patient']) for p in pair_margins if p['paired_ordering']<0],
    paired_auc_deletion_ranges=paired_delete.groupby('model').auc.agg(['min','max']).reset_index().to_dict('records'),
    histology_auc_deletion_ranges=hist_delete.groupby('model').auc.agg(['min','max']).reset_index().to_dict('records'),
    limits=['Threshold objectives on source cross-fitted scores are optimization quantities, not unbiased source performance estimates.',
            'Inner and final fitted score scales can differ; nested outer evaluation tests the full source-only rule.',
            'Cross-fitted scores are pooled here only as threshold-learning inputs, not for the original nine-cell AUROC estimand.',
            'Known target outcomes were previously inspected. This is source-only fitting within an openly exploratory post-result investigation.',
            'Source full-fit score/feature displays are in-sample references, not validation performance.',
            'Linear attribution and |z|>3 displacement do not isolate platform, procurement or endpoint causes or establish causal genes.',
            'Case deletions condition on fixed scores and are not refits, confidence intervals or independent replication.'])
(OUT/'summary.json').write_text(json.dumps(summary,indent=2),encoding='utf8')
audit=dict(created_utc=datetime.datetime.now(datetime.timezone.utc).isoformat(),elapsed_seconds=time.perf_counter()-started,
    executed_new_inner_fits=9,executed_outer_fits=3,external_models_refitted=0,
    passed=True,original_core_results_unchanged=original==after,original_result_sha256=original,
    input_sha256=inputs,outer_prediction_replay_max_error=replay,
    frozen_external_score_reconstruction_max_error=reconstruction,
    threshold_record_count=len(threshold_rows),evaluation_record_count=len(evaluations),
    paired_deletion_rows=len(paired_delete),histology_deletion_rows=len(hist_delete),
    serialized_exports_cross_checked=True,
    output_sha256={p.name:sha(p) for p in sorted(OUT.iterdir()) if p.is_file()},
    validations=['Original repetition0 covers95unique source label groups exactly once.',
      'All inner folds disjoint; held-out arterial bed absent from threshold learning.',
      'Nested outer scores reproduce original primary leave-one-bed-out scores within1e-10.',
      'Frozen coefficient accounting reconstructs saved external probabilities within1e-10.',
      'Every class and paired logit reconstruction checked within1e-10.',
      'Re-read thresholds attain independent sklearn ROC maximumYouden and statedsource sensitivityconstraint.',
      'Re-read15evaluationmetricrows match independent sklearnconfusion_matrix on188savedpredictionrecords.',
      'All original result file hashes unchanged.'])
(EVIDENCE/'threshold_analysis_audit.json').write_text(json.dumps(audit,indent=2),encoding='utf8')
print(json.dumps(dict(passed=True,elapsed_seconds=audit['elapsed_seconds'],thresholds=all_thresholds,
      external_metrics=evaluations[evaluations.dataset.isin(['GSE43292','GSE28829'])][
          ['dataset','rule','threshold','sensitivity','specificity','balanced_accuracy']].to_dict('records'),
      output=str(OUT)),indent=2))
```
