## Supplementary material for "Anatomical comparison populations and threshold transport in vascular transcriptomic classification": Reproducibility archive: code, provenance and results: export_descriptive_scores.py

```
"""Export protocol-promised descriptive averages from saved held-out scores.

No model fitting, preprocessing, or primary-estimand changes occur here. The
averaged scores mix fitted models and are not used for the nine-cell analysis.
"""
from datetime import datetime, timezone
import hashlib
import json
from pathlib import Path

import numpy as np
import pandas as pd
from sklearn.metrics import roc_auc_score


ROOT = Path(__file__).resolve().parents[1]
RESULTS = ROOT / "results"
EVIDENCE = ROOT / "evidence"
MODEL_ORDER = ["prevalence", "bed_sex", "C1Q", "Wu6", "ridge200", "centroid200", "forest200"]
SCORE_MODELS = {"C1Q", "centroid200"}


def sha256(path):
    return hashlib.sha256(path.read_bytes()).hexdigest()


def require(condition, message):
    if not condition:
        raise ValueError(message)


def main():
    prediction_path = RESULTS / "primary_predictions.csv"
    fold_path = RESULTS / "primary_fold_metrics.csv"
    cell_path = RESULTS / "primary_pair_cells.csv"
    input_hashes = {str(p.relative_to(ROOT)).replace("\\", "/"): sha256(p)
                    for p in (prediction_path, fold_path, cell_path)}
    predictions = pd.read_csv(prediction_path)
    folds = pd.read_csv(fold_path)
    cells = pd.read_csv(cell_path)
    require(set(predictions.scenario) == {"primary"}, "Unexpected prediction scenario")
    require(set(predictions.model) == set(MODEL_ORDER), "Unexpected primary models")
    require(len(predictions) == 7 * 95 * 10, "Unexpected number of predictions")
    require(np.isfinite(predictions.score).all(), "Nonfinite saved score")
    require(not predictions.duplicated(["model", "repeat", "sample"]).any(),
            "More than one held-out score per model/repetition/label group")
    require((predictions.groupby("sample")[["bed", "y"]].nunique() == 1).all().all(),
            "Inconsistent label-group metadata")
    require(predictions.groupby(["repeat", "sample"]).fold.nunique().eq(1).all(),
            "Comparator models did not share the same held-out partition")

    output_rows = []
    metric_rows = []
    for model in MODEL_ORDER:
        subset = predictions[predictions.model == model]
        require(subset["sample"].nunique() == 95, f"{model}: expected 95 label groups")
        require(subset.groupby("repeat")["sample"].nunique().eq(95).all(),
                f"{model}: incomplete repetition")
        for sample, group in subset.groupby("sample", sort=True):
            require(set(group["repeat"]) == set(range(10)) and len(group) == 10,
                    f"{model}/{sample}: incomplete ten-repeat coverage")
            output_rows.append({
                "scenario": "primary_descriptive_mean_crossfit",
                "model": model,
                "label_group": sample,
                "bed": group.bed.iloc[0],
                "y": int(group.y.iloc[0]),
                "n_held_out_scores": len(group),
                "score_type": "uncalibrated_score" if model in SCORE_MODELS else "classifier_probability",
                "mean_crossfit_score": float(group.sort_values("repeat").score.mean()),
            })
        aggregate = pd.DataFrame([row for row in output_rows if row["model"] == model])
        positive = aggregate.loc[aggregate.y == 1, "mean_crossfit_score"].to_numpy()
        negative = aggregate.loc[aggregate.y == 0, "mean_crossfit_score"].to_numpy()
        require(len(positive) == 65 and len(negative) == 30, "Unexpected class counts")
        pair_auc = float(np.mean((positive[:, None] > negative[None, :])
                                + 0.5 * (positive[:, None] == negative[None, :])))
        descriptive_auc = float(roc_auc_score(aggregate.y, aggregate.mean_crossfit_score))
        require(abs(pair_auc - descriptive_auc) < 1e-12,
                f"{model}: pair-order and standard AUROC disagree")
        model_folds = folds[folds.model == model]
        require(len(model_folds) == 30 and model_folds.groupby("repeat").size().eq(3).all(),
                f"{model}: unexpected fold metric coverage")
        primary_auc = float(model_folds.groupby("repeat").auc.mean().mean())
        metric_rows.append({
            "model": model,
            "n_label_groups": len(aggregate),
            "n_positive_label_groups": len(positive),
            "n_negative_label_groups": len(negative),
            "n_held_out_scores_per_label_group": 10,
            "descriptive_mean_crossfit_pooled_auc": descriptive_auc,
            "primary_equal_fold_mean_auc": primary_auc,
            "descriptive_minus_primary_auc": descriptive_auc - primary_auc,
            "interpretation": "descriptive_only_not_primary_nine_cell_estimand_or_independent_validation",
        })

    score_output = RESULTS / "descriptive_mean_crossfit_scores.csv"
    metrics_output = RESULTS / "descriptive_mean_crossfit_metrics.csv"
    pd.DataFrame(output_rows).to_csv(score_output, index=False, float_format="%.17g")
    pd.DataFrame(metric_rows).to_csv(metrics_output, index=False, float_format="%.17g")
    # Check the written artifact as well as the in-memory aggregation.
    saved = pd.read_csv(score_output, float_precision="round_trip")
    require(len(saved) == 665 and not saved.duplicated(["model", "label_group"]).any(),
            "Invalid exported label/model table")
    require(saved.n_held_out_scores.eq(10).all(), "Invalid exported prediction counts")

    primary_cells = cells[cells.model == "ridge200"]
    primary_folds = folds[folds.model == "ridge200"]
    mean_cells = primary_cells.groupby(["case_bed", "control_bed"]).auc.mean()
    diagonal = primary_cells[primary_cells.case_bed == primary_cells.control_bed]
    require(diagonal.auc.eq(1).all(), "Primary diagonal ceiling differs from reported result")
    verification = {
        "mean_per_fold_minimum": float(primary_folds.mix_min.mean()),
        "mean_per_fold_maximum": float(primary_folds.mix_max.mean()),
        "mean_per_fold_span": float(primary_folds.mix_span.mean()),
        "minimum_of_mean_cell_matrix": float(mean_cells.min()),
        "maximum_of_mean_cell_matrix": float(mean_cells.max()),
        "minimum_individual_fold_cell": float(primary_cells.auc.min()),
        "minimum_cell_pair_count": int(primary_cells.pairs.min()),
        "maximum_cell_pair_count": int(primary_cells.pairs.max()),
        "all_primary_diagonal_cells_equal_one": bool(diagonal.auc.eq(1).all()),
        "primary_mean_delta_mix": float(primary_folds.delta_mix.mean()),
    }
    for path in (prediction_path, fold_path, cell_path):
        key = str(path.relative_to(ROOT)).replace("\\", "/")
        require(sha256(path) == input_hashes[key], f"Input changed during export: {key}")
    audit = {
        "created_utc": datetime.now(timezone.utc).isoformat(),
        "passed": True,
        "operation": "saved-score aggregation only; no model fitting or primary-analysis change",
        "protocol_status": "Descriptive output promised by internal protocol v1; assembled after review from original saved predictions. The protocol was not a registration and did not specify a one-sided positive-inflation primary hypothesis.",
        "unit": "complete-title label group; not authenticated patient or donor",
        "input_sha256": input_hashes,
        "output_sha256": {str(p.relative_to(ROOT)).replace("\\", "/"): sha256(p)
                          for p in (score_output, metrics_output)},
        "rows": len(saved),
        "models": MODEL_ORDER,
        "metrics": metric_rows,
        "primary_cell_verification": verification,
        "limits": [
            "Every label group has one held-out score per repetition; fits and observations overlap across repetitions.",
            "Averaging scores from differently fitted models is descriptive and does not replace within-fold nine-cell calculations.",
            "Classifier probabilities are technical outputs, not population disease risks; C1Q and centroid outputs are uncalibrated scores.",
            "No confidence interval or independent-replication claim is attached to the averaged-score AUROC.",
        ],
    }
    EVIDENCE.mkdir(exist_ok=True)
    (EVIDENCE / "descriptive_mean_crossfit_export_audit.json").write_text(
        json.dumps(audit, indent=2) + "\n", encoding="utf-8")
    print(json.dumps({"passed": True, "rows": len(saved), "metrics": metric_rows,
                      "primary_cell_verification": verification}, indent=2))


if __name__ == "__main__":
    main()
```
