## Supplementary material for "Anatomical comparison populations and threshold transport in vascular transcriptomic classification": Reproducibility archive: code, provenance and results: metadata.py

```
"""Read deposited sample metadata without reading expression values."""
from pathlib import Path
import gzip,csv,json,re,datetime,collections
ROOT=Path(__file__).resolve().parents[1]
def metadata(acc):
    rows=[]
    with gzip.open(ROOT/'data'/f'{acc}_series_matrix.txt.gz','rt',encoding='utf-8') as f:
        for line in f:
            if line.startswith('!series_matrix_table_begin'): break
            if line.startswith('!Sample_'): rows.append(next(csv.reader([line],delimiter='\t')))
    n=len(rows[0])-1
    records=[{'accession':acc} for _ in range(n)]
    counts=collections.Counter()
    for row in rows:
        key=row[0].removeprefix('!Sample_'); counts[key]+=1
        if counts[key]>1: key+=f'_{counts[key]}'
        for i,val in enumerate(row[1:]):records[i][key]=val
    return records
if __name__=='__main__':
    for acc in ['GSE100927','GSE43292','GSE28829']:
        rec=metadata(acc)
        p=ROOT/'evidence'/f'{acc}_sample_metadata.json'
        p.write_text(json.dumps(rec,indent=2),encoding='utf8')
        print(acc,len(rec),'fields',list(rec[0]))
        print(json.dumps([{k:v for k,v in x.items() if k in ['title','geo_accession','source_name_ch1','description','data_processing'] or k.startswith('characteristics')} for x in rec[:2]],indent=2))
```
