## Supplementary material for "Anatomical comparison populations and threshold transport in vascular transcriptomic classification": Reproducibility archive: code, provenance and results: package_release.py

```
"""Build review-delivery source/reproduction archives from an explicit allowlist.

No publication or external upload is performed. Optional smoke checks execute in
an extracted temporary copy, downloading only the six hash-locked official inputs.
The exploratory validator performs two limited fitted-panel replays, not a full
model refit. This compact distribution is not an offline self-contained archive.
"""
from pathlib import Path
import argparse, datetime, hashlib, json, subprocess, sys, tempfile, zipfile

ROOT=Path(__file__).resolve().parents[1]
CODE=['acquire.py','metadata.py','prepare.py','analyze.py','export_descriptive_scores.py',
      'validate_results.py','check_paired_bootstrap.py','plot_figures.py','build_supplement.py',
      'explore_feature_dependence.py','explore_threshold_shift.py','validate_exploratory.py',
      'plot_feature_extension.py','plot_threshold_extension.py','plot_supplement_extension.py',
      'assemble_figure_catalog.py','build_exploratory_supplement.py',
      'assemble.py','render_review.py','package_release.py']
MANUSCRIPT=['main.tex','supplement.tex','titlepage.tex','body.tex','body_for_compilation.tex',
            'figure_pages.tex','declarations.tex','references.bib','author_metadata.json',
            'supplement_body.tex','statistical_addendum.tex','exploratory_supplement.tex']
EVIDENCE=['acquisition.json','expected_inputs_v1.json','environment.json','analysis_environment_original.json',
          'dataset_registry.csv','dataset_registry.json','preprocessing_qc.json','common_gene_vocabulary.txt',
          'GSE100927_sample_metadata.json','GSE43292_sample_metadata.json','GSE28829_sample_metadata.json',
          'GSE100927_label_grouping_audit.csv','GSE100927_repeated_label_groups.json',
          'GPL17077_unambiguous_mapping.csv','GPL570_unambiguous_mapping.csv','GPL6244_unambiguous_mapping.csv',
          'independent_references_verified.bib','independent_references_verified.json',
          'independent_reference_full_author_overlap_audit.json','protocol_freeze_sha256.json',
          'amendment_01_sha256.json','descriptive_mean_crossfit_export_audit.json',
          'independent_saved_results_validation.json','revision05_bootstrap_checks.json','analysis_main.log','analysis_robust.log',
          'citations/recommended_team_references.bib',
          'figure_revision_20261026/pre_expansion/manifest.json',
          'figure_revision_20261026/feature_analysis_audit.json',
          'figure_revision_20261026/threshold_analysis_audit.json',
          'figure_revision_20261026/independent_extension_validation.json',
          'figure_revision_20261026/exploratory_supplement_audit.json']
SCENARIOS=['primary','top50','top500','C001','C1','native','exclude_ambiguous','104_grouped_arrays','possible_shared_controls']
RESULTS=[f'{s}_{suffix}.csv' for s in SCENARIOS for suffix in ['fold_metrics','predictions','pair_cells']]
RESULTS += ['primary_analysis_units.csv','external_predictions.csv','external_metrics.csv','external_pairs.csv',
            'frozen_primary_coefficients.csv','frozen_primary_model.json','transport_metrics.csv','transport_predictions.csv',
            'sensitivity_all_fold_metrics.csv','permutation_nulls.csv','full_pipeline_subsampling.csv',
            'descriptive_mean_crossfit_scores.csv','descriptive_mean_crossfit_metrics.csv',
            'ambiguous_label_profile_similarity.csv','unmatched_within_stratum_similarity.csv']
RESULTS += ['exploratory_features/'+name for name in [
    'feature_jaccard.csv','feature_stability.csv','full_source_panel_genes.csv',
    'primary_fold_features.csv','random_panel_genes.csv','representation_cv_predictions.csv',
    'representation_external_metrics.csv','representation_external_predictions.csv',
    'representation_fold_metrics.csv','summary.json']]
RESULTS += ['exploratory_threshold/'+name for name in [
    'class_logit_decomposition.csv','evaluation_metrics.csv','evaluation_predictions.csv',
    'feature_contributions_all.csv','histology_deletion_influence.csv',
    'mean_feature_contributions.csv','paired_deletion_influence.csv',
    'paired_feature_logit_differences.csv','paired_five_rule_sign_map.csv',
    'paired_primary_margins.csv','score_geometry_samples.csv','source_crossfit_learning_scores.csv',
    'source_threshold_candidates.csv','standardization_displacement.csv','summary.json',
    'threshold_response_curves.csv','threshold_rules.csv','top20_source_coefficient_genes.csv']]
RESULTS += ['exploratory_display/source_score_stability.csv']
PROTOCOL=['analysis_protocol_v1.txt','amendment_01_before_fit.txt','implementation_note_02.txt',
          'implementation_note_03_after_review.txt','exploratory_extension_04_20261026.txt',
          'implementation_note_05_logit_ranking.txt']
FIGURE_RECORDS=['figure_provenance.json','figure_qa.json','render_bounds_checks.json',
                'feature_extension_provenance.json','threshold_extension_provenance.json',
                'threshold_extension_qa.json','supplement_extension_provenance.json']
CAPTIONS=['figure_captions.json','feature_extension_captions.json',
          'threshold_extension_captions.json','supplement_extension_captions.json']
SOURCE_ARCHIVE='VascTransport_v2_source.zip'
REPRO_ARCHIVE='VascTransport_v2_reproducibility.zip'
# Serialized local caches are regenerated from the bundled reviewable text inputs.
# They are not suitable submission attachments under the screening request.
FORBIDDEN_CACHE_SUFFIXES={'.pkl','.pickle','.joblib','.pyc','.pyo','.npy','.npz'}
CACHE_NAMES=[f'data/{acc}_{rep}.pkl' for acc in ['GSE100927','GSE43292','GSE28829']
             for rep in ['rank','native']]

def sha(path):
    h=hashlib.sha256()
    with path.open('rb') as f:
        for chunk in iter(lambda:f.read(2**20),b''):h.update(chunk)
    return h.hexdigest()

def checked_files(names):
    files=[]
    for name in sorted(set(names)):
        if Path(name).suffix.lower() in FORBIDDEN_CACHE_SUFFIXES:
            raise ValueError(f'Serialized cache is excluded from submission archives: {name}')
        path=(ROOT/name).resolve()
        if not path.is_relative_to(ROOT.resolve()) or not path.is_file():
            raise ValueError(f'Missing/unsafe allowlisted file: {name}')
        if path.is_symlink():raise ValueError(f'Symlink not allowed: {name}')
        files.append(path)
    return files

def archive(path,names,kind):
    files=checked_files(names)
    # Portal metadata stays local; public assembly needs only corresponding emails.
    metadata=json.loads((ROOT/'manuscript/author_metadata.json').read_text(encoding='utf8'))
    metadata={key:metadata[key] for key in ['authors','affiliations','equal_contribution_statement',
              'corresponding_authors','submitting_author','title','license']}
    for author in metadata['authors']:
        if not author.get('corresponding'):author.pop('email',None)
    public_metadata=(json.dumps(metadata,ensure_ascii=False,indent=2)+'\n').encode('utf8')
    def content(p):
        return public_metadata if p == (ROOT/'manuscript/author_metadata.json').resolve() else p.read_bytes()
    manifest={'created_utc':datetime.datetime.now(datetime.timezone.utc).isoformat(),
              'version':'V2 exploratory figure expansion',
              'kind':kind,'status':'Submission materials; this archive does not establish submission acceptance or public posting',
              'figure_inventory':{'main_figures':11,'main_panels':46,'supplementary_figures':2,'supplementary_panels':11},
              'license_note':'CC BY 4.0 authorized for manuscript; original public data retain source conditions. This archive does not relicense third-party material.',
              'excluded':'Third-party article full texts, private author correspondence/authorization, credentials, unrelated projects, caches and benchmark source PDFs.',
              'cache_policy':'No serialized Python or array caches are included. Run code/acquire.py to download and verify the six omitted official source inputs, then code/prepare.py to regenerate rank/native matrices locally.',
              'official_input_distribution':'Six large official GEO expression/platform files are omitted. Their exact URLs, byte sizes and SHA256 hashes are retained in evidence/expected_inputs_v1.json; online acquisition is required in a fresh extraction.',
              'metadata_transformation':'Noncorresponding author emails and internal authorization fields excluded from packaged author_metadata.json; local portal record is unchanged.',
              'files':{str(p.relative_to(ROOT)).replace('\\','/'):{'bytes':len(content(p)),'sha256':hashlib.sha256(content(p)).hexdigest()} for p in files}}
    with zipfile.ZipFile(path,'w',zipfile.ZIP_DEFLATED,compresslevel=6) as z:
        for p in files:z.writestr(str(p.relative_to(ROOT)).replace('\\','/'),content(p))
        z.writestr('RELEASE_MANIFEST.json',json.dumps(manifest,indent=2)+'\n')
    with zipfile.ZipFile(path) as z:
        forbidden=[name for name in z.namelist() if Path(name).suffix.lower() in FORBIDDEN_CACHE_SUFFIXES]
        if forbidden:raise ValueError('Serialized caches in archive: '+', '.join(forbidden))
        bad=z.testzip()
        if bad:raise ValueError('ZIP CRC failure: '+bad)
        saved=json.loads(z.read('RELEASE_MANIFEST.json'))
        for name,record in saved['files'].items():
            data=z.read(name)
            if len(data)!=record['bytes'] or hashlib.sha256(data).hexdigest()!=record['sha256']:
                raise ValueError('Archived byte mismatch: '+name)
    return {'path':str(path.relative_to(ROOT)).replace('\\','/'),'files':len(files),
            'bytes':path.stat().st_size,'sha256':sha(path),'crc_and_file_hashes_verified':True,
            'serialized_cache_members':[]}

def smoke(path,compile_pdf):
    log=[]
    # Short Windows path avoids staging/compiler MAX_PATH failures.
    temp_parent=Path(tempfile.gettempdir()).anchor if sys.platform=='win32' else None
    with tempfile.TemporaryDirectory(prefix='vt_',dir=temp_parent) as temp:
        target=Path(temp)
        with zipfile.ZipFile(path) as z:z.extractall(target)
        if any((target/name).exists() for name in CACHE_NAMES):
            raise ValueError('Relocated archive unexpectedly contains serialized caches')
        input_records=json.loads((target/'evidence/expected_inputs_v1.json').read_text(encoding='utf8'))
        absent_inputs=[str(record['path']).replace('\\','/') for record in input_records
                       if not (target/str(record['path']).replace('\\','/')).exists()]
        if len(absent_inputs)!=6:raise ValueError('Compact smoke must start with all six official inputs absent')
        commands=[['code/acquire.py'],['code/prepare.py'],['code/validate_results.py'],
                  ['code/validate_exploratory.py'],['code/export_descriptive_scores.py'],
                  ['code/plot_figures.py'],['code/plot_feature_extension.py'],
                  ['code/plot_threshold_extension.py'],['code/plot_supplement_extension.py'],
                  ['code/assemble_figure_catalog.py'],['code/build_supplement.py'],
                  ['code/build_exploratory_supplement.py'],
                  ['code/assemble.py']+(['--compile'] if compile_pdf else [])]
        immutable=['requirements.txt','evidence/acquisition.json','evidence/expected_inputs_v1.json',
                   'evidence/environment.json','evidence/analysis_environment_original.json','manuscript/author_metadata.json',
                   'evidence/figure_revision_20261026/pre_expansion/manifest.json']
        # The extension validator compares core hashes from this manifest;
        # old source/PDF snapshots are intentionally not needed or included.
        frozen=json.loads((target/immutable[-1]).read_text(encoding='utf8'))['sha256']
        immutable += [name for name in frozen if name.startswith('results/')]
        before={name:sha(target/name) for name in immutable}
        for command in commands:
            result=subprocess.run([sys.executable]+command,cwd=target,capture_output=True,text=True,
                                  encoding='utf8',errors='replace',timeout=300)
            log.append({'command':['python']+command,'exit_code':result.returncode,'output':result.stdout,'stderr':result.stderr})
            if result.returncode:break
        preserved={name:sha(target/name)==value for name,value in before.items()}
        acquisition_path=target/'evidence/input_verification_latest.json'
        acquisition=json.loads(acquisition_path.read_text(encoding='utf8')) if acquisition_path.exists() else {}
        acquired_online=(acquisition.get('offline') is False and acquisition.get('passed') is True
                         and len(acquisition.get('results',[]))==6
                         and all(x.get('status')=='downloaded_and_verified' for x in acquisition['results']))
        regenerated={name:{'exists':(target/name).is_file(),
                           'matches_canonical_cache':(target/name).is_file() and sha(target/name)==sha(ROOT/name)}
                     for name in CACHE_NAMES}
        pdf_checks=[]
        if compile_pdf and len(log)==len(commands) and all(x['exit_code']==0 for x in log):
            import fitz
            for name in ['main','supplement']:
                reference=fitz.open(ROOT/'manuscript'/f'{name}.pdf')
                rebuilt=fitz.open(target/'manuscript'/f'{name}.pdf')
                pages=[]
                for i,(a,b) in enumerate(zip(reference,rebuilt),1):
                    ap=a.get_pixmap(dpi=108,alpha=False);bp=b.get_pixmap(dpi=108,alpha=False)
                    pages.append({'page':i,'text_identical':a.get_text()==b.get_text(),
                                  'dimensions_identical':tuple(a.rect)==tuple(b.rect),
                                  'pixels_108dpi_identical':ap.width==bp.width and ap.height==bp.height and ap.samples==bp.samples})
                pdf_checks.append({'document':name,'reference_pages':len(reference),'rebuilt_pages':len(rebuilt),
                                   'passed':len(reference)==len(rebuilt) and all(all(p[k] for k in ['text_identical','dimensions_identical','pixels_108dpi_identical']) for p in pages),
                                   'pages':pages})
                reference.close();rebuilt.close()
        passed=(len(log)==len(commands) and all(x['exit_code']==0 for x in log) and all(preserved.values())
                and all(x['matches_canonical_cache'] for x in regenerated.values())
                and acquired_online
                and (not compile_pdf or len(pdf_checks)==2 and all(x['passed'] for x in pdf_checks)))
    return {'passed':passed,'scope':'Fresh short-path extraction in current installed environment; all six official inputs downloaded and hash-verified before local preparation. Exploratory validation includes two limited fitted-panel replays. No fresh dependency installation, full model refit or bootstrap regeneration.',
            'pdf_compilation_requested':compile_pdf,'immutable_source_records_preserved':preserved,
            'official_inputs_absent_at_start':absent_inputs,'online_acquisition':acquisition,
            'all_six_downloaded_and_verified':acquired_online,
            'cache_regeneration':regenerated,'pdf_comparison':pdf_checks,'commands':log}

def inventories():
    common=['README.txt','requirements.txt','requirements-qa.txt']
    source=common+['manuscript/'+x for x in MANUSCRIPT]
    source += [f'figures/Figure_{i}.{ext}' for i in range(1,12) for ext in ['pdf','svg','png']]
    source += [f'figures/Figure_S{i}.{ext}' for i in [1,2] for ext in ['pdf','svg','png']]
    source += ['figures/'+x for x in CAPTIONS]
    repro=source+['code/'+x for x in CODE]+['evidence/'+x for x in EVIDENCE]+['results/'+x for x in RESULTS]
    repro += ['protocol/'+x for x in PROTOCOL]
    repro += [f'data/{acc}_samples.csv' for acc in ['GSE100927','GSE43292','GSE28829']]
    repro += ['figures/'+x for x in FIGURE_RECORDS]
    return source,repro

def main():
    parser=argparse.ArgumentParser(description=__doc__)
    parser.add_argument('--smoke',action='store_true')
    parser.add_argument('--compile',action='store_true',help='Compile PDFs in relocated smoke copy (requires --smoke)')
    parser.add_argument('--check',action='store_true',help='Validate allowlists only; do not write archives or run analyses')
    args=parser.parse_args()
    if args.compile and not args.smoke:parser.error('--compile requires --smoke')
    if args.check and (args.smoke or args.compile):parser.error('--check cannot run smoke/compilation')
    source,repro=inventories()
    if args.check:
        for kind,names in [('source',source),('reproducibility',repro)]:
            checked_files(names)
            print(kind,':',len(set(names)),'explicitly allowlisted files present')
        return
    delivery=ROOT/'delivery';delivery.mkdir(exist_ok=True)
    expected={'main_figures':11,'main_panels':46,'supplementary_figures':2,'supplementary_panels':11}
    catalog=json.loads((ROOT/'figures/figure_provenance.json').read_text(encoding='utf8'))
    if any(catalog.get(k)!=v for k,v in expected.items()):raise ValueError('Figure catalog is not the reviewed V2 inventory')
    report={'created_utc':datetime.datetime.now(datetime.timezone.utc).isoformat(),
            'version':'V2; V1 delivery filenames are preserved','figure_inventory':expected,
            'archives':[archive(delivery/SOURCE_ARCHIVE,source,'Standalone LaTeX sources and editable figures'),
                        archive(delivery/REPRO_ARCHIVE,repro,'Compact reproducibility code, outputs and documents; six official inputs reacquired online')]}
    if args.smoke:report['relocation_smoke']=smoke(delivery/REPRO_ARCHIVE,args.compile)
    (delivery/'package_audit_v2.json').write_text(json.dumps(report,indent=2),encoding='utf8')
    print(json.dumps({'archives':report['archives'],'smoke_passed':report.get('relocation_smoke',{}).get('passed')},indent=2))
    if args.smoke and not report['relocation_smoke']['passed']:raise SystemExit(1)

if __name__=='__main__':main()
```
