## Supplementary material for "Anatomical comparison populations and threshold transport in vascular transcriptomic classification": Reproducibility archive: code, provenance and results: plot_feature_extension.py

```
"""Exploratory figures 7/8 from saved feature-control results; no fitting.

Writes only its own figures, captions and provenance. The coordinator merges
captions into the integrated manuscript after reviewing all additions.
"""
from pathlib import Path
import datetime, hashlib, json
import numpy as np
import pandas as pd
import matplotlib
matplotlib.use('Agg')
import matplotlib.pyplot as plt
from matplotlib.lines import Line2D
from matplotlib.colors import ListedColormap, BoundaryNorm
from matplotlib.ticker import NullLocator
from PIL import Image
import fitz
from sklearn.metrics import roc_auc_score, balanced_accuracy_score

ROOT=Path(__file__).resolve().parents[1]
OUT=ROOT/'figures'; OUT.mkdir(exist_ok=True)
BLUE, ORANGE, GRAY, RED='#1765A3','#D38B32','#87929E','#B54747'
W=17/2.54
plt.rcParams.update({'font.family':'DejaVu Sans','font.size':8,'axes.titlesize':9,
    'axes.labelsize':8,'xtick.labelsize':8,'ytick.labelsize':8,'legend.fontsize':8,
    'figure.titlesize':11,'axes.linewidth':.6,'lines.linewidth':1.2,'savefig.dpi':300,
    'pdf.fonttype':42,'ps.fonttype':42,'svg.fonttype':'none','axes.spines.top':False,
    'axes.spines.right':False,'axes.titlelocation':'left'})
inputs={};captions={};bounds=[];verification={}
def sha(p):return hashlib.sha256(p.read_bytes()).hexdigest()
def read(path):
    p=ROOT/path;inputs[path]=sha(p);return pd.read_csv(p)
def panel(ax,letter,title):
    ax.set_title(letter+'  '+title,loc='left',fontweight='bold',pad=9)
    ax.tick_params(width=.6,length=3)
def grid(ax,axis='x'):
    ax.grid(axis=axis,color='#E6E9EC',linewidth=.55);ax.set_axisbelow(True)
def save_figure(fig,number,title,mapping,caption,source_paths):
    stem=f'Figure_{number}';fig.canvas.draw()
    renderer=fig.canvas.get_renderer();fb=fig.bbox;outside=[]
    texts=list(fig.texts)
    for ax in fig.axes:
        texts += list(ax.texts)+[ax.title,ax._left_title,ax.xaxis.label,ax.yaxis.label]
        xlim=sorted(ax.get_xlim());ylim=sorted(ax.get_ylim())
        texts += [t for t in ax.get_xticklabels() if xlim[0]-1e-8<=t.get_position()[0]<=xlim[1]+1e-8]
        texts += [t for t in ax.get_yticklabels() if ylim[0]-1e-8<=t.get_position()[1]<=ylim[1]+1e-8]
        legend=ax.get_legend()
        if legend is not None:texts+=legend.get_texts()
    for text in texts:
        if text.get_visible() and text.get_text():
            b=text.get_window_extent(renderer)
            if b.x0<fb.x0-1 or b.y0<fb.y0-1 or b.x1>fb.x1+1 or b.y1>fb.y1+1:outside.append(text.get_text())
    for ext in ['pdf','svg','png']:fig.savefig(OUT/f'{stem}.{ext}',dpi=300,facecolor='white')
    bounds.append({'figure':stem,'text_outside_canvas':outside})
    captions[stem]={'title':title,'panel_count':len(mapping),'panels':mapping,
        'caption':caption,'source_tables':source_paths,'width_cm':17,
        'formats':['PDF','SVG','PNG_300dpi'],'analysis_status':'Exploratory; added after original results'}
    plt.close(fig)

fold=read('results/exploratory_features/representation_fold_metrics.csv')
external=read('results/exploratory_features/representation_external_metrics.csv')
external_predictions=read('results/exploratory_features/representation_external_predictions.csv')
primary=read('results/primary_fold_metrics.csv').query("model == 'ridge200'")
primary_ext=read('results/external_metrics.csv').query("model == 'ridge200'").set_index('dataset')
stability=read('results/exploratory_features/feature_stability.csv')
members=read('results/exploratory_features/primary_fold_features.csv')
jaccard=read('results/exploratory_features/feature_jaccard.csv')
coef=read('results/frozen_primary_coefficients.csv')
panelgenes=read('results/exploratory_features/random_panel_genes.csv')
randoms=sorted(fold.loc[fold.panel.str.startswith('random_'),'panel'].unique())
assert len(randoms)==20 and len(fold)==630 and len(external)==42
assert external.ranking_score.eq('linear_logit').all()
assert len(external_predictions)==21*(64+29)
for (panel_name,dataset),rows in external_predictions.groupby(['panel','dataset']):
    saved=external.loc[(external.panel==panel_name)&(external.dataset==dataset)].iloc[0]
    assert abs(roc_auc_score(rows.y,rows.linear_logit)-saved.auc)<1e-12
    assert abs(balanced_accuracy_score(rows.y,rows.score>=.5)-saved.balanced_accuracy)<1e-12
    if dataset=='GSE43292':
        paired=rows.pivot(index='patient',columns='y',values='linear_logit')
        assert paired.shape==(32,2) and not paired.isna().any().any()
        difference=paired[1]-paired[0]
        concordance=((difference>0).sum()+.5*(difference==0).sum())/32
        assert abs(concordance-saved.pair_concordance)<1e-12
assert panelgenes.groupby('panel').gene.nunique().eq(200).all()
assert len(primary)==30 and primary.groupby('repeat').size().eq(3).all()
means=fold.groupby('panel').auc.mean()
repeats=fold.groupby(['panel','repeat']).auc.mean()
primary_auc=float(primary.auc.mean())
primary_repeat=primary.groupby('repeat').auc.mean()
ext=external.pivot(index='panel',columns='dataset',values='auc')
ba=external.pivot(index='panel',columns='dataset',values='balanced_accuracy')
pair_concordance=external.query("dataset == 'GSE43292'").set_index('panel').pair_concordance
verification.update(random_panels=20,source_folds_per_panel=30,source_units_per_repeat=95,
    random_source_auc_min=float(means.loc[randoms].min()),random_source_auc_max=float(means.loc[randoms].max()),
    random_source_auc_median=float(means.loc[randoms].median()),primary_source_auc=primary_auc,
    next200_source_auc=float(means.loc['next200']),no_winner_selected=True)
verification.update(
    external_metrics_recomputed_from_saved_predictions=True,
    ranking_score='linear_logit; sigmoid probabilities retained for the 0.5 rule',
    random_external_summary={dataset:{metric:{stat:float(value) for stat,value in
        external.loc[external.panel.isin(randoms)&external.dataset.eq(dataset),metric].agg(['min','median','max']).items()}
        for metric in ['auc','balanced_accuracy']}
        for dataset in ['GSE43292','GSE28829']},
    random18_GSE43292=external.loc[(external.panel=='random_18')&external.dataset.eq('GSE43292'),
        ['auc','balanced_accuracy','pair_concordance','probability_unique','linear_logit_unique','saturated_probability_count']].iloc[0].to_dict())

# Figure 7: performance equality at source does not imply equal external behavior.
title7='Gene-panel controls separate source ranking from external transport'
fig=plt.figure(figsize=(W,7.25),layout='constrained')
gs=fig.add_gridspec(2,2,hspace=.13,wspace=.10)
axes=[fig.add_subplot(gs[i//2,i%2]) for i in range(4)]
fig.suptitle(title7,x=.015,ha='left',fontweight='bold')
ax=axes[0];panel(ax,'A','Near-ceiling source ranking\nwith different gene panels')
jitter=np.array([-.18,-.06,.06,.18]*5)
vals=means.loc[randoms].to_numpy()
ax.scatter(vals,2+jitter,s=22,color=GRAY,edgecolors='white',linewidths=.4,zorder=3)
for y,key,col,marker in [(1,'next200',ORANGE,'s'),(0,'primary',BLUE,'D')]:
    rr=repeats.loc[key] if key!='primary' else primary_repeat
    mu=means.loc[key] if key!='primary' else primary_auc
    ax.errorbar(mu,y,xerr=[[mu-rr.min()],[rr.max()-mu]],fmt=marker,color=col,ms=5,capsize=3,zorder=4)
ax.set_yticks([0,1,2],['Primary 200','Next-ranked 200','Random 200\n(20 fixed draws)'])
ax.set(xlim=(.9894,1.0004),ylim=(-.65,2.6),xlabel='Source mean held-out-fold AUROC\n(zoomed scale)')
ax.set_xticks([.990,.995,1.000],['0.990','0.995','1.000']);grid(ax)
ax.text(.991,-.47,'Bars: 10-repeat range',color='#505A64',fontsize=8)

ax=axes[1];panel(ax,'B','The same panel across\ndistinct tissue contrasts')
for name in randoms:
    v=[means.loc[name],ext.loc[name,'GSE43292'],ext.loc[name,'GSE28829']]
    ax.plot(range(3),v,color=GRAY,alpha=.50,lw=.65,marker='o',ms=2.5,zorder=2)
for name,col,mark,values in [
    ('Next-ranked 200',ORANGE,'s',[means.loc['next200'],ext.loc['next200','GSE43292'],ext.loc['next200','GSE28829']]),
    ('Primary 200',BLUE,'D',[primary_auc,primary_ext.loc['GSE43292','auc'],primary_ext.loc['GSE28829','auc']])]:
    ax.plot(range(3),values,color=col,marker=mark,ms=4.5,lw=1.7,zorder=5)
ax.set_xticks([0,1,2],['Source\nheld-out folds','Paired lesion\nGSE43292','Histology\nGSE28829'])
ax.set(ylim=(.44,1.035),xlim=(-.12,2.12),ylabel='AUROC');ax.set_yticks([.5,.6,.7,.8,.9,1.0]);grid(ax,'y')
handles=[Line2D([0],[0],color=BLUE,marker='D',ms=4,label='Primary 200'),
    Line2D([0],[0],color=ORANGE,marker='s',ms=4,label='Next-ranked 200'),
    Line2D([0],[0],color=GRAY,marker='o',ms=3,label='Random 200')]
ax.legend(handles=handles,loc='lower left',frameon=True,facecolor='white',edgecolor='none',framealpha=.95,
    handlelength=1.25,borderpad=.3,labelspacing=.22)

ax=axes[2];panel(ax,'C','Within-patient ordering\nand marginal tissue ranking')
ax.scatter(ext.loc[randoms,'GSE43292'],pair_concordance.loc[randoms],s=25,color=GRAY,edgecolors='white',linewidths=.4)
ax.scatter(ext.loc['next200','GSE43292'],pair_concordance.loc['next200'],s=42,color=ORANGE,marker='s',edgecolors='white',linewidths=.6,zorder=4)
ax.scatter(primary_ext.loc['GSE43292','auc'],primary_ext.loc['GSE43292','pair_concordance'],s=45,color=BLUE,marker='D',edgecolors='white',linewidths=.6,zorder=5)
ax.set(xlim=(.575,.90),ylim=(.50,.97),xlabel='Marginal tissue AUROC (GSE43292)',ylabel='Plaque > intact concordance (32 pairs)')
ax.set_xticks([.6,.7,.8,.9]);ax.set_yticks([.5,.6,.7,.8,.9]);grid(ax,'both')

ax=axes[3];panel(ax,'D','Unchanged-threshold\nbalanced accuracy')
for y,dataset in enumerate(['GSE43292','GSE28829']):
    vals=ba.loc[randoms,dataset].to_numpy()
    # Stack ties horizontally in y so every random panel remains visible.
    for value in np.unique(vals):
        which=np.flatnonzero(vals==value);off=np.linspace(-.16,.16,len(which)) if len(which)>1 else np.zeros(1)
        ax.scatter(np.repeat(value,len(which)),y+off,s=17,color=GRAY,edgecolors='white',linewidths=.35,zorder=3)
    ax.scatter(primary_ext.loc[dataset,'balanced_accuracy'],y-.30,s=43,marker='D',color=BLUE,zorder=4)
    ax.scatter(ba.loc['next200',dataset],y+.30,s=40,marker='s',color=ORANGE,zorder=4)
ax.axvline(.5,color='#A4ACB5',lw=.8,ls='--',zorder=0)
ax.set_yticks([0,1],['Plaque / intact','Advanced / early'])
ax.set(ylim=(-.55,1.65),xlim=(.45,1.02),xlabel='Balanced accuracy at 0.5');ax.set_xticks([.5,.6,.7,.8,.9,1.]);grid(ax)
ax.text(.985,1.47,'20 random panels per contrast',ha='right',fontsize=8,color='#505A64',
    bbox=dict(facecolor='white',edgecolor='none',alpha=1,pad=1.5))
save_figure(fig,7,title7,{
    'A':'Source mean-fold AUROC for outcome-independent random panels and the disjoint next-ranked panel',
    'B':'Every fixed panel followed from source discrimination to two distinct external contrasts',
    'C':'Marginal tissue AUROC versus within-patient ordering for each fixed panel',
    'D':'External balanced accuracy at the unchanged technical threshold'},
    'Exploratory analyses added after inspection of the original results. Twenty 200-gene panels were drawn once without using outcomes (seed 20261026); all draws are retained. The next-ranked control selects training ranks 201-400 after excluding each training set\'s top 200 genes. All trainable rules use source-only scaling and the same fixed ridge specification. New-control ranking metrics use raw linear logits to avoid artificial ties from floating-point sigmoid saturation; probabilities remain the inputs to threshold and Brier diagnostics. A, Each random point is its panel\'s mean across the original 30 held-out folds. Bars for primary and next-ranked panels show the ten-repetition range, not a confidence interval; the horizontal axis is zoomed. B, Each grey line follows the same random panel; source cross-validation and external full-source fits are different evaluations, and the three contrasts are not interchangeable endpoints. C, For each panel, marginal tissue AUROC and within-patient plaque > intact concordance answer different comparison questions in GSE43292; half credit is assigned to ties. No external winner is selected. D, Tied random-panel values are stacked only for visibility. The unchanged 0.5 rule is a technical classifier diagnostic, not a validated clinical decision threshold. Panel draws and overlapping folds are not independent cohorts; variations across panels are not confidence intervals.',
    ['results/exploratory_features/representation_fold_metrics.csv','results/exploratory_features/representation_external_metrics.csv',
     'results/exploratory_features/random_panel_genes.csv','results/primary_fold_metrics.csv','results/external_metrics.csv',
     'results/exploratory_features/representation_external_predictions.csv'])

# Figure 8: membership uncertainty is distinct from score discrimination.
assert len(stability)==15235 and stability.gene.nunique()==15235
assert len(members)==6000 and members.groupby('fit_id').size().eq(200).all()
recount=members.groupby('gene').fit_id.nunique().reindex(stability.gene,fill_value=0).to_numpy()
assert np.array_equal(recount,stability.selected_folds.to_numpy())
freq=stability.selected_folds.value_counts().reindex(range(31),fill_value=0)
assert freq.sum()==15235 and freq.loc[0]==14616 and freq.loc[30]==45
assert int((stability.selected_folds>=24).sum())==103
fit_ids=sorted(members.fit_id.unique());matrix=np.eye(30)
sets={key:set(frame.gene) for key,frame in members.groupby('fit_id')}
for row in jaccard.itertuples():
    value=len(sets[row.fit_a]&sets[row.fit_b])/len(sets[row.fit_a]|sets[row.fit_b])
    assert abs(value-row.jaccard)<1e-12
    ia,ib=fit_ids.index(row.fit_a),fit_ids.index(row.fit_b);matrix[ia,ib]=matrix[ib,ia]=value
display=(coef.assign(abs_coef=coef.coefficient.abs()).sort_values(['abs_coef','gene'],ascending=[False,True]).head(20))
display_genes=display.gene.tolist()
signed=np.zeros((20,30))
for i,gene in enumerate(display_genes):
    for row in members[members.gene==gene].itertuples():signed[i,fit_ids.index(row.fit_id)]=np.sign(row.coefficient)
counts=(signed!=0).sum(1)
for gene,n in zip(display_genes,counts):assert n==stability.set_index('gene').loc[gene,'selected_folds']
title8='Feature membership varies across fits despite stable source ranking'
fig=plt.figure(figsize=(W,8.0),layout='constrained')
gs=fig.add_gridspec(2,2,height_ratios=[1,1.55],hspace=.13,wspace=.08)
axA=fig.add_subplot(gs[0,0]);axB=fig.add_subplot(gs[0,1]);axC=fig.add_subplot(gs[1,:])
fig.suptitle(title8,x=.015,ha='left',fontweight='bold')
panel(axA,'A','Selection frequency across\nthe complete gene vocabulary')
axA.bar(range(31),freq.to_numpy(),color=[GRAY]+[BLUE]*30,width=.83,zorder=3)
axA.set_yscale('log');axA.set(ylim=(.8,60000),xlim=(-1,31),xlabel='Number of the 30 fits selecting a gene',ylabel='Number of genes (log scale)')
axA.set_xticks([0,5,10,15,20,25,30]);axA.set_yticks([1,10,100,1000,10000],['1','10','100','1,000','10,000'])
axA.yaxis.set_minor_locator(NullLocator());grid(axA,'y')
axA.text(2.0,30000,'14,616 never selected',fontsize=8,color='#505A64')
axA.text(29.8,15000,'619 selected at least once\n45 selected in every fit',ha='right',va='top',fontsize=8,color=BLUE,
    bbox=dict(facecolor='white',alpha=.95,edgecolor='none',pad=2))
panel(axB,'B','Overlap between the\n30 selected-gene sets')
im=axB.imshow(matrix,cmap='Blues',vmin=0,vmax=1,aspect='auto')
centers=np.arange(1,30,3);labels=[f'R{i}' for i in range(1,11)]
axB.set_xticks(centers,labels,rotation=90);axB.set_yticks(centers,labels)
for boundary in np.arange(2.5,29,3):
    axB.axhline(boundary,color='white',lw=.5);axB.axvline(boundary,color='white',lw=.5)
axB.set(xlabel='Repetition (three folds per group)',ylabel='Repetition')
cb=fig.colorbar(im,ax=axB,fraction=.05,pad=.02);cb.set_label('Jaccard overlap');cb.set_ticks([0,.5,1])
panel(axC,'C','Source-selected display: 20 largest full-source |coefficients|')
cmap=ListedColormap([ORANGE,'#EEF1F4',BLUE]);norm=BoundaryNorm([-1.5,-.5,.5,1.5],3)
axC.imshow(signed,cmap=cmap,norm=norm,aspect='auto',interpolation='none',extent=(-.5,29.5,19.5,-.5))
axC.set_xticks(centers,labels);axC.set_yticks(range(20),[f'{g} ({b:+.3f})' for g,b in zip(display_genes,display.coefficient)])
axC.set(xlabel='Repeated held-out fit; groups contain folds 1, 2 and 3',ylabel='Gene (frozen full-source coefficient)',xlim=(-.5,33.8),ylim=(20.55,-1.5))
for boundary in np.arange(2.5,29,3):axC.axvline(boundary,ymin=.06,ymax=.95,color='white',lw=1.1)
axC.text(31.65,-.95,'Fits',ha='center',fontsize=8,fontweight='bold')
for i,n in enumerate(counts):axC.text(31.65,i,f'{n}/30',ha='center',va='center',fontsize=8)
handles=[Line2D([0],[0],marker='s',ls='',color=BLUE,label='Positive coefficient',ms=5),
    Line2D([0],[0],marker='s',ls='',color=ORANGE,label='Negative coefficient',ms=5),
    Line2D([0],[0],marker='s',ls='',color='#DCE1E7',label='Not selected',ms=5)]
axC.legend(handles=handles,loc='lower center',bbox_to_anchor=(.5,-.18),ncol=3,frameon=False,
    handletextpad=.35,columnspacing=.9,borderaxespad=0)
verification.update(vocabulary=15235,never_selected=14616,selected_union=619,selected_in_all30=45,
    selected_at_least24=103,jaccard_offdiagonal_mean=float(jaccard.jaccard.mean()),
    jaccard_offdiagonal_min=float(jaccard.jaccard.min()),jaccard_offdiagonal_max=float(jaccard.jaccard.max()),
    named_gene_display='Top20 absolute coefficients from unchanged full-source primary model; alphabetical ties',
    displayed_genes=display_genes)
save_figure(fig,8,title8,{
    'A':'All 15235 genes, including genes never selected, grouped by selection count',
    'B':'Pairwise Jaccard overlap of the 30 source-fold feature sets',
    'C':'Membership and coefficient direction for a source-selected named-gene display'},
    'Exploratory feature-recovery analysis after the original results. The 30 primary source fits were replayed to recover selected genes and coefficients; held-out predictions were checked against the saved original predictions before interpretation. A, All 15,235 candidate genes are included; the count axis is logarithmic. Each fit selects 200 genes, with 619 distinct genes selected across fits, including 45 selected in every fit. B, Jaccard overlap is intersection divided by union. Three adjacent rows/columns constitute the three folds of a repetition; diagonal values equal one. Off-diagonal comparisons reuse training observations and are descriptive, not independent replications or confidence intervals. C, Named genes are the 20 largest absolute coefficients of the unchanged full-source primary model, selected without using external outcomes; full-source coefficients appear beside names. Each tile shows absence or coefficient sign in one recovered fold fit, with selection counts at right. These high-weight genes range from intermittent to persistent membership, while A-B retain the full vocabulary. No stable signature is refitted or revalidated using these summaries. Correlated genes and fold-specific standardization prevent interpreting coefficient magnitude or selection frequency as causal biological importance.',
    ['results/exploratory_features/feature_stability.csv','results/exploratory_features/primary_fold_features.csv',
     'results/exploratory_features/feature_jaccard.csv','results/frozen_primary_coefficients.csv'])

(OUT/'feature_extension_captions.json').write_text(json.dumps(captions,ensure_ascii=False,indent=2),encoding='utf8')
pdf_checks=[]
for i in [7,8]:
    p=OUT/f'Figure_{i}.pdf'
    with fitz.open(p) as doc:
        page=doc[0];spans=[s for b in page.get_text('dict')['blocks'] if b['type']==0 for l in b['lines'] for s in l['spans'] if s['text'].strip()]
        minimum=min(s['size'] for s in spans)
        outside=[s['text'] for s in spans if not page.rect.contains(fitz.Rect(s['bbox']))]
        assert len(doc)==1 and minimum>=7.99 and not outside,(i,minimum,outside)
    with Image.open(OUT/f'Figure_{i}.png') as im:
        pixels=im.size;dpi=im.info.get('dpi')
    pdf_checks.append(dict(figure=i,pdf_sha256=sha(p),minimum_glyph_size_pt=minimum,png_pixels=pixels,png_dpi=dpi,
        outside_pdf_text=outside,width_cm=17))
assert not any(x['text_outside_canvas'] for x in bounds),bounds
assert all(sha(ROOT/name)==digest for name,digest in inputs.items())
audit={'recorded_utc':datetime.datetime.now(datetime.timezone.utc).isoformat(),'passed':True,
    'scope':'Saved-result figure construction and independent count/overlap checks; no model fitting',
    'figures':2,'panels':7,'input_sha256':inputs,'numeric_checks':verification,'bounds':bounds,'pdf_checks':pdf_checks,
    'output_sha256':{f'Figure_{i}.{ext}':sha(OUT/f'Figure_{i}.{ext}') for i in [7,8] for ext in ['pdf','svg','png']}}
(OUT/'feature_extension_provenance.json').write_text(json.dumps(audit,ensure_ascii=False,indent=2),encoding='utf8')
print(json.dumps({'passed':True,'figures':2,'panels':7,'source_auc_random_range':[verification['random_source_auc_min'],verification['random_source_auc_max']],
                  'minimum_pdf_glyph_sizes':[x['minimum_glyph_size_pt'] for x in pdf_checks]},indent=2))
```
