## Supplementary material for "Anatomical comparison populations and threshold transport in vascular transcriptomic classification": Reproducibility archive: code, provenance and results: plot_figures.py

```
"""Publication figures from saved VascTransport results; no model refitting.

Run from any directory: python code/plot_figures.py
Every numeric panel uses saved records, or an explicitly documented algebraic
aggregation of those records. Range bars summarize repeated splits, not CIs.
"""
from pathlib import Path
import json
import hashlib
import string
import textwrap
import numpy as np
import pandas as pd
import matplotlib
matplotlib.use("Agg")
import matplotlib.pyplot as plt
from matplotlib.patches import FancyBboxPatch, FancyArrowPatch
from matplotlib.ticker import PercentFormatter, MaxNLocator, ScalarFormatter, FuncFormatter, NullFormatter
from sklearn.metrics import roc_curve, roc_auc_score
from PIL import Image, ImageOps, ImageDraw

ROOT = Path(__file__).resolve().parents[1]
RES = ROOT / "results"
OUT = ROOT / "figures"
OUT.mkdir(exist_ok=True)
W = 17 / 2.54
BEDS = ["Carotid", "Femoral", "Infrapopliteal"]
BSHORT = {"Carotid": "C", "Femoral": "F", "Infrapopliteal": "I"}
BNAMES = ["Carotid", "Femoral", "Infra."]
MODELS = ["prevalence", "bed_sex", "C1Q", "Wu6", "ridge200", "centroid200", "forest200"]
EXTMODELS = ["C1Q", "Wu6", "ridge200", "centroid200", "forest200"]
PNAMES = {"prevalence": "Intercept", "bed_sex": "Bed + sex", "C1Q": "C1Q ranks", "Wu6": "Wu6 LR", "ridge200": "Ridge-200", "centroid200": "Centroid-200", "forest200": "Forest-200"}
COLORS = {"prevalence": "#89939D", "bed_sex": "#7D5BA6", "C1Q": "#48A9A6", "Wu6": "#E6A532", "ridge200": "#1765A3", "centroid200": "#7454A3", "forest200": "#C75A3A"}
BLUE, ORANGE, GRAY, RED = "#1765A3", "#D38B32", "#77818C", "#B54747"
BEDCOLORS = dict(zip(BEDS, ["#1765A3", "#D38B32", "#4A9986"]))
plt.rcParams.update({
    "font.family": "DejaVu Sans", "font.size": 8, "axes.titlesize": 9,
    "axes.labelsize": 8, "xtick.labelsize": 8, "ytick.labelsize": 8,
    "legend.fontsize": 8, "figure.titlesize": 11, "axes.linewidth": .6,
    "lines.linewidth": 1.2, "savefig.dpi": 300, "pdf.fonttype": 42,
    "ps.fonttype": 42, "svg.fonttype": "none", "axes.spines.top": False,
    "axes.spines.right": False, "axes.titlelocation": "left",
})

inputs = {}
def read(name):
    path = RES / (name + ".csv")
    inputs[str(path.relative_to(ROOT)).replace("\\", "/")] = hashlib.sha256(path.read_bytes()).hexdigest()
    return pd.read_csv(path)

units = read("primary_analysis_units")
folds = read("primary_fold_metrics")
cells = read("primary_pair_cells")
transport = read("transport_metrics")
em = read("external_metrics")
ep = read("external_predictions")
pairs = read("external_pairs")
sens = read("sensitivity_all_fold_metrics")
null = read("permutation_nulls")
sub = read("full_pipeline_subsampling")
amb = read("ambiguous_label_profile_similarity")
unmatched = read("unmatched_within_stratum_similarity")
captions = {}
render_checks = []

def panel(ax, letter, title):
    width = 58 if ax.get_subplotspec().colspan.stop - ax.get_subplotspec().colspan.start == 2 else 27
    ax.set_title(f"{letter}  " + textwrap.fill(title, width=width, subsequent_indent="   "), fontweight="bold", pad=9)
    ax.tick_params(width=.6, length=3)

def grid(ax, axis="x"):
    ax.grid(axis=axis, color="#E6E9EC", linewidth=.55, zorder=0)
    ax.set_axisbelow(True)

def layout(title, n=4, tall=False):
    fig = plt.figure(figsize=(W, 8.5 if tall else 6.4), layout="constrained")
    gs = fig.add_gridspec(3 if n == 5 else 2, 2, hspace=.12, wspace=.10)
    axes = [fig.add_subplot(gs[i//2, i%2]) for i in range(min(n, 4))]
    if n == 5:
        axes.append(fig.add_subplot(gs[2, :]))
    fig.suptitle(textwrap.fill(title, width=68), x=.015, ha="left", fontweight="bold")
    return fig, axes

def finish(fig, number, title, panel_mapping, caption, sources):
    stem = f"Figure_{number}"
    fig.canvas.draw()
    renderer = fig.canvas.get_renderer()
    fb = fig.bbox
    candidates = list(fig.texts)
    for a in fig.axes:
        candidates += list(a.texts) + [a.title, a._left_title]
        if a.axison:
            candidates += [a.xaxis.label, a.yaxis.label]
            xlo,xhi=sorted(a.get_xlim());ylo,yhi=sorted(a.get_ylim())
            candidates += [t for t in a.get_xticklabels() if xlo-1e-10 <= t.get_position()[0] <= xhi+1e-10]
            candidates += [t for t in a.get_yticklabels() if ylo-1e-10 <= t.get_position()[1] <= yhi+1e-10]
        if a.get_legend() is not None:
            candidates += list(a.get_legend().get_texts())
        for table in a.tables:
            candidates += [c.get_text() for c in table.get_celld().values()]
    clipped=[]
    for artist in candidates:
        if artist.get_visible() and artist.get_text():
            bb=artist.get_window_extent(renderer)
            if bb.x0 < fb.x0-1 or bb.x1 > fb.x1+1 or bb.y0 < fb.y0-1 or bb.y1 > fb.y1+1:
                clipped.append(artist.get_text())
    render_checks.append({"figure":stem,"text_outside_canvas":sorted(set(clipped))})
    for ext in ("pdf", "svg", "png"):
        fig.savefig(OUT / f"{stem}.{ext}", dpi=300, facecolor="white")
    captions[stem] = {"title": title, "panel_count": len(panel_mapping), "panels": panel_mapping, "caption": caption, "source_tables": sources, "width_cm": 17, "formats": ["PDF", "SVG", "PNG_300dpi"]}
    plt.close(fig)

def heat(ax, matrix, xlabels, ylabels, fmt=".3f", vmin=0, vmax=1, cmap="Blues", xlabel=None, ylabel=None):
    im = ax.imshow(matrix, vmin=vmin, vmax=vmax, cmap=cmap, aspect="auto")
    ax.set_xticks(range(len(xlabels)), xlabels)
    ax.set_yticks(range(len(ylabels)), ylabels)
    for i in range(matrix.shape[0]):
        for j in range(matrix.shape[1]):
            val = matrix[i, j]
            if np.isfinite(val):
                ax.text(j, i, format(val, fmt), ha="center", va="center", color="white" if val > vmin + .68*(vmax-vmin) else "#142534", fontsize=8)
    if xlabel: ax.set_xlabel(xlabel)
    if ylabel: ax.set_ylabel(ylabel)
    return im

def mean_range(ax, groups, order, metric, color, offset=0, label=None):
    y = np.arange(len(order)) + offset
    z = groups.groupby(level=0)[metric].agg(["mean", "min", "max"]).reindex(order)
    ax.errorbar(z["mean"], y, xerr=np.array([z["mean"]-z["min"], z["max"]-z["mean"]]), fmt="o", ms=4, color=color, capsize=2, lw=.9, label=label, zorder=3)
    return z

def roc(ax, acc, model="ridge200"):
    d = ep[(ep.dataset == acc) & (ep.model == model)]
    fpr, tpr, _ = roc_curve(d.y, d.score)
    ax.plot(fpr, tpr, color=COLORS[model], lw=1.8)
    ax.plot([0, 1], [0, 1], color=GRAY, ls="--", lw=.8)
    row = em[(em.dataset == acc) & (em.model == model)].iloc[0]
    ax.text(.97, .05, f"AUROC {row.auc:.3f}\n95% CI {row.auc_low:.3f}–{row.auc_high:.3f}", transform=ax.transAxes, ha="right", va="bottom", fontsize=8)
    ax.set(xlim=(-.02, 1.02), ylim=(-.02, 1.03), xlabel="False-positive rate", ylabel="True-positive rate")
    ax.set_xticks([0, .5, 1]); ax.set_yticks([0, .5, 1]); grid(ax, "both")

def threshold_table(ax, acc):
    d = em[(em.dataset == acc) & em.model.isin(["Wu6", "ridge200", "forest200"])].set_index("model").reindex(["Wu6", "ridge200", "forest200"])
    cols = ["sensitivity", "specificity", "balanced_accuracy", "brier"]
    arr = d[cols].to_numpy()
    ax.axis("off")
    tab = ax.table(cellText=[[PNAMES[m]]+[f"{v:.3f}" for v in arr[i]] for i,m in enumerate(d.index)], colLabels=["Model", "Sens.", "Spec.", "Bal.\nacc.", "Brier"], loc="center", cellLoc="center", colWidths=[.28,.17,.17,.21,.17])
    tab.auto_set_font_size(False); tab.set_fontsize(8); tab.scale(1, 1.8)
    for (r,c),cell in tab.get_celld().items():
        cell.set_edgecolor("#D7DEE5"); cell.set_linewidth(.5)
        if r == 0:
            cell.set_facecolor("#EAF1F7"); cell.set_text_props(fontweight="bold"); cell.set_height(cell.get_height()*1.25)
        elif r == 2: cell.set_facecolor("#F0F6FA")
    return d

# FIGURE 1: population structure, analysis units and identity uncertainty.
fig, ax = layout("Study design and source-tissue structure")
panel(ax[0], "A", "Separate estimation and stress tests")
ax[0].axis("off")
boxes = [(.02,.64,.96,.24, "GSE100927: 104 deposited arrays\n95 full-label groups; 3 arterial beds\n10 × 3-fold source evaluation"),
         (.02,.32,.96,.23, "Freeze full-source model\n15,235 common genes; sample ranks\nNo target fitting or recalibration"),
         (.02,.02,.46,.27, "GSE43292\n32 patients\n64 tissues\nPlaque / intact"),
         (.53,.02,.45,.27, "GSE28829\n29 specimens\nAdvanced / early")]
for x,y,w,h,t in boxes:
    ax[0].add_patch(FancyBboxPatch((x,y),w,h,boxstyle="round,pad=.012",facecolor="#F0F5FA",edgecolor="#AEC2D3",lw=.8,transform=ax[0].transAxes))
    ax[0].text(x+w/2,y+h/2,t,ha="center",va="center",fontsize=8,transform=ax[0].transAxes)
for start,end in [((.5,.64),(.5,.56)),((.5,.32),(.25,.30)),((.5,.32),(.75,.30))]:
    ax[0].add_patch(FancyArrowPatch(start,end,arrowstyle="-|>",mutation_scale=9,color=GRAY,transform=ax[0].transAxes))
panel(ax[1], "B", "Deposits and conservative units")
counts = units.groupby(["bed","y"]).agg(arrays=("array_count","sum"),groups=("y","size")).reindex(pd.MultiIndex.from_product([BEDS,[0,1]]))
yy=np.arange(6)
ax[1].barh(yy-.17,counts.arrays,height=.3,color="#B8CADB",label="Arrays")
ax[1].barh(yy+.17,counts.groups,height=.3,color=BLUE,label="Label groups")
ax[1].set_yticks(yy,[f"{BNAMES[i//2]} {'control' if i%2==0 else 'disease'}" for i in range(6)])
for i,(a,g) in enumerate(zip(counts.arrays,counts.groups)):
    ax[1].text(a+.5,i-.17,str(a),va="center",fontsize=8);ax[1].text(g+.5,i+.17,str(g),va="center",fontsize=8)
ax[1].invert_yaxis(); ax[1].set(xlim=(0,36.5),xlabel="Records (not verified patients)");ax[1].legend(loc="lower right",frameon=False);grid(ax[1])
panel(ax[2], "C", "Sex distribution by source stratum")
male=units.assign(male=units.sex.eq("male")).groupby(["bed","y"]).male.mean()
x=np.arange(3)
for status,off,col,lab in [(0,-.18,GRAY,"Control"),(1,.18,BLUE,"Disease")]:
    vals=[male.loc[(b,status)] for b in BEDS]
    ax[2].bar(x+off,vals,width=.32,color=col,label=lab)
    for xx,v in zip(x+off,vals):ax[2].text(xx,v+.025,f"{100*v:.0f}%",ha="center",fontsize=8)
ax[2].set_xticks(x,BNAMES);ax[2].set(ylabel="Male proportion of label groups",ylim=(0,1.13));ax[2].yaxis.set_major_formatter(PercentFormatter(1));ax[2].legend(frameon=False,loc="lower left");grid(ax[2],"y")
panel(ax[3], "D", "Expression similarity ≠ identity")
for vals,col,label in [(unmatched.correlation.to_numpy(),GRAY,f"Other same-stratum pairs\n(n={len(unmatched):,})"),(amb.rank_profile_correlation.to_numpy(),BLUE,"Repeated labels (9 pairs)")]:
    vals=np.sort(vals);ax[3].step(vals,np.arange(1,len(vals)+1)/len(vals),where="post",color=col,label=label)
    if len(vals)==9:ax[3].scatter(vals,np.arange(1,10)/9,s=15,color=col,zorder=3)
ax[3].set(xlabel="Within-sample rank-profile correlation",ylabel="Empirical cumulative fraction",ylim=(0,1.05));ax[3].legend(fontsize=8,loc="upper left",frameon=False);grid(ax[3],"both")
finish(fig,1,"Study design and source-tissue structure",dict(zip("ABCD",["Study structure and distinct external contrasts","Arrays versus full-label groups by bed/status","Source sex distribution at label-group level","Profile similarity audit"])),"A, Study structure; the full-source frozen model is applied without target fitting. B, 104 deposited arrays are conservatively aggregated into 95 full-label groups (65 disease, 30 control), not authenticated independent patients. C, Male proportions illustrate a covariate available to the bed-and-sex control. D, Rank-profile similarity of the nine repeated-label pairs versus other pairs within the same bed and status. Pairwise comparisons are dependent and descriptive; high correlation does not authenticate donor identity. Infra., infrapopliteal.",["primary_analysis_units.csv","ambiguous_label_profile_similarity.csv","unmatched_within_stratum_similarity.csv"])

# FIGURE 2: finite-sample estimand decomposition.
fig,ax=layout("Discrimination and anatomical comparison populations")
panel(ax[0],"A","Pooled and within-bed discrimination")
rep=folds.groupby(["model","repeat"])[["auc","within_bed_auc"]].mean()
mean_range(ax[0],rep,MODELS,"auc",BLUE,-.12,"Pooled")
mean_range(ax[0],rep,MODELS,"within_bed_auc",ORANGE,.12,"Within-bed mean")
ax[0].set_yticks(range(7),[PNAMES[m] for m in MODELS]);ax[0].invert_yaxis();ax[0].set(xlim=(.32,1.025),xlabel="AUROC; mean and 10-repeat range");ax[0].legend(frameon=False,loc="lower left");grid(ax[0])
pc=cells[cells.model.eq("ridge200")].copy()
amat=pc.groupby(["case_bed","control_bed"]).auc.mean().unstack().reindex(index=BEDS,columns=BEDS).to_numpy()
panel(ax[1],"B","Ridge-200 case–control cells")
heat(ax[1],amat,BNAMES,BNAMES,vmin=.5,vmax=1,xlabel="Control arterial bed",ylabel="Disease arterial bed")
pc["weight"]=pc.pairs/pc.groupby(["repeat","fold"]).pairs.transform("sum")
wmat=pc.groupby(["case_bed","control_bed"]).weight.mean().unstack().reindex(index=BEDS,columns=BEDS).to_numpy()
panel(ax[2],"C","Comparison-pair weights")
heat(ax[2],100*wmat,BNAMES,BNAMES,fmt=".1f",vmin=0,vmax=20,xlabel="Control arterial bed",ylabel="Disease arterial bed")
ax[2].text(.5,-.24,"Mean weights (%); sum = 100% before rounding",ha="center",transform=ax[2].transAxes,fontsize=8)
panel(ax[3],"D","Reweighted anatomical mixtures")
mixgrid=np.array([[i/10,j/10,(10-i-j)/10] for i in range(11) for j in range(11-i)])
mix=mixgrid@amat@mixgrid.T
observed=folds.loc[folds.model.eq("ridge200"),"auc"].mean()
ax[3].hist(mix.ravel(),bins=18,color="#9BBBD6",edgecolor="white")
ax[3].axvline(observed,color=BLUE,lw=1.5,label=f"Observed pooled mean: {observed:.3f}")
ax[3].set(xlabel="Mean fold AUROC under specified mixture",ylabel="Grid scenarios (66 × 66)");ax[3].legend(frameon=False,loc="upper left");grid(ax[3],"y")
finish(fig,2,"Discrimination and anatomical comparison populations",dict(zip("ABCD",["Seven fixed models: pooled and diagonal-mean AUROC","Mean nine-cell primary AUROC matrix","Mean nine-cell pair weights","Mixture grid using mean cell AUROCs"])),"A, Fold metrics are averaged within each repetition, then summarized across ten repetitions; bars are the observed split range, not confidence intervals. B, Each cell compares case and control scores generated by the same held-out-fold model; cells are averaged over 30 folds. C, Mean fractions of all case–control pairs; within each fold the pair-weighted sum of cell AUROCs exactly equals pooled AUROC. D, The two 0.1-step three-bed simplexes each contain 66 mixtures, giving 4,356 deterministic reweighting scenarios. This histogram is the distribution of scenario values from the mean cell matrix, not a sampling distribution or observed population. It differs from the protocol's mean per-fold extrema, reported numerically in results.",["primary_fold_metrics.csv","primary_pair_cells.csv"])

# FIGURE 3: genuinely separate anatomical target fits.
fig,ax=layout("Transport to held-out arterial beds")
lobo=transport[(transport.source=="other_two") & ~transport.purge_possible_controls]
panel(ax[0],"A","Leave-one-bed-out AUROC")
for j,m in enumerate(EXTMODELS):
    d=lobo[lobo.model==m].set_index("target").reindex(BEDS)
    ax[0].plot(range(3),d.auc,marker=["^","s","o","D","x"][j],ms=[4,4,6,4,4][j],mfc="white" if m=="ridge200" else COLORS[m],color=COLORS[m],label=PNAMES[m],lw=1.1)
ax[0].set_xticks(range(3),BNAMES);ax[0].set(xlabel="Held-out arterial bed",ylabel="AUROC",ylim=(.45,1.03));ax[0].legend(frameon=False,fontsize=8,loc="lower left");grid(ax[0],"y")
panel(ax[1],"B","Leave-one-bed-out threshold transport")
for m in ["Wu6","ridge200","forest200"]:
    d=lobo[lobo.model==m].set_index("target").reindex(BEDS)
    ax[1].plot(range(3),d.balanced_accuracy,marker="o",ms=3.5,color=COLORS[m],label=PNAMES[m])
ax[1].axhline(.5,ls="--",color=GRAY,lw=.8);ax[1].set_xticks(range(3),BNAMES);ax[1].set(xlabel="Held-out arterial bed",ylabel="Balanced accuracy at 0.5",ylim=(.45,1.03));ax[1].legend(frameon=False,loc="lower left",bbox_to_anchor=(0,.10));grid(ax[1],"y")
panel(ax[2],"C","Single-bed source → target")
direct=transport[(transport.source!="other_two") & ~transport.purge_possible_controls]
arr=np.full((3,3),np.nan)
for _,r in direct.iterrows():arr[BEDS.index(r.source),BEDS.index(r.target)]=r.auc
im=ax[2].imshow(arr,vmin=.5,vmax=1,cmap="Blues",aspect="auto")
for i,s in enumerate(BEDS):
    for j,t in enumerate(BEDS):
        if s==t:ax[2].text(j,i,"—",ha="center",va="center",color=GRAY)
        else:
            r=direct[(direct.source==s)&(direct.target==t)].iloc[0]
            ax[2].text(j,i,f"{r.auc:.3f}\n({r.balanced_accuracy:.3f})",ha="center",va="center",fontsize=8,color="white" if r.auc>.84 else "#142534")
ax[2].set_xticks(range(3),BNAMES);ax[2].set_yticks(range(3),BNAMES);ax[2].set(xlabel="Target bed: AUROC (balanced accuracy)",ylabel="Source arterial bed")
panel(ax[3],"D","Possible-shared-control sensitivity")
r=transport[transport.model=="ridge200"]
wide=r.pivot(index=["source","target"],columns="purge_possible_controls",values=["auc","balanced_accuracy","n_removed"])
labels=[]
for s,t in wide.index:
    ss="".join(BSHORT[b] for b in BEDS if b!=t) if s=="other_two" else BSHORT[s]
    labels.append(f"{ss}→{BSHORT[t]} ({int(wide.loc[(s,t),('n_removed',True)])})")
y=np.arange(len(wide))
for metric,col,off,lab in [("auc",BLUE,-.13,"Δ AUROC"),("balanced_accuracy",ORANGE,.13,"Δ balanced accuracy")]:
    diff=wide[(metric,True)]-wide[(metric,False)]
    ax[3].scatter(diff,y+off,c=col,s=18,label=lab,zorder=3)
ax[3].axvline(0,color=GRAY,lw=.8);ax[3].set_yticks(y,labels);ax[3].set_ylim(len(wide)+2.2,-.7);ax[3].set(xlabel="Purged minus original metric",ylabel="Source→target (controls removed)");ax[3].legend(frameon=False,fontsize=8,loc="lower left");grid(ax[3])
finish(fig,3,"Transport to held-out arterial beds",dict(zip("ABCD",["Four source-fitted expression models and the fixed C1Q rule","Probability-model threshold transport","Six directional primary-model transports","Proxy-control purging: metric differences and exclusions"])),"A–B, For each trainable expression specification, a separate model is fitted on two source beds and evaluated in the remaining bed; the fixed C1Q score is applied without fitting. C, Ridge-200 is trained in a single bed for each of six directed source–target comparisons; cells show AUROC and balanced accuracy at the fixed 0.5 threshold. D, Changes after excluding source controls with a numeric suffix/sex proxy matching target controls; labels give the number removed. This deliberately conservative sensitivity does not authenticate shared donors. C, carotid; F, femoral; I, infrapopliteal. C1Q and centroid scores are not probabilities and are excluded from fixed-probability-threshold diagnostics. No uncertainty bars are inferred from these three or six dependent tasks.",["transport_metrics.csv"])

# FIGURE 4: paired external plaque/intact target.
fig,ax=layout("Paired external tissue discrimination and threshold failure",n=5,tall=True)
pp=pairs[pairs.model=="ridge200"].sort_values("patient")
panel(ax[0],"A","Frozen ridge scores in 32 pairs")
for _,r in pp.iterrows():
    color=BLUE if r.difference>0 else RED
    ax[0].plot([0,1],[r.intact_score,r.plaque_score],marker="o",ms=2.5,color=color,alpha=.55,lw=.8)
ax[0].axhline(.5,color=GRAY,ls="--",lw=.9,label="Source threshold 0.5")
ax[0].set_yscale("log");ax[0].yaxis.set_major_formatter(FuncFormatter(lambda x, pos: f"{x:.0e}".replace("e-0", "e-").replace("e+0", "e+")));ax[0].yaxis.set_minor_formatter(NullFormatter());ax[0].set_xticks([0,1],["Intact tissue","Plaque"]);ax[0].set(xlim=(-.25,1.25),ylabel="Ridge probability score (log scale)",ylim=(min(pp.intact_score.min(),pp.plaque_score.min())*.6,.85));ax[0].legend(frameon=True,facecolor="white",framealpha=1,edgecolor="none",fontsize=8,loc="upper left");grid(ax[0],"y")
panel(ax[1],"B","Plaque versus intact ROC")
roc(ax[1],"GSE43292")
panel(ax[2],"C","Within-patient score differences")
d=pp.sort_values("difference");xx=np.arange(32)
ax[2].vlines(xx,0,1000*d.difference,color=np.where(d.difference>0,BLUE,RED),lw=1.2)
ax[2].scatter(xx,1000*d.difference,c=np.where(d.difference>0,BLUE,RED),s=10,zorder=3)
ax[2].axhline(0,color=GRAY,lw=.8);ax[2].set(xlabel="Patients ordered by paired difference",ylabel="(Plaque − intact) score × 1,000");ax[2].set_xticks([0,15,31],["1","16","32"]);grid(ax[2],"y")
panel(ax[3],"D","Ranking and paired concordance")
ee=em[em.dataset=="GSE43292"].set_index("model").reindex(EXTMODELS);yy=np.arange(5)
for met,lo,hi,col,off,lab in [("auc","auc_low","auc_high",GRAY,-.13,"AUROC, bootstrap CI"),("pair_concordance","pair_exact_low","pair_exact_high",BLUE,.13,"Pair concordance, exact CI")]:
    ax[3].errorbar(ee[met],yy+off,xerr=np.array([ee[met]-ee[lo],ee[hi]-ee[met]]),fmt="o",ms=3.5,color=col,capsize=2,lw=.9,label=lab)
ax[3].set_yticks(yy,[PNAMES[m] for m in EXTMODELS]);ax[3].set_ylim(6,-.5);ax[3].set(xlim=(.5,1.02),xlabel="Metric with conditional 95% interval");ax[3].legend(frameon=False,fontsize=8,loc="lower left");grid(ax[3])
panel(ax[4],"E","Probability-model diagnostics at the unchanged 0.5 threshold")
threshold_table(ax[4],"GSE43292")
finish(fig,4,"Paired external tissue discrimination and threshold failure",dict(zip("ABCDE",["All 32 paired primary-model scores on original probability/log axis","Primary-model external ROC","All paired raw-score differences","Model AUROC and paired concordance with conditional intervals","Threshold and Brier diagnostics for probability models"])),"Frozen source models are applied to GSE43292: 64 carotid tissues from 32 paired hypertensive endarterectomy patients, plaque versus macroscopically intact Stary I/II tissue. A, Lines join tissues from the same patient; red indicates a lower plaque score. The log axis displays the original positive scores and the unchanged threshold. B, The primary-model AUROC interval uses 2,000 patient-block bootstrap draws. C, Raw paired differences, scaled only by 1,000 for legibility. D, AUROC intervals use the patient bootstrap; pair-concordance intervals are exact binomial intervals because there are no ties. These are conditional on the frozen model. E, All primary-model scores fall below 0.5 despite ranking discrimination. Brier scores describe selected tissue samples and are not clinical-risk calibration. Nonprobability scores have no threshold/Brier entries.",["external_predictions.csv","external_pairs.csv","external_metrics.csv"])

# FIGURE 5: distinct histological-severity target; not a disease/control validation.
fig,ax=layout("External histology contrast: advanced versus early lesions")
panel(ax[0],"A","Primary-model histology ROC")
roc(ax[0],"GSE28829")
panel(ax[1],"B","Probability scale after transport")
dd=ep[(ep.dataset=="GSE28829")&(ep.model=="ridge200")].copy()
for y,col in [(0,GRAY),(1,BLUE)]:
    vals=np.sort(dd.loc[dd.y==y,"score"].to_numpy());jit=np.linspace(-.16,.16,len(vals))
    ax[1].scatter(y+jit,vals,s=22,color=col,edgecolor="white",lw=.4,zorder=3)
    ax[1].hlines(np.median(vals),y-.2,y+.2,color=col,lw=1.5)
ax[1].axhline(.5,color=RED,ls="--",lw=.9)
ax[1].annotate("Threshold 0.5", xy=(.04,.5), xycoords=("axes fraction","data"), xytext=(0,5), textcoords="offset points", va="bottom", color=RED, fontsize=8, bbox=dict(facecolor="white",edgecolor="none",pad=.6))
ax[1].set_yscale("log");ax[1].yaxis.set_major_formatter(FuncFormatter(lambda x, pos: f"{x:.0e}".replace("e-0", "e-").replace("e+0", "e+")));ax[1].yaxis.set_minor_formatter(NullFormatter())
ax[1].set_xticks([0,1],[f"Early (n={(dd.y==0).sum()})",f"Advanced (n={(dd.y==1).sum()})"]);ax[1].set(ylabel="Ridge probability score (log scale)",xlim=(-.4,1.4),ylim=(dd.score.min()*.6,.95));grid(ax[1],"y")
panel(ax[2],"C","Frozen-model ranking comparison")
ee=em[em.dataset=="GSE28829"].set_index("model").reindex(EXTMODELS)
for i,m in enumerate(EXTMODELS):
    row=ee.loc[m];ax[2].errorbar(row.auc,i,xerr=[[row.auc-row.auc_low],[row.auc_high-row.auc]],fmt="o",ms=4,capsize=2,lw=.9,color=COLORS[m])
ax[2].set_yticks(range(5),[PNAMES[m] for m in EXTMODELS]);ax[2].invert_yaxis();ax[2].set(xlim=(.5,1.025),xlabel="AUROC; conditional 95% bootstrap CI");grid(ax[2])
panel(ax[3],"D","Unchanged threshold diagnostics")
threshold_table(ax[3],"GSE28829")
finish(fig,5,"External histology contrast: advanced versus early lesions",dict(zip("ABCD",["Primary-model severity ROC","All primary probability scores by histology","Five fixed-model AUROCs with intervals","Probability-model threshold diagnostics"])),"GSE28829 contains 29 autopsy carotid specimens (13 early, 16 advanced lesions); deposited metadata do not establish an independent donor count. This is a changed histological contrast, not healthy-control discrimination. A, Primary frozen-model ROC. B, Original probability scores on a log axis; horizontal marks are within-class medians. C, Conditional percentile intervals from 2,000 class-stratified specimen bootstraps; unresolved donor dependence is not captured. A perfect observed AUROC can have a degenerate fixed-score empirical interval, which does not establish zero uncertainty. D, Threshold diagnostics without target recalibration; Brier values do not estimate clinical-risk calibration. No best external model was selected for the primary conclusion.",["external_predictions.csv","external_metrics.csv"])

# FIGURE 6: prespecified sensitivity, pipeline nulls, subset stability.
fig,ax=layout("Specification, negative-control and subset stability analyses",n=5,tall=True)
primary=folds[folds.model.eq("ridge200")].copy();primary["scenario"]="primary"
ss=pd.concat([primary,sens],ignore_index=True)
order=["primary","top50","top500","C001","C1","native","exclude_ambiguous","104_grouped_arrays","possible_shared_controls"]
snames={"primary":"Primary (95 groups)","top50":"Top 50 genes","top500":"Top 500 genes","C001":"Ridge C = 0.01","C1":"Ridge C = 1","native":"Native expression","exclude_ambiguous":"Exclude ambiguity (86)","104_grouped_arrays":"All 104; grouped folds","possible_shared_controls":"Control-proxy groups"}
rr=ss.groupby(["scenario","repeat"])[["auc","delta_mix"]].mean()
panel(ax[0],"A","Specification sensitivity: AUROC")
mean_range(ax[0],rr,order,"auc",BLUE)
ax[0].set_yticks(range(9),[snames[s] for s in order]);ax[0].invert_yaxis();ax[0].set(xlim=(min(.95,rr.auc.min()-.005),1.003),xlabel="Repeat mean and range");grid(ax[0])
panel(ax[1],"B","Specification sensitivity: Δmix")
mean_range(ax[1],rr,order,"delta_mix",ORANGE)
ax[1].axvline(0,color=GRAY,lw=.8);ax[1].set_yticks(range(9),[""]*9);ax[1].invert_yaxis();ax[1].set(xlabel="Pooled − within-bed mean AUROC");grid(ax[1])
ax[1].xaxis.set_major_locator(MaxNLocator(3))
obs=primary[primary.repeat==0].auc.mean()
for aa,scheme,letter,title in [(ax[2],"within_bed","C","Within-bed permutation null"),(ax[3],"unrestricted","D","Unrestricted permutation null")]:
    vals=null[null.scheme==scheme].mean_fold_auc
    panel(aa,letter,title);aa.hist(vals,bins=np.linspace(.25,.8,15),color="#AAC2D8",edgecolor="white")
    aa.set_ylim(0, aa.get_ylim()[1]*1.22)  # Reserve clear headroom for the opaque legend.
    aa.axvline(.5,color=GRAY,ls="--",lw=.8);aa.axvline(obs,color=BLUE,lw=1.5,label=f"Observed {obs:.3f}")
    aa.set(xlim=(min(.25,vals.min()-.02),1.02),xlabel="Mean held-out-fold AUROC",ylabel="Permutations (n=99)");aa.legend(frameon=True,facecolor="white",framealpha=1,edgecolor="none",loc="upper left");grid(aa,"y")
panel(ax[4],"E","Complete-pipeline stability in 30 stratified subsets")
idx=np.arange(len(sub))
ax[4].plot(idx,sub.auc,"o-",color=BLUE,ms=3,label="Pooled AUROC",lw=.9)
ax[4].plot(idx,sub.within_bed_auc,"s-",color=ORANGE,ms=3,label="Mean within-bed AUROC",lw=.9)
ax[4].fill_between(idx,sub.mix_min,sub.mix_max,color="#DCE8F1",alpha=.8,label="Per-subset mixture extrema",zorder=0)
ax[4].set(xlabel="Stratified subset / pipeline rerun",ylabel="Mean fold AUROC",ylim=(max(.85,sub.mix_min.min()-.01),1.004));ax[4].set_xticks([0,9,19,29],["1","10","20","30"]);ax[4].legend(frameon=False,loc="lower left",ncol=2);grid(ax[4],"y")
finish(fig,6,"Specification, negative-control and subset stability analyses",dict(zip("ABCDE",["Eight alternative specifications plus primary AUROC","The same specifications' primary composition contrast","Within-bed label-permutation pipeline null","Unrestricted label-permutation pipeline null","Thirty full-pipeline 80% stratum subsamples"])),"A–B, Each point is the mean of ten repetition means, each formed from three held-out folds; bars show the ten-repeat range, not confidence intervals. Each alternative was specified before its dependent analysis. Identity conventions include excluding all repeated-label records (86 arrays), retaining 104 arrays with full-label groups kept in one fold, and conservatively grouping possible shared-control suffix/sex proxies. C–D, Ninety-nine label permutations per scheme rerun feature selection, scaling and fitting; the observed line is the first fixed three-fold scheme. These are pipeline-null diagnostics, not tests of anatomical causation or independent replications. E, Thirty stratified 80% subsets (flooring within six strata gives 75 label groups) rerun the complete primary pipeline. Shading shows mean per-fold mixture extrema within each subset, not a confidence band.",["primary_fold_metrics.csv","sensitivity_all_fold_metrics.csv","permutation_nulls.csv","full_pipeline_subsampling.csv"])

captions["Figure_2"]["caption"] += " C1Q and Wu6 are retrospective literature controls with prior cohort exposure, not independent biomarker discovery validation."
(OUT/"figure_captions.json").write_text(json.dumps(captions,indent=2,ensure_ascii=False),encoding="utf-8")
(OUT/"render_bounds_checks.json").write_text(json.dumps(render_checks,indent=2,ensure_ascii=False),encoding="utf-8")
manifest={"script":"code/plot_figures.py","input_sha256":inputs,"figures":{},"main_figures":6,"main_panels":sum(v["panel_count"] for v in captions.values()),"font":"DejaVu Sans, minimum 8 pt at 17 cm width","display_transformations":["Original ridge probability on logarithmic axes, no recalibration","Paired raw differences multiplied by 1000 for display","Split ranges are not confidence intervals","Mixture-grid figure algebraically aggregates saved nine-cell AUROCs"]}
for p in sorted(OUT / f"Figure_{i}.{ext}" for i in range(1,7) for ext in ["pdf","svg","png"]):
    manifest["figures"][p.name]={"bytes":p.stat().st_size,"sha256":hashlib.sha256(p.read_bytes()).hexdigest()}
(OUT/"figure_provenance.json").write_text(json.dumps(manifest,indent=2),encoding="utf-8")

# Contact sheet is a visual QA overview, not a publication result panel.
thumbs=[]
for i in range(1,7):
    im=Image.open(OUT/f"Figure_{i}.png").convert("RGB")
    im.thumbnail((850,1150))
    tile=Image.new("RGB",(900,1200),"#E5E9ED")
    tile.paste(im,((900-im.width)//2,35))
    ImageDraw.Draw(tile).text((20,10),f"Figure {i}",fill="black")
    thumbs.append(tile)
sheet=Image.new("RGB",(2700,2400),"white")
for i,t in enumerate(thumbs):sheet.paste(t,((i%3)*900,(i//3)*1200))
sheet.save(OUT/"contact_sheet.png")
print(json.dumps({"figures":6,"panels":manifest["main_panels"],"output_directory":str(OUT),"formats":["PDF","SVG","300dpi PNG"],"input_tables":len(inputs)},indent=2))
```
