## Supplementary material for "Anatomical comparison populations and threshold transport in vascular transcriptomic classification": Reproducibility archive: code, provenance and results: plot_supplement_extension.py

```
"""Additional displays of original saved scores; no new fitting or inferential claim."""
from pathlib import Path
import hashlib,json,textwrap
import numpy as np
import pandas as pd
import matplotlib
matplotlib.use('Agg')
import matplotlib.pyplot as plt

ROOT=Path(__file__).resolve().parents[1];OUT=ROOT/'figures'
W=17/2.54;BLUE='#1765A3';ORANGE='#D38B32';GRAY='#77818C';RED='#B54747'
plt.rcParams.update({'font.family':'DejaVu Sans','font.size':8,'axes.titlesize':9,'axes.labelsize':8,
    'xtick.labelsize':8,'ytick.labelsize':8,'legend.fontsize':8,'axes.linewidth':.6,
    'pdf.fonttype':42,'svg.fonttype':'none','axes.spines.top':False,'axes.spines.right':False})
inputs={};caps={};bounds=[]
def read(name):
    p=ROOT/'results'/name;inputs[str(p.relative_to(ROOT)).replace('\\','/')]=hashlib.sha256(p.read_bytes()).hexdigest()
    return pd.read_csv(p)
def panel(a,l,t):a.set_title(l+'  '+t,loc='left',fontweight='bold',pad=9)
def finish(fig,n,title,panels,caption):
    fig.canvas.draw();renderer=fig.canvas.get_renderer();fb=fig.bbox;outside=[]
    for t in fig.findobj(matplotlib.text.Text):
        if not t.get_visible() or not t.get_text():continue
        bb=t.get_window_extent(renderer)
        # Inspect actual visible objects; unused off-range auto ticks are ignored below.
        if bb.x0<fb.x0-1 or bb.x1>fb.x1+1 or bb.y0<fb.y0-1 or bb.y1>fb.y1+1:outside.append(t.get_text())
    for ext in ['pdf','svg','png']:fig.savefig(OUT/f'Figure_S{n}.{ext}',dpi=300,facecolor='white')
    caps[f'Figure_S{n}']={'title':title,'panel_count':len(panels),'panels':panels,'caption':caption,
        'status':'Additional display of original saved results, assembled after the primary analyses',
        'width_cm':17,'formats':['PDF','SVG','PNG_300dpi']}
    bounds.append({'figure':f'Figure_S{n}','outside_candidate_text':outside})
    plt.close(fig)

cells=read('primary_pair_cells.csv');cells=cells[cells.model=='ridge200']
beds=['Carotid','Femoral','Infrapopliteal'];names=['Carotid','Femoral','Infrapopliteal']
fig,axes=plt.subplots(3,3,figsize=(W,8.1),layout='constrained')
fig.suptitle('Fold-level resolution of anatomical comparisons',fontsize=11,fontweight='bold',x=.02,ha='left')
paneldefs={}
for ia,a in enumerate(beds):
    for ib,b in enumerate(beds):
        k=ia*3+ib;ax=axes[ia,ib];d=cells[(cells.case_bed==a)&(cells.control_bed==b)]
        letter=chr(65+k);paneldefs[letter]=f'{a} cases versus {b} controls'
        panel(ax,letter,f'{names[ia]} cases\n{names[ib]} controls')
        ax.scatter(d.repeat+1+(d.fold-1)*.16,d.auc,s=15,color=BLUE if ia==ib else ORANGE,alpha=.75,lw=0)
        ax.set(ylim=(.85,1.018),xlim=(.4,10.6),xticks=[1,5,10],yticks=[.85,.9,.95,1])
        ax.text(.04,.08,f'{int(d.pairs.min())}–{int(d.pairs.max())} pairs/fold',transform=ax.transAxes,fontsize=8)
        ax.grid(axis='y',color='#E6E9EC',lw=.5);ax.set_axisbelow(True)
        if ia==2:ax.set_xlabel('Source repetition')
        if ib==0:ax.set_ylabel('Within-fold cell AUROC')
finish(fig,1,'Fold-level resolution of anatomical comparisons',paneldefs,
    'A–I, All 30 held-out folds for each primary-model case-bed/control-bed comparison. Horizontal jitter separates the three folds of each repetition. Blue denotes diagonal within-bed comparisons; orange denotes cross-bed comparisons. The AUROC axis is deliberately truncated at 0.85 to display the smallest observed cell (0.875). Labels give the actual pair-count range in each cell; pairs share observations, and folds/repetitions share source data. Points are not independent studies, and no confidence interval is implied. These original saved results complement the mean matrix in Figure 2; no new model was fitted for this display.')

p=read('primary_predictions.csv');p=p[p.model=='ridge200']
units=read('primary_analysis_units.csv').set_index('label_group')
rows=[]
for sample,d in p.groupby('sample'):
    assert len(d)==10 and d.repeat.nunique()==10
    row=units.loc[sample]
    rows.append(dict(sample=sample,bed=row.bed,y=int(row.y),mean=d.score.mean(),minimum=d.score.min(),
        maximum=d.score.max(),wrong_at_half=int(((d.score>=.5).astype(int)!=d.y).sum())))
a=pd.DataFrame(rows);a['bed_order']=a.bed.map(dict(zip(beds,range(3))))
a=a.sort_values(['bed_order','y','mean','sample']).reset_index(drop=True);a['display_position']=np.arange(1,len(a)+1)
(ROOT/'results/display_source_score_stability.csv').parent.mkdir(exist_ok=True)
# Store display products separately so original core result inventory remains untouched.
display=ROOT/'results/exploratory_display';display.mkdir(exist_ok=True)
a.to_csv(display/'source_score_stability.csv',index=False)
fig=plt.figure(figsize=(W,7.1),layout='constrained');gs=fig.add_gridspec(2,1,height_ratios=[1.45,1])
ax=fig.add_subplot(gs[0]);bx=fig.add_subplot(gs[1])
fig.suptitle('Repeated source scores expose threshold discrepancies',fontsize=11,fontweight='bold',x=.02,ha='left')
panel(ax,'A','All 95 label groups: ten held-out scores per group')
centres=[];labels=[]
for j,((bed,y),g) in enumerate(a.groupby(['bed_order','y'],sort=True)):
    left=g.display_position.min()-.5;right=g.display_position.max()+.5
    ax.axvspan(left,right,color='#F0F3F6' if j%2==0 else '#FFFFFF',zorder=0)
    c=BLUE if y==1 else GRAY
    ax.vlines(g.display_position,g.minimum,g.maximum,color=c,lw=.8,alpha=.8)
    ax.scatter(g.display_position,g['mean'],s=9,color=c,zorder=3)
    centres.append((left+right)/2);labels.append(['Carotid','Femoral','Infra.'][bed]+'\n'+('Disease' if y else 'Control'))
    ax.axvline(right,color='#DBE2E9',lw=.6,zorder=0)
bad=a[a.wrong_at_half==10]
ax.scatter(bad.display_position,bad['mean'],s=35,facecolors='none',edgecolors=RED,lw=1.2,zorder=4,label='Wrong in all 10 repeats')
ax.axhline(.5,color=RED,ls='--',lw=1,label='Technical threshold 0.5')
ax.set(xlim=(0,96),ylim=(-.03,1.04),ylabel='Held-out probability score',xticks=centres,xticklabels=labels)
ax.legend(loc='center right',framealpha=.95,fontsize=8)
panel(bx,'B','Error frequency counts groups, not repeated experiments')
cnt=a.wrong_at_half.value_counts().reindex(range(11),fill_value=0)
bx.bar(cnt.index,cnt.values,color=[GRAY if i==0 else (RED if i==10 else ORANGE) for i in cnt.index],width=.7)
for i,n in cnt.items():
    if n:bx.text(i,n+.7,str(n),ha='center',fontsize=8)
bx.set(xlabel='Number of incorrect classifications in ten repetitions',ylabel='Complete-label groups',xticks=range(11),ylim=(0,cnt.max()*1.14))
bx.grid(axis='y',color='#E6E9EC',lw=.5);bx.set_axisbelow(True)
finish(fig,2,'Repeated source scores expose threshold discrepancies',{'A':'All 95 group score ranges','B':'Frequency of 0.5 threshold errors'},
    'A, Each point is one group’s mean of ten original held-out primary scores, with its minimum-to-maximum range. Groups are ordered within bed/status by mean score; this ordering is descriptive. Red outlines identify groups misclassified in every repetition at 0.5. The display-position-to-label mapping is supplied in results/exploratory_display/source_score_stability.csv. B, Counts of groups by their number of threshold errors. Source groups are unverified donor units, repeated fits are dependent, and ranges are not confidence intervals. These pooled score summaries do not replace the original within-fold AUROC estimand. No new fitting or threshold selection was performed for this display.')
(OUT/'supplement_extension_captions.json').write_text(json.dumps(caps,indent=2,ensure_ascii=False),encoding='utf8')
(OUT/'supplement_extension_provenance.json').write_text(json.dumps({'input_sha256':inputs,'output_sha256':{p.name:hashlib.sha256(p.read_bytes()).hexdigest() for i in [1,2] for p in [OUT/f'Figure_S{i}.{e}' for e in ['pdf','svg','png']]},'bounds_candidates':bounds},indent=2),encoding='utf8')
print(json.dumps({'supplementary_figures':2,'panels':11,'persistent_wrong':bad['sample'].tolist(),'error_frequency':cnt.to_dict()},indent=2))
```
