## Supplementary material for "Anatomical comparison populations and threshold transport in vascular transcriptomic classification": Reproducibility archive: code, provenance and results: plot_threshold_extension.py

```
"""Exploratory Figures9-11 from saved threshold/score/failure outputs only."""
from pathlib import Path
import hashlib, json, textwrap
import numpy as np
import pandas as pd
import matplotlib
matplotlib.use('Agg')
import matplotlib.pyplot as plt
from matplotlib.colors import LinearSegmentedColormap, ListedColormap, TwoSlopeNorm
from matplotlib.lines import Line2D
from matplotlib.patches import Patch
from matplotlib.ticker import PercentFormatter, NullFormatter
from PIL import Image, ImageDraw
import fitz

ROOT=Path(__file__).resolve().parents[1]
RES=ROOT/'results/exploratory_threshold';OUT=ROOT/'figures'
W=17/2.54
BLUE,ORANGE,GRAY,RED,PURPLE,TEAL='#1765A3','#D38B32','#77818C','#B54747','#7454A3','#48A9A6'
MODEL_ORDER=['C1Q','Wu6','ridge200','centroid200','forest200']
MODEL_NAMES={'C1Q':'C1Q','Wu6':'Wu6','ridge200':'Ridge-200','centroid200':'Centroid','forest200':'Forest'}
RULE_ORDER=['fixed_0.5','source_max_youden','source_sensitivity_0.90']
RULE_NAMES={'fixed_0.5':'Fixed 0.5','source_max_youden':'Source Youden','source_sensitivity_0.90':'Source sens. >=0.90'}
RULE_COLORS=dict(zip(RULE_ORDER,[GRAY,PURPLE,TEAL]))
plt.rcParams.update({'font.family':'DejaVu Sans','font.size':8,'axes.titlesize':9,
    'axes.labelsize':8,'xtick.labelsize':8,'ytick.labelsize':8,'legend.fontsize':8,
    'figure.titlesize':11,'axes.linewidth':.6,'lines.linewidth':1.2,
    'savefig.dpi':300,'pdf.fonttype':42,'ps.fonttype':42,'svg.fonttype':'none',
    'axes.spines.top':False,'axes.spines.right':False,'axes.titlelocation':'left'})
inputs={};captions={};checks=[]

def read(name):
    p=RES/(name+'.csv');inputs[str(p.relative_to(ROOT)).replace('\\','/')]=hashlib.sha256(p.read_bytes()).hexdigest()
    return pd.read_csv(p)

def panel(ax,letter,title,width=24):
    ax.set_title(letter+'  '+textwrap.fill(title,width=width,subsequent_indent='   '),fontweight='bold',pad=9)
    ax.tick_params(width=.6,length=3)

def grid(ax,axis='x'):
    ax.grid(axis=axis,color='#E6E9EC',linewidth=.55,zorder=0);ax.set_axisbelow(True)

def title(fig,text):
    fig.suptitle(textwrap.fill(text,width=68),x=.015,ha='left',fontweight='bold')

def finish(fig,number,name,panels,caption,sources):
    fig.canvas.draw(); renderer=fig.canvas.get_renderer();bounds=fig.bbox;bad=[]
    artists=list(fig.texts)
    for ax in fig.axes:
        artists+=list(ax.texts)+[ax._left_title,ax.title,ax.xaxis.label,ax.yaxis.label]
        if ax.axison:
            lo,hi=sorted(ax.get_xlim());yl,yh=sorted(ax.get_ylim())
            artists += [t for t in ax.get_xticklabels() if lo-1e-10<=t.get_position()[0]<=hi+1e-10]
            artists += [t for t in ax.get_yticklabels() if yl-1e-10<=t.get_position()[1]<=yh+1e-10]
        if ax.get_legend():artists+=ax.get_legend().get_texts()
    for item in artists:
        if not item.get_visible() or not item.get_text():continue
        b=item.get_window_extent(renderer)
        if b.x0<bounds.x0-1 or b.y0<bounds.y0-1 or b.x1>bounds.x1+1 or b.y1>bounds.y1+1:
            bad.append({'text':item.get_text(),'bbox':[float(v) for v in b.bounds]})
    for ext in ['pdf','svg','png']:fig.savefig(OUT/f'Figure_{number}.{ext}',dpi=300)
    plt.close(fig)
    with fitz.open(OUT/f'Figure_{number}.pdf') as doc:
        sizes=[];outside=[]
        for page in doc:
            for block in page.get_text('dict')['blocks']:
                for line in block.get('lines',[]):
                    for span in line['spans']:
                        sizes.append(span['size']);b=fitz.Rect(span['bbox'])
                        if not (page.rect+(-1,-1,1,1)).contains(b):outside.append(span['text'])
        qa={'figure':number,'pdf_pages':len(doc),'minimum_text_size_pt':min(sizes),
            'matplotlib_outside_canvas':bad,'pdf_outside_text':outside}
    checks.append(qa)
    assert not bad and not outside, qa
    assert qa['minimum_text_size_pt']>=7.95,qa
    captions[f'Figure_{number}']={'title':name,'panel_count':len(panels),'panels':panels,
        'caption':caption,'source_tables':['exploratory_threshold/'+s+'.csv' for s in sources],
        'analysis_status':'Post-result exploratory diagnostics, extension04','width_cm':17,
        'formats':['PDF','SVG','PNG_300dpi']}

geo=read('score_geometry_samples');dec=read('class_logit_decomposition')
mc=read('mean_feature_contributions');top=read('top20_source_coefficient_genes')
thresholds=read('threshold_rules');ev=read('evaluation_metrics');curves=read('threshold_response_curves')
signs=read('paired_five_rule_sign_map');pdels=read('paired_deletion_influence');hdels=read('histology_deletion_influence')
pdiffs=read('paired_feature_logit_differences');margins=read('paired_primary_margins')
GROUPS=[('GSE100927',0,'Source donor (30)'),('GSE100927',1,'Source disease (65)'),
        ('GSE43292',0,'Intact tissue (32)'),('GSE43292',1,'Plaque tissue (32)'),
        ('GSE28829',0,'Early lesion (13)'),('GSE28829',1,'Advanced lesion (16)')]
SHORT=['Src C','Src D','Pair I','Pair P','Hist E','Hist A']
diverge=LinearSegmentedColormap.from_list('signed_source_terms',[ORANGE,'#FFFFFF',BLUE])

# Figure9: individual score support, exact score accounting and representation shift.
name='Frozen score geometry under changed tissue comparisons'
fig=plt.figure(figsize=(W,7.8),layout='constrained');gs=fig.add_gridspec(2,2,hspace=.11,wspace=.09)
ax=[fig.add_subplot(gs[i//2,i%2]) for i in range(4)];title(fig,name)
panel(ax[0],'A','Class-specific frozen logits')
rng=np.random.default_rng(20261026)
for i,(acc,yy,label) in enumerate(GROUPS):
    v=geo.loc[(geo.dataset==acc)&(geo.y==yy),'linear_logit'].to_numpy()
    color=BLUE if yy else GRAY
    ax[0].scatter(v,np.full(len(v),i)+rng.uniform(-.19,.19,len(v)),s=8,color=color,alpha=.75,linewidths=0)
    ax[0].plot([np.median(v)]*2,[i-.28,i+.28],color='black',lw=1.3)
ax[0].axvline(0,color=RED,lw=.9,ls='--');ax[0].set_yticks(range(6),[v[2] for v in GROUPS]);ax[0].invert_yaxis()
ax[0].set(xlabel='Frozen linear logit\n(0 = probability 0.5)');grid(ax[0])
ax[0].text(.02,.01,'Source: in-sample reference',transform=ax[0].transAxes,fontsize=8,color=GRAY,
           bbox={'facecolor':'white','edgecolor':'none','pad':1})

panel(ax[1],'B','Exact class-mean score accounting')
for i,(acc,yy,_) in enumerate(GROUPS):
    row=dec[(dec.dataset==acc)&(dec.class_state.astype(str)==str(yy))].iloc[0]
    ax[1].barh(i,row.positive_mean_feature_sum,color=BLUE,height=.56)
    ax[1].barh(i,row.negative_mean_feature_sum,color=ORANGE,height=.56)
    ax[1].barh(i,row.intercept,left=row.positive_mean_feature_sum,color=GRAY,height=.56)
    ax[1].plot(row.mean_logit,i,'D',color='black',ms=3)
ax[1].axvline(0,color='#B6BEC6',lw=.7);ax[1].set_yticks(range(6),SHORT);ax[1].invert_yaxis()
ax[1].set(xlabel='Mean weighted feature terms\nplus intercept');grid(ax[1])
ax[1].legend(handles=[Patch(color=BLUE,label='Positive terms'),Patch(color=ORANGE,label='Negative terms'),
    Patch(color=GRAY,label='Intercept'),Line2D([0],[0],marker='D',color='black',ls='',ms=3,label='Mean logit')],
    loc='lower left',frameon=True,facecolor='white',framealpha=1,edgecolor='none',ncol=1,bbox_to_anchor=(.0,.005))
ax[1].set_ylim(9.2,-.6)

panel(ax[2],'C','Source-standardized displacement')
for i,(acc,yy,label) in enumerate(GROUPS):
    v=geo.loc[(geo.dataset==acc)&(geo.y==yy),'z_abs_gt3_fraction'].to_numpy()
    ax[2].scatter(v,np.full(len(v),i)+rng.uniform(-.18,.18,len(v)),s=9,color=BLUE if yy else GRAY,
                  alpha=.75,linewidths=0)
    ax[2].plot(np.mean(v),i,'D',color='black',ms=3)
ax[2].set_yticks(range(6),SHORT);ax[2].invert_yaxis();ax[2].xaxis.set_major_formatter(PercentFormatter(1))
ax[2].set(xlabel='Fraction of 200 selected genes\nwith |z| > 3 in each specimen');grid(ax[2])
ax[2].text(.02,.01,'Descriptive reference only',transform=ax[2].transAxes,fontsize=8,
           color=GRAY,bbox={'facecolor':'white','edgecolor':'none','pad':1})

panel(ax[3],'D','Source-selected feature contributions')
genes=top.sort_values('source_absolute_coefficient_rank').gene.tolist()
matrix=np.column_stack([mc[(mc.dataset==acc)&(mc.class_state.astype(str)==str(yy))].set_index('gene').loc[genes,
                         'mean_weighted_contribution'].to_numpy() for acc,yy,_ in GROUPS])
lim=float(np.max(abs(matrix)))
im=ax[3].imshow(matrix,aspect='auto',cmap=diverge,norm=TwoSlopeNorm(vmin=-lim,vcenter=0,vmax=lim))
ax[3].set_yticks(range(len(genes)),genes);ax[3].set_xticks(range(6),SHORT,rotation=40,ha='right')
cb=fig.colorbar(im,ax=ax[3],orientation='horizontal',pad=.03,shrink=.85);cb.set_label('Mean weighted contribution\n(logit units)')
finish(fig,9,name,dict(zip('ABCD',['Observed class score distributions','Exact class-mean linear accounting',
    'Source-standardized feature displacement','Source-selected named-feature display'])),
    'Post-result exploratory diagnostics. A, Each point is one specimen or source label group scored with the same original full-source ridge model; source values are in-sample descriptive references, not held-out performance. Black marks denote class medians. B, Positive and negative sums of the 200 class-mean coefficient-weighted standardized features, plus the intercept, exactly reconstruct the black mean-logit markers. Classes are shown separately because overall cohort means also depend on class composition. C, Each point gives the fraction of 200 selected feature values outside absolute source-standardized z = 3; diamonds show class means. This descriptive reference does not establish an out-of-distribution detector or patient error rate. D, Genes are displayed in descending absolute full-source coefficient order, fixed without target outcomes; the heatmap shows only these 20 genes, whereas B uses all 200. Src C/D, source donor control/surgical disease; Pair I/P, paired intact/plaque tissues; Hist E/A, early/advanced histology. These are distinct endpoints. The accounting does not identify causal genes or isolate platform, procurement, normalization or comparator effects. GSE28829 donor independence is unresolved.',
    ['score_geometry_samples','class_logit_decomposition','mean_feature_contributions','top20_source_coefficient_genes'])

# Figure10: source-only selection contrasted with unchanged target outcomes.
name='Tested source thresholds leave external sensitivity at zero'
fig=plt.figure(figsize=(W,8.1),layout='constrained');gs=fig.add_gridspec(3,2,height_ratios=[1,1,0.72],hspace=.12,wspace=.09)
ax=[fig.add_subplot(gs[i//2,i%2]) for i in range(4)];ax.append(fig.add_subplot(gs[2,:]));title(fig,name)
contexts=['all_source','LOBO_to_Carotid','LOBO_to_Femoral','LOBO_to_Infrapopliteal']
panel(ax[0],'A','Thresholds learned only in source data')
for rule,offset in zip(RULE_ORDER,[-.13,0,.13]):
    vals=[thresholds[(thresholds.learning_context==c)&(thresholds.rule==rule)].threshold.iloc[0] for c in contexts]
    ax[0].plot(np.arange(4)+offset,vals,'o',color=RULE_COLORS[rule],ms=4,label=RULE_NAMES[rule])
ax[0].set_xticks(range(4),['All source','To carotid','To femoral','To infra.'],rotation=30,ha='right')
ax[0].set(ylabel='Source-derived probability threshold',ylim=(0,1.03));grid(ax[0],'y')
ax[0].legend(loc='upper left',frameon=False)
panel(ax[1],'B','Nested anatomical operating performance')
datasets=['LOBO_Carotid','LOBO_Femoral','LOBO_Infrapopliteal']
for rule,offset in zip(RULE_ORDER,[-.13,0,.13]):
    vals=[ev[(ev.dataset==d)&(ev.rule==rule)].balanced_accuracy.iloc[0] for d in datasets]
    ax[1].plot(np.arange(3)+offset,vals,'o-',color=RULE_COLORS[rule],ms=4,label=RULE_NAMES[rule],lw=.8)
ax[1].set_xticks(range(3),['Carotid','Femoral','Infra.']);ax[1].set(ylabel='Outer-bed balanced accuracy',ylim=(.5,1.02));grid(ax[1],'y')
ax[1].text(.02,.02,'Threshold selection excludes the target bed',transform=ax[1].transAxes,fontsize=8,color=GRAY)

for a,letter,dataset,heading in [(ax[2],'C','GSE43292','Paired target threshold response'),
                                (ax[3],'D','GSE28829','Histology target threshold response')]:
    panel(a,letter,heading)
    cc=curves[(curves.dataset==dataset)&(curves.threshold>0)].sort_values('threshold')
    a.step(cc.threshold,cc.sensitivity,where='post',color=BLUE,label='Sensitivity')
    a.step(cc.threshold,cc.specificity,where='post',color=ORANGE,label='Specificity')
    for rule,ls in zip(RULE_ORDER,['-','--',':']):
        t=thresholds[(thresholds.learning_context=='all_source')&(thresholds.rule==rule)].threshold.iloc[0]
        a.axvline(t,color=RULE_COLORS[rule],ls=ls,lw=1)
    a.set(xscale='log',xlim=(1e-7,1.02),ylim=(-.03,1.05),xlabel='Probability threshold (descriptive sweep)',ylabel='Fraction')
    a.set_xticks([1e-7,1e-5,1e-3,.1,1],['1e-7','1e-5','1e-3','0.1','1']);a.xaxis.set_minor_formatter(NullFormatter())
    a.legend(loc='upper left',frameon=True,facecolor='white',framealpha=1,edgecolor='none',bbox_to_anchor=(0,.73));grid(a,'both')

panel(ax[4],'E','Frozen external confusion counts for all three source rules',width=78);ax[4].axis('off')
table_rows=[]
for dataset,short in [('GSE43292','Paired tissues'),('GSE28829','Histology')]:
    for rule in RULE_ORDER:
        row=ev[(ev.dataset==dataset)&(ev.rule==rule)].iloc[0]
        table_rows.append([short,RULE_NAMES[rule],f'{row.threshold:.3f}',f'{int(row.tp)}/{int(row.fn)}',
            f'{int(row.tn)}/{int(row.fp)}',f'{row.sensitivity:.2f}',f'{row.specificity:.2f}',f'{row.balanced_accuracy:.2f}'])
table=ax[4].table(cellText=table_rows,colLabels=['Target','Source rule','Cutoff','TP / FN','TN / FP','Sens.','Spec.','Bal. acc.'],
    colWidths=[.15,.25,.09,.11,.11,.09,.09,.11],loc='center',cellLoc='center',bbox=[0,.02,1,.94])
table.auto_set_font_size(False);table.set_fontsize(8)
for (r,c),cell in table.get_celld().items():
    cell.set_edgecolor('#E1E5E9');cell.set_linewidth(.4)
    if r==0:cell.set_facecolor('#E8F0F7');cell.set_text_props(fontweight='bold')
    elif r%2==0:cell.set_facecolor('#F5F7F9')
finish(fig,10,name,dict(zip('ABCDE',['Source-only selected thresholds','Nested arterial-bed threshold evaluation',
    'Paired target descriptive threshold sweep','Histology target descriptive threshold sweep','Frozen external confusion counts'])),
    'Post-result exploratory diagnostics; no target threshold optimization or recalibration. A, Source threshold rules are fixed 0.5, maximum Youden J and the largest candidate meeting source sensitivity at least 0.90. Candidates are adjacent score midpoints plus endpoints 0 and 1; Youden ties choose the value closest to 0.5, then the smaller value. The 0.90 constraint is illustrative, not a clinical target. The all-source rule uses the original first three-fold repetition, one held-out score per label group. Each anatomical rule uses a new inner three-fold cross-fit confined to the other two beds. B, Each full outer-training fit is evaluated only in its excluded bed; no source fitting or threshold selection used that bed. C–D, Outcome-dependent sweeps describe the already observed target scores; vertical lines mark the three unchanged source rules, whose values are shown in A/E. No target-best cutoff is promoted. E, TP/FN and TN/FP are tissue counts, not numbers of independent patients across cohorts. Source maxima/constraints are optimization quantities, not unbiased source performance estimates. The frozen external model and original primary 0.5 results are unchanged. Previously inspected ten-repeat averaged-source threshold calculations are separately disclosed and are not this first-repetition recipe.',
    ['threshold_rules','evaluation_metrics','threshold_response_curves'])

# Figure11: all paired failures, patient/specimen influence and exact failure accounting.
name='Individual failures and fixed-score influence boundaries'
fig=plt.figure(figsize=(W,8.0),layout='constrained');gs=fig.add_gridspec(2,2,hspace=.13,wspace=.09)
a=fig.add_subplot(gs[0,0]);sub=gs[0,1].subgridspec(2,1,height_ratios=[1,1],hspace=.23)
b1=fig.add_subplot(sub[0]);b2=fig.add_subplot(sub[1]);c=fig.add_subplot(gs[1,0]);d=fig.add_subplot(gs[1,1]);title(fig,name)
panel(a,'A','Every paired ordering across five rules')
mat=signs.pivot(index='patient',columns='model',values='paired_ordering').sort_index().loc[:,MODEL_ORDER]
a.imshow(mat.to_numpy(),aspect='auto',interpolation='none',cmap=ListedColormap([RED,'#FFFFFF',BLUE]),vmin=-1,vmax=1)
tick_ids=[1,4,8,12,17,21,24,28,32]
tick_positions=[list(mat.index).index(p) for p in tick_ids]
a.set_yticks(tick_positions,[str(p) for p in tick_ids]);a.set_xticks(range(5),[MODEL_NAMES[v] for v in MODEL_ORDER],rotation=35,ha='right')
a.set(ylabel='Known paired patient identifier')
for tick,p in zip(a.get_yticklabels(),tick_ids):
    if margins.loc[margins.patient==p,'retrospective_reversed_pair'].iloc[0]:tick.set_color(RED);tick.set_fontweight('bold')
a.legend(handles=[Patch(color=BLUE,label='Plaque > intact'),Patch(color=RED,label='Plaque < intact')],
         loc='upper left',bbox_to_anchor=(-.04,-.20),frameon=False,ncol=1)
panel(b1,'B','Delete one paired patient; keep scores fixed')
for i,model in enumerate(MODEL_ORDER):
    bb=pdels[pdels.model==model]
    jit=rng.uniform(-.14,.14,len(bb))
    b1.scatter(np.full(len(bb),i)+jit,bb.auc_change,s=8,color=BLUE,alpha=.65,linewidths=0)
    b2.scatter(np.full(len(bb),i)+jit,bb.pair_concordance_change,s=8,color=ORANGE,alpha=.65,linewidths=0)
for b in [b1,b2]:
    b.axhline(0,color=GRAY,lw=.7,ls='--');b.set_xticks(range(5),[MODEL_NAMES[v] for v in MODEL_ORDER],rotation=35,ha='right');grid(b,'y')
b1.set(ylabel='Change in marginal AUROC');b2.set(ylabel='Change in paired concordance')
b1.tick_params(labelbottom=False)
panel(c,'C','Delete one histology specimen; keep scores fixed')
for i,model in enumerate(MODEL_ORDER):
    bb=hdels[hdels.model==model]
    c.scatter(bb.auc_change,np.full(len(bb),i)+rng.uniform(-.16,.16,len(bb)),s=9,color=BLUE,alpha=.65,linewidths=0)
c.axvline(0,color=GRAY,lw=.7,ls='--');c.set_yticks(range(5),[MODEL_NAMES[v] for v in MODEL_ORDER]);c.invert_yaxis()
c.set(xlabel='Change in advanced-versus-early AUROC',ylabel='Frozen scoring rule',ylim=(5.0,-.5));grid(c)
c.text(.02,.01,'29 specimens; donors unverified',transform=c.transAxes,color=GRAY,fontsize=8,
       bbox={'facecolor':'white','edgecolor':'none','pad':1})
panel(d,'D','Accounting for three reversed primary pairs')
failed=margins.loc[margins.retrospective_reversed_pair,'patient'].tolist()
display_genes=genes[:5];rows=display_genes+['Other 195','Total']
matrix=[]
for gene in rows:
    vals=[]
    for patient in failed:
        pp=pdiffs[pdiffs.patient==patient]
        if gene=='Total':value=pp.weighted_logit_difference.sum()
        elif gene=='Other 195':value=pp.loc[~pp.gene.isin(display_genes),'weighted_logit_difference'].sum()
        else:value=pp.loc[pp.gene==gene,'weighted_logit_difference'].iloc[0]
        vals.append(value)
    matrix.append(vals)
matrix=np.array(matrix);assert np.allclose(matrix[:-1].sum(axis=0),matrix[-1],atol=1e-10)
lim=float(np.max(abs(matrix)));im=d.imshow(matrix,aspect='auto',cmap=diverge,norm=TwoSlopeNorm(vmin=-lim,vcenter=0,vmax=lim))
d.set_xticks(range(len(failed)),[str(int(v)) for v in failed]);d.set_yticks(range(len(rows)),rows)
d.set(xlabel='Retrospectively identified reversed pair',ylabel='Source-selected terms + remainder')
for ii in range(len(rows)):
    for jj in range(len(failed)):
        d.text(jj,ii,f'{matrix[ii,jj]:.2f}',ha='center',va='center',fontsize=8,
               color='white' if abs(matrix[ii,jj])>.7*lim else '#18212A',fontweight='bold' if ii==len(rows)-1 else 'normal')
cb=fig.colorbar(im,ax=d,orientation='horizontal',pad=.04,shrink=.9);cb.set_label('Plaque minus intact logit contribution')
finish(fig,11,name,dict(zip('ABCD',['Complete paired rule-ordering map','Patient-block deletion influence for two estimands',
    'Histology specimen deletion influence','Exact linear accounting of retrospective primary failures'])),
    'Post-result exploratory failure diagnostics. A, All 32 known patient pairs and five fixed expression rules are shown in numerical patient order; a subset of axis identifiers is labeled for legibility. Blue means plaque scores above intact tissue and red means the reverse. No ties occurred. Bold red identifiers mark the three primary ridge reversals, selected retrospectively as failures; colors compare ordering signs, not raw score scales across models. B, Each point deletes both tissues from one patient while retaining every other frozen score; upper and lower displays concern marginal AUROC and within-patient concordance, respectively. These are influence descriptions, not refits, bootstrap draws or confidence intervals. C, Analogous fixed-score deletion removes one of 29 histology specimens; donor dependence remains unknown. D, Coefficient-weighted paired rank differences exactly reconstruct the plaque-minus-intact logit margin. Five named genes are the largest absolute coefficients in the full-source model, not chosen from these failed patients. The other 195 genes are summed; the intercept and source center cancel in paired differences. This algebra does not establish biological causes of failure. The three failed pairs do not represent an independently selected validation set.',
    ['paired_five_rule_sign_map','paired_deletion_influence','histology_deletion_influence',
     'paired_feature_logit_differences','paired_primary_margins','top20_source_coefficient_genes'])

(OUT/'threshold_extension_captions.json').write_text(json.dumps(captions,ensure_ascii=False,indent=2),encoding='utf8')
(OUT/'threshold_extension_qa.json').write_text(json.dumps(checks,indent=2),encoding='utf8')
manifest={'script':'code/plot_threshold_extension.py','analysis_status':'Post-result exploratory, not prespecified',
    'input_sha256':inputs,'main_figures':3,'main_panels':sum(c['panel_count'] for c in captions.values()),
    'font':'DejaVu Sans; minimum8pt at17cm','files':{}}
for i in [9,10,11]:
    for ext in ['pdf','svg','png']:
        p=OUT/f'Figure_{i}.{ext}';manifest['files'][p.name]={'sha256':hashlib.sha256(p.read_bytes()).hexdigest(),'bytes':p.stat().st_size}
(OUT/'threshold_extension_provenance.json').write_text(json.dumps(manifest,indent=2),encoding='utf8')
sheet=Image.new('RGB',(2700,1250),'#E5E9ED')
for j,i in enumerate([9,10,11]):
    with Image.open(OUT/f'Figure_{i}.png') as source:im=source.convert('RGB')
    im.thumbnail((850,1200));sheet.paste(im,(j*900+(900-im.width)//2,35))
    ImageDraw.Draw(sheet).text((j*900+15,10),f'Figure{i}',fill='black')
sheet.save(OUT/'threshold_extension_contact.png')
print(json.dumps({'figures':3,'panels':manifest['main_panels'],'qa':checks},indent=2))
```
