## Supplementary material for "Anatomical comparison populations and threshold transport in vascular transcriptomic classification": Reproducibility archive: code, provenance and results: prepare.py

```
"""Deterministic annotation, sample-wise representation, and declared identity units."""
from pathlib import Path
import pandas as pd, numpy as np, gzip,io,json,re,hashlib,datetime
from metadata import metadata
ROOT=Path(__file__).resolve().parents[1]
PLAT={'GSE100927':'GPL17077','GSE43292':'GPL6244','GSE28829':'GPL570'}

def matrix(acc):
    with gzip.open(ROOT/'data'/f'{acc}_series_matrix.txt.gz','rt') as f: text=f.read()
    tab=text.split('!series_matrix_table_begin\n',1)[1].split('!series_matrix_table_end',1)[0]
    x=pd.read_csv(io.StringIO(tab),sep='\t',index_col=0)
    x.index=x.index.astype(str)
    return x.apply(pd.to_numeric,errors='coerce')

def annotation(gpl):
    text=(ROOT/'data'/f'{gpl}_platform_table.txt').read_text(encoding='utf8')
    tab=text.split('!platform_table_begin\n',1)[1].split('!platform_table_end',1)[0]
    a=pd.read_csv(io.StringIO(tab),sep='\t',dtype=str,keep_default_na=False)
    key={'GPL17077':'GENE_SYMBOL','GPL570':'Gene Symbol','GPL6244':'gene_assignment'}[gpl]
    out=[]
    for probe,v in zip(a.ID,a[key]):
        if gpl=='GPL6244':
            vals={p.split(' // ')[1].strip() for p in v.split(' /// ') if len(p.split(' // '))>1}
        else: vals={p.strip() for p in re.split(r'\s*///\s*|\s*//\s*|\s*;\s*',v)}
        vals={s for s in vals if s and s not in ['---','NA','nan']}
        if len(vals)==1:
            s=next(iter(vals))
            if re.fullmatch(r'[A-Za-z0-9_.-]+',s):out.append((probe,s))
    aout=pd.DataFrame(out,columns=['probe','gene'])
    assert not aout.probe.duplicated().any()
    aout.to_csv(ROOT/'evidence'/f'{gpl}_unambiguous_mapping.csv',index=False)
    return aout

def sample_table(acc):
    rec=metadata(acc);out=[]
    for r in rec:
        title=r['title']
        if acc=='GSE100927':
            suffix=title.split(': ',1)[1]
            bed=next(b for b in ['Carotid','Femoral','Infrapopliteal'] if b.lower() in r['source_name_ch1'].lower().replace('-',''))
            y=int('Atherosclerotic' in r['source_name_ch1'])
            sex=r['characteristics_ch1_2'].split(': ')[1]
            label=suffix
            proxy=('AS_' if y else 'Control_')+suffix.split('_')[-1].replace('Control','')
            patient='unknown'
        elif acc=='GSE43292':
            bed='Carotid';y=int('Atheroma plaque' in title);sex='unknown'
            patient=r['characteristics_ch1'].split(': ')[1];label=title;proxy=patient
        else:
            bed='Carotid';y=int('advanced' in title);sex='unknown';patient='unknown';label=title;proxy=title
        out.append(dict(sample=r['geo_accession'],title=title,bed=bed,y=y,sex=sex,label_group=label,identity_proxy=proxy,patient=patient))
    return pd.DataFrame(out).set_index('sample')

if __name__=='__main__':
    mapped={};meta={};qc={}
    for acc,gpl in PLAT.items():
        x=matrix(acc);a=annotation(gpl).set_index('probe')
        ids=x.index.intersection(a.index)
        gx=x.loc[ids].groupby(a.loc[ids,'gene'].values).median()
        mapped[acc]=gx;meta[acc]=sample_table(acc)
        assert list(x.columns)==list(meta[acc].index)
        q=np.nanpercentile(x.to_numpy(),[0,1,50,99,100])
        qc[acc]={'matrix_probes':len(x),'platform_mapped_probes':len(a),'matched_probes':len(ids),'unique_genes':len(gx),'arrays':x.shape[1],'quantiles':q.tolist(),'missing_values':int(x.isna().sum().sum())}
    common=sorted(set.intersection(*(set(x.index) for x in mapped.values())))
    (ROOT/'evidence'/'common_gene_vocabulary.txt').write_text('\n'.join(common)+'\n')
    qc['common_genes']=len(common)
    for acc,gx in mapped.items():
        gx=gx.loc[common].T
        # Monotone intensity transformation only for native-expression sensitivity.
        log2=qc[acc]['quantiles'][3]>100
        native=np.log2(gx.clip(lower=0)+1) if log2 else gx
        rank=gx.rank(axis=1,pct=True).fillna(.5)
        native.to_pickle(ROOT/'data'/f'{acc}_native.pkl')
        rank.to_pickle(ROOT/'data'/f'{acc}_rank.pkl')
        meta[acc].to_csv(ROOT/'data'/f'{acc}_samples.csv')
        qc[acc]['native_log2_transformed']=bool(log2)
    qc['prepared_utc']=datetime.datetime.now(datetime.timezone.utc).isoformat()
    (ROOT/'evidence'/'preprocessing_qc.json').write_text(json.dumps(qc,indent=2))
    print(json.dumps(qc,indent=2),flush=True)
```
