## Supplementary material for "Anatomical comparison populations and threshold transport in vascular transcriptomic classification": Reproducibility archive: code, provenance and results: render_review.py

```
"""Render current manuscript PDFs using Poppler, with hash-keyed page directories.

Requires requirements-qa.txt and pdftoppm on PATH or --pdftoppm PATH.
"""
from pathlib import Path
import argparse, hashlib, json, shutil, subprocess
from PIL import Image, ImageDraw
from pypdf import PdfReader
import fitz

ROOT=Path(__file__).resolve().parents[1]

def main():
    parser=argparse.ArgumentParser(description=__doc__)
    parser.add_argument('--pdftoppm', default=shutil.which('pdftoppm'))
    args=parser.parse_args()
    if not args.pdftoppm:
        parser.error('Install Poppler and add pdftoppm to PATH, or pass --pdftoppm PATH')
    for name in ['main','supplement']:
        pdf=ROOT/'manuscript'/f'{name}.pdf'
        digest=hashlib.sha256(pdf.read_bytes()).hexdigest()
        d=ROOT/'qa'/f'{name}_{digest[:12]}'
        d.mkdir(parents=True,exist_ok=True)
        subprocess.run([args.pdftoppm,'-r','85','-png',str(pdf),str(d/'page')],check=True,stdout=subprocess.DEVNULL,stderr=subprocess.PIPE)
        files=sorted(d.glob('page-*.png'), key=lambda p:int(p.stem.split('-')[-1]))
        reader=PdfReader(pdf)
        if len(files)!=len(reader.pages): raise ValueError('Rendered page count differs from current PDF')
        tiles=[]
        for p in files:
            with Image.open(p) as source: im=source.convert('RGB')
            im.thumbnail((340,480))
            tile=Image.new('RGB',(360,510),'#dfe4e8');tile.paste(im,((360-im.width)//2,25))
            ImageDraw.Draw(tile).text((12,7),p.stem,fill='black');tiles.append(tile)
        ncol=4;nrow=(len(tiles)+ncol-1)//ncol
        sheet=Image.new('RGB',(ncol*360,nrow*510),'#dfe4e8')
        for i,t in enumerate(tiles):sheet.paste(t,((i%ncol)*360,(i//ncol)*510))
        sheet.save(ROOT/'qa'/f'{name}_contact.png')
        text='\n'.join(p.extract_text() or '' for p in reader.pages)
        (ROOT/'qa'/f'{name}_extracted.txt').write_text(text,encoding='utf8')
        problems=[]
        with fitz.open(pdf) as doc:
            skipped=[]
            for i,p in enumerate(doc):
                if p.rotation:
                    skipped.append(i+1);continue
                for b in p.get_text('blocks'):
                    if b[0]<-1 or b[1]<-1 or b[2]>p.rect.width+1 or b[3]>p.rect.height+1:
                        problems.append({'page':i+1,'bbox':list(b[:4]),'text':str(b[4])[:150]})
            report={'pages':len(doc),'pdf_sha256':digest,'render_directory':str(d.relative_to(ROOT)).replace('\\','/'),
                    'outside_page_text_blocks':problems,'rotated_pages_not_checked_by_bounds_heuristic':skipped,
                    'scope':'Text-bound heuristic only; visual inspection remains necessary.'}
        (ROOT/'qa'/f'{name}_bounds.json').write_text(json.dumps(report,indent=2),encoding='utf8')
        print(name,len(reader.pages),'pages',len(problems),'outside blocks; rendered to',d)

if __name__=='__main__': main()
```
