## Supplementary material for "Anatomical comparison populations and threshold transport in vascular transcriptomic classification": Reproducibility archive: code, provenance and results: validate_exploratory.py

```
"""Independent, read-only verification of the post-result exploratory extension.

Does not import either exploration implementation. Saved-score recomputation plus
limited independent fitting checks do not constitute a clean-environment refit.
"""
from pathlib import Path
import datetime, hashlib, json, itertools
import numpy as np
import pandas as pd
from scipy.special import expit
from sklearn.metrics import roc_auc_score, average_precision_score, brier_score_loss
from sklearn.model_selection import StratifiedKFold
from sklearn.preprocessing import StandardScaler
from sklearn.linear_model import LogisticRegression

ROOT=Path(__file__).resolve().parents[1]
F=ROOT/'results/exploratory_features'; T=ROOT/'results/exploratory_threshold'
E=ROOT/'evidence/figure_revision_20261026'
errors=[]; checks={}; inputs={}; discrepancies=[]
def sha(p):return hashlib.sha256(p.read_bytes()).hexdigest()
def require(ok,msg):
    if not bool(ok):errors.append(msg)
def close(a,b,msg,tol=1e-9):
    a,b=np.asarray(a,dtype=float),np.asarray(b,dtype=float)
    d=float(np.max(np.abs(a-b))) if a.size else 0.
    discrepancies.append(d)
    require(a.shape==b.shape and np.isfinite(d) and d<tol, msg+f'; max_abs_error={d}')
def read(path):
    inputs[str(path.relative_to(ROOT)).replace('\\','/')]=sha(path)
    return pd.read_csv(path)
def metric(y,prob,rank):
    y=np.asarray(y);prob=np.asarray(prob);rank=np.asarray(rank)
    pred=prob>=.5
    sen=float(pred[y==1].mean());spe=float((~pred[y==0]).mean())
    return dict(auc=roc_auc_score(y,rank),ap=average_precision_score(y,rank),sensitivity=sen,
        specificity=spe,balanced_accuracy=(sen+spe)/2,brier=brier_score_loss(y,prob))
def independent_source():
    xx=pd.read_pickle(ROOT/'data/GSE100927_rank.pkl')
    mm=read(ROOT/'data/GSE100927_samples.csv').set_index('sample')
    return xx.groupby(mm.label_group,sort=True).median(),mm.groupby('label_group',sort=True).first()
def gene_order(x,y):
    a=x.to_numpy(); y=np.asarray(y)
    t=(a[y==1].mean(0)-a[y==0].mean(0))/np.sqrt(
        a[y==1].var(0,ddof=1)/sum(y==1)+a[y==0].var(0,ddof=1)/sum(y==0)+1e-12)
    return np.argsort(-np.abs(t),kind='stable')
def independent_fit(x,y,genes,seed):
    scale=StandardScaler().fit(x[genes])
    model=LogisticRegression(C=.1,class_weight='balanced',solver='liblinear',max_iter=2000,random_state=seed)
    model.fit(scale.transform(x[genes]),y)
    return scale,model
def operating(y,score,t):
    y=np.asarray(y);p=np.asarray(score)>=t
    tp=int(np.sum(p&(y==1)));tn=int(np.sum(~p&(y==0)))
    fn=int(np.sum(~p&(y==1)));fp=int(np.sum(p&(y==0)))
    sen=tp/(tp+fn);spe=tn/(tn+fp)
    return dict(tp=tp,tn=tn,fp=fp,fn=fn,sensitivity=sen,specificity=spe,
        balanced_accuracy=(sen+spe)/2,youden_j=sen+spe-1,positive_predictions=int(p.sum()),n=len(y))

# Historical output bytes must be identical to the pre-extension snapshot.
snap=json.loads((E/'pre_expansion/manifest.json').read_text())['sha256']
old={n:h for n,h in snap.items() if n.startswith('results/')}
for n,h in old.items():require(sha(ROOT/n)==h,'Original result changed: '+n)
checks['original_core_result_hashes_unchanged']=len(old)
x,m=independent_source(); require(x.shape==(95,15235),'Source dimensions')
genes=read(F/'random_panel_genes.csv')
rng=np.random.default_rng(20261026)
panel_genes={}
for i in range(20):
    p=f'random_{i+1:02d}'
    expected=sorted(rng.choice(x.columns.tolist(),200,replace=False))
    observed=genes.loc[genes.panel==p,'gene'].tolist()
    require(observed==expected,'Random panel differs from label-independent seeded draw '+p)
    panel_genes[p]=observed
require(len(genes)==4000 and genes.groupby('panel').gene.nunique().eq(200).all(),'Random panel counts')

pred=read(F/'representation_cv_predictions.csv'); fm=read(F/'representation_fold_metrics.csv')
ranking_column='linear_logit'
has_logit=ranking_column in pred
require(has_logit,'Feature-extension predictions must retain linear_logit to avoid sigmoid saturation in ranking')
if not has_logit:ranking_column='score'
if has_logit:
    close(pred.score,expit(pred.linear_logit),'Source probability/logit consistency')
    require(np.array_equal(pred.score.to_numpy()>=.5,pred.linear_logit.to_numpy()>=0),
            'Source probability0.5 and logit0 operating decisions differ')
original_pred=read(ROOT/'results/primary_predictions.csv')
original_pred=original_pred[original_pred.model=='ridge200']
fold_index=fm.set_index(['panel','repeat','fold'])
require(len(fm)==630 and not fold_index.index.duplicated().any(),'630 unique panel-fold records')
for (panel,rep,fold),block in pred.groupby(['panel','repeat','fold']):
    r=fold_index.loc[(panel,rep,fold)]
    ref=original_pred[(original_pred['repeat']==rep)&(original_pred.fold==fold)]
    require(set(block['sample'])==set(ref['sample']),f'Unmatched source fold {panel}/{rep}/{fold}')
    vals=metric(block.y,block.score,block[ranking_column])
    for name,value in vals.items():close(r[name],value,f'Source {panel}/{rep}/{fold}/{name}')
    beds=['Carotid','Femoral','Infrapopliteal'];mat=np.zeros((3,3));w=np.zeros((3,3))
    for a,ca in enumerate(beds):
        for b,cb in enumerate(beds):
            case=block.loc[(block.y==1)&(block.bed==ca),ranking_column].to_numpy()
            control=block.loc[(block.y==0)&(block.bed==cb),ranking_column].to_numpy()
            dif=case[:,None]-control[None,:]
            mat[a,b]=np.mean((dif>0)+.5*(dif==0));w[a,b]=dif.size
    pooled=np.sum(mat*w)/w.sum();diag=np.trace(mat)/3
    for name,value in dict(auc=pooled,within_bed_auc=diag,delta_mix=pooled-diag,
            mix_min=mat.min(),mix_max=mat.max(),mix_span=mat.max()-mat.min(),
            cross_bed_pair_fraction=1-np.trace(w)/w.sum()).items():
        close(r[name],value,f'Source nine-cell {panel}/{rep}/{fold}/{name}')
    require(len(block)==r.n_test and r.n_train+r.n_test==95,'Train/test count differs')
for (panel,rep),block in pred.groupby(['panel','repeat']):
    require(len(block)==95 and block['sample'].nunique()==95,'Source repeat partition overlap/missing')
checks['source_fold_records_recomputed']=len(fm)

# Recover membership from each original training complement; check all 6000 rows.
features=read(F/'primary_fold_features.csv'); sets={}; limited_refits=0
for rep in range(10):
    strata=m.bed.astype(str)+'_'+m.y.astype(str)
    sk=StratifiedKFold(3,shuffle=True,random_state=20261025+rep)
    for fold,(tr,te) in enumerate(sk.split(m,strata)):
        xx=x.iloc[tr]; yy=m.y.iloc[tr]
        order=gene_order(xx,yy); expected=x.columns[order[:200]].tolist()
        rows=features[(features['repeat']==rep)&(features.fold==fold)].sort_values('rank')
        require(rows.gene.tolist()==expected,f'Training-only membership {rep}/{fold}')
        scale=StandardScaler().fit(xx[expected])
        close(rows.source_mean,scale.mean_,f'Training means {rep}/{fold}')
        close(rows.source_sd,scale.scale_,f'Training scales {rep}/{fold}')
        sets[rows.fit_id.iloc[0]]=set(expected)
        if rep==0 and fold==0:
            for panel,gs in [('next200',x.columns[order[200:400]].tolist()),('random_01',panel_genes['random_01'])]:
                scale,model=independent_fit(xx,yy,gs,20261025)
                actual=model.predict_proba(scale.transform(x.iloc[te][gs]))[:,1]
                logits=model.decision_function(scale.transform(x.iloc[te][gs]))
                observed=pred[(pred.panel==panel)&(pred['repeat']==rep)&(pred.fold==fold)].set_index('sample').loc[m.index[te]]
                close(observed.score,actual,'Limited independent refit '+panel)
                if has_logit:close(observed.linear_logit,logits,'Limited independent logit refit '+panel)
                limited_refits+=1
stability=read(F/'feature_stability.csv').set_index('gene')
frequency=features.gene.value_counts().reindex(x.columns,fill_value=0)
require(stability.index.tolist()==x.columns.tolist(),'Full vocabulary absent from stability export')
close(stability.selected_folds,frequency,'All-gene selection counts')
jac=read(F/'feature_jaccard.csv')
require(len(jac)==435,'Feature Jaccard pair count')
for row in jac.itertuples():
    a,b=sets[row.fit_a],sets[row.fit_b]
    close(row.jaccard,len(a&b)/len(a|b),'Feature Jaccard')
checks['feature_membership']={'training_fits':30,'rows':len(features),'union':int(sum(frequency>0)),
    'all_30':int(sum(frequency==30)),'all_vocabulary_genes':len(stability),'limited_independent_refits':limited_refits}

ep=read(F/'representation_external_predictions.csv'); em=read(F/'representation_external_metrics.csv').set_index(['panel','dataset'])
require('linear_logit' in ep,'External panel predictions lack unsaturated linear logits')
erank='linear_logit' if 'linear_logit' in ep else 'score'
if erank=='linear_logit':
    close(ep.score,expit(ep.linear_logit),'External probability/logit consistency')
    require(np.array_equal(ep.score.to_numpy()>=.5,ep.linear_logit.to_numpy()>=0),
            'External probability0.5 and logit0 operating decisions differ')
saturation=[]
for (panel,dataset),block in ep.groupby(['panel','dataset']):
    r=em.loc[(panel,dataset)]
    vals=metric(block.y,block.score,block[erank])
    for name,value in vals.items():close(r[name],value,f'External {panel}/{dataset}/{name}')
    if dataset=='GSE43292':
        diffs=[]
        require(block.groupby('patient').size().eq(2).all(),'External pair sizes')
        for patient,pair in block.groupby('patient'):
            require(pair.y.sum()==1,'External pair classes')
            diffs.append(pair.loc[pair.y==1,erank].iloc[0]-pair.loc[pair.y==0,erank].iloc[0])
        d=np.asarray(diffs)
        close(r.pair_concordance,np.mean((d>0)+.5*(d==0)),f'External pair concordance {panel}')
    if block.score.nunique()<len(block):
        saturation.append(dict(panel=panel,dataset=dataset,probability_unique=int(block.score.nunique()),
            ranking_unique=int(block[erank].nunique()),at_zero=int(sum(block.score==0)),at_one=int(sum(block.score==1)),
            probability_auc=roc_auc_score(block.y,block.score),linear_logit_auc=roc_auc_score(block.y,block[erank])))
checks['external_panel_records_recomputed']=len(em)
checks['probability_saturation_diagnostic']=saturation

# Source-only threshold selection and held-out anatomy structure.
learning=read(T/'source_crossfit_learning_scores.csv')
rules=read(T/'threshold_rules.csv'); tc=read(T/'source_threshold_candidates.csv')
require(len(learning)==285 and len(rules)==12,'Threshold source denominators')
thresholds={}
for context,block in learning.groupby('learning_context'):
    require(block['sample'].nunique()==len(block),'Repeated threshold-learning label')
    if context=='all_source':
        require(set(block['sample'])==set(m.index),'Full source OOF coverage')
        saved=original_pred[original_pred['repeat']==0].set_index('sample').loc[block['sample']]
        close(block.score,saved.score,'Full source OOF exact original repeat0')
    else:
        target=context.replace('LOBO_to_','')
        require(set(block['sample'])==set(m.index[m.bed!=target]),'Outer target contamination '+context)
        require(not (block.bed==target).any(),'Outer target bed enters threshold learning')
        expected_seed=20261026+['Carotid','Femoral','Infrapopliteal'].index(target)
        sub=m.loc[block['sample']].sort_index()
        strata=sub.bed.astype(str)+'_'+sub.y.astype(str)
        for fold,(tr,te) in enumerate(StratifiedKFold(3,shuffle=True,random_state=expected_seed).split(sub,strata)):
            require(set(sub.index[te])==set(block.loc[block.fold==fold,'sample']), 'Inner fold partition mismatch '+context)
            require(not set(sub.index[tr]) & set(sub.index[te]),'Inner overlap')
    u=np.sort(block.score.unique()); candidates=np.unique(np.r_[0.,(u[:-1]+u[1:])/2,1.])
    values=pd.DataFrame([dict(threshold=t,**operating(block.y,block.score,t)) for t in candidates])
    stored=tc[tc.learning_context==context].sort_values('threshold')
    close(stored.threshold,values.threshold,'Candidate grid '+context)
    for col in ['sensitivity','specificity','balanced_accuracy','youden_j']:
        close(stored[col],values[col],'Source candidate metric '+context+'/'+col)
    tied=values[np.isclose(values.youden_j,values.youden_j.max(),rtol=0,atol=1e-12)].copy()
    tied['distance']=abs(tied.threshold-.5)
    best=float(tied.sort_values(['distance','threshold']).iloc[0].threshold)
    high=float(values.loc[values.sensitivity>=.90-1e-12,'threshold'].max())
    expected={'fixed_0.5':.5,'source_max_youden':best,'source_sensitivity_0.90':high}
    thresholds[context]=expected
    for rule,value in expected.items():
        row=rules[(rules.learning_context==context)&(rules.rule==rule)].iloc[0]
        close(row.threshold,value,'Chosen threshold '+context+'/'+rule)
        for col,number in operating(block.y,block.score,value).items():close(row[col],number,'Chosen threshold source quantity')
evalp=read(T/'evaluation_predictions.csv'); evalm=read(T/'evaluation_metrics.csv')
for (dataset,context),block in evalp.groupby(['dataset','learning_context']):
    if dataset.startswith('LOBO_'):
        require(not set(block['sample'])&set(learning.loc[learning.learning_context==context,'sample']), 'Threshold train/test overlap')
    for rule,t in thresholds[context].items():
        r=evalm[(evalm.dataset==dataset)&(evalm.rule==rule)].iloc[0]
        close(r.threshold,t,'Evaluation threshold')
        for col,value in operating(block.y,block.score,t).items():close(r[col],value,'Target operating result')
        close(r.auc,roc_auc_score(block.y,block.score),'Target ranking')
        close(block[rule+'_prediction'],(block.score>=t).astype(int),'Saved target predicted class')
curves=read(T/'threshold_response_curves.csv')
for dataset,block in evalp.groupby('dataset'):
    for row in curves[curves.dataset==dataset].itertuples():
        for name,value in operating(block.y,block.score,row.threshold).items():close(getattr(row,name),value,'Descriptive curve '+name)
checks['source_only_thresholds']={'learning_rows':len(learning),'rules':len(rules),'evaluations':len(evalm),
    'all_source':thresholds['all_source'],'curve_rows_recomputed':len(curves),
    'scope':'Code and exact split audit confirms threshold learning excludes outer target bed; curves use targets descriptively only'}

# Independent arithmetic from actual rank matrices, not saved contributions alone.
coef=read(ROOT/'results/frozen_primary_coefficients.csv')
frozen=json.loads((ROOT/'results/frozen_primary_model.json').read_text())
intercept=frozen['intercept']
geometry=read(T/'score_geometry_samples.csv')
contrib=read(T/'feature_contributions_all.csv')
means=read(T/'mean_feature_contributions.csv')
decomp=read(T/'class_logit_decomposition.csv')
zstats=read(T/'standardization_displacement.csv')
for dataset in ['GSE100927','GSE43292','GSE28829']:
    xx,mm=(x,m) if dataset=='GSE100927' else (pd.read_pickle(ROOT/'data'/f'{dataset}_rank.pkl'),
        read(ROOT/'data'/f'{dataset}_samples.csv').set_index('sample'))
    z=(xx[coef.gene].to_numpy()-coef.source_mean.to_numpy())/coef.source_sd.to_numpy()
    c=z*coef.coefficient.to_numpy(); eta=intercept+c.sum(1)
    gs=geometry[geometry.dataset==dataset].set_index('sample').loc[xx.index]
    close(gs.linear_logit,eta,'Frozen logit reconstruction '+dataset)
    close(gs.probability,expit(eta),'Frozen probability reconstruction '+dataset)
    saved=contrib[contrib.dataset==dataset].pivot(index='sample',columns='gene',values='weighted_contribution').loc[xx.index,coef.gene]
    close(saved,c,'Every sample-gene contribution '+dataset)
    for state,mask in [('all',np.ones(len(mm),bool)),('0',mm.y.to_numpy()==0),('1',mm.y.to_numpy()==1)]:
        cm=c[mask].mean(0)
        row=decomp[(decomp.dataset==dataset)&(decomp.class_state==state)].iloc[0]
        close(row.mean_logit,eta[mask].mean(),'Class mean logit '+dataset+'/'+state)
        close(row.positive_mean_feature_sum,np.maximum(cm,0).sum(),'Sum positive gene means')
        close(row.negative_mean_feature_sum,np.minimum(cm,0).sum(),'Sum negative gene means')
        zr=zstats[(zstats.dataset==dataset)&(zstats.class_state==state)].iloc[0]
        close(zr.fraction_abs_z_gt3,np.mean(abs(z[mask])>3),'Outlier fraction')
        close(zr.median_abs_z,np.median(abs(z[mask])),'Median abs z')
    row=decomp[(decomp.dataset==dataset)&(decomp.class_state=='class_balanced')].iloc[0]
    close(row.mean_logit,(eta[mm.y==0].mean()+eta[mm.y==1].mean())/2,'Equal-class mean logit')
checks['frozen_geometry']={'sample_records':len(geometry),'gene_contribution_records':len(contrib),
    'scope':'Exact classifier arithmetic; source in-sample reference, no platform-specific cause or causal biology established'}
top=read(T/'top20_source_coefficient_genes.csv')
expected_top=coef.assign(abs_coefficient=abs(coef.coefficient)).sort_values(['abs_coefficient','gene'],ascending=[False,True]).head(20)
require(top.gene.tolist()==expected_top.gene.tolist(),'Display genes were not selected only by source absolute coefficient')
pm=read(T/'paired_primary_margins.csv'); pc=read(T/'paired_feature_logit_differences.csv')
sg=geometry[geometry.dataset=='GSE43292']
for row in pm.itertuples():
    block=sg[sg.patient.astype(str)==str(row.patient)]
    difference=float(block.loc[block.y==1,'linear_logit'].iloc[0]-block.loc[block.y==0,'linear_logit'].iloc[0])
    close(row.logit_difference,difference,'Paired primary logit margin')
    close(pc.loc[pc.patient==row.patient,'weighted_logit_difference'].sum(),difference,'Paired gene contributions sum')
checks['paired_geometry']={'known_pairs':len(pm),'pair_gene_rows':len(pc),'source_only_display_genes':len(top)}

ext=read(ROOT/'results/external_predictions.csv')
epairs=read(ROOT/'results/external_pairs.csv')
pdels=read(T/'paired_deletion_influence.csv'); hdels=read(T/'histology_deletion_influence.csv')
for row in pdels.itertuples():
    block=ext[(ext.dataset=='GSE43292')&(ext.model==row.model)]
    keep=block[block.patient.astype(str)!=str(row.omitted_patient)]
    pairs=epairs[(epairs.model==row.model)&(epairs.patient.astype(str)!=str(row.omitted_patient))]
    require(len(keep)==62 and len(pairs)==31,'Patient block deletion size')
    close(row.auc,roc_auc_score(keep.y,keep.score),'Patient deletion AUROC')
    close(row.auc_change,row.auc-roc_auc_score(block.y,block.score),'Patient deletion AUROC change')
    close(row.pair_concordance,np.mean((pairs.difference>0)+.5*(pairs.difference==0)),'Patient deletion concordance')
for row in hdels.itertuples():
    block=ext[(ext.dataset=='GSE28829')&(ext.model==row.model)]
    keep=block[block['sample']!=row.omitted_sample]
    require(len(keep)==28,'Histology deletion size')
    close(row.auc,roc_auc_score(keep.y,keep.score),'Specimen deletion AUROC')
checks['fixed_score_case_influence']={'paired_rows':len(pdels),'histology_rows':len(hdels),
    'interpretation':'Deletes fixed known patient blocks or specimens, not model fitting/CV/CI'}

for name,digest in inputs.items():require(sha(ROOT/name)==digest,'Input changed during independent audit: '+name)
report={'checked_utc':datetime.datetime.now(datetime.timezone.utc).isoformat(),'passed':not errors,'errors':errors,
    'maximum_absolute_numerical_discrepancy':max(discrepancies,default=0),'checks':checks,'input_sha256':inputs,
    'scope':'Independent saved-output recomputation, source-only split/selection audit and two independent fitted-panel replays. No clean environment install, full refit or human peer-review endorsement.'}
(E/'independent_extension_validation.json').write_text(json.dumps(report,indent=2),encoding='utf8')
print(json.dumps({'passed':not errors,'errors':errors,'maximum_error':report['maximum_absolute_numerical_discrepancy'],
    'report':str(E/'independent_extension_validation.json')},indent=2))
raise SystemExit(0 if not errors else 1)
```
