## Supplementary material for "Anatomical comparison populations and threshold transport in vascular transcriptomic classification": Reproducibility archive: code, provenance and results: validate_results.py

```
"""Independent saved-output checks; performs no model fitting or source mutation."""
from pathlib import Path
import datetime, hashlib, json
import numpy as np
import pandas as pd
from scipy.special import expit
from sklearn.metrics import roc_auc_score, average_precision_score, brier_score_loss

ROOT = Path(__file__).resolve().parents[1]
OUT = ROOT / "results"
TOL = 1e-10
BEDS = ["Carotid", "Femoral", "Infrapopliteal"]
SCORE_MODELS = {"C1Q", "centroid200"}
MODELS = {"prevalence", "bed_sex", "C1Q", "Wu6", "ridge200", "centroid200", "forest200"}
EXPRESSION_MODELS = MODELS - {"prevalence", "bed_sex"}
SCENARIOS = {"primary", "top50", "top500", "C001", "C1", "native", "exclude_ambiguous", "104_grouped_arrays", "possible_shared_controls"}
errors, checks, snapshots = [], {}, {}


def require(condition, message):
    if not bool(condition):
        errors.append(message)


def read(name):
    path = OUT / name
    snapshots[name] = hashlib.sha256(path.read_bytes()).hexdigest()
    return pd.read_csv(path)


def near(actual, expected, message):
    delta = abs(float(actual) - float(expected))
    require(np.isfinite(delta) and delta < TOL, message + f"; discrepancy={delta}")
    return delta


def score_metrics(y, score, probability):
    y, score = np.asarray(y), np.asarray(score)
    result = {"auc": roc_auc_score(y, score), "ap": average_precision_score(y, score)}
    if probability:
        require(np.all((score >= 0) & (score <= 1)), "Probability outside [0,1]")
        yes = score >= .5
        sensitivity = yes[y == 1].mean()
        specificity = (~yes[y == 0]).mean()
        result.update(sensitivity=sensitivity, specificity=specificity,
                      balanced_accuracy=(sensitivity + specificity) / 2,
                      brier=brier_score_loss(y, score))
    return result


required_files = {f"{s}_{suffix}.csv" for s in SCENARIOS
                  for suffix in ["fold_metrics", "predictions", "pair_cells"]}
required_files.update({"primary_analysis_units.csv", "external_predictions.csv", "external_metrics.csv",
                      "external_pairs.csv", "frozen_primary_coefficients.csv", "frozen_primary_model.json",
                      "transport_metrics.csv", "transport_predictions.csv", "sensitivity_all_fold_metrics.csv",
                      "permutation_nulls.csv", "full_pipeline_subsampling.csv",
                      "descriptive_mean_crossfit_scores.csv", "descriptive_mean_crossfit_metrics.csv"})
missing = sorted(name for name in required_files if not (OUT / name).is_file())
present_scenarios = {p.name.removesuffix("_fold_metrics.csv") for p in OUT.glob("*_fold_metrics.csv")} - {"sensitivity_all"}
if missing or present_scenarios != SCENARIOS:
    failure = {"passed": False, "errors": ["Required output inventory mismatch"], "missing_files": missing,
               "expected_scenarios": sorted(SCENARIOS), "found_scenarios": sorted(present_scenarios)}
    (ROOT/"evidence"/"independent_saved_results_validation.json").write_text(json.dumps(failure, indent=2), encoding="utf8")
    print(json.dumps(failure, indent=2))
    raise SystemExit(1)
checks["required_output_inventory"] = {"files": len(required_files), "scenarios": sorted(SCENARIOS)}

units = read("primary_analysis_units.csv")
unit_key = "label_group"
require(len(units) == 95 and units[unit_key].nunique() == 95, "Primary units not 95 unique labels")
source_meta = pd.read_csv(ROOT / "data" / "GSE100927_samples.csv")
require(len(source_meta) == 104, "Source arrays not 104")
sizes = source_meta.groupby("label_group").size()
require((sizes == 2).sum() == 9 and sizes.max() == 2, "Ambiguous full-label pair structure changed")
require((sizes == 1).sum() == 86, "Discard-ambiguous count not 86")
checks["source_denominators"] = {"arrays": len(source_meta), "label_groups": len(units),
    "ambiguous_two_array_groups": int((sizes == 2).sum()), "unambiguous_arrays": int((sizes == 1).sum()),
    "group_counts": units.groupby(["bed", "y"]).size().rename("n").reset_index().to_dict("records"),
    "interpretation": "Label groups and arrays, not authenticated distinct patients. Original source reports96arteries."}

primary = None
fold_report = {}
for metric_path in sorted(OUT.glob("*_fold_metrics.csv")):
    scenario = metric_path.name.removesuffix("_fold_metrics.csv")
    if scenario == "sensitivity_all":
        continue
    metrics = read(metric_path.name)
    pred = read(scenario + "_predictions.csv")
    cells = read(scenario + "_pair_cells.csv")
    expected_models = MODELS if scenario == "primary" else {"ridge200"}
    expected_keys = {(model, repeat, fold) for model in expected_models for repeat in range(10) for fold in range(3)}
    actual_keys = set(metrics[["model", "repeat", "fold"]].itertuples(index=False, name=None))
    require(actual_keys == expected_keys and len(metrics) == len(expected_keys), f"Incomplete fold/model inventory: {scenario}")
    require(set(pred[["model", "repeat", "fold"]].itertuples(index=False, name=None)) == expected_keys,
            f"Incomplete prediction fold/model inventory: {scenario}")
    max_error, n_complete, n_missing = 0., 0, 0
    require(np.isfinite(pred.score).all(), f"Nonfinite scores in {scenario}")
    for key, block in pred.groupby(["model", "repeat", "fold"]):
        model, repeat, fold = key
        mr = metrics[(metrics.model == model) & (metrics.repeat == repeat) & (metrics.fold == fold)]
        require(len(mr) == 1, f"Nonunique metric key: {scenario}/{key}")
        if len(mr) != 1:
            continue
        mr = mr.iloc[0]
        require(len(block) == int(mr.n_test), f"n_test mismatch: {scenario}/{key}")
        for name, value in score_metrics(block.y, block.score, model not in SCORE_MODELS).items():
            max_error = max(max_error, near(mr[name], value, f"{scenario}/{key}/{name}"))
        complete = all(np.any((block.y == label) & (block.bed == bed)) for label in [0, 1] for bed in BEDS)
        cr = cells[(cells.model == model) & (cells.repeat == repeat) & (cells.fold == fold)]
        if not complete:
            n_missing += 1
            require(len(cr) == 0, f"Saved cells despite missing stratum: {scenario}/{key}")
            continue
        n_complete += 1
        mat, weights = np.empty((3, 3)), np.empty((3, 3))
        for ia, case_bed in enumerate(BEDS):
            for ib, control_bed in enumerate(BEDS):
                case = block.loc[(block.y == 1) & (block.bed == case_bed), "score"].to_numpy()
                control = block.loc[(block.y == 0) & (block.bed == control_bed), "score"].to_numpy()
                diff = case[:, None] - control[None, :]
                value = ((diff > 0) + .5 * (diff == 0)).mean()
                mat[ia, ib], weights[ia, ib] = value, diff.size
                row = cr[(cr.case_bed == case_bed) & (cr.control_bed == control_bed)]
                require(len(row) == 1, f"Nonunique pair cell: {scenario}/{key}/{case_bed}/{control_bed}")
                if len(row) == 1:
                    near(row.iloc[0].auc, value, f"Saved pair-cell AUC: {scenario}/{key}")
                    require(row.iloc[0].pairs == diff.size, f"Saved pair count: {scenario}/{key}")
        weighted = np.sum(mat * weights) / weights.sum()
        max_error = max(max_error, near(mr.auc, weighted, f"Weighted identity: {scenario}/{key}"))
        diag = np.trace(mat) / 3
        values = {"within_bed_auc": diag, "delta_mix": weighted - diag,
                  "mix_min": mat.min(), "mix_max": mat.max(), "mix_span": mat.max() - mat.min(),
                  "cross_bed_pair_fraction": 1 - np.trace(weights) / weights.sum()}
        for name, value in values.items():
            max_error = max(max_error, near(mr[name], value, f"{scenario}/{key}/{name}"))
    fold_report[scenario] = {"fold_model_records": len(metrics), "complete_cell_records": n_complete,
                            "missing_cell_records": n_missing, "maximum_absolute_error": max_error}
    if scenario == "primary":
        primary = pred
        require(len(metrics) == 30 * 7 and n_complete == 30 * 7 and n_missing == 0,
                "Primary does not contain all30folds*7models with complete cells")
        require(pred.groupby(["model", "repeat"]).size().eq(95).all(), "Primary repeat not95 rows")
        require(pred.groupby(["model", "repeat"])["sample"].nunique().eq(95).all(), "Primary test-fold overlap")
        require(pred.groupby(["repeat", "sample"])["fold"].nunique().eq(1).all(), "Models use different test partitions")
        for (_, _), block in pred.groupby(["model", "repeat"]):
            require(set(block["sample"]) == set(units.label_group), "Primary partition does not cover exact labels")
        baseline = metrics[metrics.model == "prevalence"]
        require(np.allclose(baseline.auc, .5, atol=TOL) and np.allclose(baseline.delta_mix, 0, atol=TOL),
                "Intercept negative baseline fails chance-ordering check")
    if scenario == "104_grouped_arrays":
        merged = pred.merge(source_meta[["sample", "label_group"]], on="sample", validate="many_to_one")
        require(merged.groupby(["model", "repeat", "label_group"]).fold.nunique().eq(1).all(),
                "A repeated label crosses folds in104-array sensitivity")
checks["independent_fold_recomputation"] = fold_report
checks["primary_partition_scope"] = "Test folds are disjoint and cover95labels per repetition. Actual training arrays are not independently logged; code defines training as the complement. This checks partitions, not hidden fitting state."

ext = read("external_predictions.csv")
ext_metrics = read("external_metrics.csv")
ext_pairs = read("external_pairs.csv")
external_keys = {(dataset, model) for dataset in ["GSE43292", "GSE28829"] for model in EXPRESSION_MODELS}
require(set(ext[["dataset", "model"]].itertuples(index=False, name=None)) == external_keys,
        "External prediction inventory not exact five-model by two-cohort grid")
require(set(ext_metrics[["dataset", "model"]].itertuples(index=False, name=None)) == external_keys and len(ext_metrics) == 10,
        "External metric inventory not exact five-model by two-cohort grid")
require(len(ext_pairs) == 160 and set(ext_pairs.model) == EXPRESSION_MODELS, "External pair inventory not five models by32patients")
ext_report = {}
for (dataset, model), block in ext.groupby(["dataset", "model"]):
    row = ext_metrics[(ext_metrics.dataset == dataset) & (ext_metrics.model == model)]
    require(len(row) == 1, "Nonunique external metric row")
    if len(row) != 1:
        continue
    row = row.iloc[0]
    require(len(block) == (64 if dataset == "GSE43292" else 29), "External denominator changed")
    delta = 0.
    for name, value in score_metrics(block.y, block.score, model not in SCORE_MODELS).items():
        delta = max(delta, near(row[name], value, f"External {dataset}/{model}/{name}"))
    if dataset == "GSE43292":
        require(block.patient.nunique() == 32, "External paired patient count not32")
        require(block.groupby("patient").size().eq(2).all(), "Patient block does not have2tissues")
        require(block.groupby("patient").y.sum().eq(1).all(), "Patient block lacks one tissue of each class")
        differences = []
        for patient, pair in block.groupby("patient"):
            plaque = pair.loc[pair.y == 1, "score"].iloc[0]
            intact = pair.loc[pair.y == 0, "score"].iloc[0]
            differences.append(plaque - intact)
            pr = ext_pairs[(ext_pairs.model == model) & (ext_pairs.patient.astype(str) == str(patient))]
            require(len(pr) == 1, "Saved external pair missing")
            if len(pr) == 1:
                near(pr.iloc[0].difference, plaque - intact, "Paired difference")
        differences = np.array(differences)
        near(row.pair_concordance, ((differences > 0) + .5 * (differences == 0)).mean(), "Pair concordance")
        near(row.paired_mean_difference, differences.mean(), "Mean paired difference")
    ext_report[dataset + "/" + model] = {"n": len(block), "maximum_metric_error": delta}
checks["external_recomputed_metrics"] = ext_report

coef = read("frozen_primary_coefficients.csv")
frozen_path = OUT / "frozen_primary_model.json"
snapshots[frozen_path.name] = hashlib.sha256(frozen_path.read_bytes()).hexdigest()
frozen = json.loads(frozen_path.read_text())
require(len(coef) == 200 and coef.gene.nunique() == 200, "Frozen coefficient feature count")
reconstruction = {}
for dataset in ["GSE43292", "GSE28829"]:
    matrix = pd.read_pickle(ROOT / "data" / (dataset + "_rank.pkl"))
    scaled = (matrix[coef.gene].to_numpy() - coef.source_mean.to_numpy()) / coef.source_sd.to_numpy()
    probability = expit(scaled @ coef.coefficient.to_numpy() + frozen["intercept"])
    expected = pd.Series(probability, index=matrix.index)
    block = ext[(ext.dataset == dataset) & (ext.model == "ridge200")]
    discrepancy = np.max(np.abs(block.score.to_numpy() - expected.loc[block["sample"]].to_numpy()))
    require(discrepancy < TOL, "Frozen coefficient reconstruction fails for " + dataset)
    reconstruction[dataset] = {"n": len(block), "maximum_absolute_score_error": float(discrepancy)}
checks["frozen_primary_external_reconstruction"] = reconstruction

transport = read("transport_metrics.csv")
transport_pred = read("transport_predictions.csv")
keycols = ["source", "target", "purge_possible_controls", "model"]
expected_transport = set()
for target in BEDS:
    for source in ["other_two"] + [bed for bed in BEDS if bed != target]:
        for purge in [False, True]:
            for model in (EXPRESSION_MODELS if source == "other_two" and not purge else {"ridge200"}):
                expected_transport.add((source, target, purge, model))
require(len(transport) == 30 and set(transport[keycols].itertuples(index=False, name=None)) == expected_transport,
        "Transport metric inventory mismatch")
require(set(transport_pred[keycols].itertuples(index=False, name=None)) == expected_transport, "Transport prediction inventory mismatch")
transport_error = 0.
for key, block in transport_pred.groupby(keycols):
    selected = np.ones(len(transport), dtype=bool)
    for name, value in zip(keycols, key): selected &= transport[name].eq(value).to_numpy()
    rows = transport[selected]
    require(len(rows) == 1, "Nonunique transport row")
    if len(rows) != 1: continue
    row = rows.iloc[0]
    require(len(block) == int(row.n_test) and block['sample'].nunique() == len(block), "Transport target count/duplicate mismatch")
    for name, value in score_metrics(block.y, block.score, key[-1] not in SCORE_MODELS).items():
        transport_error = max(transport_error, near(row[name], value, f"Transport {key}/{name}"))
checks["transport_saved_metric_recomputation"] = {"rows": len(transport), "maximum_absolute_error": transport_error}

aggregate = read("sensitivity_all_fold_metrics.csv")
require(len(aggregate) == 240 and set(aggregate.scenario) == SCENARIOS - {"primary"}, "Sensitivity aggregate inventory mismatch")
for scenario in sorted(SCENARIOS - {"primary"}):
    original = read(scenario + "_fold_metrics.csv").sort_values(["model", "repeat", "fold"]).reset_index(drop=True)
    saved = aggregate[aggregate.scenario == scenario].sort_values(["model", "repeat", "fold"]).reset_index(drop=True)
    try: pd.testing.assert_frame_equal(saved[original.columns], original, check_dtype=False, atol=TOL, rtol=0)
    except AssertionError as exc: require(False, f"Sensitivity aggregate mismatch {scenario}: {exc}")
checks["sensitivity_aggregate"] = {"rows": len(aggregate), "scenarios": 8, "matches_individual_tables": True}

descriptive = read("descriptive_mean_crossfit_scores.csv")
descriptive_metrics = read("descriptive_mean_crossfit_metrics.csv")
require(len(descriptive) == 665 and set(descriptive.model) == MODELS and not descriptive.duplicated(["model", "label_group"]).any(),
        "Descriptive mean-score inventory mismatch")
require(len(descriptive_metrics) == 7 and set(descriptive_metrics.model) == MODELS, "Descriptive metric inventory mismatch")
for model in sorted(MODELS):
    mean_scores = primary[primary.model == model].groupby("sample").score.mean()
    saved = descriptive[descriptive.model == model].set_index("label_group")
    require(set(saved.index) == set(units.label_group) and saved.n_held_out_scores.eq(10).all(), "Descriptive labels/count mismatch")
    if set(saved.index) == set(mean_scores.index):
        require(np.allclose(saved.mean_crossfit_score, mean_scores.loc[saved.index], atol=TOL, rtol=0), "Descriptive mean-score mismatch")
        metric = descriptive_metrics[descriptive_metrics.model == model]
        if len(metric) == 1:
            near(metric.iloc[0].descriptive_mean_crossfit_pooled_auc,
                 roc_auc_score(saved.y, saved.mean_crossfit_score), "Descriptive pooled AUROC mismatch")
checks["descriptive_mean_crossfit"] = {"rows": len(descriptive), "models": len(descriptive_metrics), "scope": "Saved-score aggregation, separate from within-model primary estimand"}

subsets = read("full_pipeline_subsampling.csv")
require(len(subsets) == 30 and set(subsets.iteration) == set(range(30)), "Subsampling iteration inventory mismatch")
require(subsets.n.eq(75).all() and np.isfinite(subsets.auc).all() and subsets.auc.between(0,1).all(), "Invalid subset sizes/AUROCs")
checks["subsampling_output_structure"] = {"iterations": len(subsets), "actual_units_per_subset": 75, "scope": "Saved-output structure only; fitting not rerun"}

nulls = read("permutation_nulls.csv")
require(nulls.groupby("scheme").size().to_dict() == {"unrestricted": 99, "within_bed": 99}, "Permutation counts")
require(np.isfinite(nulls.mean_fold_auc).all() and nulls.mean_fold_auc.between(0, 1).all(), "Invalid null output")
checks["null_output_structure"] = {"within_bed": 99, "unrestricted": 99,
    "scope": "Counts/ranges only. Permutations not rerun; no requirement that every null AUC equals0.5."}
report = {"created_utc": datetime.datetime.now(datetime.timezone.utc).isoformat(),
          "tolerance": TOL, "passed": len(errors) == 0, "errors": errors, "checks": checks,
          "result_file_sha256": snapshots,
          "scope": "Read-only recomputation from saved scores, metadata and coefficients; no training, cohort reacquisition or donor authentication."}
path = ROOT / "evidence" / "independent_saved_results_validation.json"
path.write_text(json.dumps(report, indent=2), encoding="utf-8")
print(json.dumps({"passed": report["passed"], "errors": errors, "report": str(path),
                  "frozen_reconstruction": reconstruction}, indent=2))
raise SystemExit(0 if report["passed"] else 1)
```
