## Supplementary material for "Anatomical comparison populations and threshold transport in vascular transcriptomic classification": Reproducibility archive: code, provenance and results: recommended_team_references.bib

@article{flow,
  author = {You, Lingsen and Chen, Yuheng and Zhang, Zeyang and Wang, Yu and Gu, Zhongze and Shen, Li and Ge, Junbo},
  title = {{The FLOW Framework: a panvascular-on-a-chip platform to model systemic disease and guide panvascular interventional device suitcordance}},
  journal = {Science Bulletin},
  year = {2026},
  doi = {10.1016/j.scib.2025.12.051},
  url = {https://doi.org/10.1016/j.scib.2025.12.051},
  volume = {71},
  pages = {654--670}
}

@article{tri_ecological,
  author = {You, Lingsen and Li, Yaxuan and Luo, Yuchen and Cheng, Qiang and Feng, Liuliu and Shen, Li and Ge, Junbo},
  title = {{Vascular-device Suitcordance: a tri-ecological framework for eco-rebalancing}},
  journal = {Trends in Molecular Medicine},
  year = {2026},
  doi = {10.1016/j.molmed.2026.09.007},
  url = {https://doi.org/10.1016/j.molmed.2026.09.007}
}

@article{photonics,
  author = {You, Lingsen and Yao, Jiaxin and Qiu, Yaoqing and Wang, Yu and Sun, Yunlu and Zhang, Rongjun and Shen, Li and Ge, Junbo},
  title = {{From intravascular imaging to adaptive vascular care: intelligent photonics and digital twins in panvascular disease}},
  journal = {Light: Science \& Applications},
  year = {2026},
  doi = {10.1038/s41377-026-02410-6},
  url = {https://doi.org/10.1038/s41377-026-02410-6},
  volume = {15},
  pages = {335}
}

@article{full_watershed,
  author = {You, Lingsen and Chen, Yuheng and Zhang, Zeyang and Wang, Yu and Shen, Li and Ge, Junbo},
  title = {{High Suitcordance for Panvascular Full-Watershed Organs: A New Interventional Perspective}},
  journal = {Research},
  year = {2025},
  doi = {10.34133/research.0974},
  url = {https://doi.org/10.34133/research.0974},
  volume = {8},
  pages = {0974}
}

@misc{cui2026iwm,
  author = {Cui, Taoyong and Wang, Xi and Li, Zonghang and Ding, Jinchao and You, Lingsen and Xu, Yuzhi and Xu, Wanghan and Wu, Fang and Ying, Kejun and Ouyang, Wanli and Heng, Pheng Ann and Yang, Ling and Yin, Zhenfei and Wu, Yingcheng},
  title = {{An immune world model for multiscale forecasting and therapeutic hypothesis generation}},
  year = {2026},
  eprint = {2609.14709},
  archivePrefix = {arXiv},
  primaryClass = {cs.LG},
  note = {Preprint, version 1, submitted 13 September 2026},
  doi = {10.48550/arXiv.2609.14709},
  url = {https://arxiv.org/abs/2609.14709v1}
}
