## Supplementary material for "Anatomical comparison populations and threshold transport in vascular transcriptomic classification": Reproducibility archive: code, provenance and results: independent_references_verified.bib

@article{steenman2018,
  author = {Steenman, Marja and Espitia, Olivier and Maurel, Blandine and Guyomarch, Beatrice and Heymann, Marie-Françoise and Pistorius, Marc-Antoine and Ory, Benjamin and Heymann, Dominique and Houlgatte, Rémi and Gouëffic, Yann and Quillard, Thibaut},
  title = {{Identification of genomic differences among peripheral arterial beds in atherosclerotic and healthy arteries}},
  journal = {Scientific Reports},
  year = {2018},
  volume = {8},
  number = {1},
  pages = {3940},
  doi = {10.1038/s41598-018-22292-y},
  url = {https://doi.org/10.1038/s41598-018-22292-y},
}

@article{ayari2013,
  author = {Ayari, Hanène and Bricca, Giampiero},
  title = {{Identification of two genes potentially associated in iron-heme homeostasis in human carotid plaque using microarray analysis}},
  journal = {Journal of Biosciences},
  year = {2013},
  volume = {38},
  number = {2},
  pages = {311--315},
  doi = {10.1007/s12038-013-9310-2},
  url = {https://doi.org/10.1007/s12038-013-9310-2},
}

@article{doring2012,
  author = {Döring, Yvonne and Manthey, Helga D and Drechsler, Maik and Lievens, Dirk and Megens, Remco T A and Soehnlein, Oliver and Busch, Martin and Manca, Marco and Koenen, Rory R and Pelisek, Jaroslav and Daemen, Mat J and Lutgens, Esther and Zenke, Martin and Binder, Christoph J and Weber, Christian and Zernecke, Alma},
  title = {{Auto-Antigenic Protein-DNA Complexes Stimulate Plasmacytoid Dendritic Cells to Promote Atherosclerosis}},
  journal = {Circulation},
  year = {2012},
  volume = {125},
  number = {13},
  pages = {1673--1683},
  doi = {10.1161/circulationaha.111.046755},
  url = {https://doi.org/10.1161/circulationaha.111.046755},
}

@article{wu2024,
  author = {Wu, Xize and Pan, Xue and Zhou, Yi and Pan, Jiaxiang and Kang, Jian and Yu, J J Jiajia and Cao, Yingyue and Quan, Chao and Gong, Lihong and Li, Yue},
  title = {{Identification of key genes for atherosclerosis in different arterial beds}},
  journal = {Scientific Reports},
  year = {2024},
  volume = {14},
  number = {1},
  pages = {6543},
  doi = {10.1038/s41598-024-55575-8},
  url = {https://doi.org/10.1038/s41598-024-55575-8},
}

@article{geo,
  author = {Edgar, Ron and Domrachev, Michael and Lash, Alex E},
  title = {{Gene Expression Omnibus: NCBI gene expression and hybridization array data repository}},
  journal = {Nucleic Acids Research},
  year = {2002},
  volume = {30},
  number = {1},
  pages = {207--210},
  doi = {10.1093/nar/30.1.207},
  url = {https://doi.org/10.1093/nar/30.1.207},
}

@article{hanley1982,
  author = {Hanley, J A and McNeil, B J},
  title = {{The meaning and use of the area under a receiver operating characteristic (ROC) curve.}},
  journal = {Radiology},
  year = {1982},
  volume = {143},
  number = {1},
  pages = {29--36},
  doi = {10.1148/radiology.143.1.7063747},
  url = {https://doi.org/10.1148/radiology.143.1.7063747},
}

@article{delong1988,
  author = {DeLong, E R and DeLong, D M and Clarke-Pearson, D L},
  title = {{Comparing the Areas under Two or More Correlated Receiver Operating Characteristic Curves: A Nonparametric Approach}},
  journal = {Biometrics},
  year = {1988},
  volume = {44},
  number = {3},
  pages = {837},
  doi = {10.2307/2531595},
  url = {https://doi.org/10.2307/2531595},
}

@article{obuchowski1997,
  author = {Obuchowski, N A},
  title = {{Nonparametric Analysis of Clustered ROC Curve Data}},
  journal = {Biometrics},
  year = {1997},
  volume = {53},
  number = {2},
  pages = {567--578},
  doi = {10.2307/2533958},
  url = {https://doi.org/10.2307/2533958},
}

@article{janes2008,
  author = {Janes, Holly and Pepe, Margaret S},
  title = {{Matching in Studies of Classification Accuracy: Implications for Analysis, Efficiency, and Assessment of Incremental Value}},
  journal = {Biometrics},
  year = {2008},
  volume = {64},
  number = {1},
  pages = {1--9},
  doi = {10.1111/j.1541-0420.2007.00823.x},
  url = {https://doi.org/10.1111/j.1541-0420.2007.00823.x},
}

@article{ambroise2002,
  author = {Ambroise, Christophe and McLachlan, Geoffrey J},
  title = {{Selection bias in gene extraction on the basis of microarray gene-expression data}},
  journal = {Proceedings of the National Academy of Sciences},
  year = {2002},
  volume = {99},
  number = {10},
  pages = {6562--6566},
  doi = {10.1073/pnas.102102699},
  url = {https://doi.org/10.1073/pnas.102102699},
}

@article{varma2006,
  author = {Varma, Sudhir and Simon, Richard},
  title = {{Bias in error estimation when using cross-validation for model selection}},
  journal = {BMC Bioinformatics},
  year = {2006},
  volume = {7},
  number = {1},
  pages = {91},
  doi = {10.1186/1471-2105-7-91},
  url = {https://doi.org/10.1186/1471-2105-7-91},
}

@article{roberts2017,
  author = {Roberts, David R. and Bahn, Volker and Ciuti, Simone and Boyce, Mark S. and Elith, Jane and Guillera‐Arroita, Gurutzeta and Hauenstein, Severin and Lahoz‐Monfort, José J. and Schröder, Boris and Thuiller, Wilfried and Warton, David I. and Wintle, Brendan A. and Hartig, Florian and Dormann, Carsten F.},
  title = {{Cross‐validation strategies for data with temporal, spatial, hierarchical, or phylogenetic structure}},
  journal = {Ecography},
  year = {2017},
  volume = {40},
  number = {8},
  pages = {913--929},
  doi = {10.1111/ecog.02881},
  url = {https://doi.org/10.1111/ecog.02881},
}

@article{soneson2014,
  author = {Soneson, Charlotte and Gerster, Sarah and Delorenzi, Mauro},
  title = {{Batch Effect Confounding Leads to Strong Bias in Performance Estimates Obtained by Cross-Validation}},
  journal = {PLoS ONE},
  year = {2014},
  volume = {9},
  number = {6},
  pages = {e100335},
  doi = {10.1371/journal.pone.0100335},
  url = {https://doi.org/10.1371/journal.pone.0100335},
}

@article{tripod2024,
  author = {Collins, Gary S and Moons, Karel G M and Dhiman, Paula and Riley, Richard D and Beam, Andrew L and Van Calster, Ben and Ghassemi, Marzyeh and Liu, Xiaoxuan and Reitsma, Johannes B and van Smeden, Maarten and Boulesteix, Anne-Laure and Camaradou, Jennifer Catherine and Celi, Leo Anthony and Denaxas, Spiros and Denniston, Alastair K and Glocker, Ben and Golub, Robert M and Harvey, Hugh and Heinze, Georg and Hoffman, Michael M and Kengne, André Pascal and Lam, Emily and Lee, Naomi and Loder, Elizabeth W and Maier-Hein, Lena and Mateen, Bilal A and McCradden, Melissa D and Oakden-Rayner, Lauren and Ordish, Johan and Parnell, Richard and Rose, Sherri and Singh, Karandeep and Wynants, Laure and Logullo, Patricia},
  title = {{TRIPOD+AI statement: updated guidance for reporting clinical prediction models that use regression or machine learning methods}},
  journal = {BMJ},
  year = {2024},
  volume = {385},
  pages = {e078378},
  doi = {10.1136/bmj-2023-078378},
  url = {https://doi.org/10.1136/bmj-2023-078378},
}

@article{cui2023,
  author = {Cui, Hong-Kai and Tang, Chao-Jie and Gao, Yu and Li, Zi-Ang and Zhang, Jian and Li, Yong-Dong},
  title = {{An integrative analysis of single-cell and bulk transcriptome and bidirectional mendelian randomization analysis identified C1Q as a novel stimulated risk gene for Atherosclerosis}},
  journal = {Frontiers in Immunology},
  year = {2023},
  volume = {14},
  pages = {1289223},
  doi = {10.3389/fimmu.2023.1289223},
  url = {https://doi.org/10.3389/fimmu.2023.1289223},
}

@article{zhong2025,
  author = {Zhong, Yiming and Chen, Yifan and Wang, Yuehong and Pan, Wenhai and Huang, Honghao and Cao, Lin Lin and Wang, Yili and Zhang, Jiawen and Liang, Yunyi and Qian, Xiangyue and Shi, Chenxi and Lu, Xiyuan and Pu, Jun},
  title = {{Deciphering the panvascular immune landscape of atherosclerosis}},
  journal = {VIEW},
  year = {2025},
  volume = {6},
  number = {4},
  pages = {20250041},
  doi = {10.1002/viw.20250041},
  url = {https://doi.org/10.1002/viw.20250041},
}

@article{venet2011,
  author = {Venet, David and Dumont, Jacques E. and Detours, Vincent},
  title = {{Most Random Gene Expression Signatures Are Significantly Associated with Breast Cancer Outcome}},
  journal = {PLoS Computational Biology},
  year = {2011},
  volume = {7},
  number = {10},
  pages = {e1002240},
  doi = {10.1371/journal.pcbi.1002240},
  url = {https://doi.org/10.1371/journal.pcbi.1002240}
}
