## Supplementary material for "Anatomical comparison populations and threshold transport in vascular transcriptomic classification": Reproducibility archive: code, provenance and results: Figure_4.pdf

### Paired external tissue discrimination and threshold failure

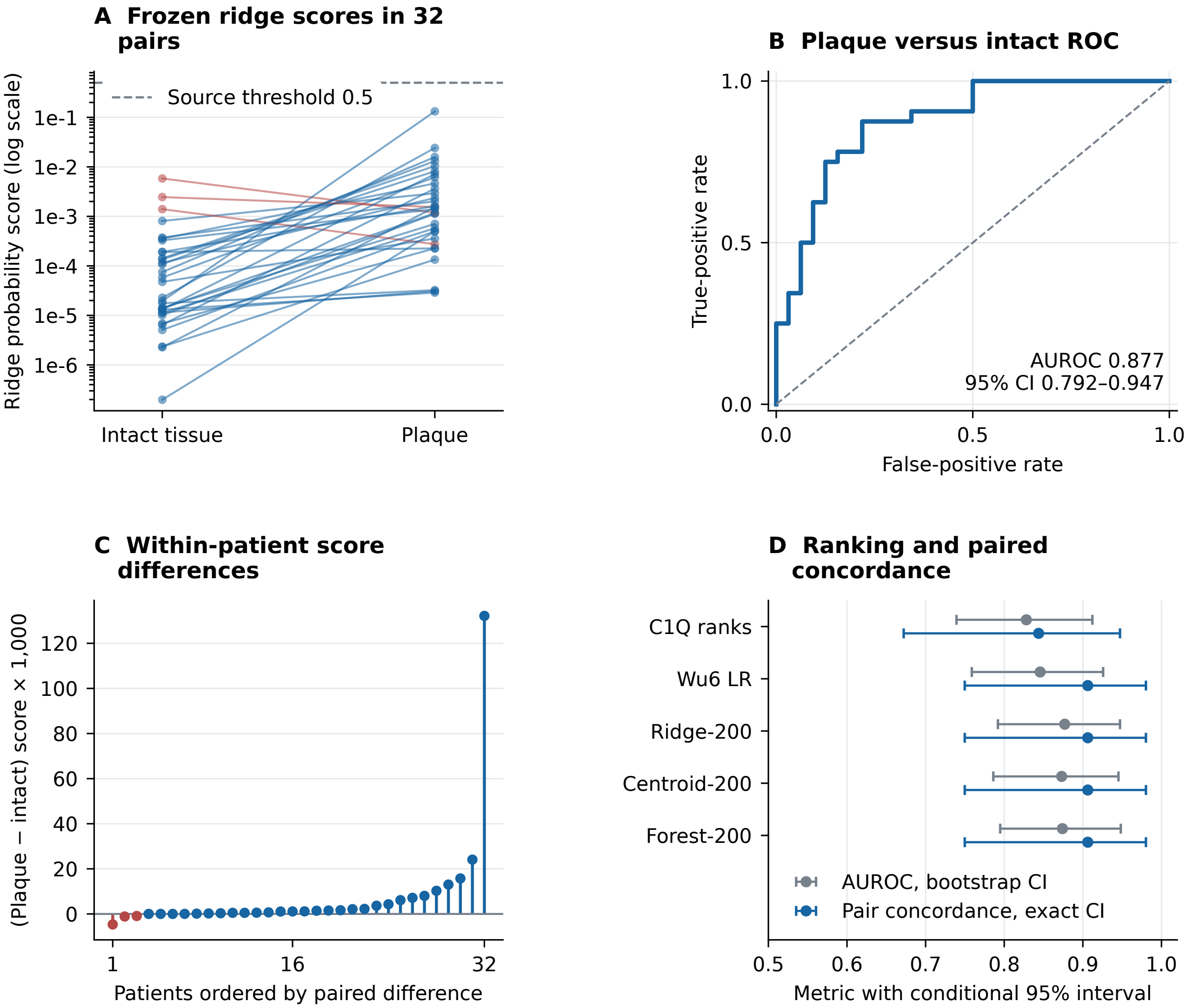


#### E Probability-model diagnostics at the unchanged 0.5 threshold

| Model | Sens. | Spec. | Bal. acc. | Brier |
|---|---|---|---|---|
| Wu6 LR | 0.938 | 0.531 | 0.734 | 0.180 |
| Ridge-200 | 0.000 | 1.000 | 0.500 | 0.493 |
| Forest-200 | 0.125 | 1.000 | 0.562 | 0.219 |
