## Supplementary material for "Anatomical comparison populations and threshold transport in vascular transcriptomic classification": Reproducibility archive: code, provenance and results: Figure_5.pdf

### External histology contrast: advanced versus early lesions

##### A Primary-model histology ROC

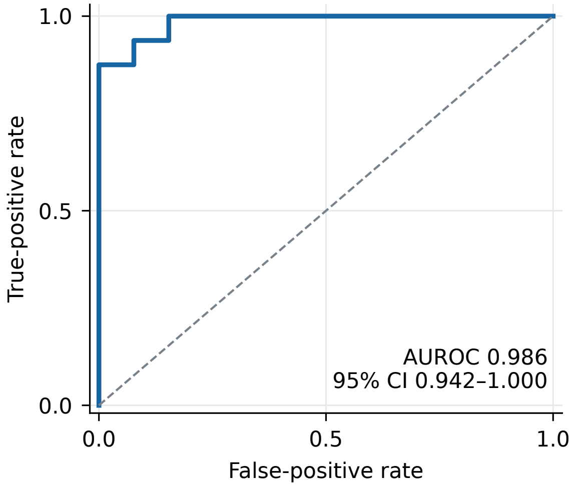


##### B Probability scale after transport

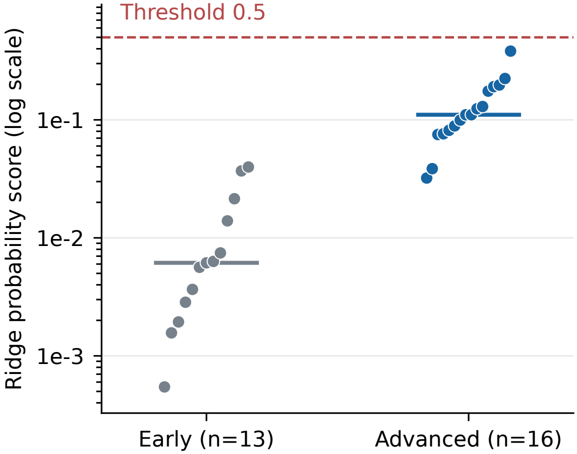


##### C Frozen-model ranking comparison

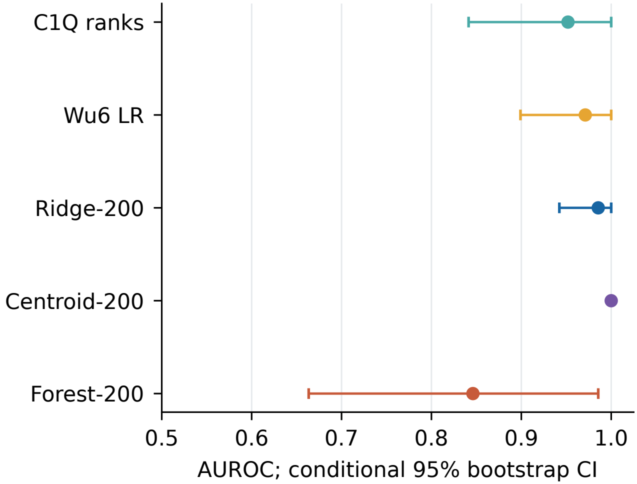


##### D Unchanged threshold diagnostics

| Model | Sens. | Spec. | Bal. acc. | Brier |
|---|---|---|---|---|
| Wu6 LR | 0.000 | 1.000 | 0.500 | 0.258 |
| Ridge-200 | 0.000 | 1.000 | 0.500 | 0.418 |
| Forest-200 | 0.000 | 1.000 | 0.500 | 0.256 |
