## Supplementary material for "Anatomical comparison populations and threshold transport in vascular transcriptomic classification": Reproducibility archive: code, provenance and results: Figure_10.pdf

### Tested source thresholds leave external sensitivity at zero

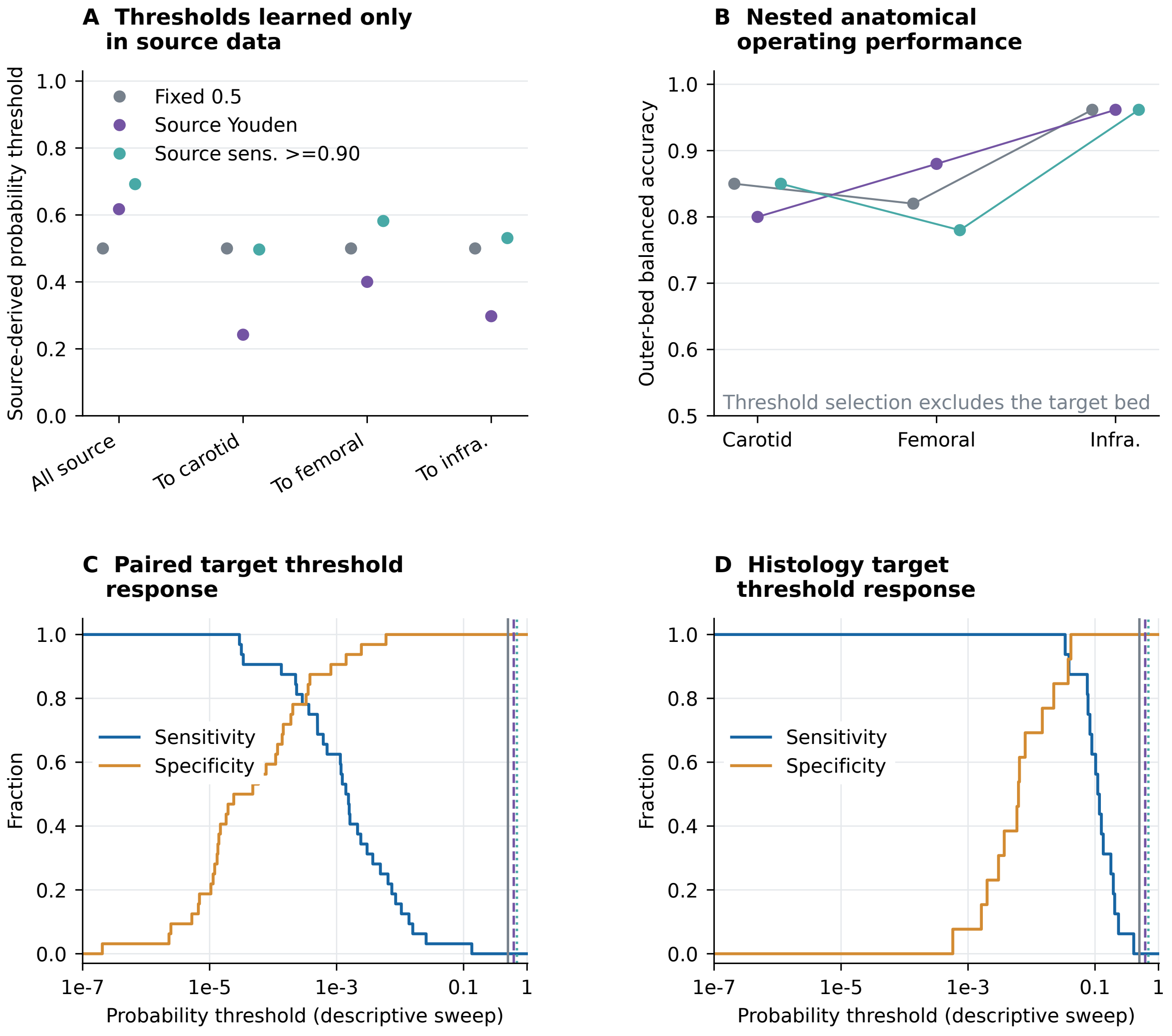


**E Frozen external confusion counts for all three source rules**

| Target | Source rule | Cutoff | TP / FN | TN / FP | Sens. | Spec. | Bal. acc. |
|---|---|---|---|---|---|---|---|
| Paired tissues | Fixed 0.5 | 0.500 | 0/32 | 32/0 | 0.00 | 1.00 | 0.50 |
| Paired tissues | Source Youden | 0.617 | 0/32 | 32/0 | 0.00 | 1.00 | 0.50 |
| Paired tissues | Source sens. >=0.90 | 0.692 | 0/32 | 32/0 | 0.00 | 1.00 | 0.50 |
| Histology | Fixed 0.5 | 0.500 | 0/16 | 13/0 | 0.00 | 1.00 | 0.50 |
| Histology | Source Youden | 0.617 | 0/16 | 13/0 | 0.00 | 1.00 | 0.50 |
| Histology | Source sens. >=0.90 | 0.692 | 0/16 | 13/0 | 0.00 | 1.00 | 0.50 |
