## Supplementary material for "Anatomical comparison populations and threshold transport in vascular transcriptomic classification": Reproducibility archive: code, provenance and results: declarations.tex

\section*{Data and code availability}
Official processed data are available from GEO accessions GSE100927, GSE43292, and GSE28829, with platforms GPL17077, GPL6244, and GPL570. The accompanying reproducibility archive contains acquisition code, source hashes, annotation mappings, the internal protocol and amendments, sample-label audits, analysis code, frozen coefficients, predictions, figures, and machine-readable result tables. Inputs can be reacquired from the recorded official URLs. The archive does not confer rights beyond those applying to the original data. A separate public archival identifier for the reproducibility archive has not yet been assigned.

\section*{Ethics, consent, and study registration}
This work analyzes existing public tissue-expression records and does not recruit participants, intervene, or access direct personal identifiers. Original cohort ethics and consent are described in the source publications. The authors confirm that applicable ethical and consent requirements for this secondary analysis have been met. No new institutional approval or exemption is claimed. This is not a clinical trial, and no trial registration is claimed. The internal analysis plan was not a prospective registry entry.

\section*{Author contributions}
Lingsen You, Zisu Peng, and Yujun Guo contributed equally to this work. All authors reviewed and approved the manuscript.

\section*{Funding}
This work was supported by the National Natural Science Foundation of China (T2288101 and 82170342), the Shanghai Advanced Materials Key Technology Project Fund (25CL2900500), the Medical Engineering Joint Fund of Fudan University (yg2023-01), and the AI for Science Foundation of Fudan University (FudanX24AI003).

\section*{Competing interests}
The authors declare no competing interests.
