## Supplementary material for "Anatomical comparison populations and threshold transport in vascular transcriptomic classification": Reproducibility archive: code, provenance and results: supplement.tex

\documentclass[11pt,a4paper]{article}
\usepackage[margin=20mm]{geometry}
\usepackage[T1]{fontenc}
\usepackage[utf8]{inputenc}
\usepackage{lmodern,amsmath,amssymb,graphicx,booktabs,longtable,array,xcolor,pdflscape}
\usepackage{microtype,setspace,caption,float}
\usepackage[hidelinks]{hyperref}
\usepackage[numbers,sort&compress]{natbib}
\setstretch{1.12}
\setlength{\parindent}{0pt}
\setlength{\parskip}{5pt}
\captionsetup{font=small,labelfont=bf}
\hypersetup{pdftitle={Supplementary material: Anatomical comparison populations and threshold transport in vascular transcriptomic classification},pdfauthor={Lingsen You, Zisu Peng, Yujun Guo, Li Shen, Junbo Ge},pdfsubject={medRxiv submission manuscript}}
\begin{document}
{\Large\bfseries Supplementary material\par}
Anatomical comparison populations and threshold transport in vascular transcriptomic classification\par\vspace{3mm}
Computational tables and provenance. No independent-patient total is constructed across cohorts.\par
\input{supplement_body.tex}
\clearpage\input{exploratory_supplement.tex}
\clearpage\renewcommand{\thefigure}{S\arabic{figure}}\setcounter{figure}{0}\newgeometry{left=20mm,right=20mm,top=12mm,bottom=14mm}
\clearpage\begin{figure}[p]\centering\setstretch{1}
\includegraphics[width=17cm]{../figures/Figure_S1.pdf}
\caption{\textbf{Fold-level resolution of anatomical comparisons}. A--I, All 30 held-out folds for each primary-model case-bed/control-bed comparison. Horizontal jitter separates the three folds of each repetition. Blue denotes diagonal within-bed comparisons; orange denotes cross-bed comparisons. The AUROC axis is deliberately truncated at 0.85 to display the smallest observed cell (0.875). Labels give the actual pair-count range in each cell; pairs share observations, and folds/repetitions share source data. Points are not independent studies, and no confidence interval is implied. These original saved results complement the mean matrix in Figure 2; no new model was fitted for this display.}\label{fig:s1}
\end{figure}\clearpage
\clearpage\begin{figure}[p]\centering\setstretch{1}
\includegraphics[width=17cm]{../figures/Figure_S2.pdf}
\caption{\textbf{Repeated source scores expose threshold discrepancies}. A, Each point is one group’s mean of ten original held-out primary scores, with its minimum-to-maximum range. Groups are ordered within bed/status by mean score; this ordering is descriptive. Red outlines identify groups misclassified in every repetition at 0.5. The display-position-to-label mapping is supplied in results/exploratory\_display/source\_score\_stability.csv. B, Counts of groups by their number of threshold errors. Source groups are unverified donor units, repeated fits are dependent, and ranges are not confidence intervals. These pooled score summaries do not replace the original within-fold AUROC estimand. No new fitting or threshold selection was performed for this display.}\label{fig:s2}
\end{figure}\clearpage
\restoregeometry\end{document}
