## Supplementary material for "Anatomical comparison populations and threshold transport in vascular transcriptomic classification": Reproducibility archive: code, provenance and results: titlepage.tex

\begin{titlepage}\vspace*{2mm}{\Large\bfseries\raggedright\hyphenpenalty=10000 Anatomical comparison populations and threshold transport in vascular transcriptomic classification\par}\vspace{5mm}
Lingsen You\textsuperscript{a,b,c,d,e,$\dagger$,$*$}, Zisu Peng\textsuperscript{a,b,d,e,f,g,$\dagger$}, Yujun Guo\textsuperscript{h,$\dagger$}, Li Shen\textsuperscript{a,b,c,d,e}, Junbo Ge\textsuperscript{a,b,c,d,e,$*$}\par\vspace{3mm}
{\small
\textsuperscript{a}Department of Cardiology, Zhongshan Hospital, Fudan University, Shanghai Institute of Cardiovascular Diseases, Shanghai 200032, China.\par
\textsuperscript{b}National Clinical Research Center for Interventional Medicine, Shanghai 200032, China.\par
\textsuperscript{c}Oriental Pan-Vascular Devices Innovation College, University of Shanghai for Science and Technology, Shanghai 200093, China.\par
\textsuperscript{d}State Key Laboratory of Cardiovascular Diseases, Zhongshan Hospital, Fudan University, Shanghai 200032, China.\par
\textsuperscript{e}NHC Key Laboratory of Ischemic Heart Diseases, Shanghai 200032, China.\par
\textsuperscript{f}Key Laboratory of Viral Heart Diseases, Chinese Academy of Medical Sciences, Shanghai 200032, China.\par
\textsuperscript{g}Institutes of Biomedical Sciences, Fudan University, Shanghai 200030, China.\par
\textsuperscript{h}College of Biomedical Engineering, Fudan University, Shanghai 200438, China.\par
}
\vspace{4mm}\textsuperscript{$\dagger$}Lingsen You, Zisu Peng, and Yujun Guo contributed equally to this work.\par
\vspace{3mm}\begingroup\small\setstretch{1}
\noindent\textbf{Abstract}\par
\textbf{Background:} High vascular transcriptomic discrimination may obscure dependence on comparison populations, gene selection, and score scale. We examined these properties separately.

\textbf{Methods:} GSE100927 compared surgical atherosclerotic tissue with lesion-free deceased-donor arteries: 104 arrays formed 95 complete-label groups, versus 96 arteries reported originally; independent donor identities remained unresolved. Within held-out folds, pooled area under the receiver-operating-characteristic curve (AUROC) was decomposed into nine case-bed--control-bed comparisons. Seven fixed rules underwent ten repeated three-fold splits. The primary ridge model used sample-wise gene ranks and training-only selection of 200 genes. External tests involved 32 paired plaque/intact patients in GSE43292 and 29 early/advanced specimens in GSE28829. Post-result explorations added random and disjoint gene-panel controls, feature stability, frozen-score accounting, source-only threshold learning, and individual-failure analyses.

\textbf{Results:} Primary mean held-out-fold pooled AUROC was 0.9968, versus a within-bed mean of 1.0000 and contrast $-0.0032$; the empirical ceiling constrained this contrast to be nonpositive. Paired external AUROC was 0.877 (conditional 95\% interval 0.792--0.947), with plaque ranked higher in 29/32 patients. Histological AUROC was 0.986 (0.942--1.000). All primary external probabilities fell below 0.5. Both source-derived thresholds also gave zero external sensitivity. Twenty outcome-independent random panels retained source AUROCs of 0.9932--0.9992, while external ranking varied substantially. Across 30 primary fits, 619 genes were selected at least once and 45 every time. Fixed-score failure analyses distinguished shared from model-specific discordance.

\textbf{Conclusions:} Strong source ranking did not establish a unique gene signature or transferable technical thresholds. Procurement confounding, uncertain donor identities, inherited preprocessing, changed endpoints, and the exploratory status of added analyses limit interpretation. This is a comparison-population audit, not a clinically calibrated risk model.

\noindent\textbf{Keywords:} atherosclerosis; arterial bed; transcriptomics; transportability; paired tissues; classification.

\endgroup
\end{titlepage}
