## Supplementary material for "Anatomical comparison populations and threshold transport in vascular transcriptomic classification": Reproducibility archive: code, provenance and results: amendment_01_before_fit.txt

2026-10-25, after outcome-agnostic preprocessing QC and independent protocol review, before first fitted model/result.
1. 'Materially' in the question is descriptive only; no equivalence margin or clinical threshold is claimed. Report magnitude and uncertainty continuously.
2. Brier score, sensitivity/specificity and probability threshold0.5 apply only to logistic/RF/intercept outputs. C1Q rank and centroid are uncalibrated scores and receive ranking metrics only; no sigmoid is passed off as clinical probability.
3. All three deposited matrices have finite log-range intensities (P99 10.09,11.913,11.077). Native-expression sensitivity will use the deposited values unchanged, with source-only feature scaling, not an adaptive target transformation. The general parser logged the originally planned rule but transformed no matrix.
4. Conservative potentially shared-control suffix sensitivity will use sex plus numeric control suffix, and is explicitly an identity uncertainty scenario. Primary title groups have no conflicting sex. Case suffix groups are not treated as verified donors; observed cross-bed sex conflicts preclude that assumption.
5. Within-fold nine-cell reweighting avoids comparisons of independently trained score scales. Split ranges are descriptive. Any fixed-prediction bootstrap is marked conditional and excludes fitting uncertainty; full-pipeline subsampling provides separate stability evidence, not a confidence interval.
6. Annotation/QC found 15,235 common unambiguous genes; include all of this vocabulary before train-only feature ranking. Absence of cohort gene-level missingness means no fitted imputation needed. Sample-wise ranks are computed before conservative median identity aggregation; this ordering is fixed for every run.
