## Supplementary material for "Anatomical comparison populations and threshold transport in vascular transcriptomic classification": Reproducibility archive: code, provenance and results: exploratory_extension_04_20261026.txt

EXPLORATORY FIGURE AND SCIENTIFIC-EVIDENCE EXTENSION
26 October 2026. This record is written AFTER inspection of the original study results and a preliminary saved-score diagnostic. It is not preregistration, a blinded external validation, or a replacement for the original primary analysis.

Trigger and prior exposure
The user requested a substantive improvement in figure quality/quantity after studying original papers linked by Ling Yang and arXiv:2609.14709. The original six figures/26 panels, numerical results and author-review PDFs are retained in a dated snapshot. The original primary hypothesis and fitted external model remain unchanged.
A design-audit agent already inspected a preliminary threshold based on each source label group's average repeated held-out score: maximum-balanced-accuracy midpoint 0.22512067247436887; external balanced accuracy0.5 inGSE43292 and0.53125 inGSE28829. The same audit inspected mean external logits approximately -8.3072 and -3.4185, source intercept0.28075, and source-standardized absolute-z>3 proportions. These observations are disclosed prior exposure; no confirmatory status is claimed for subsequent analyses.

Questions
1. Does high source and external ranking require outcome-guided selection of the original200genes, or is it also present in label-independent random panels and a disjoint second-ranked panel?
2. How stable is gene membership/sign across the exact original30sourcefits, and what does the frozen linear model's arithmetic reveal about shifted score scales?
3. Does threshold failure persist for thresholds derived only in source-training data, evaluated in nested anatomical holdouts and applied unchanged externally?
4. Which known patient pairs and specimens are discordant across fixed scoring rules, and how dependent are aggregate external results on individual observations?

Specifications for additions
Retain original input vocabulary, sample-wise ranks,95source label groups, source-specific scaling, Welch-type feature ordering, primarylogistic C0.1 and originalsplit seeds. Refit the exact30primaryfits only to recover feature membership/coefficients; verify their held-out scores against saved predictions before interpreting new outputs.
Draw20random200gene panels without consulting any outcome using seed20261026, once from the15235gene vocabulary. Use the same20panels in every original fold and in full-source external fits. These are representation draws, not independent cohorts. Record every panel and seed; do not select a winning panel. Report20panel mean-fold values and panel ranges, not a confidence interval or600independent replicates. Include a source-only disjoint alternative selecting ranks201–400 in each training fold after excluding its top200. No primary-model replacement.
Threshold selection uses only training-source outcomes. Evaluate fixed0.5, maximumYoudenJ with a documented deterministic tie rule, and a source high-sensitivity rule. In each anatomical holdout, source innercross-fitting selects thresholds and the fulloutertraining fit supplies target scores. Full-source thresholds use source cross-fitting and are applied to the unchanged existing frozen external scores. Target-label threshold-response curves are descriptive only; no target-optimal threshold, target recalibration, or clinical utility recommendation will be promoted.
Feature and logit decompositions are arithmetic properties of the fitted classifier: intercept plus sum beta_g*(x_g-mu_g)/sd_g. They neither separate platform/procurement/endpoint causes nor identify causal molecular mechanisms. Source full-fit predictions are in-sample reference diagnostics, not performance validation. Displayed named genes are chosen by source coefficient magnitude or source-fold frequency, with deterministic ties; no target-outcome-driven gene discovery.
Paired failure displays use all32patients, allfiveexpression rules, and fixed-score leave-one-patient-out influence. The corresponding histology deletion uses specimens because donor identity is unknown. These are sensitivity/influence descriptions, not new independent validation or confidence intervals. Any failed-pair case display explicitly states retrospective failure selection.

Outputs and interpretation
Write new code/results under separately named exploratory files/directories, retaining all original core outputs and hashes. Label every new scientific panel/caption and Methods subsection exploratory. Preserve negative/null findings, no extra human-agent/RL/wet-lab claims. Only retain panels that answer distinct questions; supplementary placement is determined by scientific role and legibility. ExportPDF/SVG/300dpiPNG and compile allmanuscript/source/reproducibility packages after review.
