## Supplementary material for "Anatomical comparison populations and threshold transport in vascular transcriptomic classification": Reproducibility archive: code, provenance and results: implementation_note_02.txt

2026-10-25, after main and robustness results first observed.
Independent code review identified a terminology mismatch: executed feature ranking is an absolute Welch-type t statistic, |mean_case-mean_control|/sqrt(var_case/n_case+var_control/n_control+1e-12), rather than a Cohen standardized mean difference. The implemented algorithm is retained; this note and manuscript state the exact formula. No model or outcome was selected after inspection.
The decomposition error handler was also narrowed: missing bed-by-status cells are explicitly recorded; a failure of the weighted pair-cell algebraic identity now raises an error instead of being silently suppressed. Saved predictions/results from the initial successful run are retained. Subsequent validation independently recomputes every complete-fold identity and documents any missing cells. This change does not alter predictions.
