## Supplementary material for "Anatomical comparison populations and threshold transport in vascular transcriptomic classification": Reproducibility archive: code, provenance and results: implementation_note_03_after_review.txt

Implementation note 03, 26 October 2026, after the original model results and internal review.

The paired-bootstrap function now checks its existing input contract before drawing: known patient identifiers, binary labels, and exactly two tissues with one of each class per patient. This defensive check makes the assumption used by its two-column reshaping explicit. It does not change sampling, metrics, seeds, intervals, or any valid-data computation and is not a new prespecified analysis.

The existing GSE43292 input has 32 complete patient pairs and passes. A focused saved-score check reproduces the prior algorithm exactly over 23 draws, reproduces all primary GSE43292 2,000-draw interval endpoints within 1.12e-16, and rejects incomplete, single-class, missing-ID, and nonbinary patient-block inputs. No fitted model or stored result was changed. Details: code/check_paired_bootstrap.py and evidence/revision05_bootstrap_checks.json.
