## Supplementary material for "Anatomical comparison populations and threshold transport in vascular transcriptomic classification": Reproducibility archive: code, provenance and results: implementation_note_05_logit_ranking.txt

NUMERICAL-STABILITY CORRECTION AFTER EXPLORATORY RESULTS
26 October2026. During independent checking of extension04, random_panel18 inGSE43292 was found to output64probabilities rounded to exactly1.0. The original exploratory AUROC0.5 and pairconcordance0.5 therefore reflected artificial sigmoid floating-point ties. This was an implementation-level rank-statistic problem, not evidence that the linear model lost its ranking signal.
The complete pre-correction extensionoutputs and code were preserved in evidence/figure_revision_20261026/pre_logit_correction/. Original core study outputs were not altered.
All new random/disjoint logistic-panel ranking statistics(AUROC,averageprecision andwithin-patient ordering) nowuse the classifier's unrounded decision_function(linear_logit). Every new saved prediction retains both that logit and the original probability. Technical threshold/Brier metrics stilluse probability. The samecorrection isapplieduniformly toallnewpanels/folds/targets, notjust theaffectedpanel; all20panels andsourcehyperparameters are retained. The random panel drawseed andgene lists are unchanged. No target-selectedmodel ispromoted.
This is not a new biologicalanalysis orpreregisteredchange. The reviewer discovered it after newtargetresults were inspected, andthe correction and resultingvalues mustbe reported transparently. The original primarymodel and its externalranking conclusions remainunchanged; its savedscore reconstruction is separatelyverified. Literal probability saturation remains a scoredistribution/operating-point diagnostic, while inference fromexactly tied roundedprobabilities is avoided.
