## Supplementary material for "Anatomical comparison populations and threshold transport in vascular transcriptomic classification": Reproducibility archive: code, provenance and results: README.txt

VascTransport: anatomical comparison populations and threshold transport
Version: V2 submission manuscript; scientific expansion 26 October 2026, declarations finalized 27 October 2026

Attachment-format correction, 29 October 2026: six serialized rank/native matrix
caches are excluded from the reproducibility ZIP in response to medRxiv screening.
Scientific data, saved results, manuscript text and reviewed figures are unchanged.
Compact-upload fallback, 29 October 2026: six large official GEO source inputs
are also omitted from this distribution. Their URLs, exact sizes and SHA256 hashes
remain frozen in evidence/expected_inputs_v1.json. A fresh extraction requires
online acquisition; this is not a self-contained offline package.

STATUS
This is a public-data computational study prepared for medRxiv. This archive does not establish submission acceptance or public posting. CC BY 4.0 is authorized for the manuscript. Current-study author review, assent, no competing interests, applicable ethics/consent requirements, the contribution statement, and all five funding awards have been confirmed. Final submission is authorized. Original public data retain their original conditions. No approval, exemption, trial registration or public archival DOI is invented.

READ FIRST
manuscript/main.pdf: submission manuscript, delivered separately from source archives
manuscript/supplement.pdf: supplementary methods and detailed tables
protocol/analysis_protocol_v1.txt and subsequent amendments
manuscript/statistical_addendum.tex: reporting clarifications and descriptive averaging
results/descriptive_mean_crossfit_scores.csv: all 665 model/label averages
results/descriptive_mean_crossfit_metrics.csv: seven descriptive pooled AUROCs
results/primary_fold_metrics.csv: the distinct primary within-model estimand
evidence/dataset_registry.csv, acquisition.json and expected_inputs_v1.json
evidence/independent_saved_results_validation.json
figures/figure_captions.json
figures/supplement_extension_captions.json
manuscript/exploratory_supplement.tex: additional exploratory tables and interpretation limits
protocol/exploratory_extension_04_20261026.txt: decisions made after viewing original results
protocol/implementation_note_05_logit_ranking.txt: documented sigmoid-saturation correction
evidence/figure_revision_20261026/independent_extension_validation.json

V2 FIGURE AND ANALYSIS INVENTORY
There are 11 main figures with 46 lettered panels, plus 2 supplementary figures with 11 panels. Figures 1-6 retain the original analyses. Figures 7-11 add explicitly exploratory controls for gene-panel dependence, feature membership, source-learned thresholds, frozen-score geometry and fixed-score case influence. Supplementary Figures S1-S2 expose variation already present in the original saved scores. Figure counts describe actual lettered panels, not independent experiments or evidence of expert endorsement. Every figure has vector PDF/SVG and 300 dpi PNG outputs, plus plotting code and source-table provenance.
The twenty fixed random panels are representation draws, not independent cohorts. The 30 overlapping fits and 435 pairwise gene-set comparisons are descriptive. New-control ranking uses raw linear logits to avoid artificial sigmoid-saturation ties; probabilities are retained for threshold/Brier diagnostics. No external winning panel or replacement primary signature is selected. The original core result hashes remain unchanged.

ENVIRONMENT AND HISTORICAL RECORDS
Python 3.11.9 and the installed versions pinned in requirements.txt were retained. V2 figure export/QA also imports PyMuPDF; install requirements-qa.txt, which includes requirements.txt, for the complete supported V2 rebuild. The original requirements file is not retrospectively altered.
evidence/environment.json is the earliest retained post-analysis environment snapshot, recorded during manuscript assembly on 25 October 2026 at 16:16 UTC. Its unchanged copy is analysis_environment_original.json. This is not a pre-fit lock or an independent reconstruction of the execution-time environment. requirements.txt is similarly retained without changes. Later PDF rebuilds write manuscript_build_environment.json; supplementary assembly writes supplement_environment_snapshot.json. They do not replace these historical records.

FULL REPRODUCTION FROM THE FROZEN INPUTS
From this directory, using Python 3.11:
  python -m pip install -r requirements-qa.txt
  python code/acquire.py
  python code/metadata.py
  python code/prepare.py
  python code/analyze.py main
  python code/analyze.py robust
  python code/export_descriptive_scores.py
  python code/validate_results.py
  python code/explore_feature_dependence.py
  python code/explore_threshold_shift.py
  python code/validate_exploratory.py
  python code/plot_figures.py
  python code/plot_feature_extension.py
  python code/plot_threshold_extension.py
  python code/plot_supplement_extension.py
  python code/assemble_figure_catalog.py
  python code/build_supplement.py
  python code/build_exploratory_supplement.py
  python code/assemble.py --compile

The first acquisition command downloads the six omitted official source files and verifies their exact bytes against the frozen expected-input manifest. In a populated local workspace, python code/acquire.py --offline verifies existing inputs without network access; it is not the first command for this compact package. Existing or downloaded hash mismatches fail without rewriting the expected manifest or original acquisition.json. Changed upstream data require a separate deliberately versioned study; they are not silently accepted. Official platform family archives are streamed only through the first complete platform table; the entire multi-gigabyte archive is neither downloaded nor hash-verified.

SAVED-OUTPUT REBUILD AND VERIFICATION
After installing requirements-qa.txt, use the following sequence in the extracted reproducibility archive:
  python code/acquire.py
  python code/prepare.py
  python code/validate_results.py
  python code/validate_exploratory.py
  python code/export_descriptive_scores.py
  python code/plot_figures.py
  python code/plot_feature_extension.py
  python code/plot_threshold_extension.py
  python code/plot_supplement_extension.py
  python code/assemble_figure_catalog.py
  python code/build_supplement.py
  python code/build_exploratory_supplement.py
  python code/assemble.py --compile

All mapping CSVs, sample tables and saved scores/results are retained. Three large source expression matrices and three complete platform text tables are omitted and reacquired from their official URLs by acquire.py. The six rank/native .pkl files are deterministic processed-matrix caches, not trained models, and are intentionally absent from submission ZIPs. The prepare.py command regenerates them locally from the downloaded, hash-verified text inputs before validation. It performs no model fitting or network retrieval; its preprocessing-QC timestamp describes this local regeneration. Do not upload the regenerated caches. No scientific estimator or input definition was changed for this attachment-format correction. validate_results.py checks exact original output inventories, partitions, metrics/cells, frozen coefficient reconstruction, sensitivity concatenation, descriptive averages and null/subset structure without refitting. validate_exploratory.py independently recomputes the new metrics and source-only selections, verifies original core hashes, and performs TWO limited fitted-panel replays (next-ranked and random_01 in one source fold). It is not a full independent model refit or a clean-environment experiment. The plotters, catalog, supplement builders and assembler do not fit models. code/check_paired_bootstrap.py supplies a further primary paired-bootstrap interval/contract check without source refitting.
The validation dependency evidence/figure_revision_20261026/pre_expansion/manifest.json is included as a historical hash record only. Old PDFs, old code copies and the pre-correction output directory are excluded. Implementation note 05 discloses the numerical correction; the V2 archive contains the corrected operative results. A fresh refit must still match the frozen original-result hashes before it is accepted as an exact reproduction.

TEX AND VISUAL QA
PDF compilation requires pdflatex and bibtex on PATH. Required LaTeX packages include geometry, fontenc, inputenc, lmodern, amsmath, amssymb, graphicx, booktabs, longtable, array, xcolor, pdflscape, microtype, setspace, caption, float, hyperref and natbib. MiKTeX is detected and its automatic installer is disabled; install required packages before compiling. Other pdfLaTeX distributions do not receive that MiKTeX-specific option.
Additional page rendering uses:
  python -m pip install -r requirements-qa.txt
  python code/render_review.py --pdftoppm PATH_TO_POPPLER_PDFTOPPM
The argument can be omitted when pdftoppm is on PATH. Page render directories are keyed by PDF SHA256, preventing stale pages from an earlier longer PDF. Automated text-bound checks skip rotated pages and do not substitute for visual review. Local historical evidence helper scripts, including build_registry_and_benchmark_audit.py, are excluded from the supported public pipeline and archives; the verified registry is included as a source record.

RELEASE PACKAGES
After the final source/PDF review, run:
  python code/package_release.py --check
  python code/package_release.py --smoke --compile
The check command validates the explicit file allowlists without writing archives. Packaging creates delivery/VascTransport_v2_source.zip and VascTransport_v2_reproducibility.zip, with an internal RELEASE_MANIFEST.json hashing every included file. delivery/package_audit_v2.json records archive hashes, CRC checks and relocation smoke logs. V1 archive names are not overwritten. The source archive contains the assembled LaTeX files, all 13 figures in three formats and captions; it compiles directly from manuscript/main.tex and supplement.tex using pdflatex/bibtex. The reproducibility archive additionally supplies the explicit code/data/results/protocol/evidence allowlists.
The smoke check runs all commands in the saved-output sequence above in a fresh short-path temporary extraction, optionally including compilation. It first confirms all six official inputs and caches are absent, actually downloads and hash-verifies the inputs, regenerates the caches and compares their bytes with the retained local reference caches. It checks immutable source records and original core-result hashes remain unchanged. With --compile it also compares all regenerated main and supplement pages with the reviewed PDFs by text, page dimensions and 108 dpi pixels. Its exploratory validator includes two limited fitted-panel replays; it does not install a fresh environment, rerun the complete experiment or regenerate bootstrap draws. Network availability and unchanged official input bytes are required; any mismatch stops the check. Archived PDFs/figures are the reviewed delivery artifacts; later recompiled PDF bytes can differ because of compiler metadata or environment changes even when scientific content agrees. On Windows, extract under a short path such as C:/VascTransport to avoid toolchain path-length limits.
Noncorresponding author emails and private authorization fields are omitted from public scientific-archive metadata. The full local portal metadata is unchanged. Main and supplementary PDFs are delivered separately. After the integrated release audit and smoke checks pass, the local code/package_author_review.py creates delivery/VascTransport_author_review_20261026_v2.zip and author_review_package_audit_v2.json. That PRIVATE author-review bundle contains local contact/portal metadata, the PDFs, review records and the two minimized scientific ZIPs. It must not be confused with a public-upload archive. Package generation does not post or submit the work.

INFERENCE
The primary 95 units are conservative specimen-title groups, not verified independent patients. The original publication reports 96 arteries and GEO contains 104 arrays; the discrepancy remains unresolved. GSE43292 supplies 32 paired patients. GSE28829 supplies 29 tissue specimens, with donor independence unresolved. Never sum these into one independent-patient total.
Within-fold AUROC decomposition uses one fitted model per held-out fold. Repeat ranges are not confidence intervals. Averaged cross-fitted scores are descriptive and mix fitted models. External bootstrap intervals condition on the frozen source fit; they omit source fitting and unresolved donor uncertainty. Anatomical holdouts are not authenticated donor-disjoint validation. Processed inputs retain original cohort-wide normalization.
Primary ranking is near perfect in the source and high in the two external contrasts. The frozen 0.5 rule labels every external sample negative. This is a technical threshold-transport finding, not a deployed diagnostic test result. Target comparisons differ, and platform/procurement/comparator changes cannot be disentangled causally.

PRIOR EXPOSURE
These datasets had prior public use. GSE43292 was descriptively analyzed in the separate VascFlux-GEO manuscript. The present whole-transcriptome scores, comparisons and results were newly executed. Literature controls C1Q and Wu6 have feature-selection exposure in the same public datasets.

FILES AND RIGHTS
Archives use an explicit allowlist, excluding private correspondence, credentials, unrelated manuscript histories, copyrighted third-party full texts, benchmark source PDFs and caches. Local saved reference PDFs are for scholarly review, not redistributed. Public GEO data retain their original repository conditions; no third-party rights are waived by the manuscript CC BY authorization. No passwords or verification codes are stored.
The packager additionally rejects .pkl, .pickle, .joblib, .pyc, .pyo, .npy and .npz members; only the explicit reviewable input/output formats and document/figure formats are distributed. Local use of deterministic caches is separate from their inclusion in submission attachments.

AI ASSISTANCE
Actual AI assistance extended to literature/provenance checks, code, execution, debugging, plotting and review. The requested declaration appears once in the main manuscript, with a factual Methods disclosure. AI reviews are separate tasks, not independent expert endorsement. Human review was confirmed by the authors; final review of the revised submission files is still required before Submit.
