## Supplementary figures and images for "Anatomical comparison populations and threshold transport in vascular transcriptomic classification"

### Figure_1.pdf

#### Study design and source-tissue structure

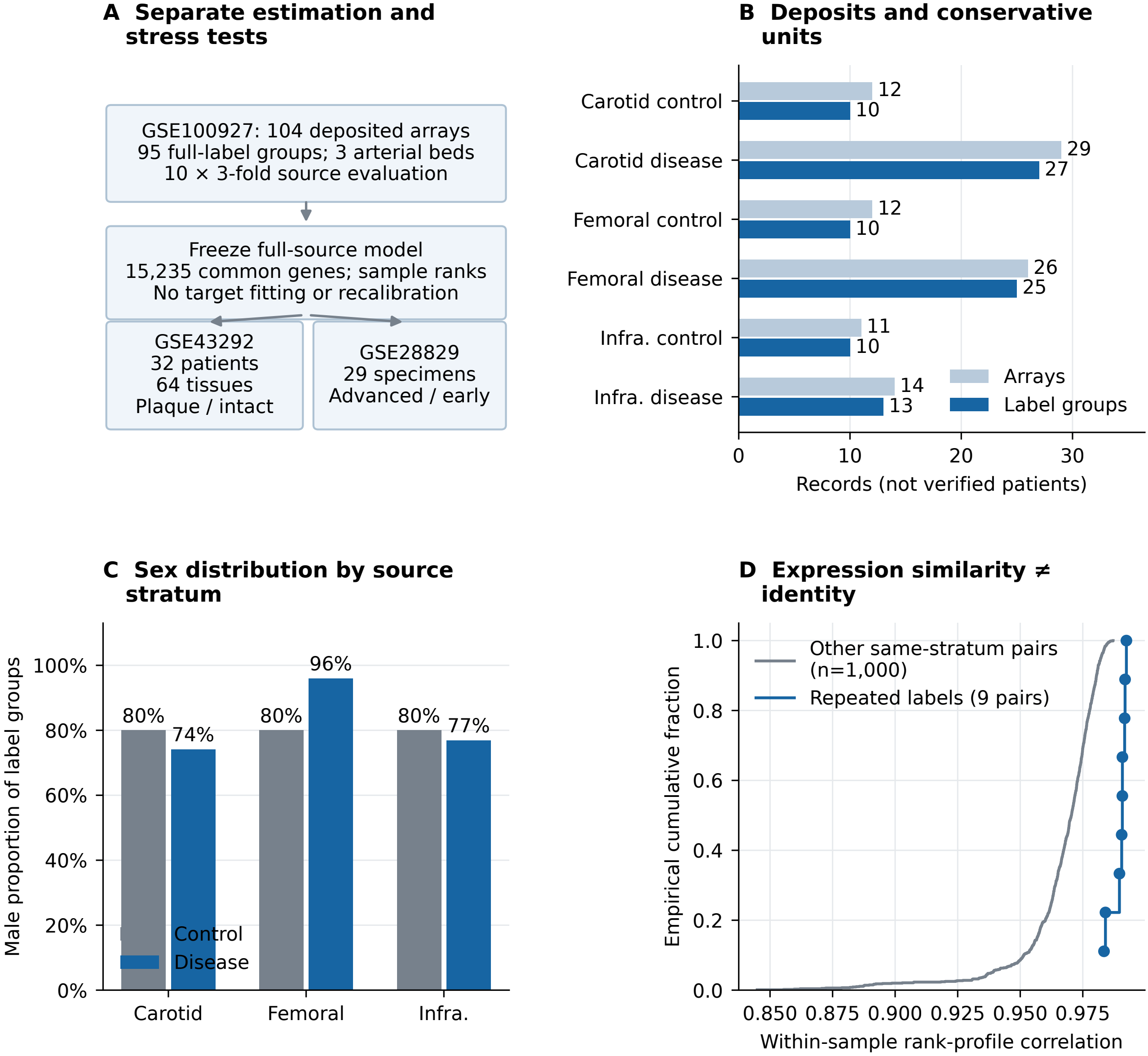

### Figure_1.png

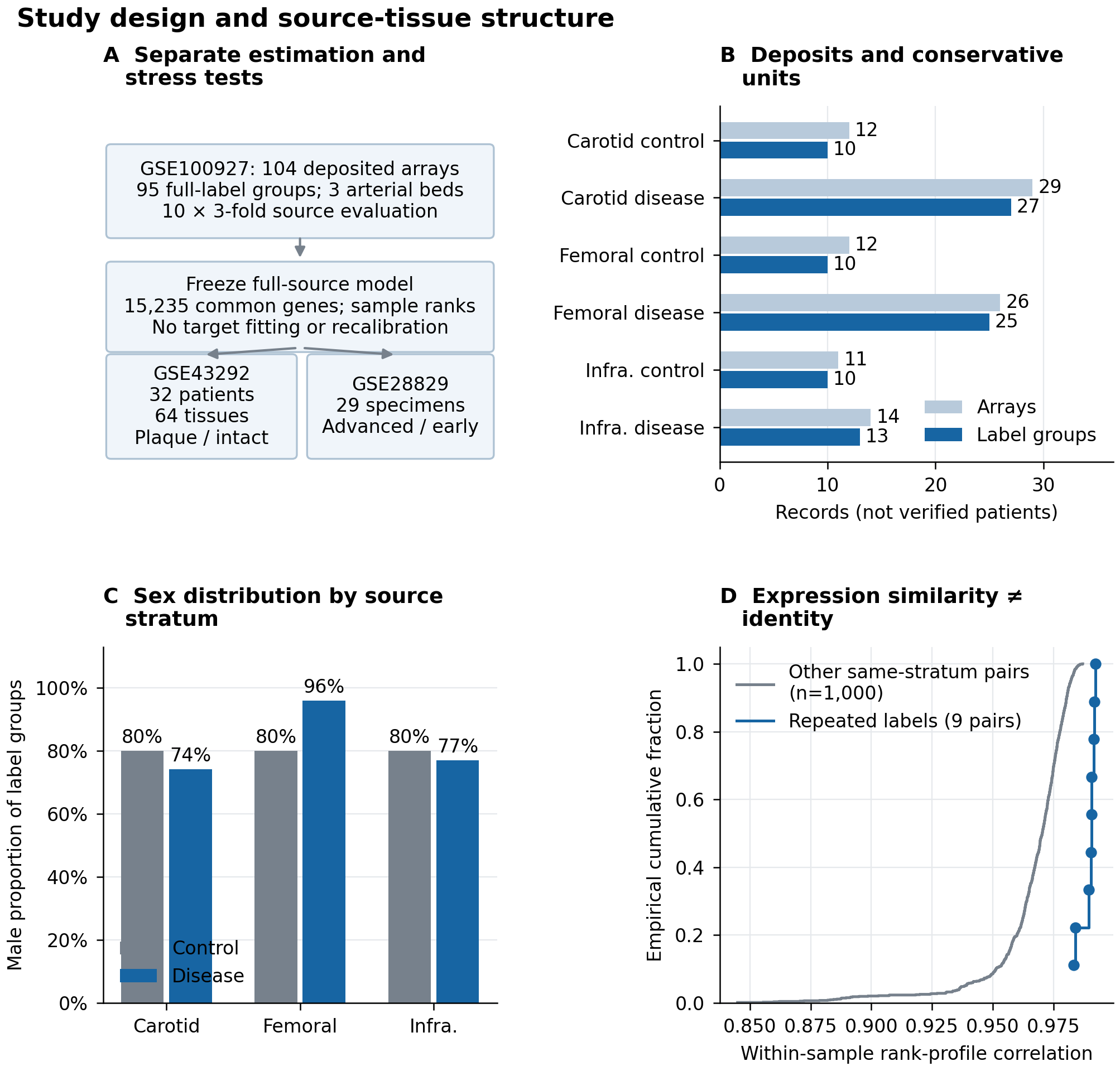

### Figure_2.pdf

##### Discrimination and anatomical comparison populations

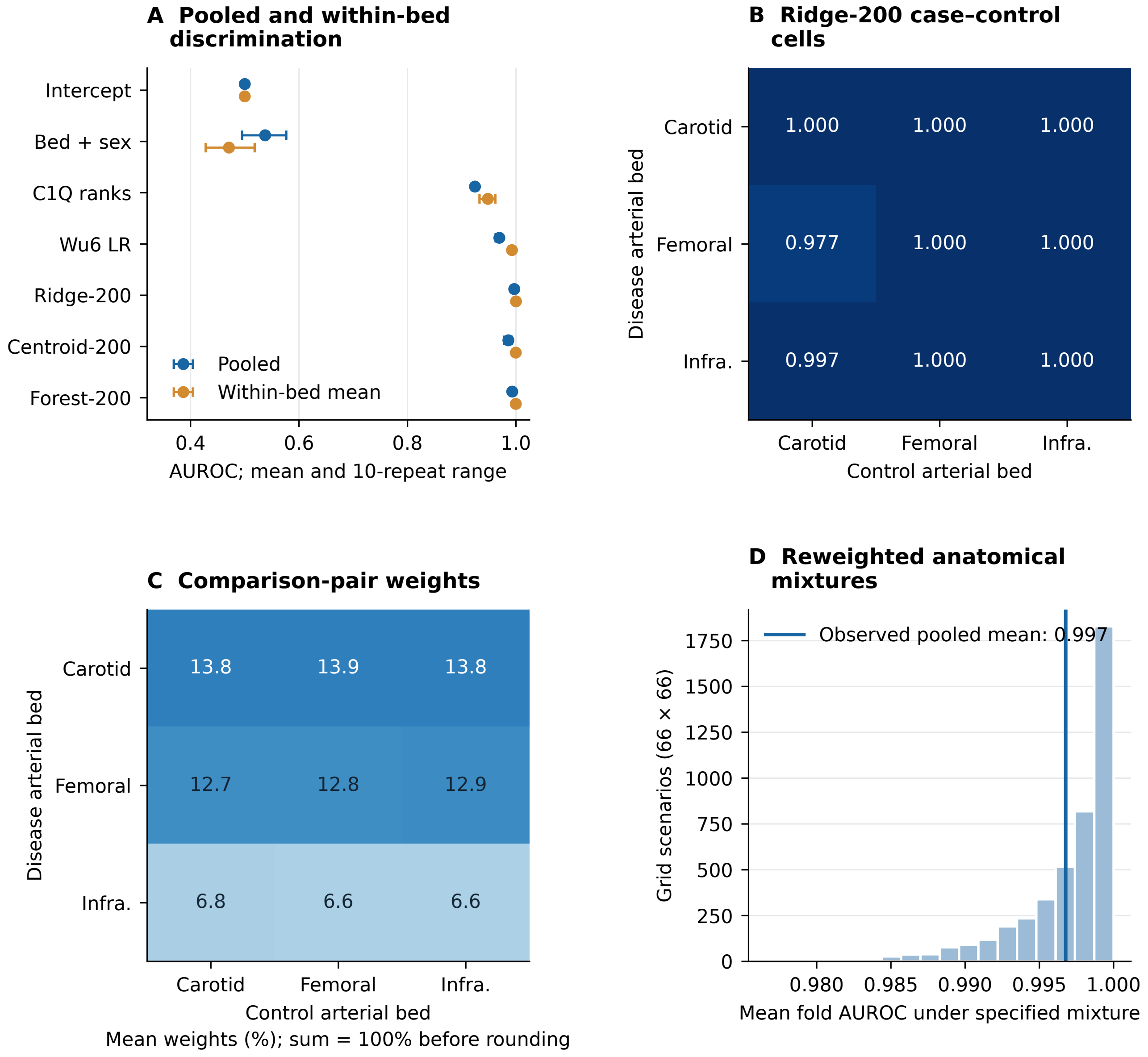

### Figure_2.png

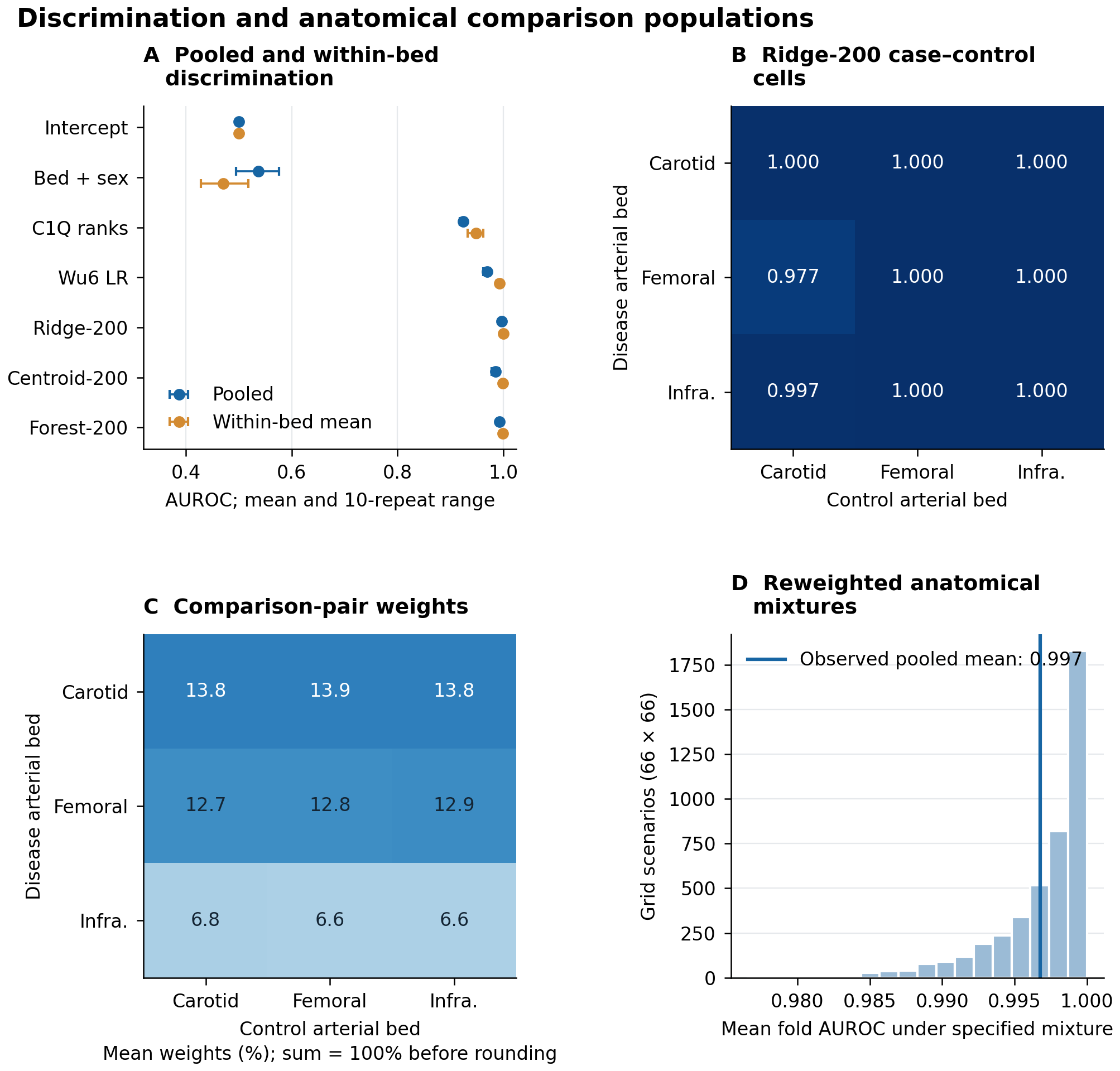

### Figure_3.pdf

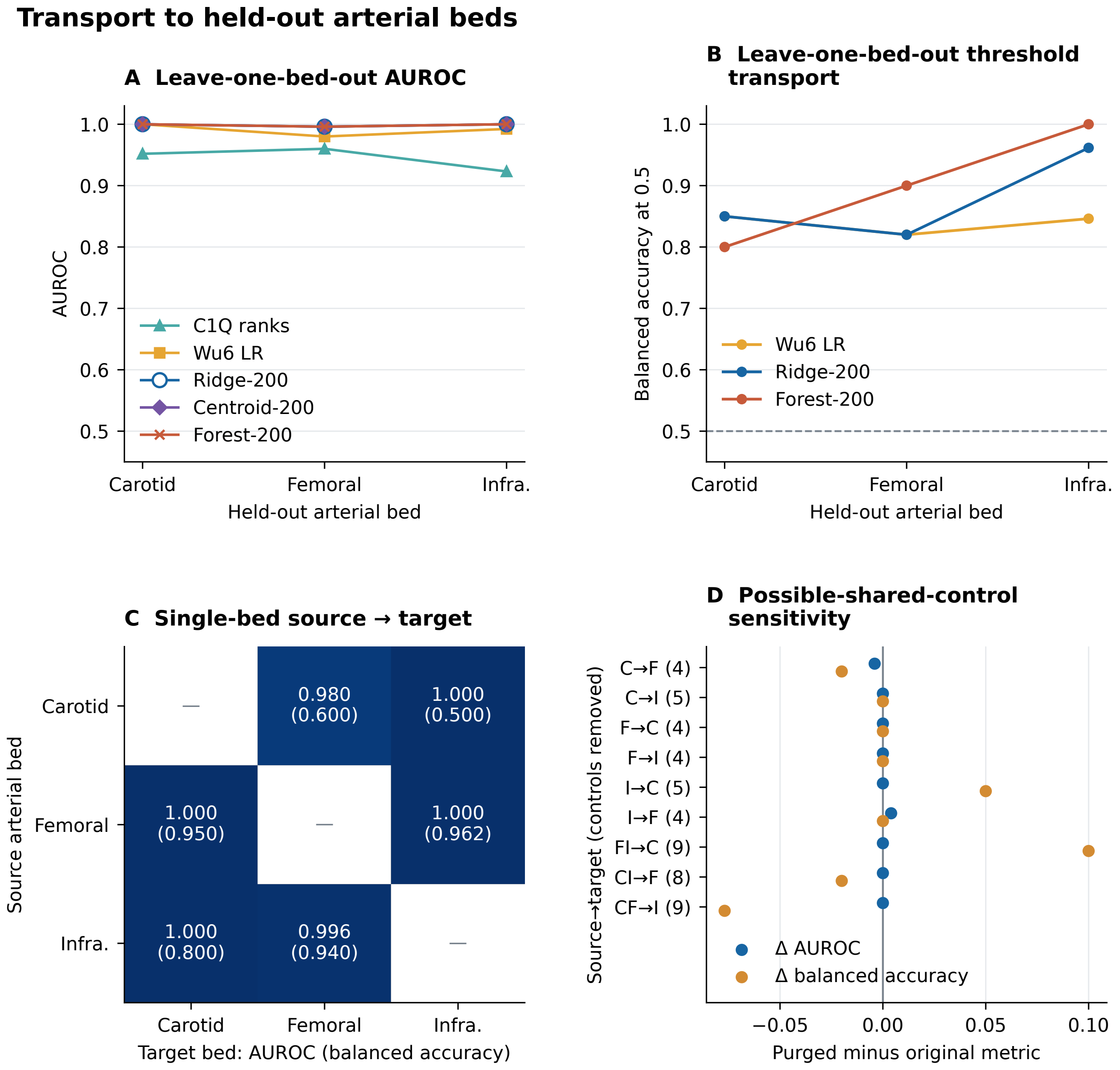

### Figure_3.png

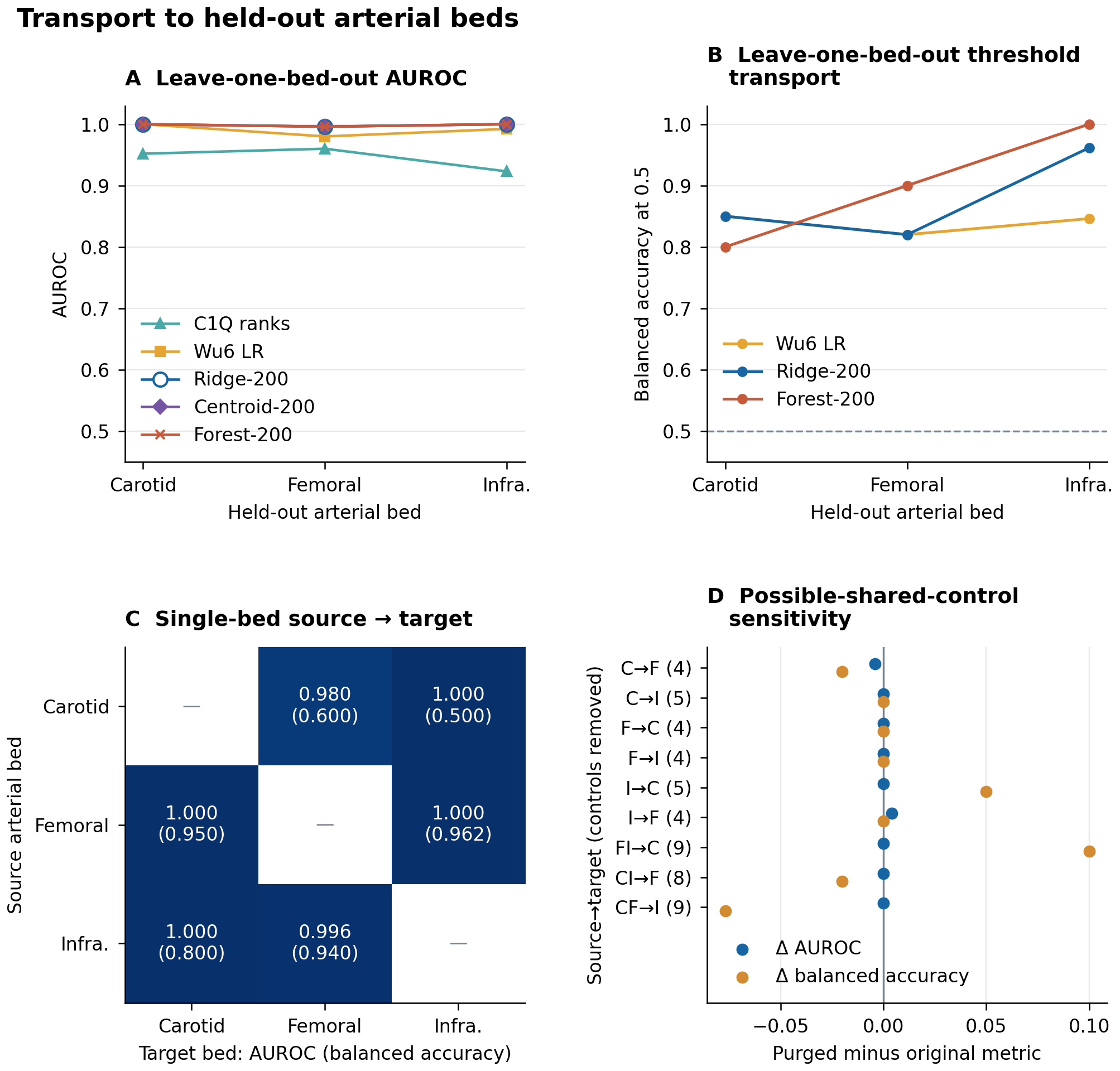

### Figure_4.png

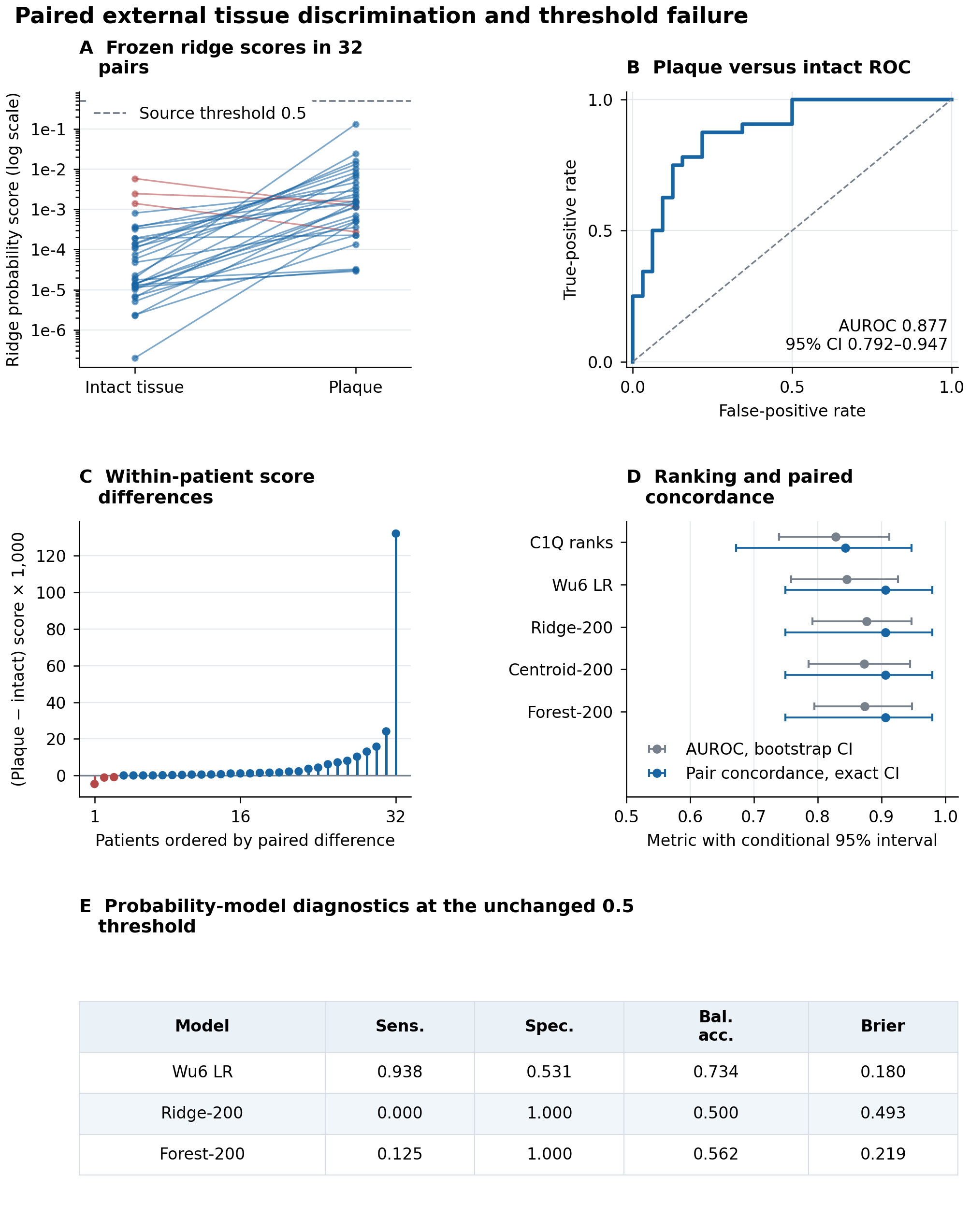

### Figure_5.png

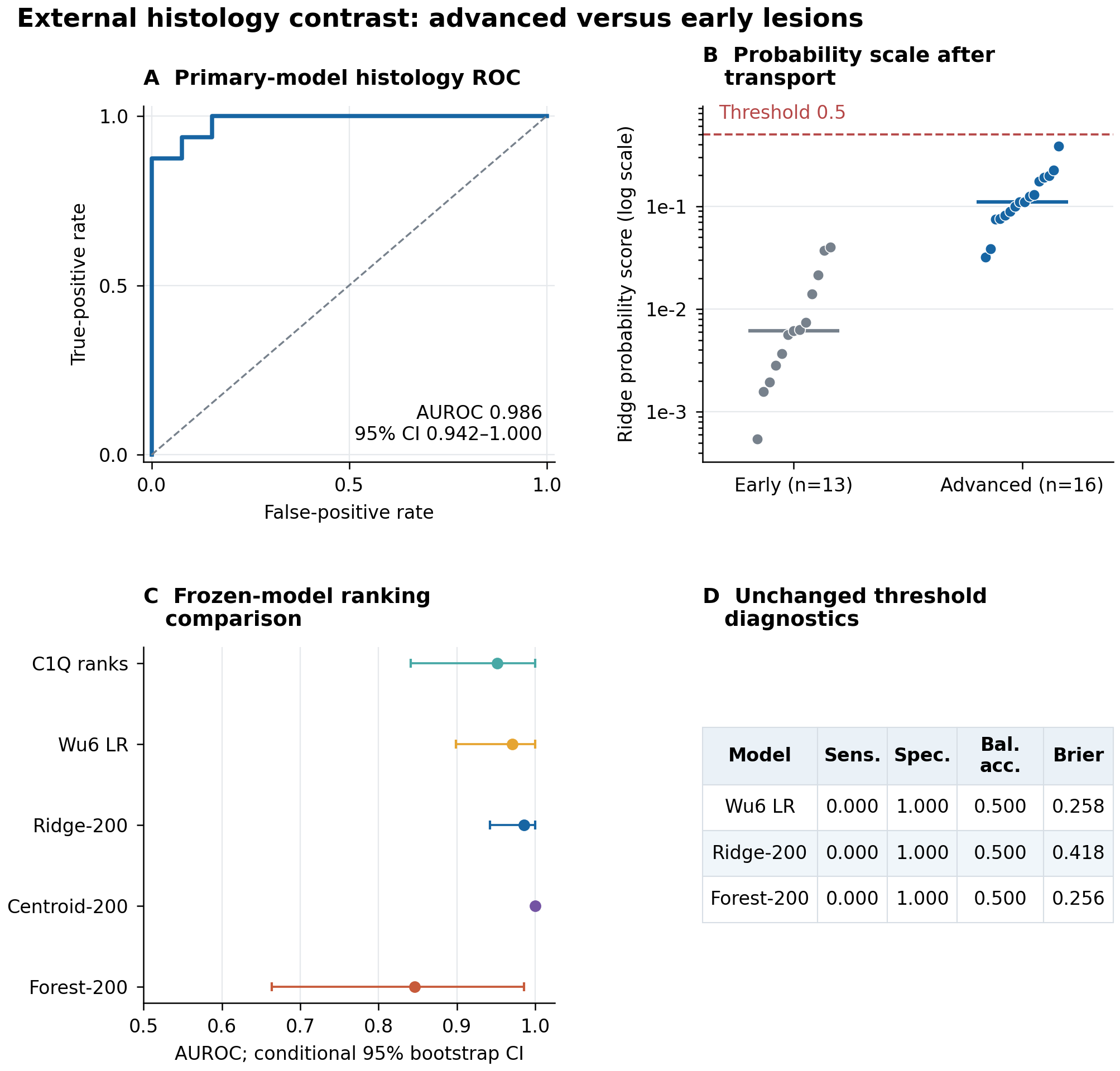

### Figure_6.pdf

#### Specification, negative-control and subset stability analyses

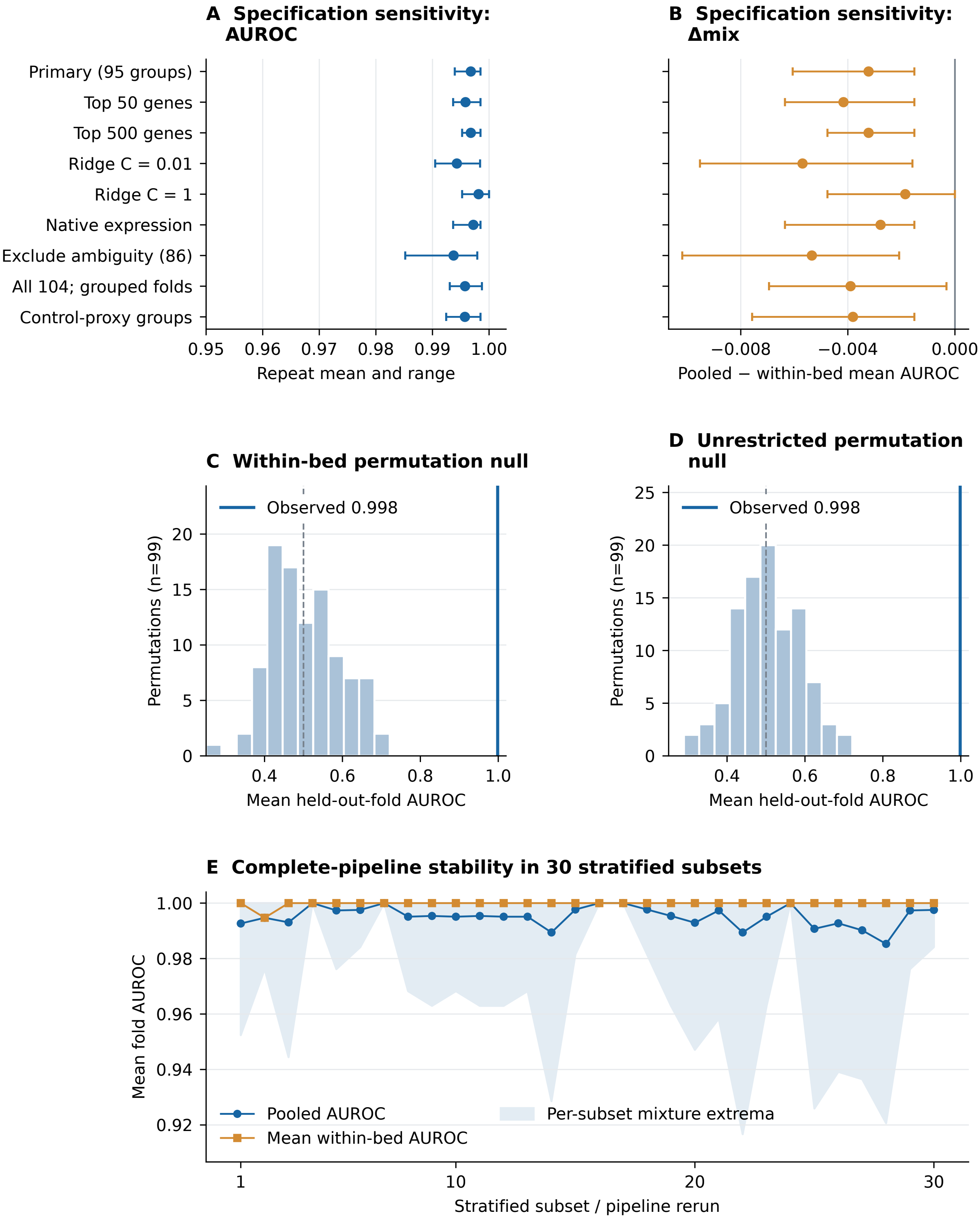

### Figure_6.png

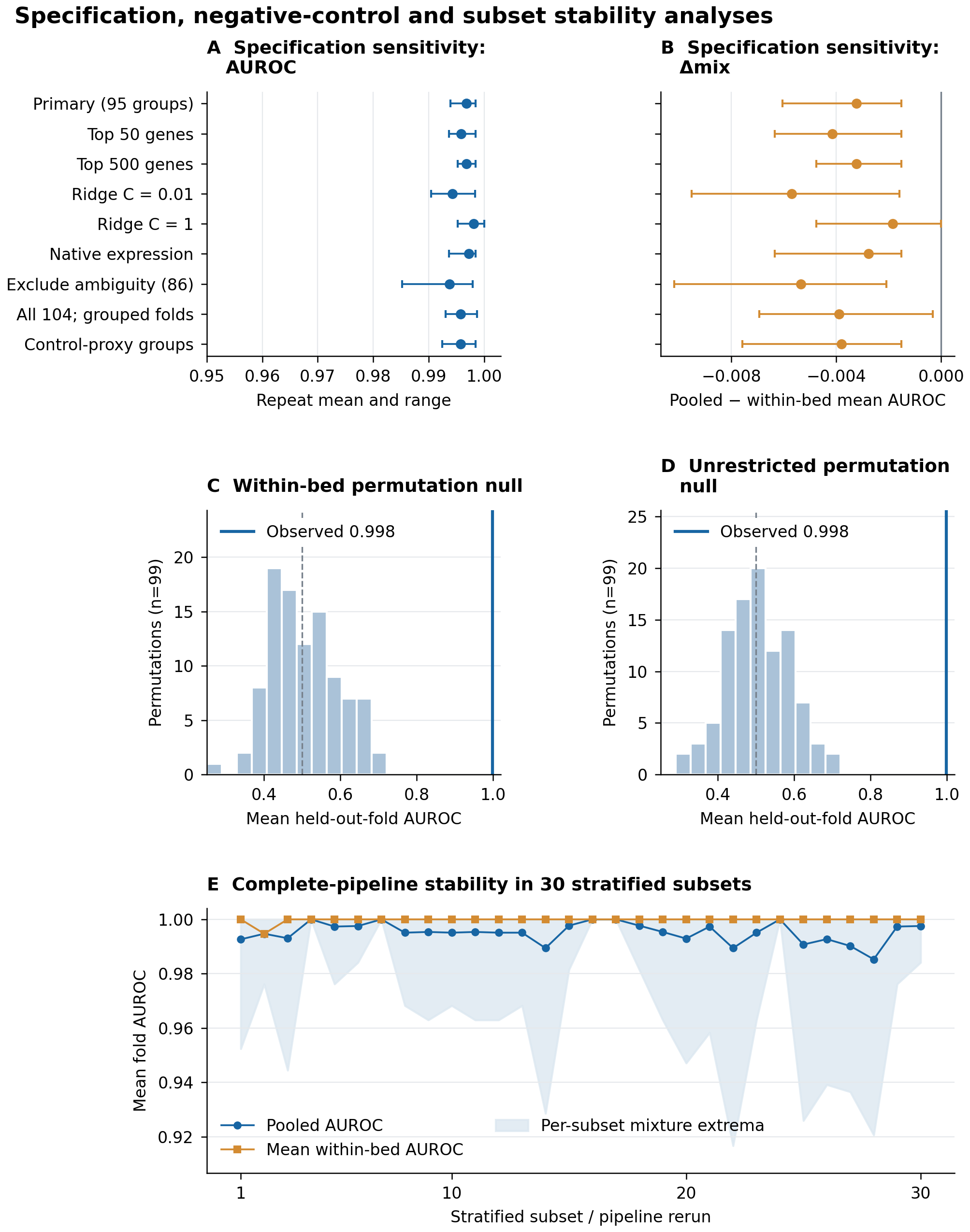

### Figure_7.pdf

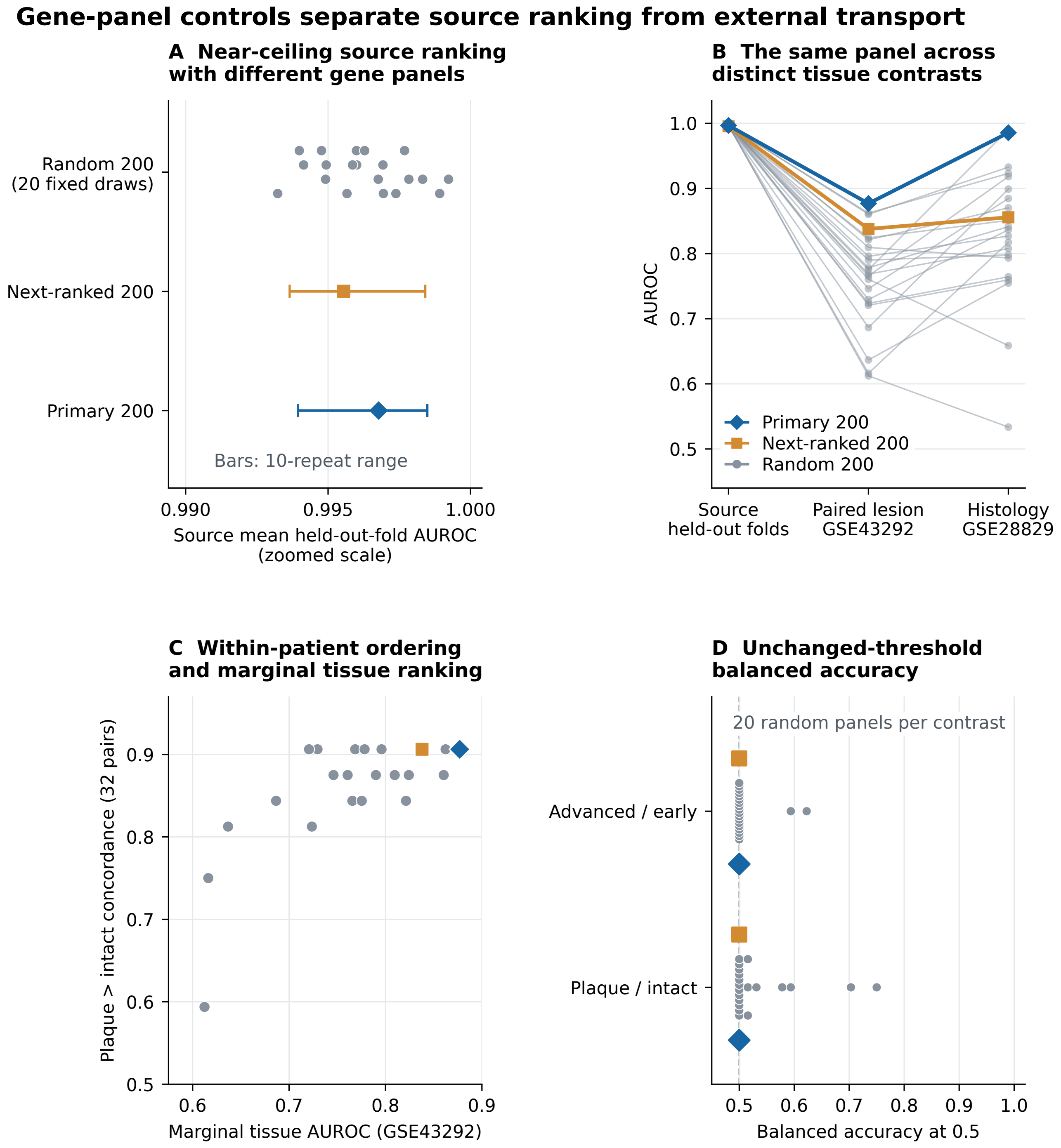

### Figure_7.png

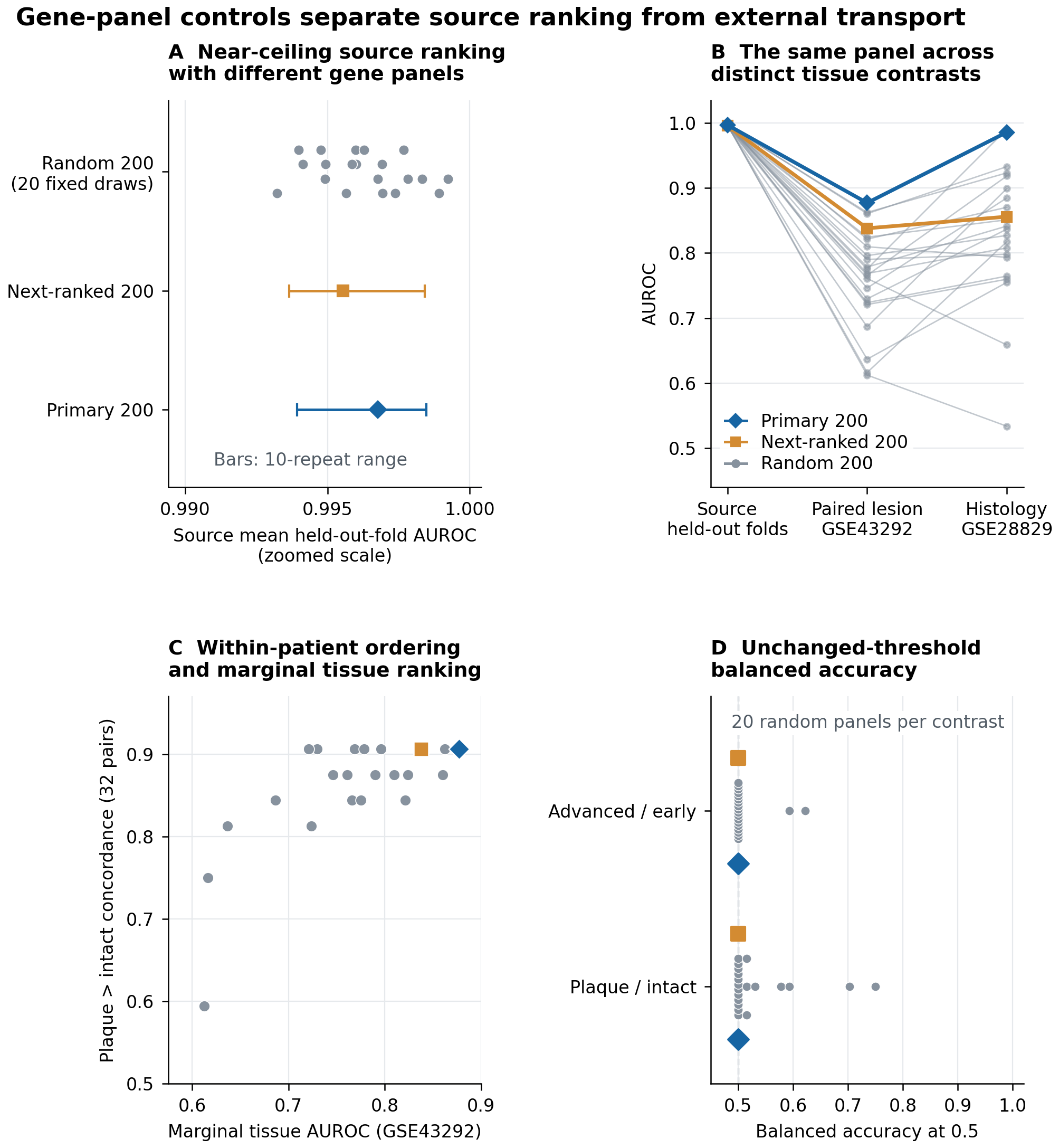

### Figure_8.pdf

#### Feature membership varies across fits despite stable source ranking

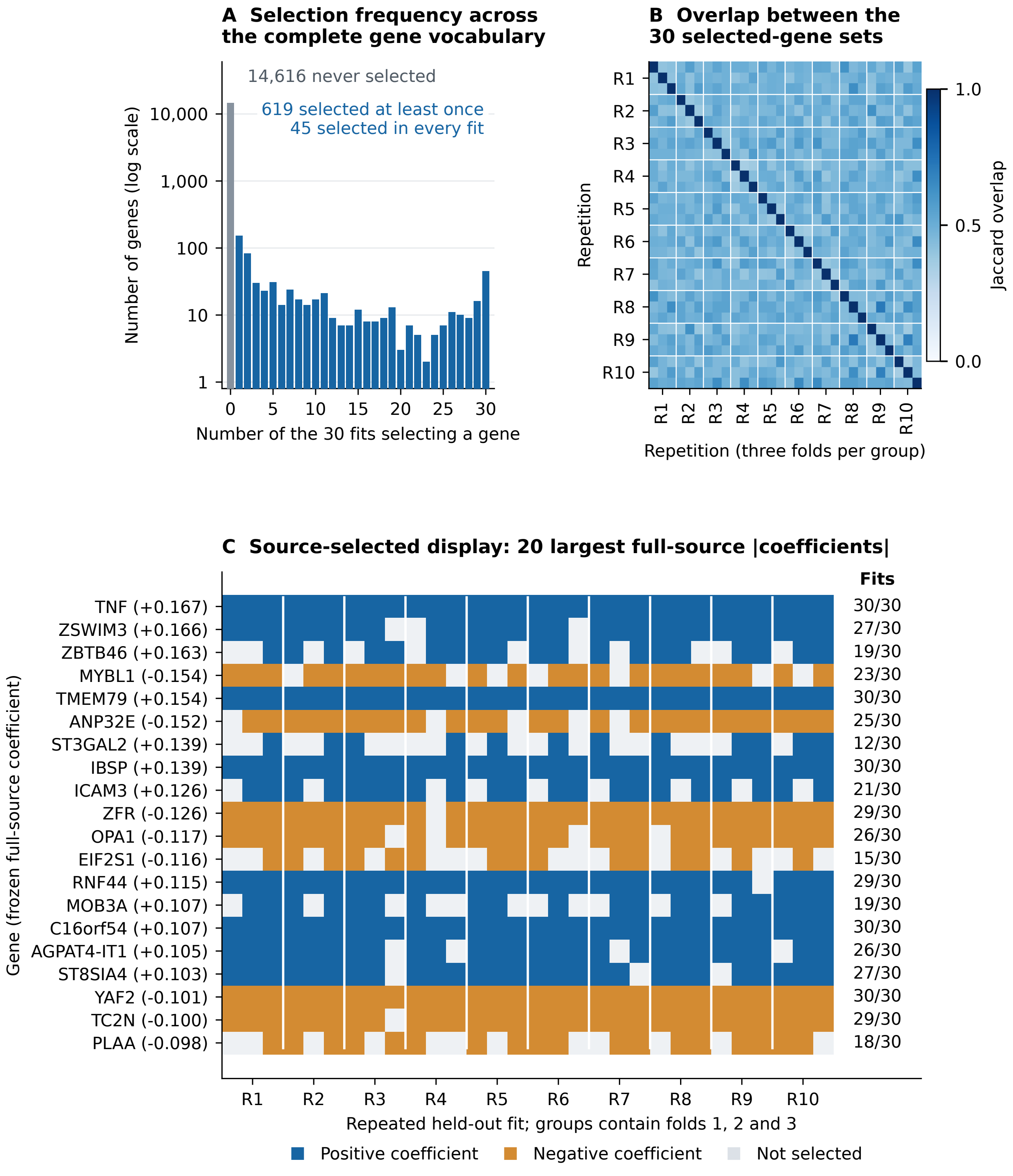

### Figure_8.png

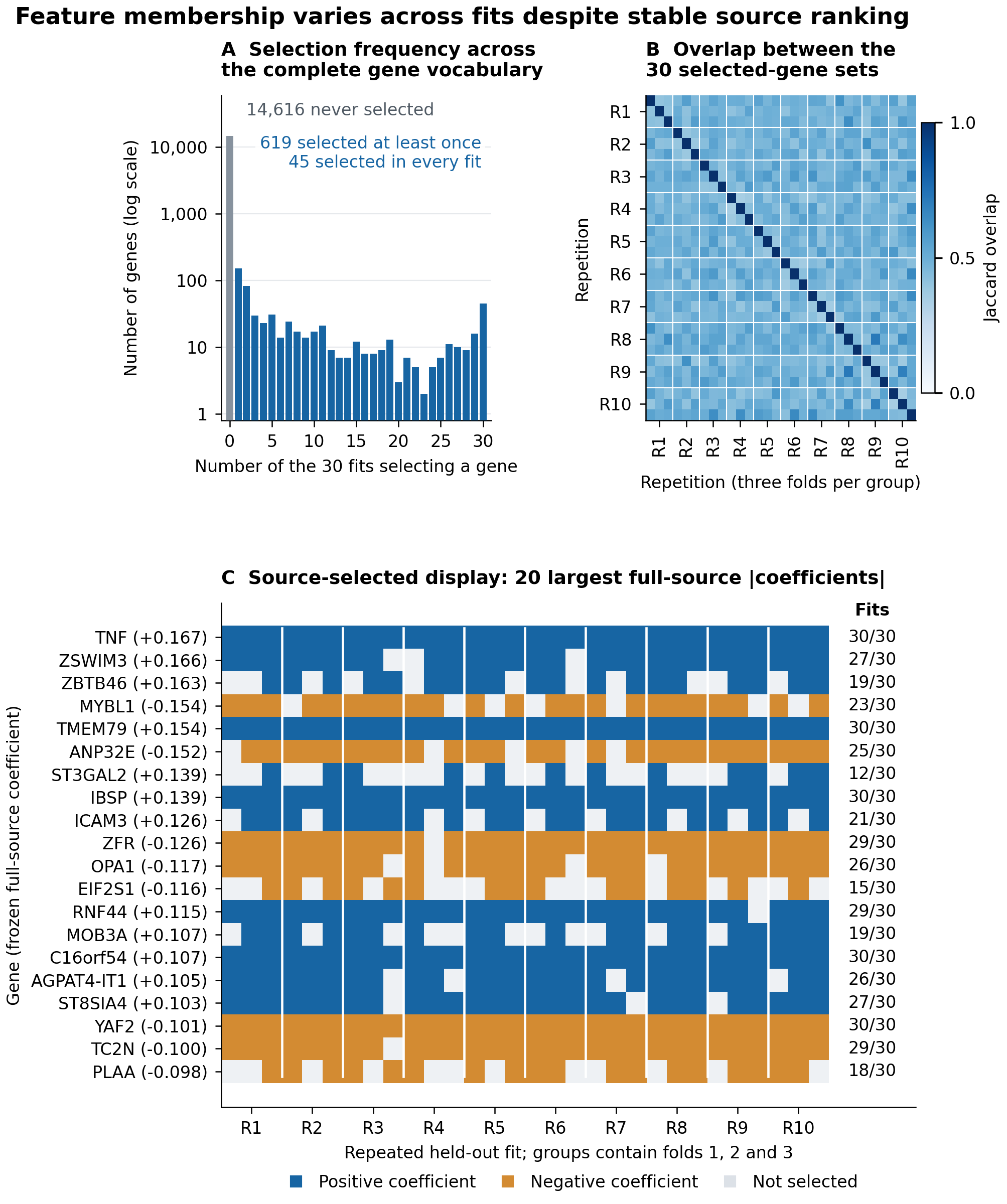

### Figure_9.pdf

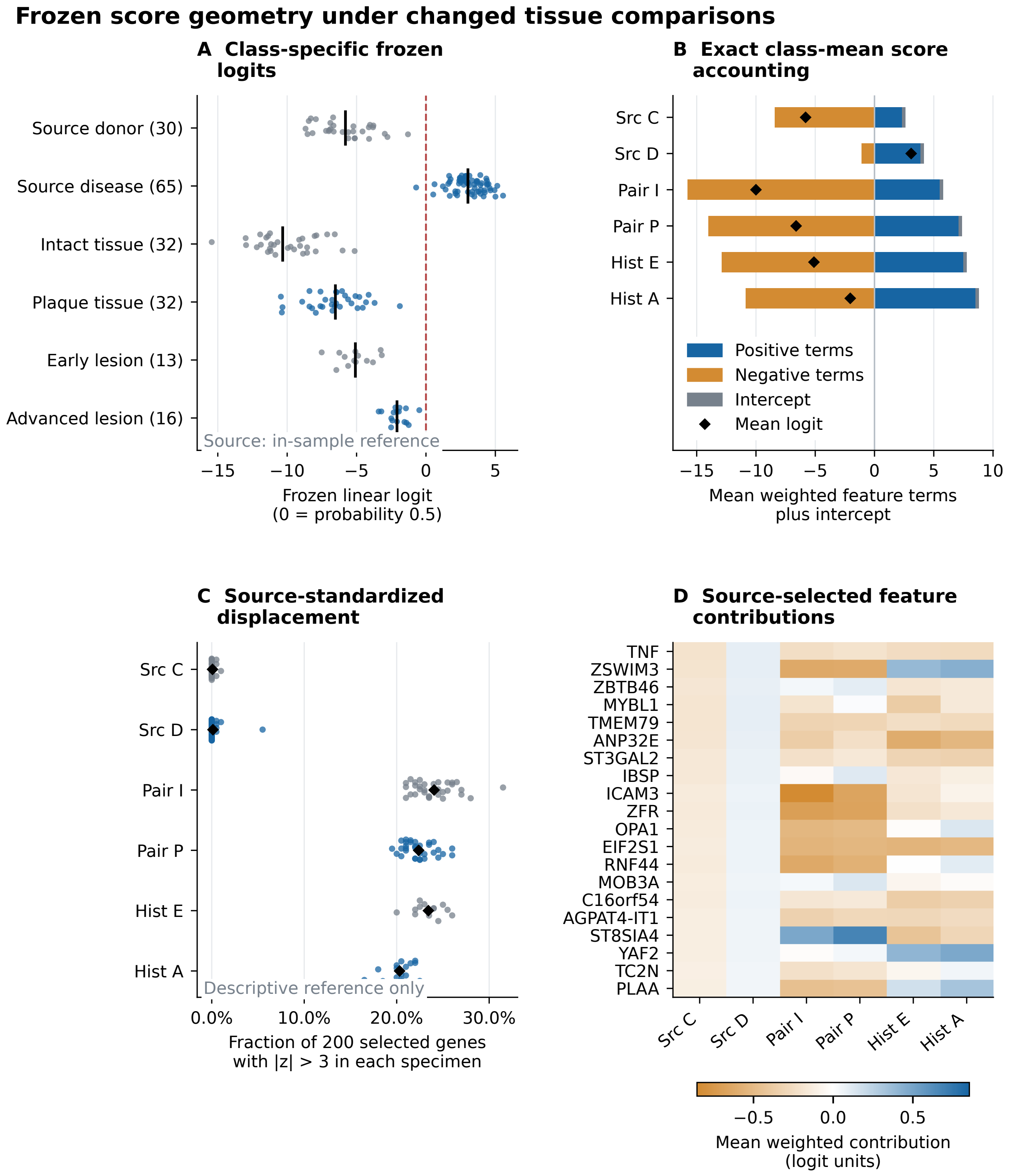

### Figure_9.png

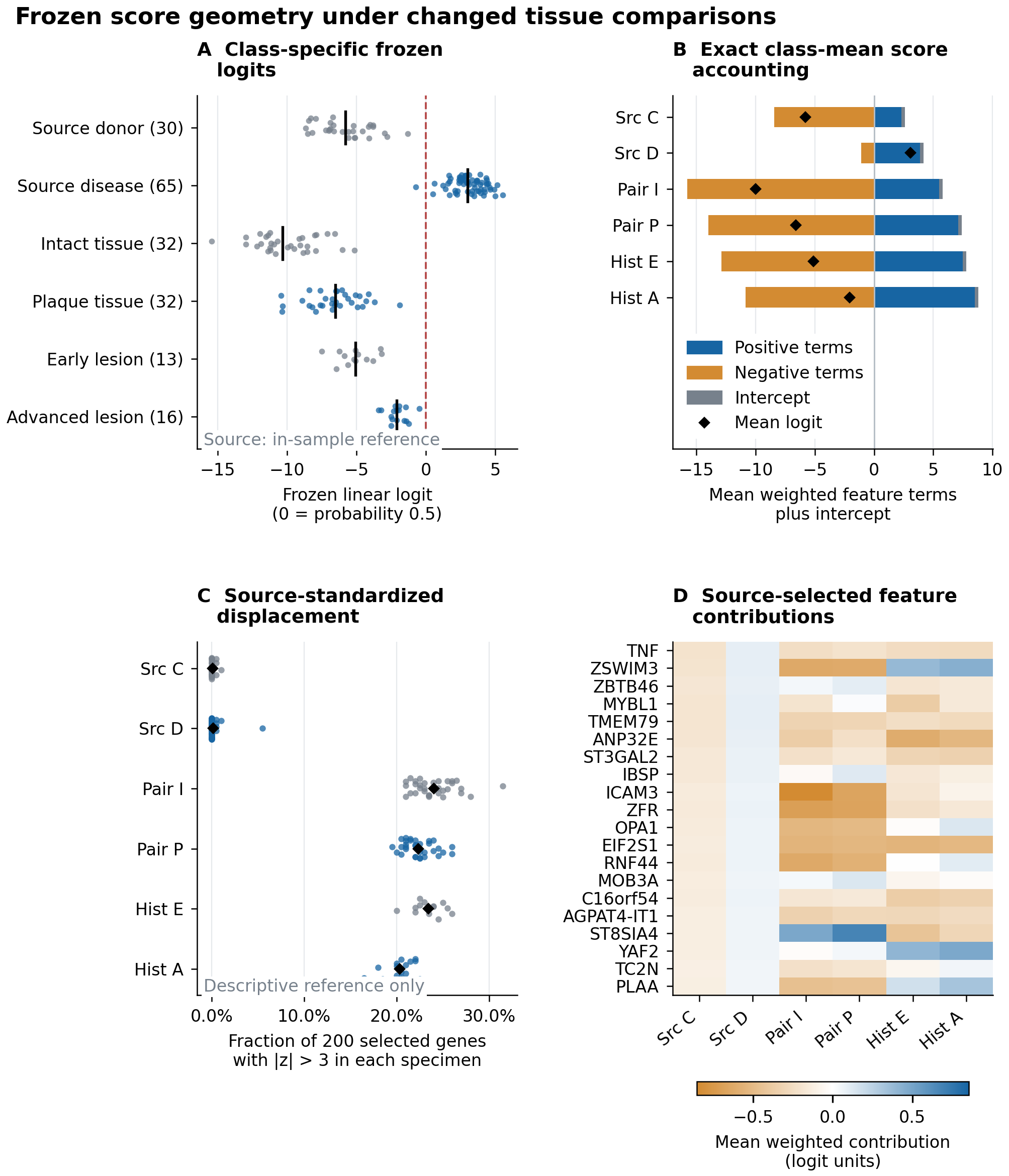

### Figure_10.png

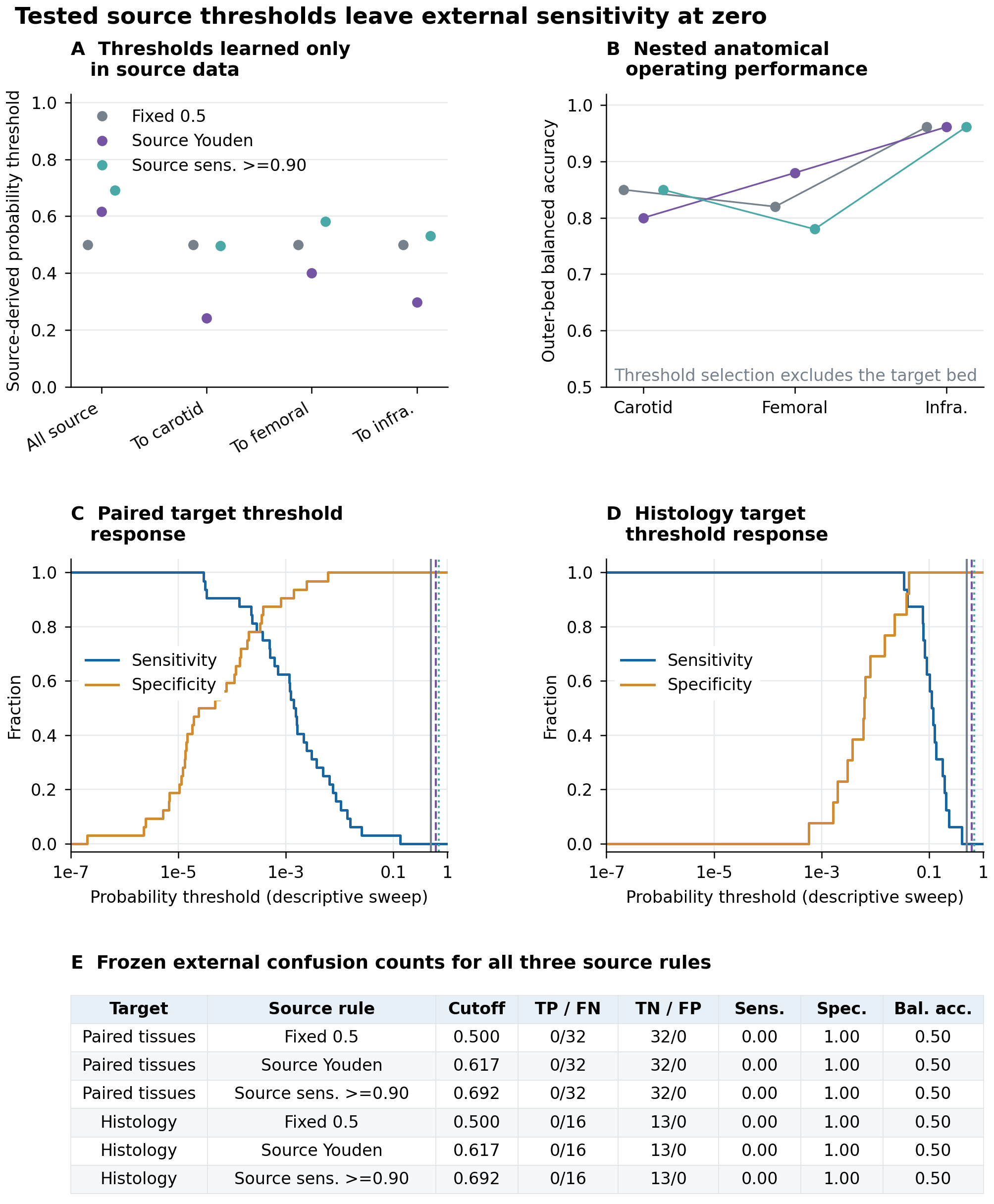

### Figure_11.pdf

#### Individual failures and fixed-score influence boundaries

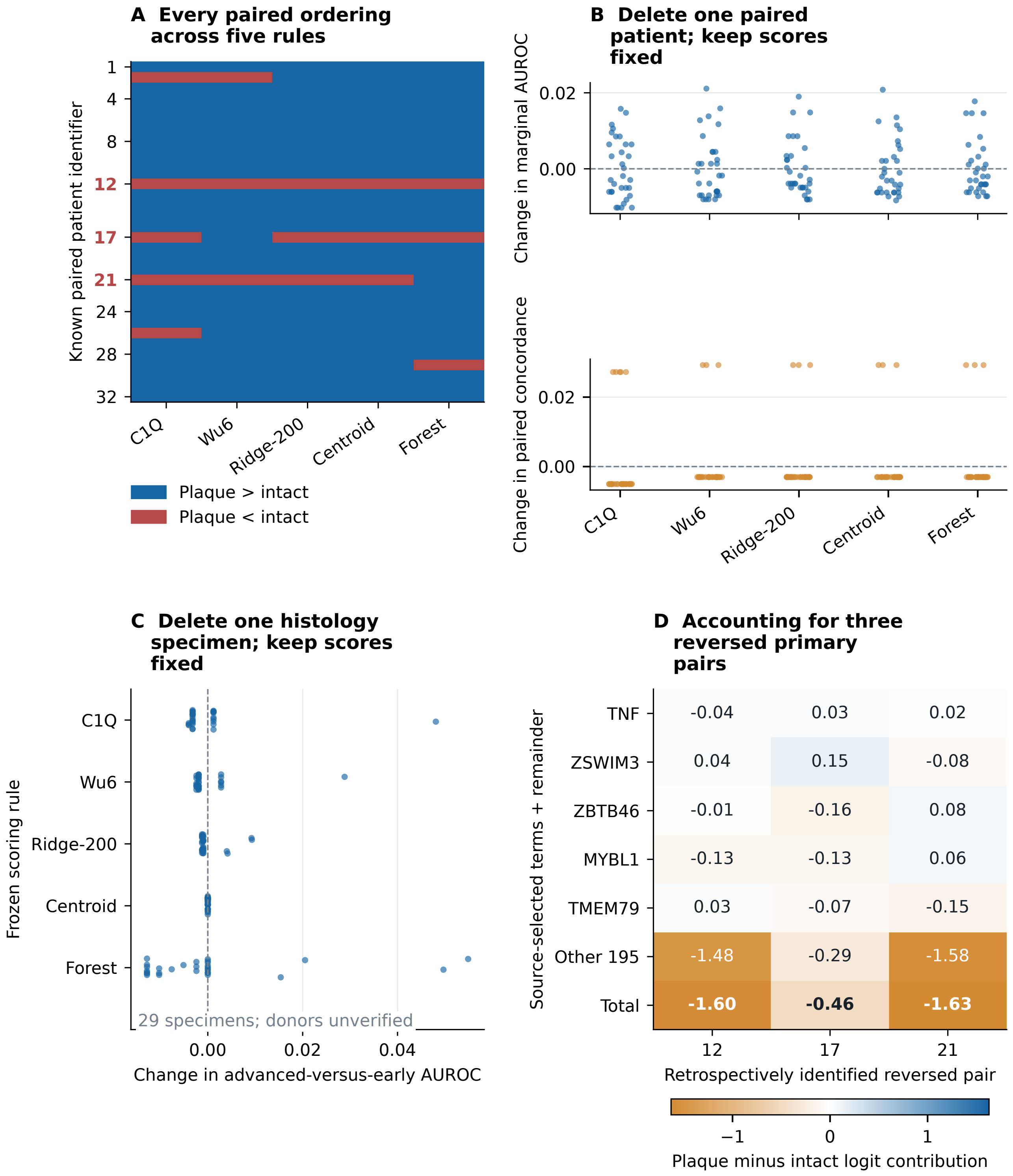

### Figure_11.png

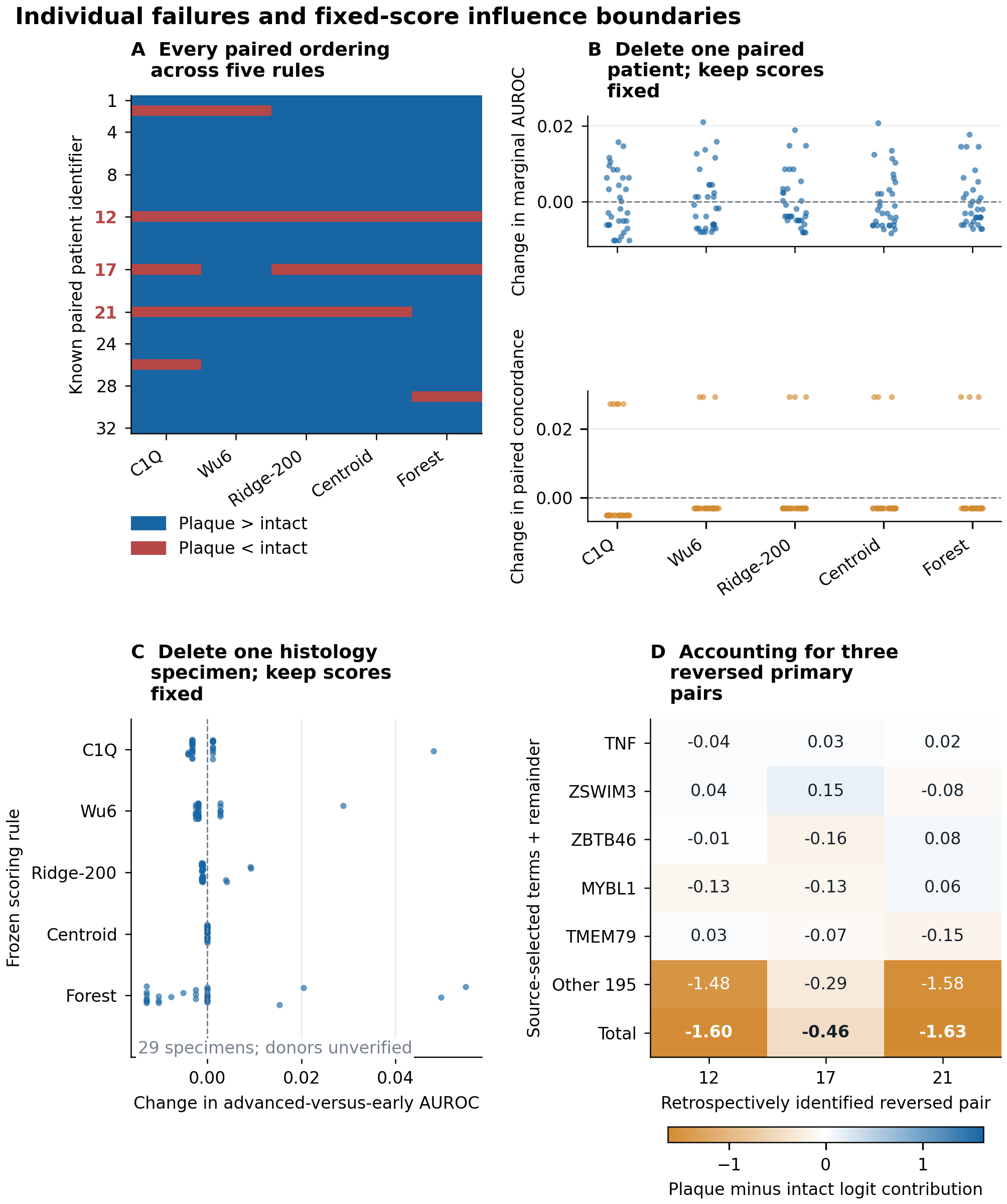

### Figure_S1.pdf

#### Fold-level resolution of anatomical comparisons

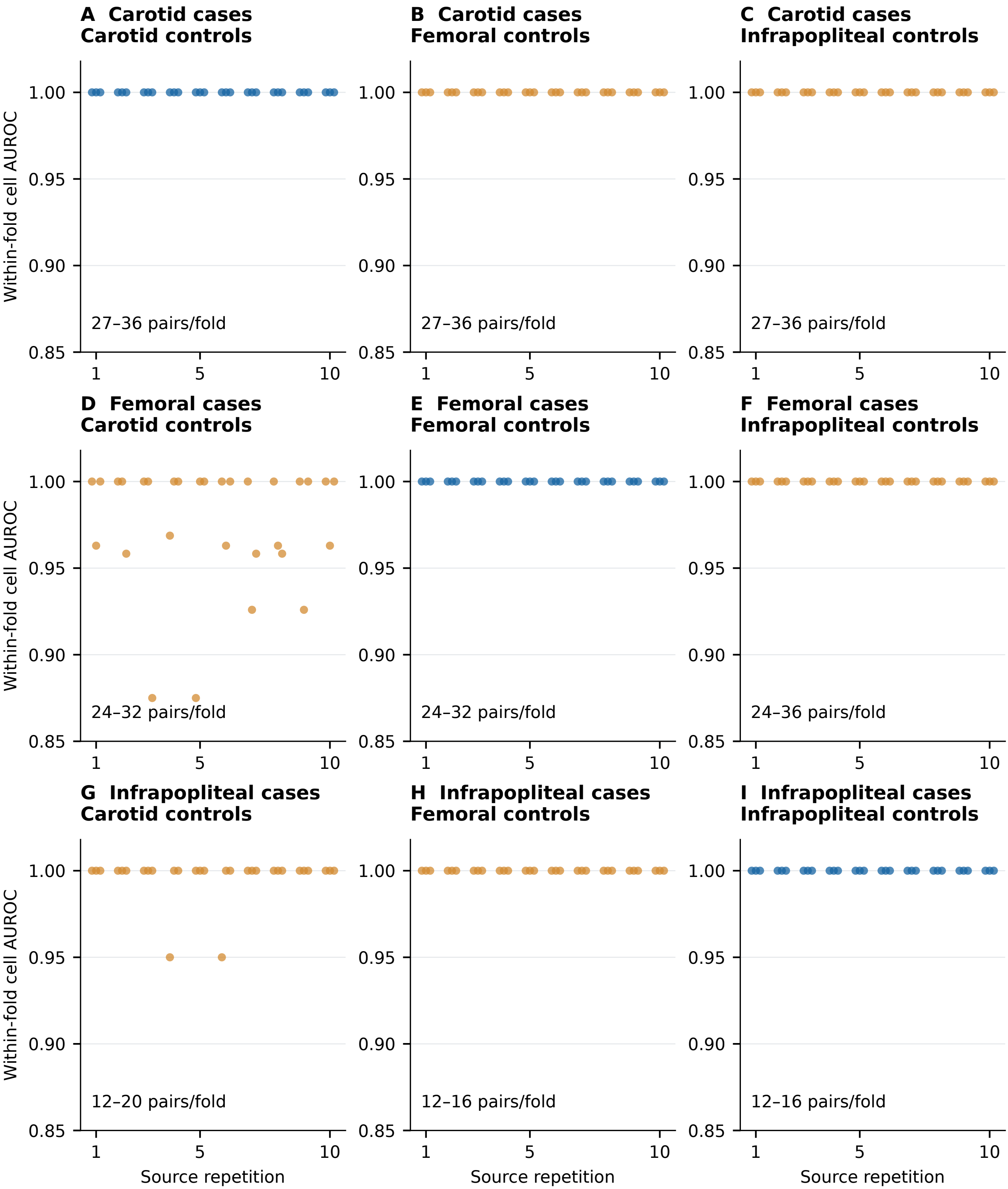

### Figure_S1.png

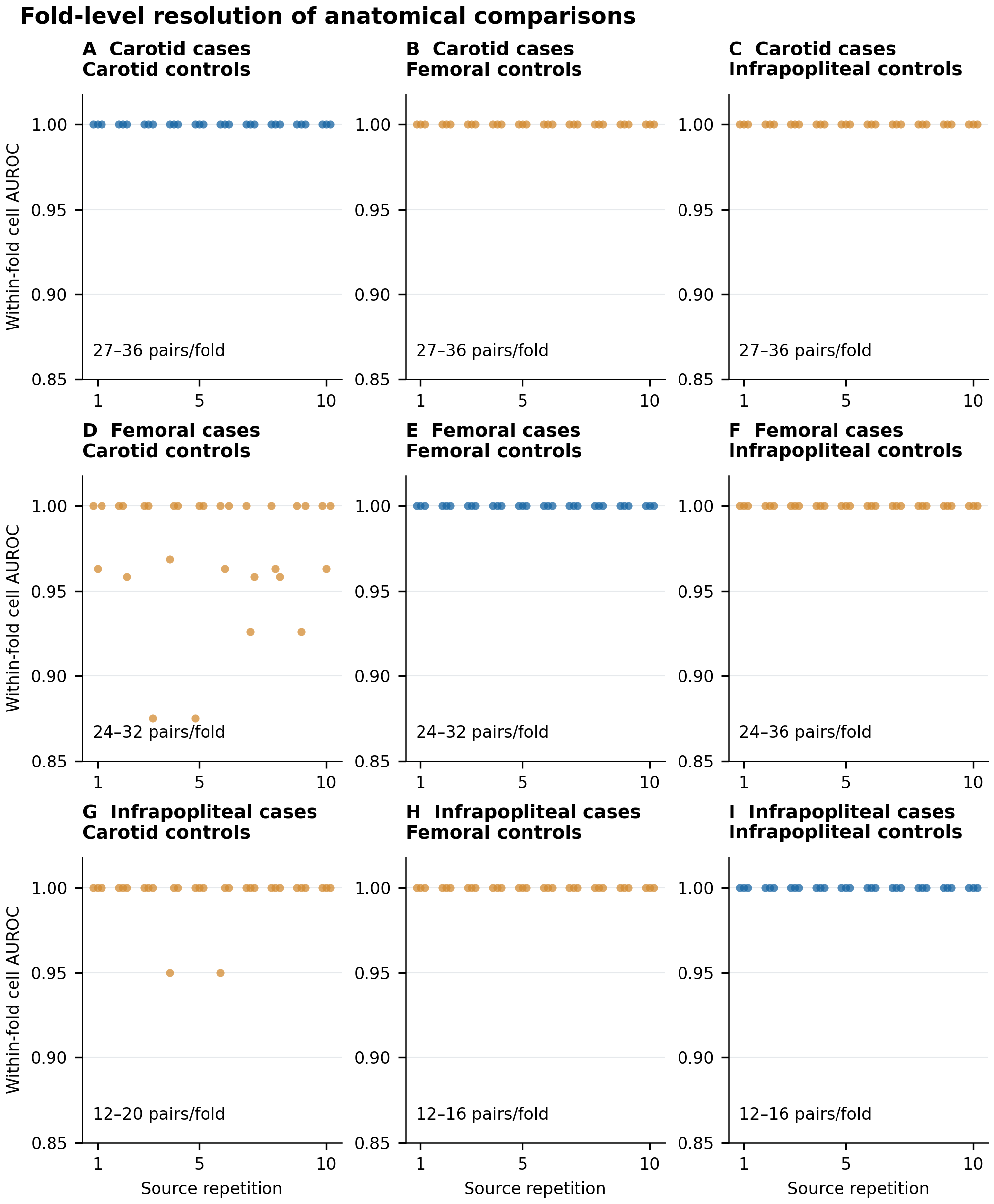

### Figure_S2.pdf

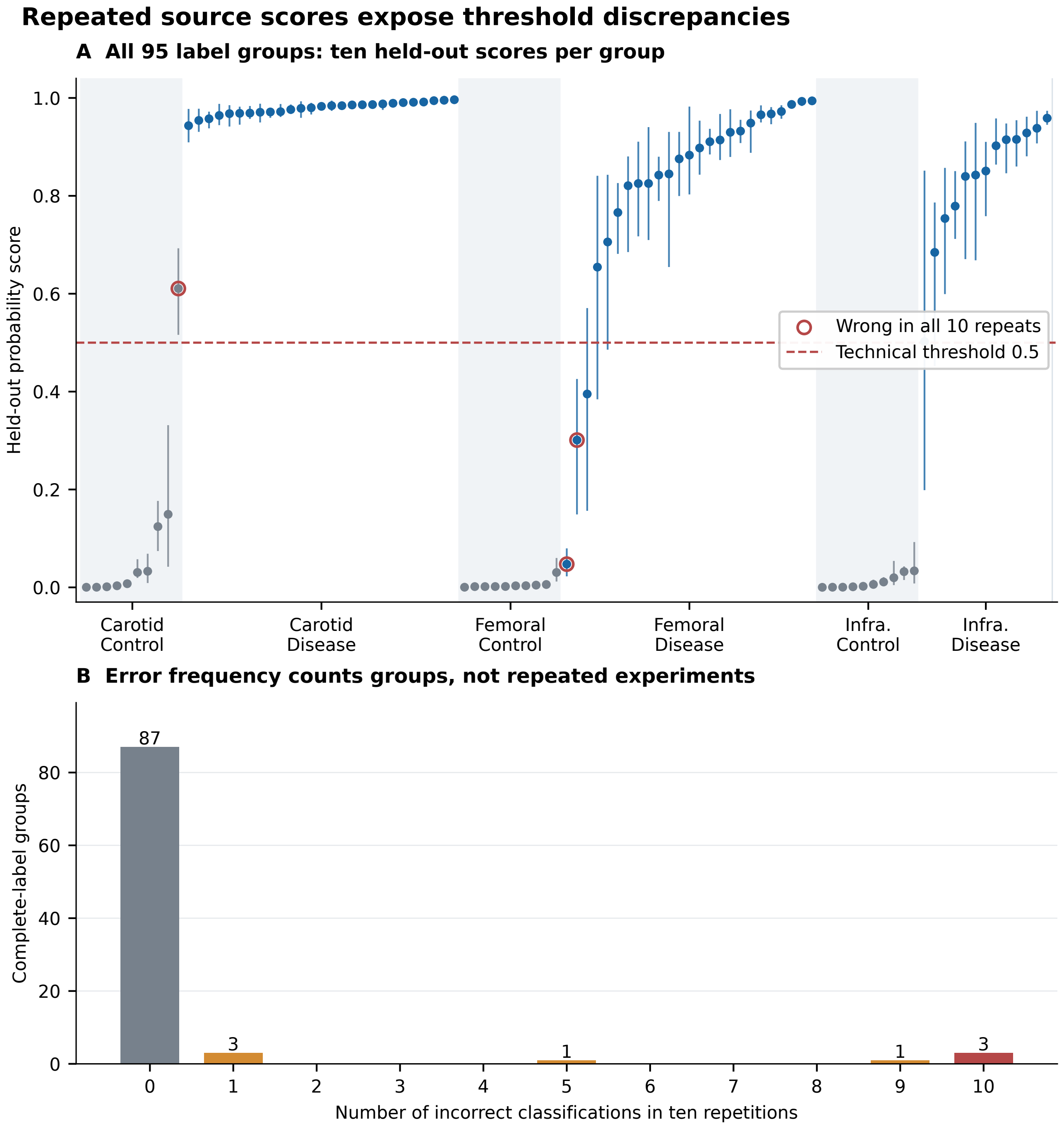

### Figure_S2.png

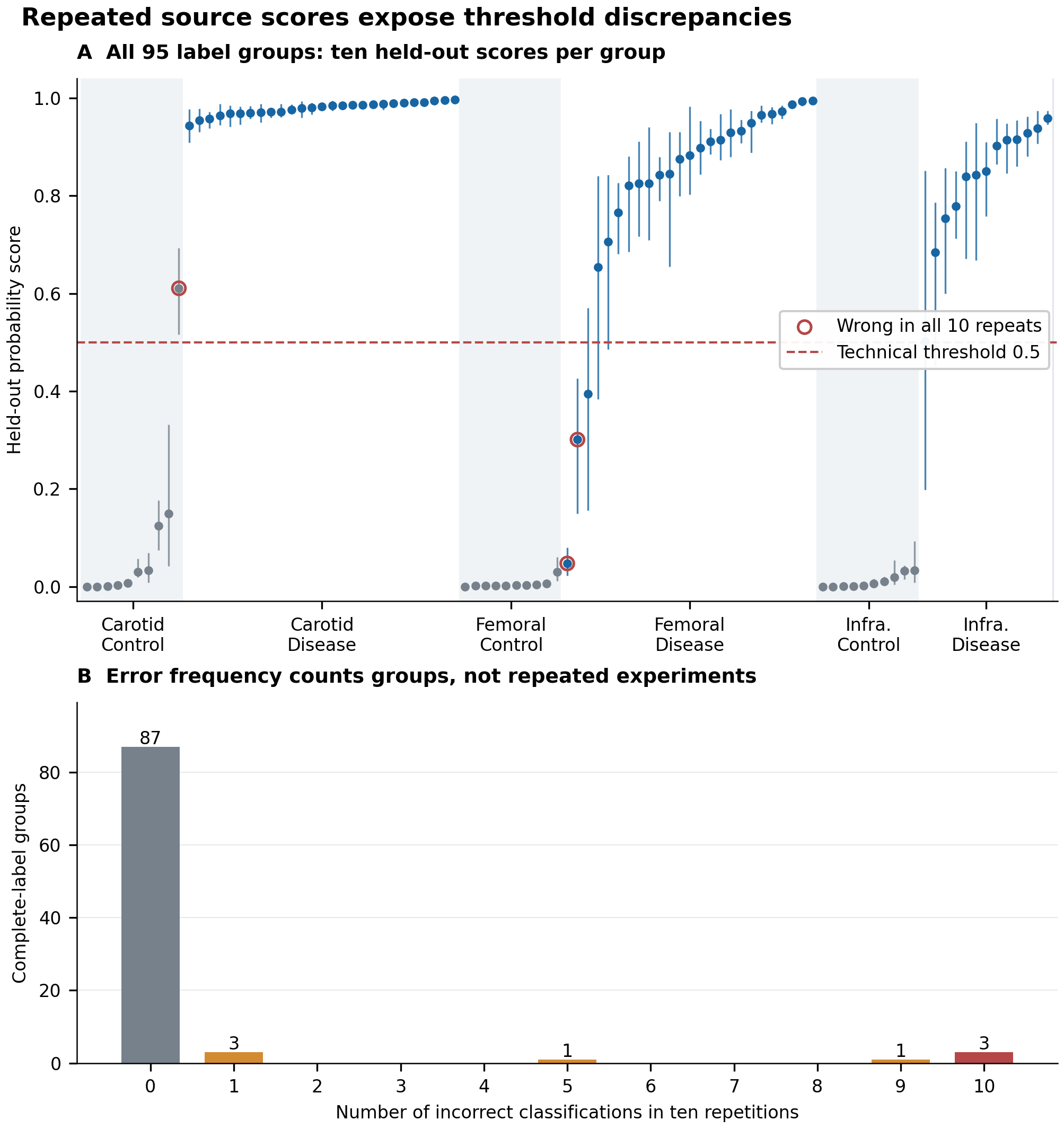
